# Emergence and Persistent Dominance of Omicron BA.2.3.7 Variant in Community Outbreaks in Taiwan

**DOI:** 10.1101/2022.09.27.22280238

**Authors:** Pei-Lan Shao, Hsiao-Chen Tu, Yu-Nong Gong, Hung-Yu Shu, Ralph Kirby, Li-Yun Hsu, Hui-Yee Yeo, Han-Yueh Kuo, Yi-Chia Huang, Yung-Feng Lin, Hui-Ying Weng, Yueh-Lin Wu, Chien-Chih Chen, Tzen-Wen Chen, Kuo-Ming Lee, Chung-Guei Huang, Shin-Ru Shih, Wei J Chen, Chen-Chi Wu, Chong-Jen Yu, Shih-Feng Tsai

## Abstract

Since April 2022, waves of Omicron cases have surfaced in Taiwan and spread throughout the island. Using high-throughput sequencing of the SARS-CoV-2 genome, we have analyzed 2405 PCR-positive swab samples from 2339 individuals and identified the Omicron BA.2.3.7 variant as a major lineage within the recent community outbreak in Taiwan.

## THE STUDY

The coronavirus disease 2019 (COVID-19) pandemic, caused by severe acute respiratory syndrome coronavirus 2 (SARS-CoV-2), originated in the People’s Republic of China (PRC) in late 2019, probably in the city of Wuhan^1–2^. The outbreak of this unusual respiratory disease led to a wide variety of different responses by various countries across the world^3–6^. The response of Taiwan was rapid and based on both its proximity to the PRC and its experiences during the SARS pandemic almost two decades earlier^5,7,8^. The introduction of almost complete travel restrictions on incoming air and sea passengers, long compulsory quarantine periods for the few residents who were allowed to enter Taiwan and a very wide acceptance by the population of social distancing, temperature checks, mask wearing, etc., in all social situations, meant that, unlike many other countries, the entry of the COVID-19 pandemic was delayed significantly^5,9,10^. Until April 2022, there were only limited breakthroughs, all of which were quickly contained. Hence, Taiwan provides a unique opportunity to explore what happened when the Omicron variant finally evaded the controls put in place by the Taiwan government and began to spread though the country’s population. The Taiwanese population had not been exposed on a large scale to any of the virus variants before Omicron. By the time the virus began to spread widely in April 2022, there had been around seventeen thousand recorded cases of COVID-19, and most of them were infected by the Alpha variant. Almost all of the cases in our current study had not been infected with COVID-19 before. Furthermore, the vaccination rates of Taiwan’s population at that time, were 82.7%, 78.0%, and 59.1% with single dose, double doses, and triple doses, respectively. The vaccines used in Taiwan before May 2022 were the Oxford–AstraZeneca COVID-19 vaccine, the Pfizer-BioNTech vaccine, the Moderna COVID-19 vaccine, the Johnson & Johnson’s Janssen (J&J/Janssen) COVID-19 vaccine, and The Median COVID-19 vaccine (a protein subunit COVID-19 vaccine made in Taiwan). Most Taiwanese received doses of the first three vaccines.

Very few COVID-19 cases had occurred in Taiwan during 2020 and 2021. Clustered infections were reported in May and June 2021, mainly in northern Taiwan. Even at their peak, only hundreds of positive cases were recorded by Taiwan’s Centers for Disease Control (CDC). At the beginning of 2022, a number of Omicron infection clusters were noted, first in northern Taiwan, and very soon after new cases began to exceed fifty thousand per day with outbreaks affecting the entire country (Figure 1A) (https://nidss.cdc.gov.tw/nndss/disease?id=19CoV). To gain insights into community transmission and to monitor viral evolution, we deployed a genomic surveillance protocol at National Taiwan University Hospital Hsinchu Branch (NTU-HCH) whereby left-over nasal swab samples that had been detected by PCR to be positive were whole-genome sequenced (see Appendix methods for details).

**Figure 1:**
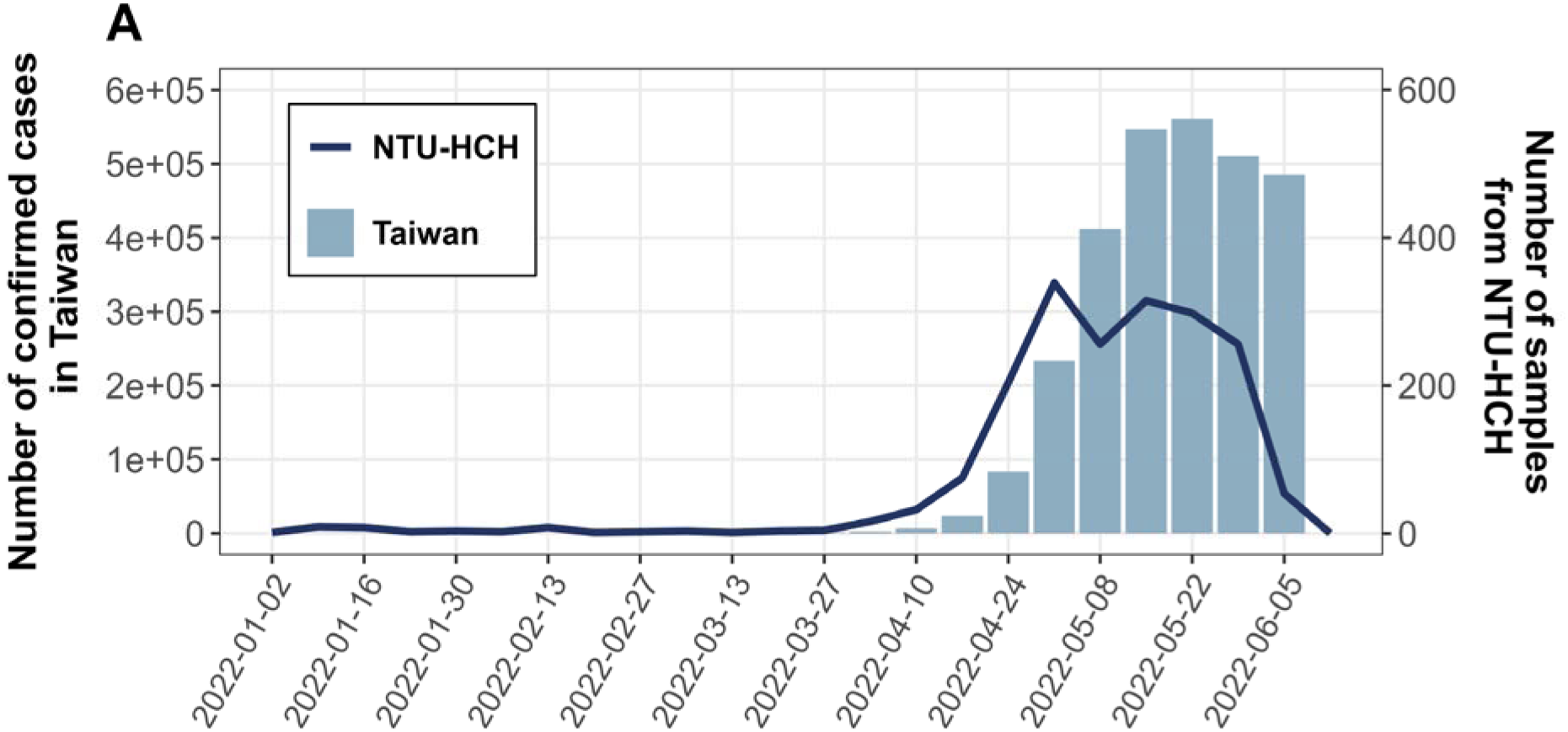
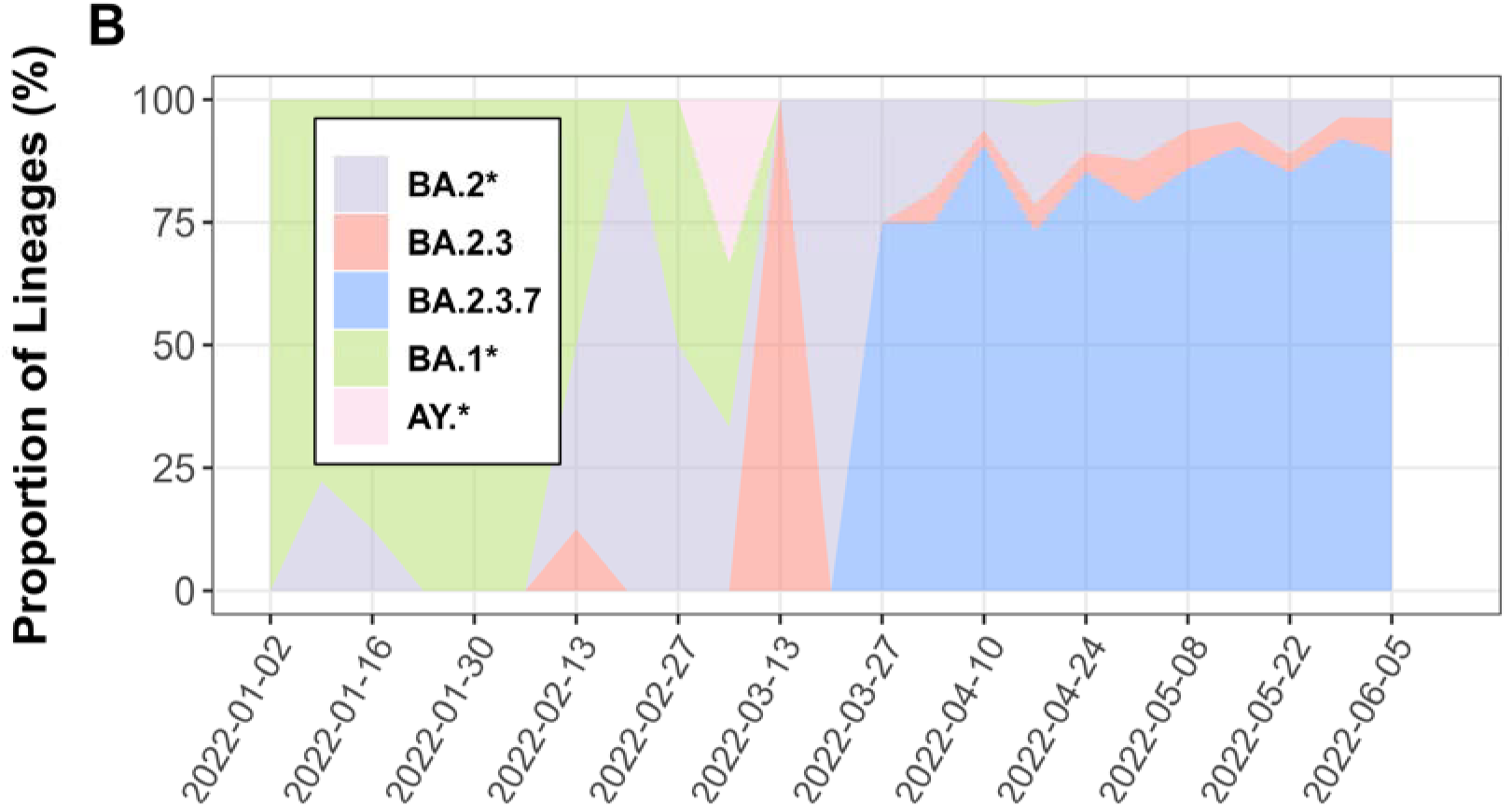
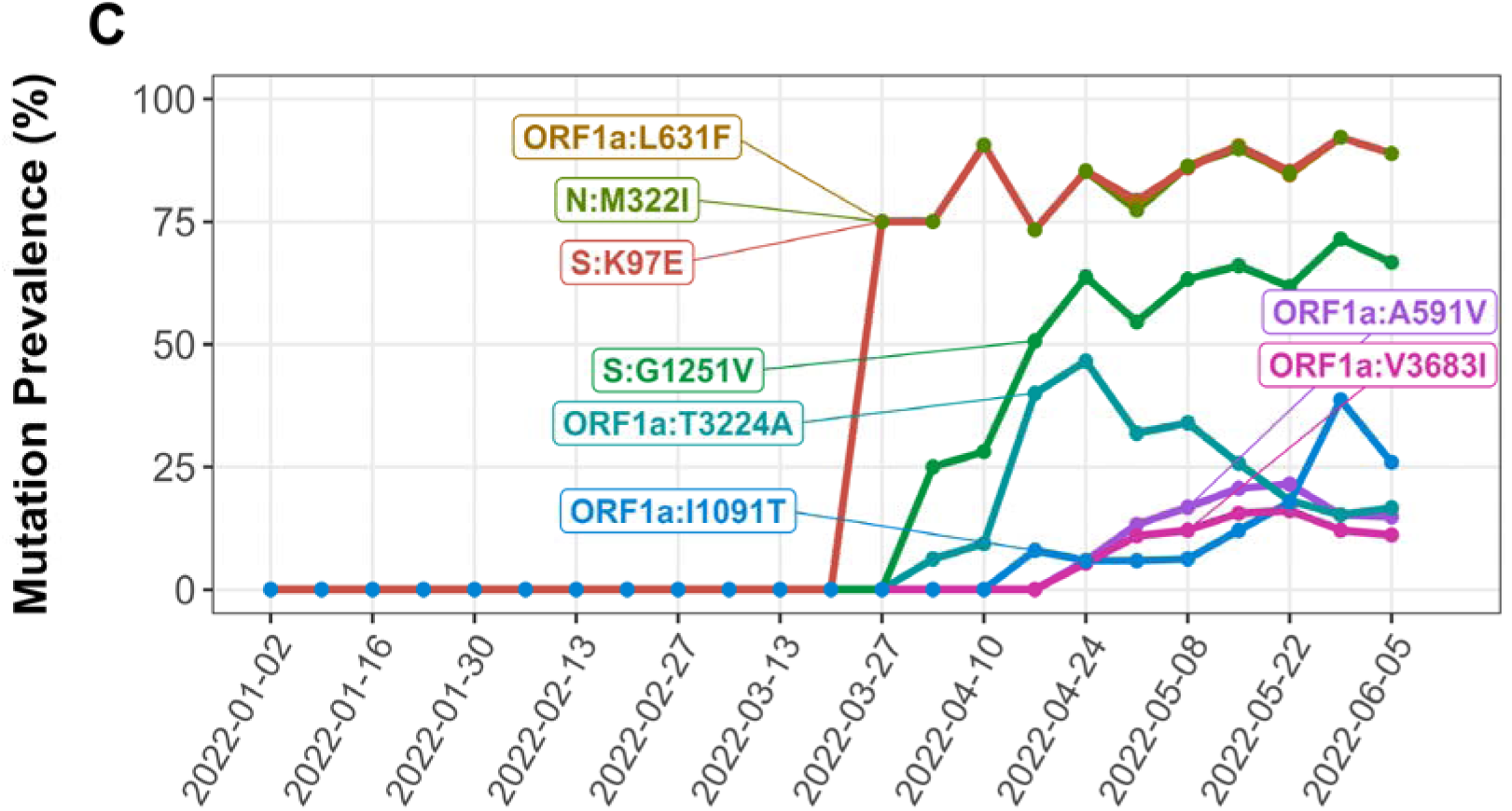
Weekly statistics for confirmed cases and sequenced samples, lineages distribution, and mutations prevalence in the NTU-HCH surveillance program. This figure was constructed using the publicly available data of Taiwan CDC (https://nidss.cdc.gov.tw/nndss/disease?id=19CoV). **1A.** The number of COVID-19 confirmed cases in Taiwan and the sequenced samples of NTU-HCH from January (the first week) to early June (the 23^rd^ week) in 2022. **1B.** Lineages distribution of NTU-HCH strains. Most of them were identified as BA.1 or BA.2 lineages and sub-lineages annotated with an asterisk sign (*) before the wave of community outbreak in end of March. Since then, the BA.2.3.7 lineage became the dominant variant circulating in Taiwan. **1C.** Proportion of the different signature mutations in the period of January 1^st^ to June 6^th^, 2022. Note that there was a sharp increase of the Omicron variant 2.3.7 from week 13 (March 27^th^– April 2^nd^) onwards, and the proportion of this Omicron variant remained high for at least ten weeks. The three signature mutations (S:K97E, N:M322I, ORF1a:L631F) of BA.2.3.7 are completely overlapped. A steady increase of sequences positive for S:G1251V was noted from week 14 (April 3^rd^–9^th^) and reached plateau at week 17 (April 24^th^–30^th^).

To ensure data quality, we have only submitted genomic data to GISAID on those sequences that had a greater than 98% coverage of the 29903 bp SARS-CoV-2 target genome. The same set of high-quality sequences were used for tracking the signature mutations in the viral samples (**Table 1**) and for phylogenetic analysis (**Figure 2**, see Appendix methods for details). Overall, 2405 samples in all five batches met the above criterion and this generated 2043 sequences (84.9% pass rate), and 1966 sequences were selected for GISAID submission. (**Appendix Table 1**)

**Figure 2.**
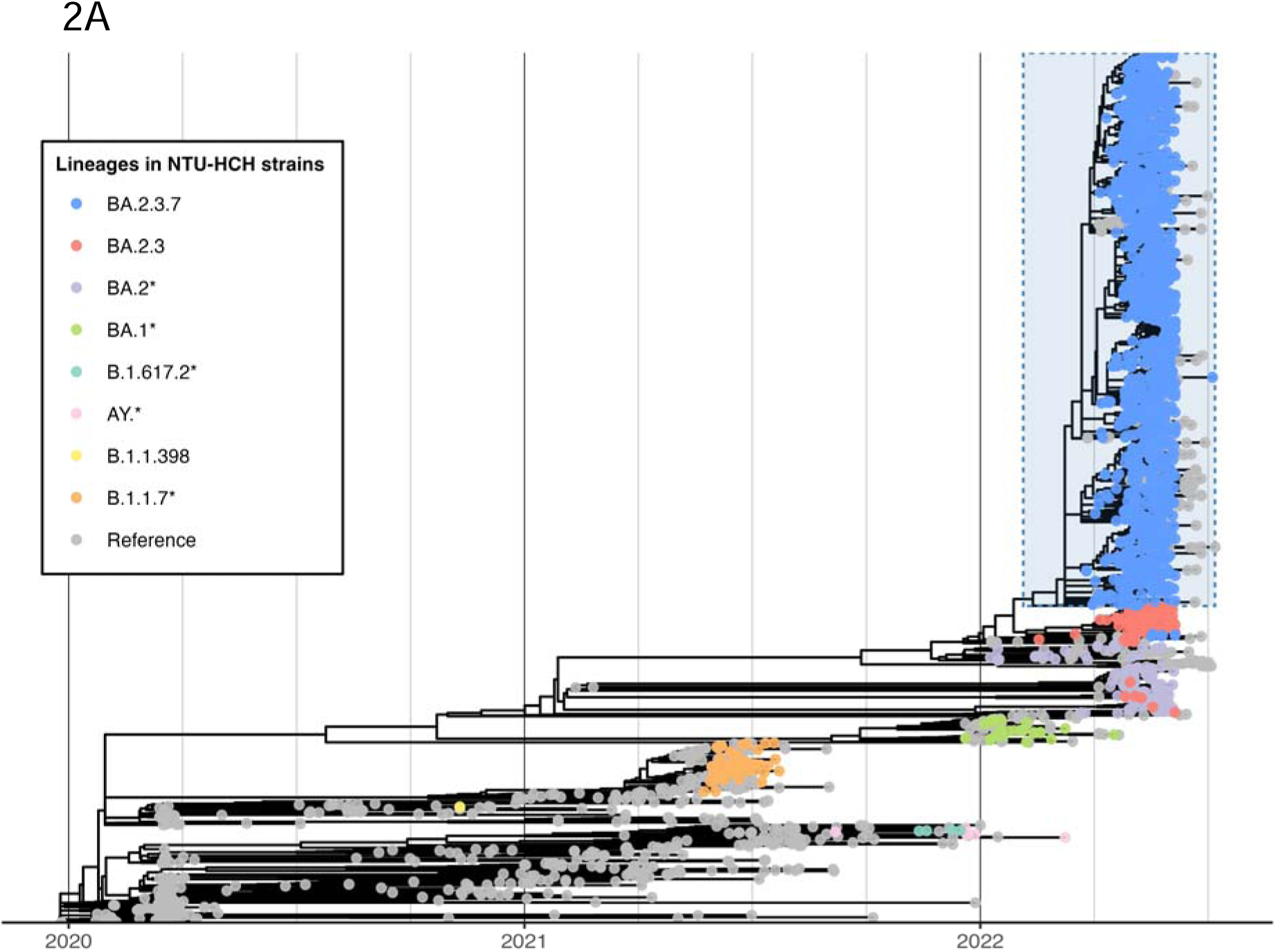
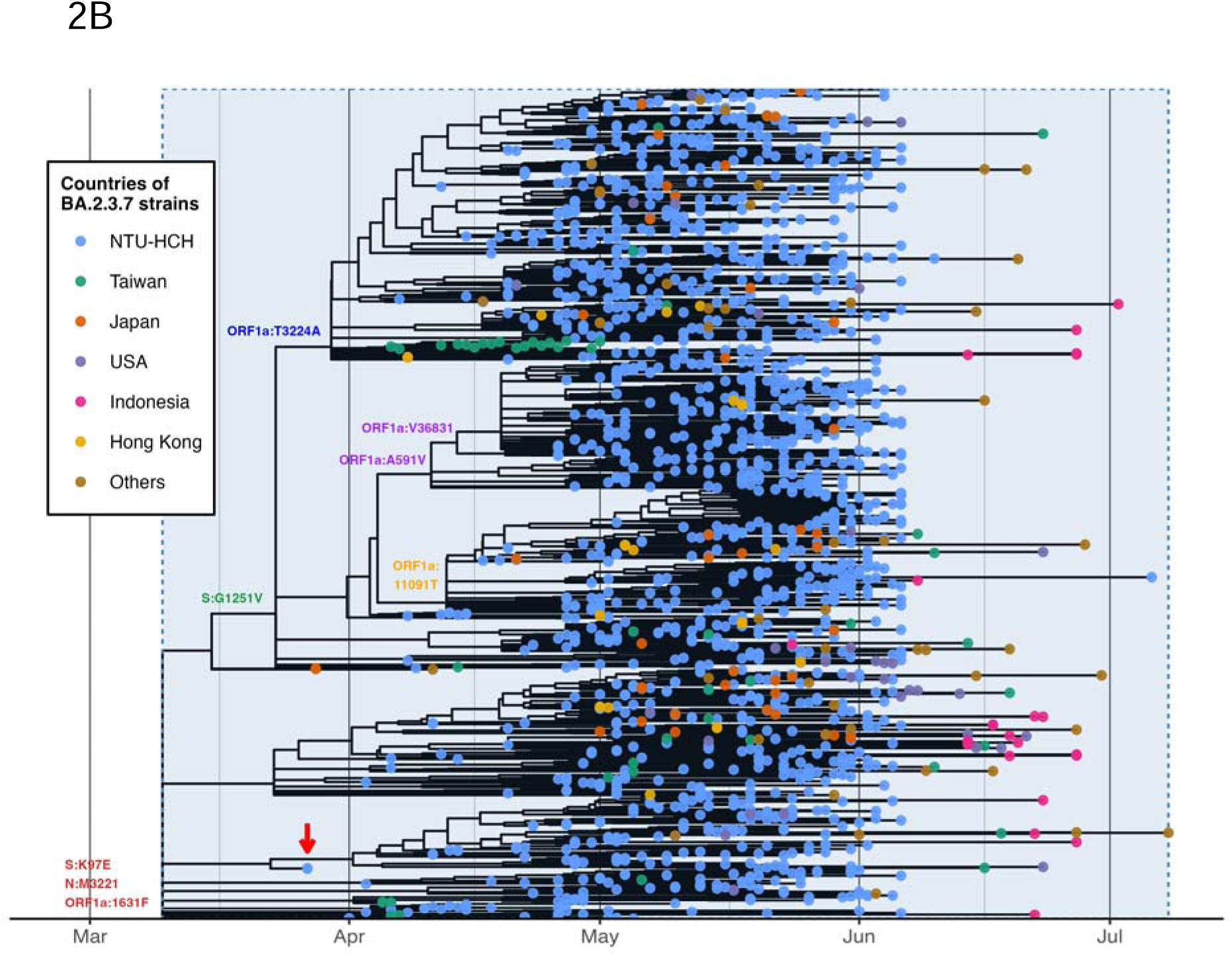
Phylogenetic analysis of the SARS-CoV-2 sequences. **2A**. We used 1966 sequences from NTU-HCH and 881 sequences from GISAID as a framework for constructing the phylogenetic tree. Lineages of NTU-HCH strains are annotated in different colors, and an asterisk sign (*) represents the collection of a specific lineage with its sub-lineage. The BA.2.3.7 strains were dominantly circulating in Taiwan from March 2022, highlighted by light blue in the tree. **2B**. We analyzed 1584 BA.2.3.7 sequences submitted to GISAID from the current study together with 228 BA.2.3.7 sequences deposited to GISAID by other groups. The position of signature mutations are indicated; three (S:K97E, N:M322I, ORF 1a:L631F) are located at the origin of the B.A.2.3.7 lineages. S:G1251V is mapped at a major branch in the upper trunk of the tree, under which three minor branches can be defined by mutations in ORF1a:T3224A in blue, ORF1a:A591V and ORF1a:V3683I in purple, and ORF1a:I1091T in orange. Sample origins are color coded. An arrow in red denotes the index case collected from Taiwan on March 27^th^, 2022.

**Table 1.**
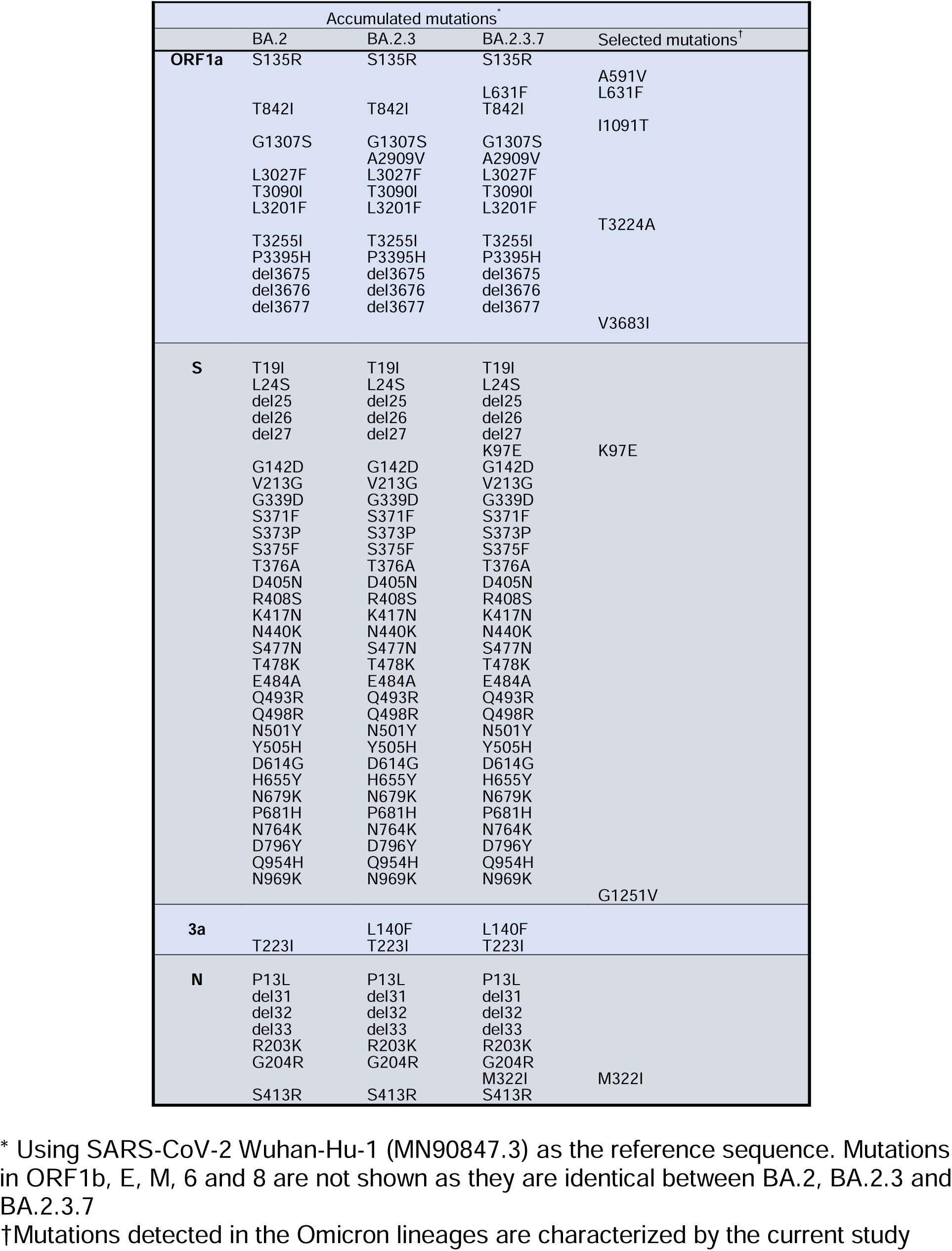
Mutations in the BA.2.3.7 sublineages.

We analyzed the assembled viral genome sequences (**Appendix Table 2**) and tracked the lineages and significant amino acid changes in the Omicron samples collected during year 2022 (**Figures 1B and 1C**). Comparing the later three data sets (batch 3 to batch 5), we discovered that three amino acid changes (ORF1a:L631F, S:K97E, N:M322I) occurred only after the fourth sequencing batch. The percentage of sequences containing the signatures progressed steadily from 62 % in batch 4 to 85% in batch 5. All batch 3 isolates belonged to the BA.1 or BA.2 classification. This suggests that the rapid increase of cases in Taiwan in April and May, from previously zero cases per day, to almost reaching a hundred thousand per day, came from a strain, BA.2.3.7, that might have been involved in a founder effect.

To construct the framework of the phylogenetic tree, first we took 1966 genome sequences from this study and analyzed them in the global context of 872 GISAID reference sequences (**Figure 2A**). On this basis, we then zoomed in and compared the 1584 Omicron sequences of the current study against the 228 Omicron BA.2.3.7 strains from the GISAID database. These sequences were reported from 21 countries, including 51 from Taiwan (**Table 2**). We have conducted phylogenetic analysis using the Pango-dynamic nomenclature system^11^.

**Table 2.** Submitted sequences of SARS-CoV-2 BA2.3.7 from different countries.

| Country | Count |
| --- | --- |
| Taiwan | 51 |
| Japan | 44 |
| USA | 37 |
| Indonesia | 27 |
| Hong Kong | 22 |
| Australia | 7 |
| Denmark | 6 |
| Canada | 5 |
| Singapore | 4 |
| South Korea | 4 |
| Philippines | 3 |
| Thailand | 3 |
| France | 3 |
| Cambodia | 2 |
| Austria | 2 |
| Sweden | 2 |
| Germany | 2 |
| Spain | 1 |
| Vietnam | 1 |
| Slovenia | 1 |
| New Zealand | 1 |
| Total | 228 |

\* The case number does not include the contribution from this study.

It is evident that this novel lineage BA.2.3.7 with three amino acid changes (ORF1a:L631F, ORF1a:S:K97E, and N:M322I) was circulating dominantly in Taiwan over the study period. Notably, the first BA.2.3.7 strain identified in the epidemic in Taiwan was collected on March 27^th^, 2022, and, since that point, we were able to detect several genomic changes affecting this Omicron lineage. For example, a new mutation, G1251V (**Figure 2B**, marked in green) in the Spike protein, was detected from April onwards, and this particular circulating lineage then rapidly spread across Taiwan.

While this paper was in preparation, we became aware that a number of viral sequences with the same signature mutations had been reported from Taiwan. Even though the number was relatively small (51 cases) as compared to our serial investigation at a single medical center, the four locations in Taiwan of the reported cases are different from our collection point at the hospital. Thus, this new lineage would seem to be detectable broadly across Taiwan.

Notably, over the same time period, other Asia Pacific countries also reported a significant cumulative prevalence of the BA.2.3.7 variant (**Table 2**). Among the 44 Omicron BA.2.3.7 strains reported from Japan, two of the affected individuals had travel history to Vietnam and 41 to Taiwan, suggesting significant silent outward transmission from Taiwan. In contrast, BA.2.3.7 forms less than 0.5% of the sequences reported in either California, USA, or globally. The emergence of Omicron BA.2.3.7 in Asia is remarkable. While there is no reliable genomic data from the early cases in Malaysia and Vietnam, our phylogenic analysis and the metadata from GISAID suggests that travel between Asian countries contributed to the rapid spread of this unique Omicron lineage.

Finally, this study has limitation, as we have conducted the genomic surveillance in only one medical center, therefore, the observed dominance of BA.2.3.7 might be due to clustering of cases.

## CONCLUSIONS

In conclusion, our genomic dataset is uniquely valuable for understanding how a major COVID-19 outbreak occurs in a naïve and vaccinated population in Taiwan with a very limited number of entry events, and the dominant circulation of BA.2.3.7 in Taiwan probably is due to genetic drift or a founder effect, although it is also possible that increased transmissibility or vaccine evasion played some part. As Asian countries began to move from zero tolerance to more open COVID-19 policies, continued surveillance of SARS-CoV-2 using NGS is important. Early detection of significant viral evolution events in endemic areas will help to minimize any future disruption due to a new variant.

## Data Availability

All data produced in the present study are available upon reasonable request to the authors

## ACKNOWLEDGEMENTS

This work was supported by Ministry of Health and Welfare of Taiwan (MOHW110-TDU-C-222-000010) to S.F.T., by the Research Center for Emerging Viral Infections from The Featured Areas Research Center Program within the framework of the Higher Education Sprout Project by the Ministry of Education (MOE) of Taiwan, the Ministry of Science and Technology (MOST) of Taiwan (MOST 111-2321-B-182-001, MOST 111-2634-F-182-001, MOST 110-2222-E-182-004, and MOST 111-2221-E-182-053-MY3), and the National Institute Of Allergy And Infectious Diseases of the National Institutes of Health under Award Number 5U01AI151698-02 to Y.N.G. and S.R.S.. The authors wish to thank Y Henry Sun and Hsiao-Hui Tso for critical reading of the manuscript, to Yi-Chun Tsai, Shuo-Peng Chou, Jhen-Rong Huang, and Yuan-Yu You for sample collection and nucleic acid preparation, to Tai-Yun Lin, Yu-Chen Huang and Chiao-Chan Wang for library construction and sequencing operation.

## AUTHOR CONTRIBUTION

P.L.S. Collecting clinical specimens and performed clinical data analysis.

H.C.T. Performing COVIDseq and manuscript preparation

Y.N.G. Phylogenetic analyses and manuscript preparation.

H.Y.S. Mutation signature analyses and manuscript preparation.

R.K. Interpretation and manuscript preparation

L.Y.H. Collecting clinical samples, nucleic acid extraction

H.Y.Y. Collecting clinical samples, nucleic acid extraction

H.Y.K. Clinical data management

Y.C.H. Clinical data interpretation

Y.F.L. Genomic data analysis

H.Y.W. Genomic data analysis

Y.L.W. Data interpretation and manuscript preparation

C.C.C. Collecting samples and viral analysis

T.W.C. Collecting samples and clinical data analysis

K.M.L. Establishing protocols and sample processing

C.G.H. Establishing protocols and sample processing

S.R.S. Virology study and functional analysis

W.J.C. Data interpretation and manuscript preparation

C.C.W. Project management and manuscript preparation

C.J.Y. Project management and manuscript preparation

S.F.T. Project management and manuscript preparation

## ABOUT THE AUTHORS

Dr. Shao is an assistant professor in the Department of Laboratory Medicine and Pediatrics, National Taiwan University College of Medicine, Taiwan. Her main research interests include infectious disease, clinical virology, clinical microbiology, and vaccines.

Dr. Tu is postdoctoral fellow in the Institute of Molecular and Genomic Medicine, National Health Research Institutes, Taiwan. Her research interests include NGS technology, cancer and genomic research.

Dr. Gong is an assistant professor in Research Center for Emerging Viral Infections and International Master Degree Program for Molecular Medicine in Emerging Viral Infections, Chang Gung University, Taiwan. His research interests include bioinformatics, phylogenetics, machine learning, and viral evolution.

The authors wish to thank Y Henry Sun and Hsiao-Hui Tso for critical reading of the manuscript, Yi-Chun Tsai, Shuo-Peng Chou, Jhen-Rong Huang, and Yuan-Yu You for sample collection and nucleic acid preparation, and Tai-Yun Lin, Yu-Chen Huang and Chiao-Chan Wang for library construction and sequencing operation.

## APPENDIX

### METHODS

We applied Illumina COVIDSeq amplicon sequencing technology to the whole-genome analysis of SARS-CoV-2. RNA was extracted from virus-inactivated swab samples. Complementary DNA synthesis was carried out using the reagents provided by the manufacturer, by which the SARS-CoV-2 genome underwent reverse transcription and was then amplified in 98 overlapping amplicons, together with appropriate human controls. In the final optimized procedure, we processed 384 samples at a time, and analyzed the pooled libraries on a single lane of a S4 chip using a NovaSeq6000.

### Patients and RNA extraction

Representative samples were collected from National Taiwan University Hospital Hsinchu Branch (NTU-HCH). Before the major outbreak, nearly 300 samples were surveyed in January-April, 2022. On the other hand, we sequenced close to 2000 samples during the major outbreak in May, representing approximately 0.1% of the estimated 2 million confirmed cases of Taiwan. Samples for RNA extraction were collected in 3 mL sterile viral transport medium (VTM) tubes and consisted of 2405 nasopharyngeal swabs belonging to 2339 patients. RNA was prepared by automated extraction using TANBead Nucleic Acid Extraction kits REF M665A46 (Taiwan Advanced Nanotech Inc.) and the QIAsymphony SP protocol (QIAGEN). This study was reviewed and approved by the Research Ethics Committee (110-110-E) of NTU-HCH.

### COVIDseq

We carried out the sequencing using Illumina COVIDSeq Test kits (RUO version) according to the manufacturer’s instructions. The workflow consists the following steps: cDNA synthesis, then virus target amplification using V3 nCov-2019 primers, followed by library preparation and library pooling. Subsequently, 98 SARS-CoV-2 targets and 11 human targets, the latter acting as controls, were analyzed on a NovaSeq 6000 instrument using 2×151-bp paired-end reads. Next, we used Illumina DRAGEN COVID Lineage app version 3.5.9 (base on pangolin 4.1.2 pangolin-data 1.12 and NextClade 1.11.0) in the BaseSpace Sequence Hub for rapid analysis.

### Phylogenetic analysis

A set of 1966 NTU-HCH sequences were deposited to the Global Initiative on Sharing All Influenza Data (GISAID)^1^ with the following epi accession numbers: EPI_ISL_14192849 to EPI_ISL_14192840, EPI_ISL_14191496 to EPI_ISL_14191488, EPI_ISL_14191320 to EPI_ISL_14190364, and EPI_ISL_14190353 to EPI_ISL_14189364. To investigate the global transmission, 249 BA.2.3.7, 282 global, and 403 additional sequences from Taiwanese were retrieved from GISAID as of July 2022. A total of 2847 sequences, including 1966 from NTU-HCH and 881 reference sequences (Appendix Table 3: GISAID_Acknowlegement) that had met the quality standard, were used for the analysis. The phylogenetic tree was initially constructed using Nextstrain CLI (command-line interface) (version 3.2.5)^2^, and annotated and visualized using ggtree package^3^.

---

**Appendix Table 1.** Sequences selected for GISAID submission and phylogenetics analysis.

| Batch | Run | Coverage |  |  |  |  |  | Total |
| --- | --- | --- | --- | --- | --- | --- | --- | --- |
|  |  | >=98 | 90-97 | 80-89 | 70-79 | 60-69 | <60 |  |
| 1 | 1 | 77 | 12 | 0 | 4 | 0 | 1 | 94 |
| 2 | 2 | 63 | 0 | 5 | 3 | 4 | 18 | 93 |
| 3 | 3 | 43 | 14 | 4 | 2 | 5 | 38 | 106 |
| 4 | 4 | 140 | 17 | 2 | 3 | 0 | 30 | 192 |
| 5 | 5A | 360 | 15 | 5 | 2 | 0 | 2 | 384 |
|  | 5B | 331 | 30 | 11 | 6 | 1 | 5 | 384 |
|  | 5C | 304 | 47 | 15 | 14 | 4 | 0 | 384 |
|  | 5D | 366 | 16 | 2 | 0 | 0 | 0 | 384 |
|  | 5E | 359 | 24 | 0 | 1 | 0 | 0 | 384 |
|  | NTU-HCH* | 1966 | 163 | 44 | 31 | 14 | 93 | 2311 |
|  | Total<sup>#</sup> | 2043 | 175 | 44 | 35 | 14 | 94 | 2405 |

\* Count of NTU-HCH cases, from Batch (Run-2) through Batch 5 (Runs 5A-5E)
<sup>#</sup> Total count of all three hospitals

**Appendix Table 2.** Samples and cases collected for this study as analyzed by the Illumina COVIDSeq system on NovaSeq6000.

| Batch | Sampling time period | Hospital source | Sample number | Case number | Sequencing run |
| --- | --- | --- | --- | --- | --- |
| 1st | 2021 (May) | WKH 12 CGMH 82 | 94 | 93 | SP 2x51bp |
| 2nd | 2021 (May-July) | WKH 12* CGMH 82*<br>NTU-HCH 93 | 93# | 69# | S4 lane 2x151bp |
| 3rd | 2021 (Dec)~2022 (Feb)+2020(Nov)~2021(April~Nov)11 cases | NTU-HCH 106 | 106 | 66 | S4 lane 2x151bp |
| 4th | 2022 (Feb 25~April 27)<br>April 27 (1 case) | NTU-HCH 192 | 192 | 191 | S4 lane 2x151bp<br>(288 sample/ lane) |
| 5th | 5A_2022<br>(April 26~May 3) | NTU-HCH 384 | 384 | 384 | S4 lane 2x151bp |
|  | 5B_2022<br>(May 3~May 23) | NTU-HCH 384 | 384 | 384 | S4 lane 2x151bp |
|  | 5C_2022<br>(May 17~June 6) | NTU-HCH 384 | 384 | 384 | S4 lane 2x151bp |
|  | 5D_2022<br>(May 4~June 6) | NTU-HCH 384 | 384 | 384 | S4 lane 2x151bp |
|  | 5E_2022<br>(May 18~June 6)<br>+ July 6 (1 case) | NTU-HCH 384 | 384 | 384 | S4 lane 2x151bp |

\*Repeat from the first batch
#New samples from NTU-HCH

**Appendix Table 3.**
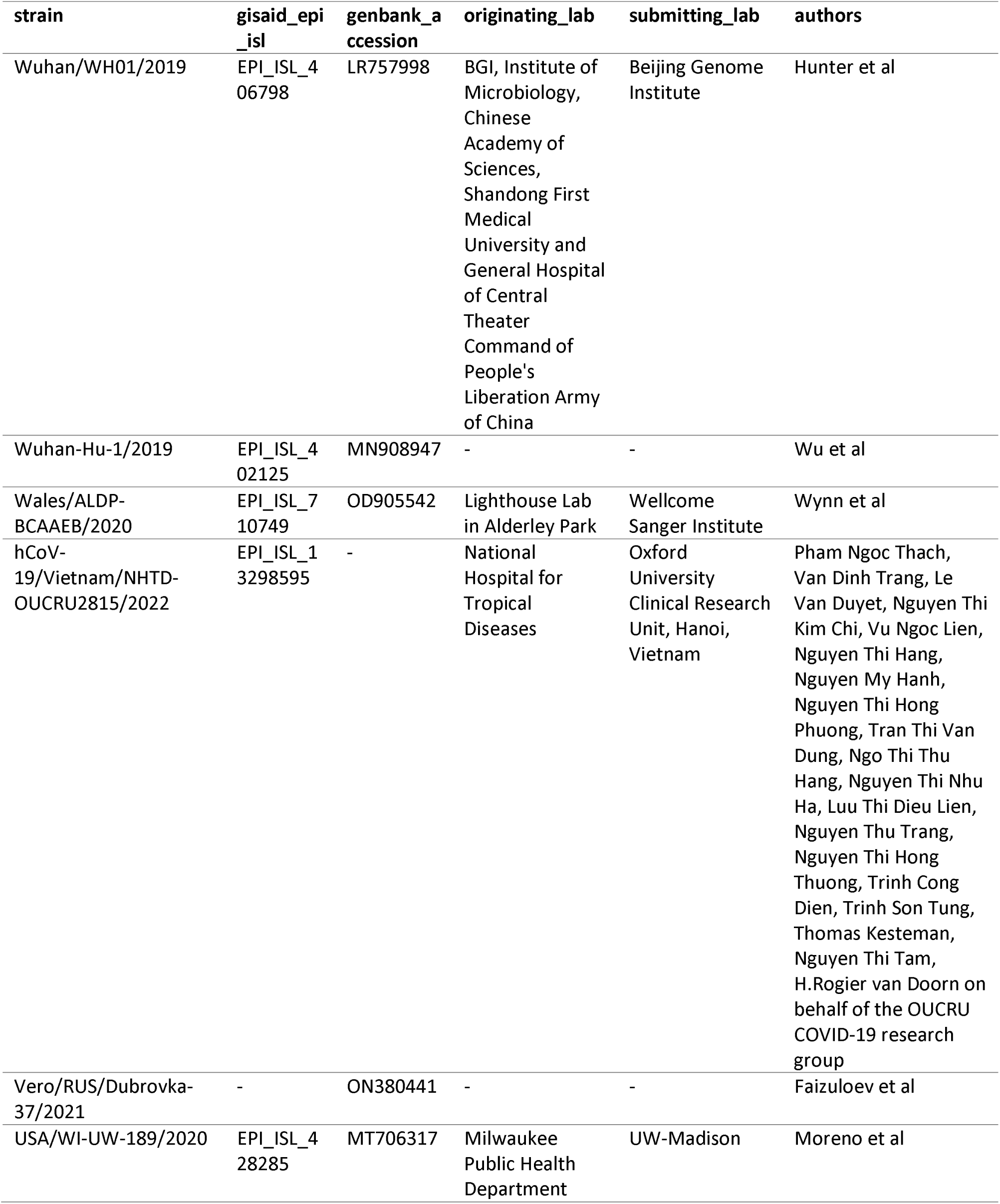
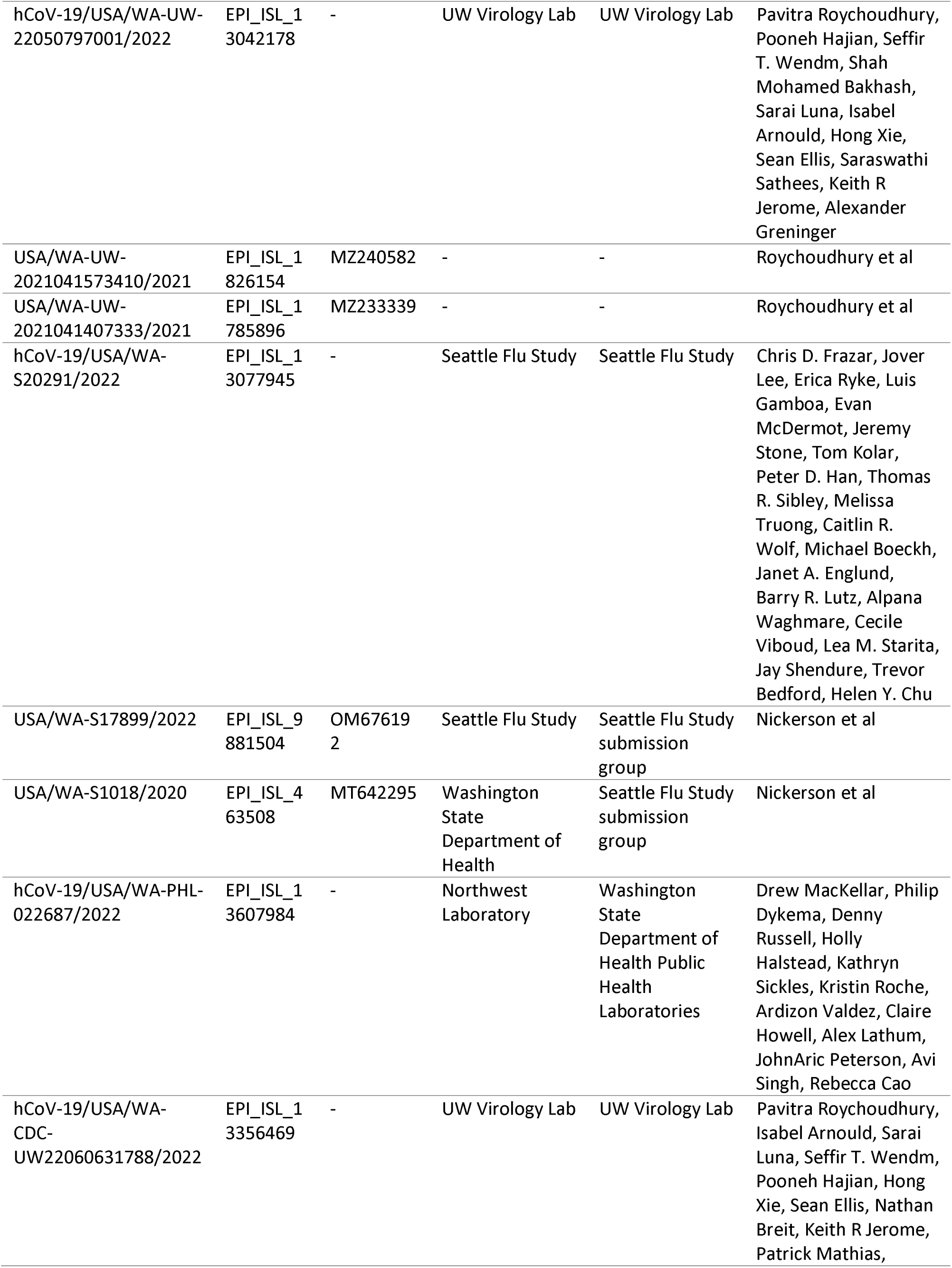
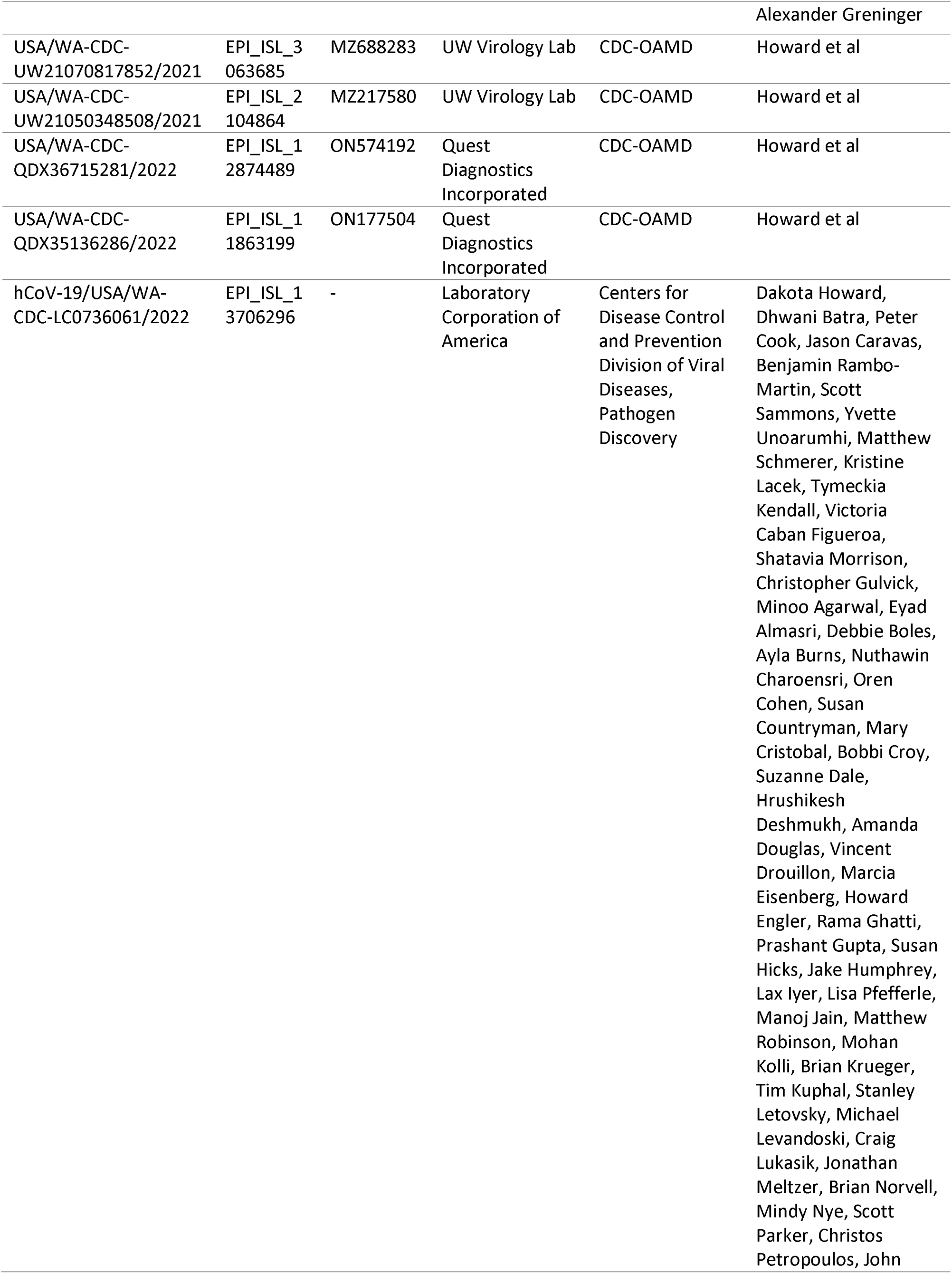
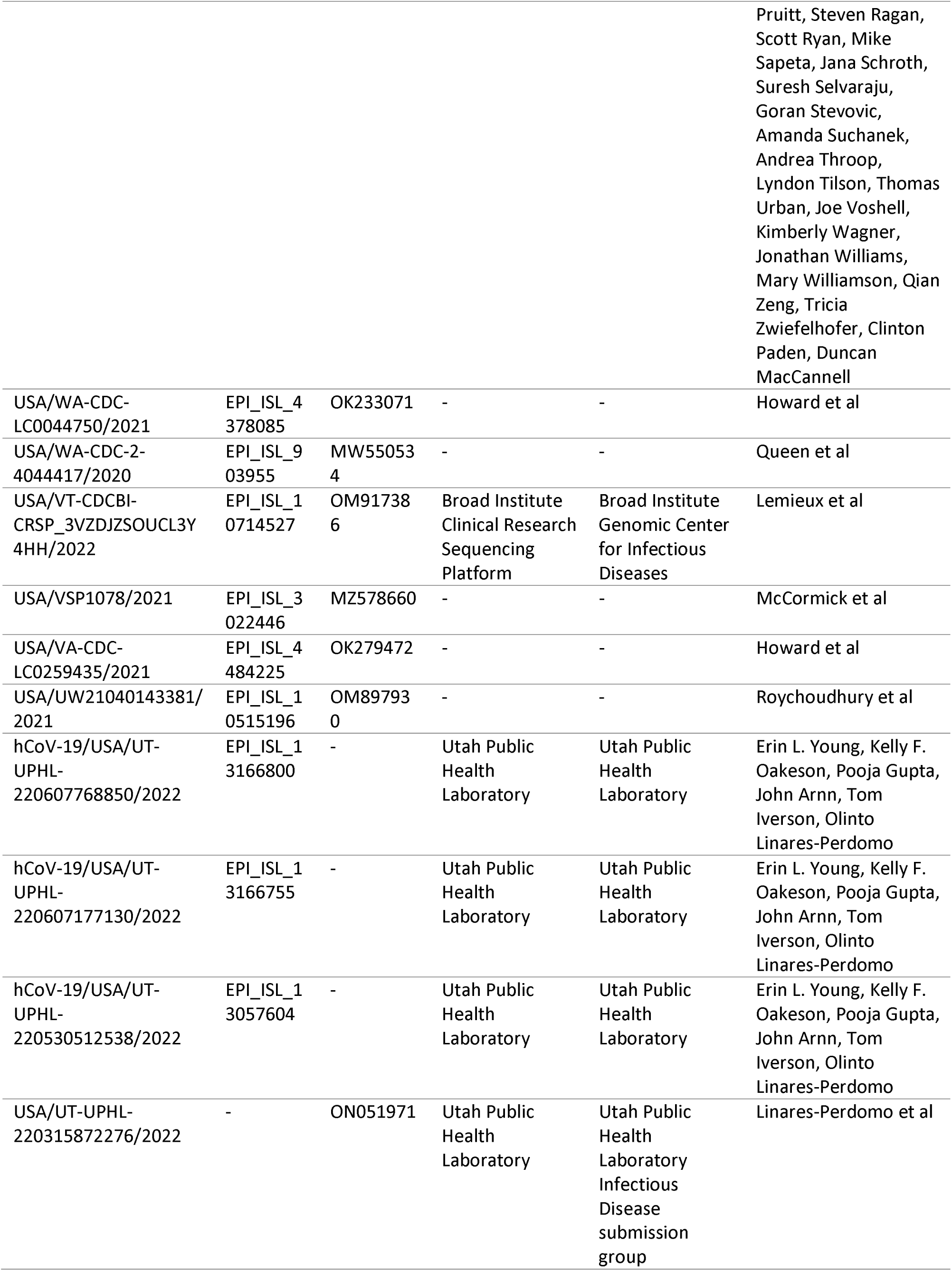
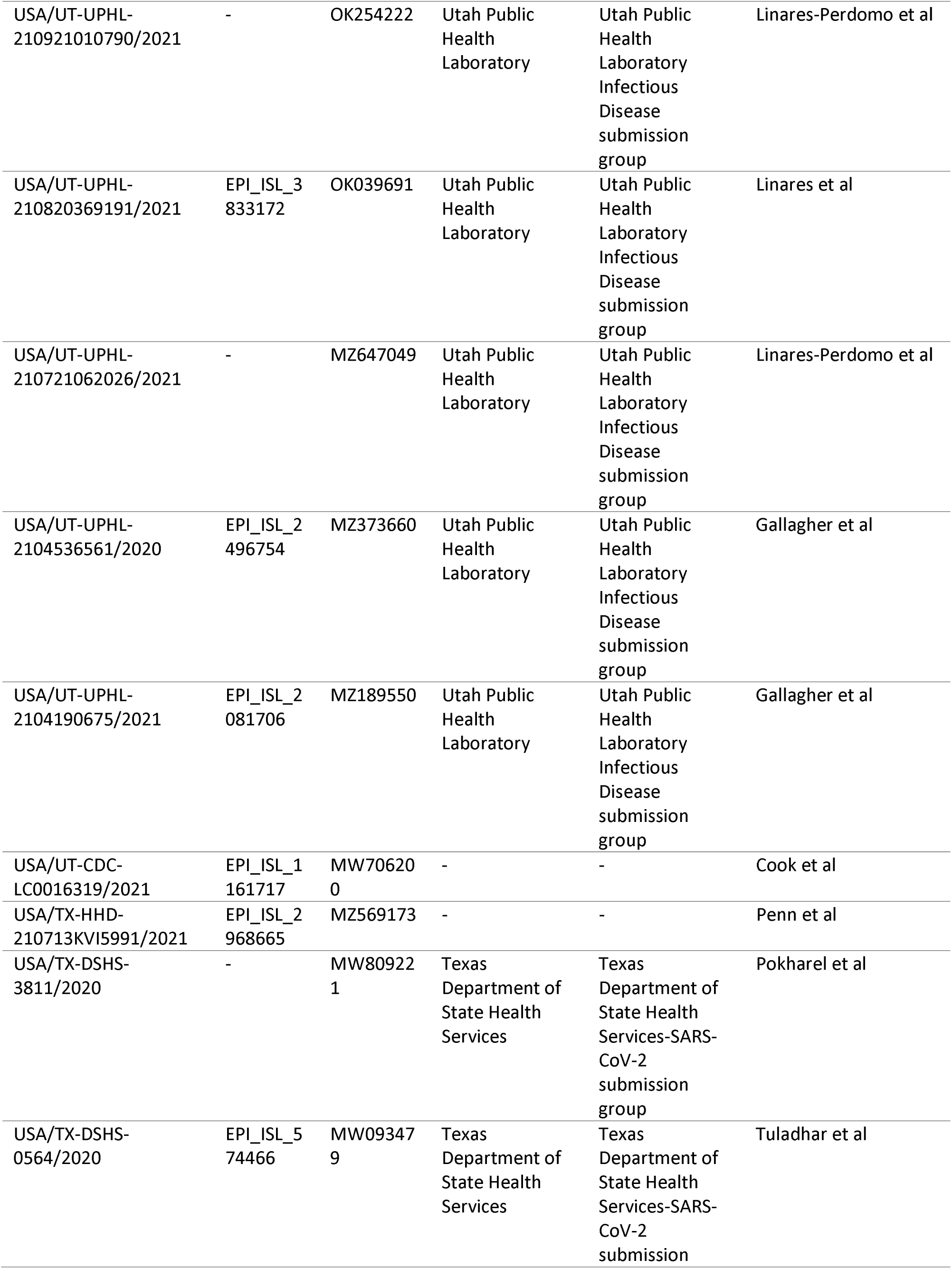
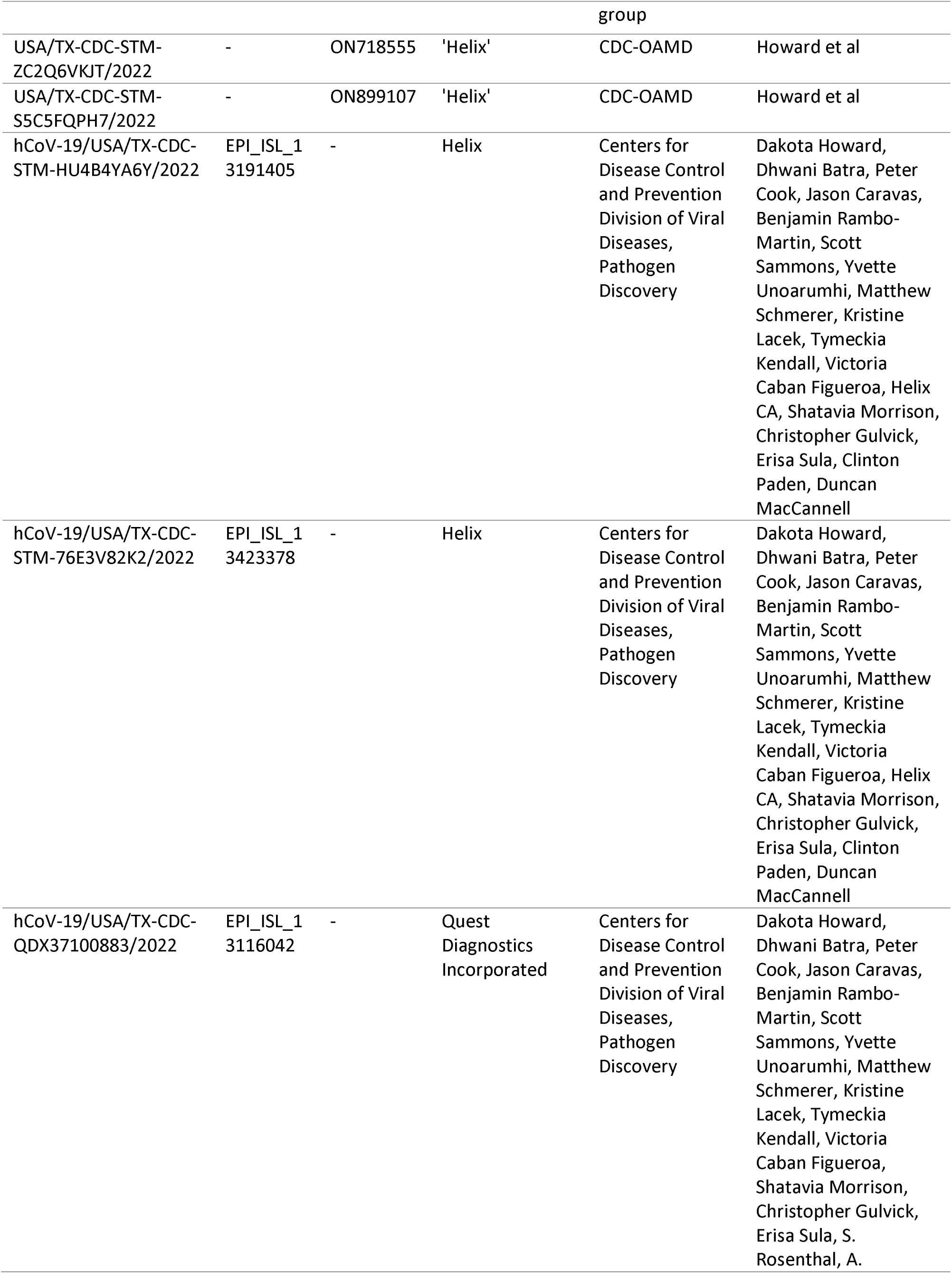
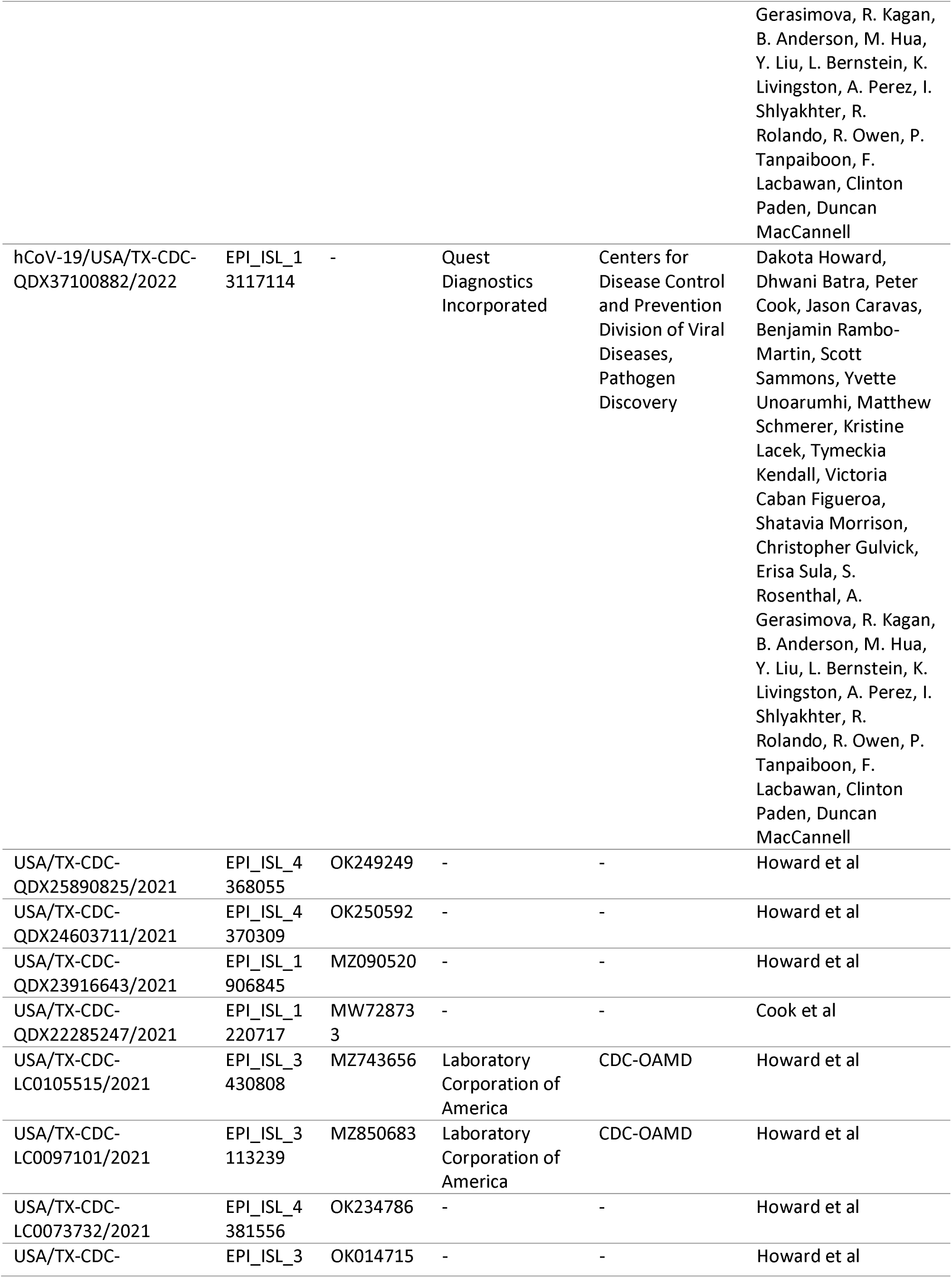
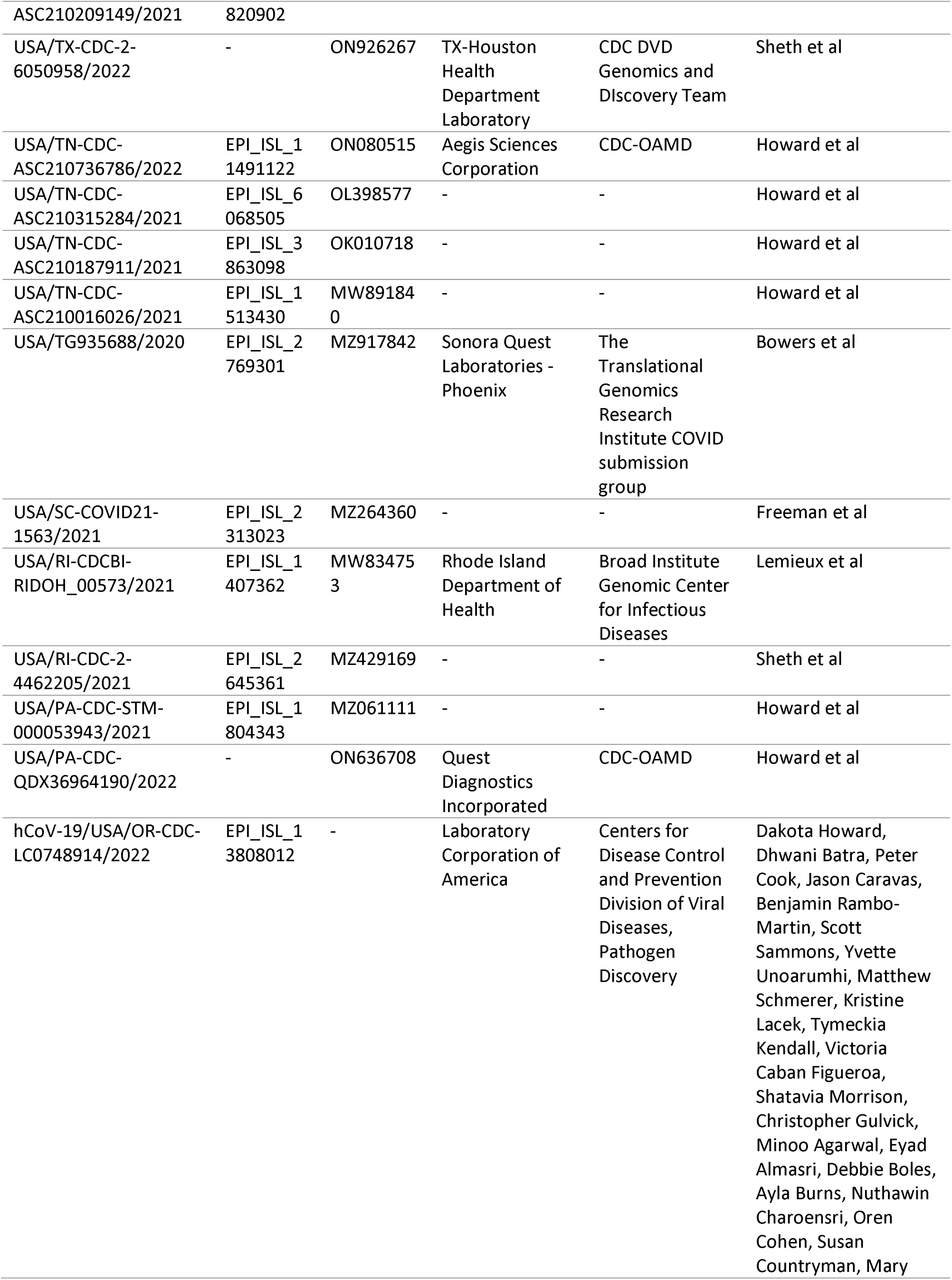
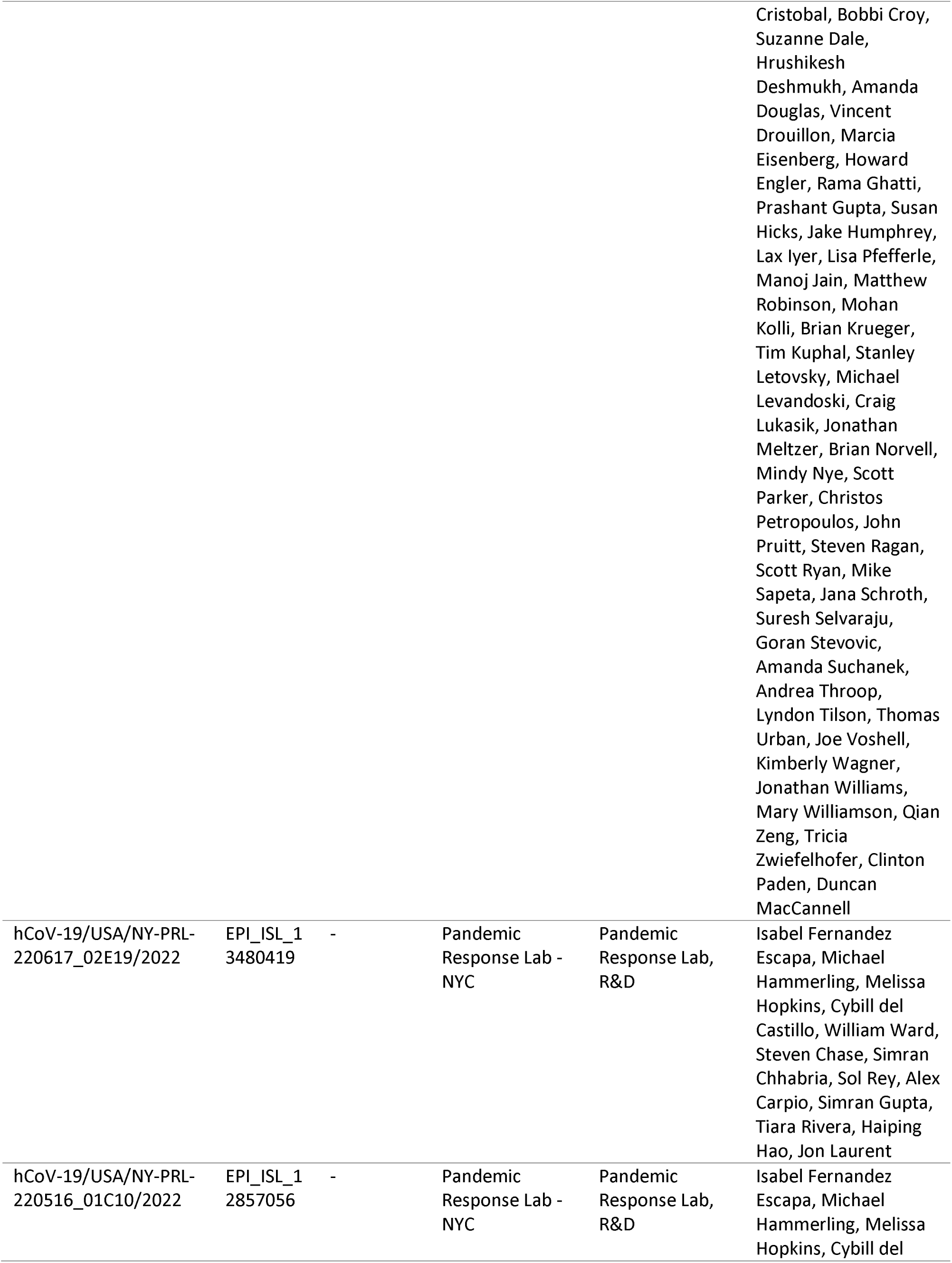
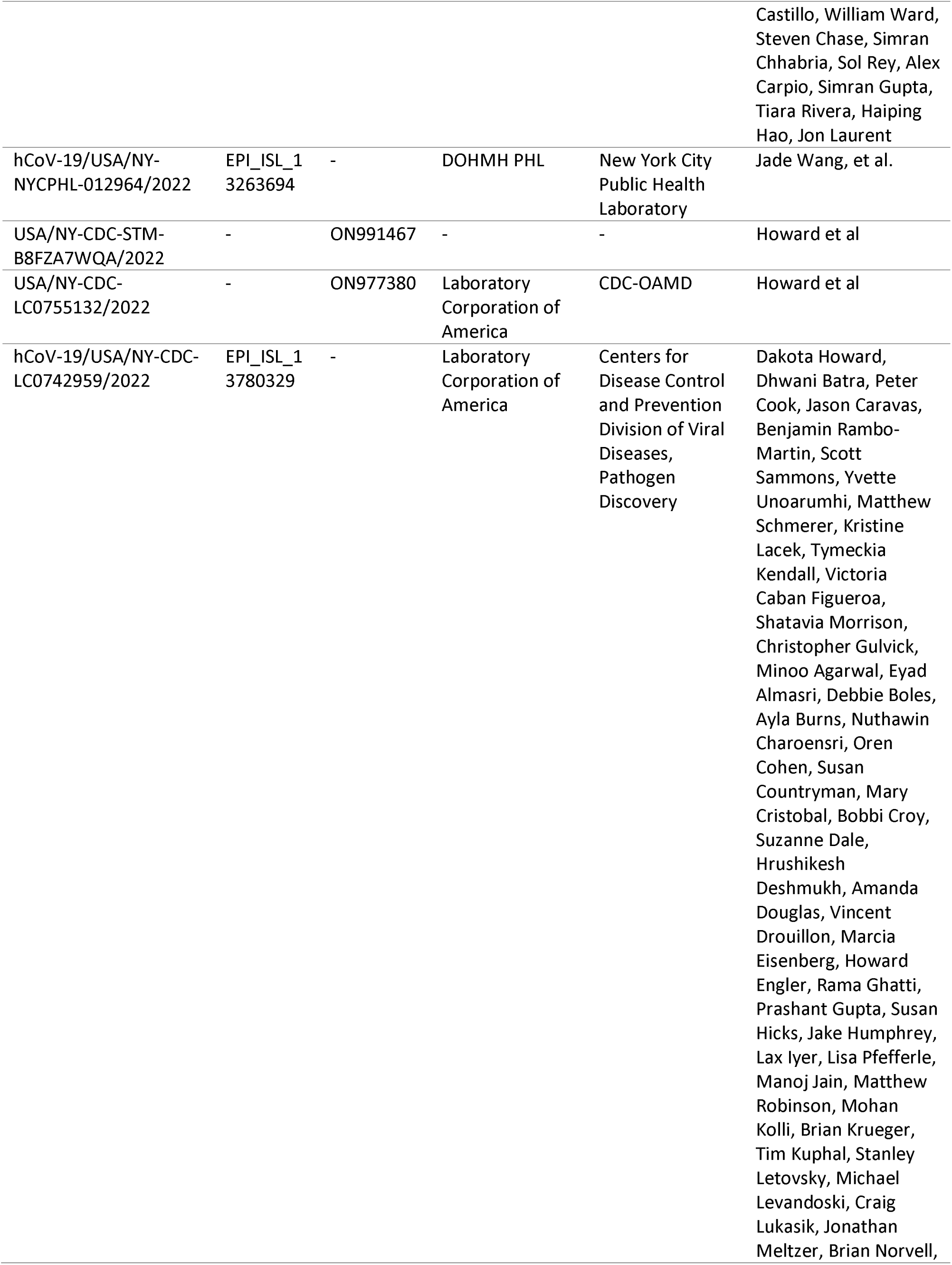
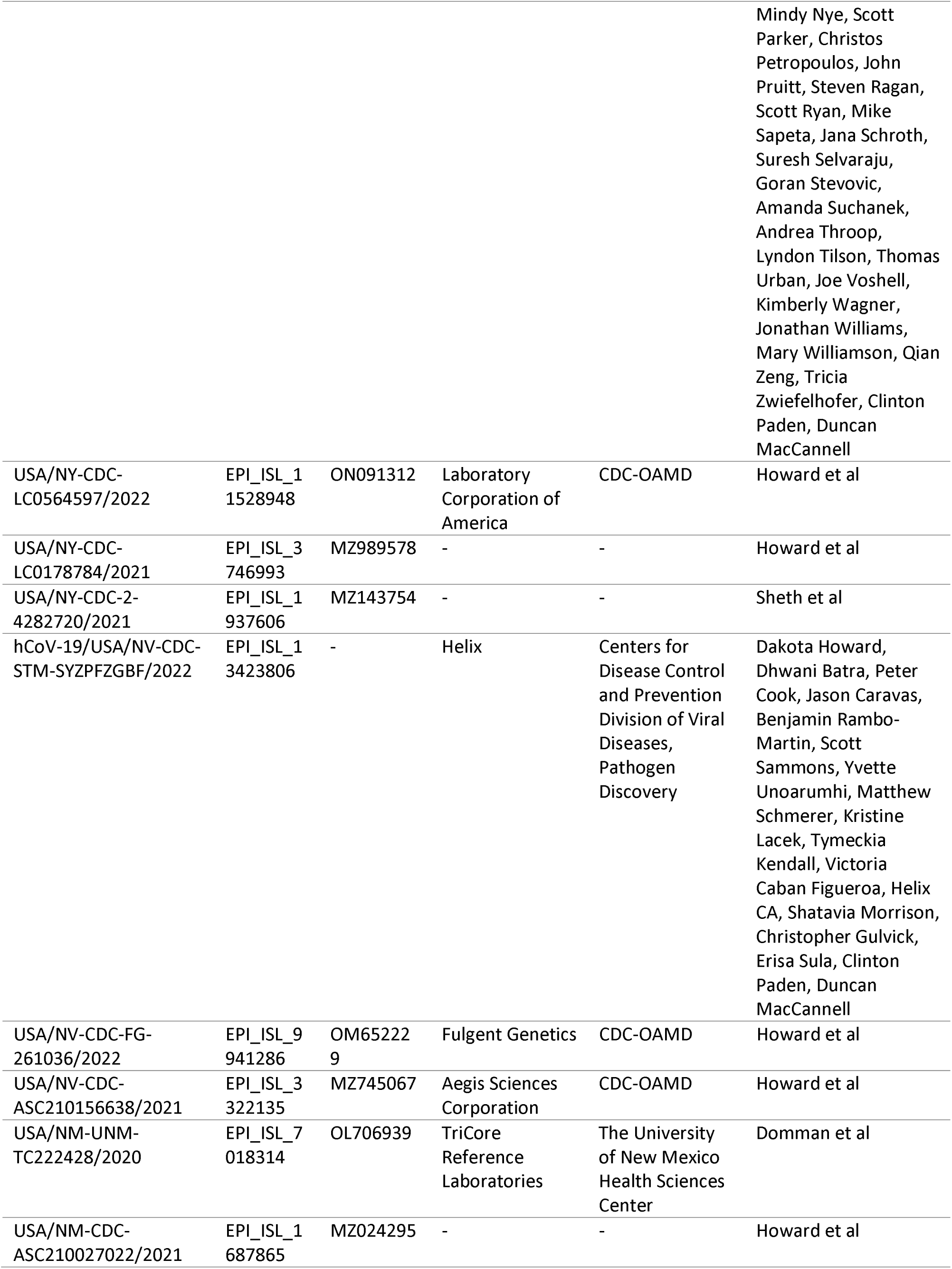
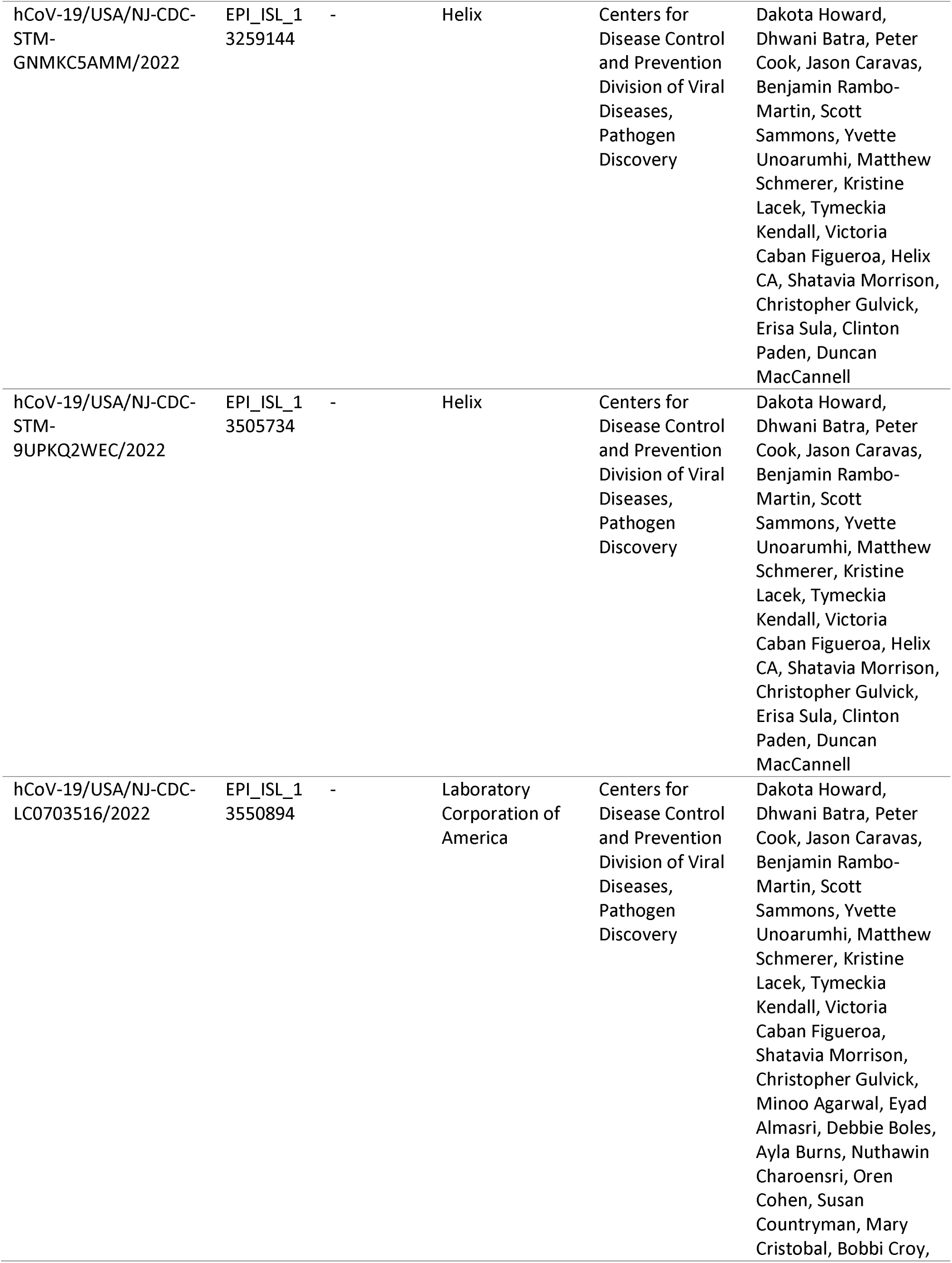
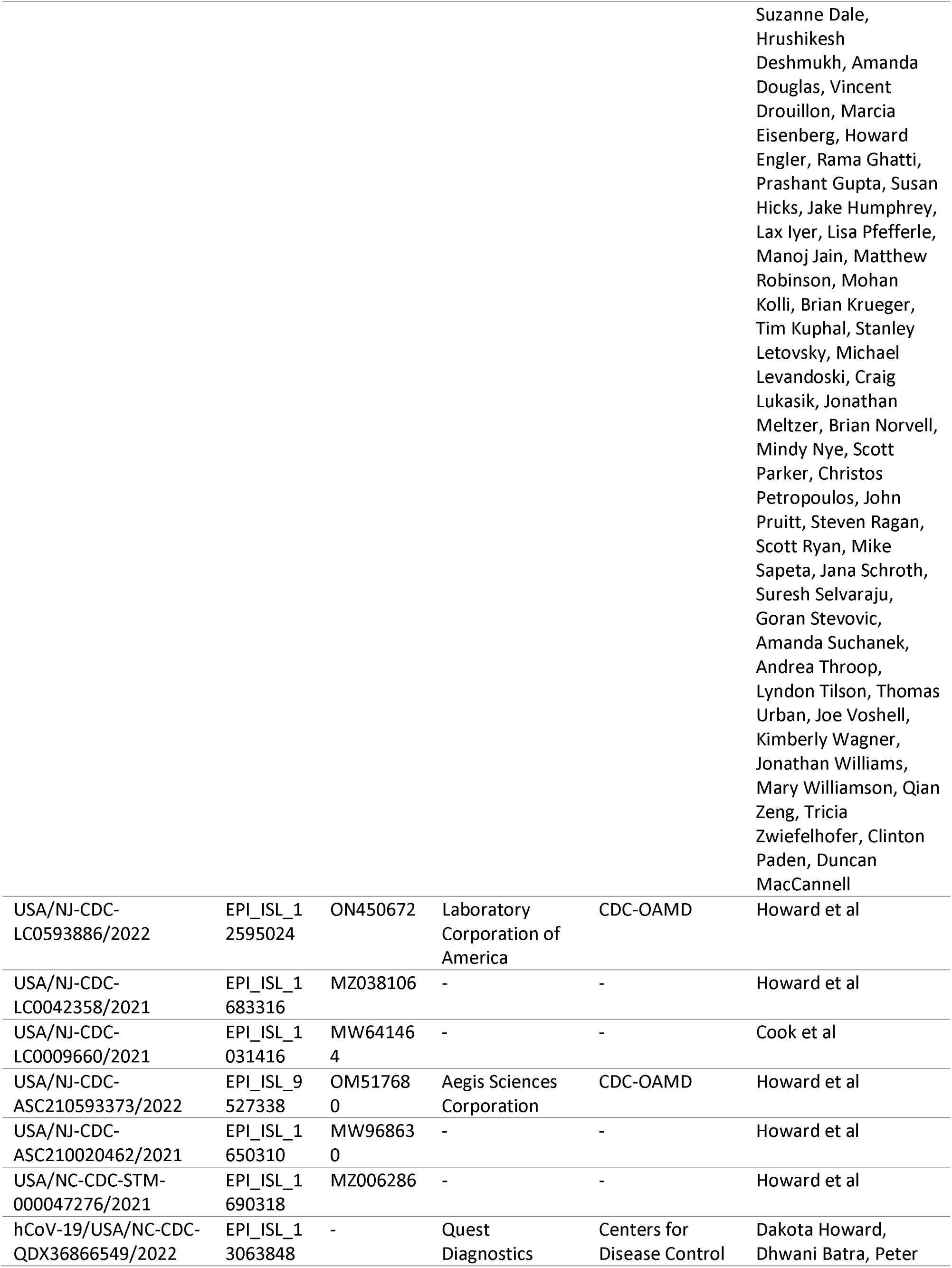
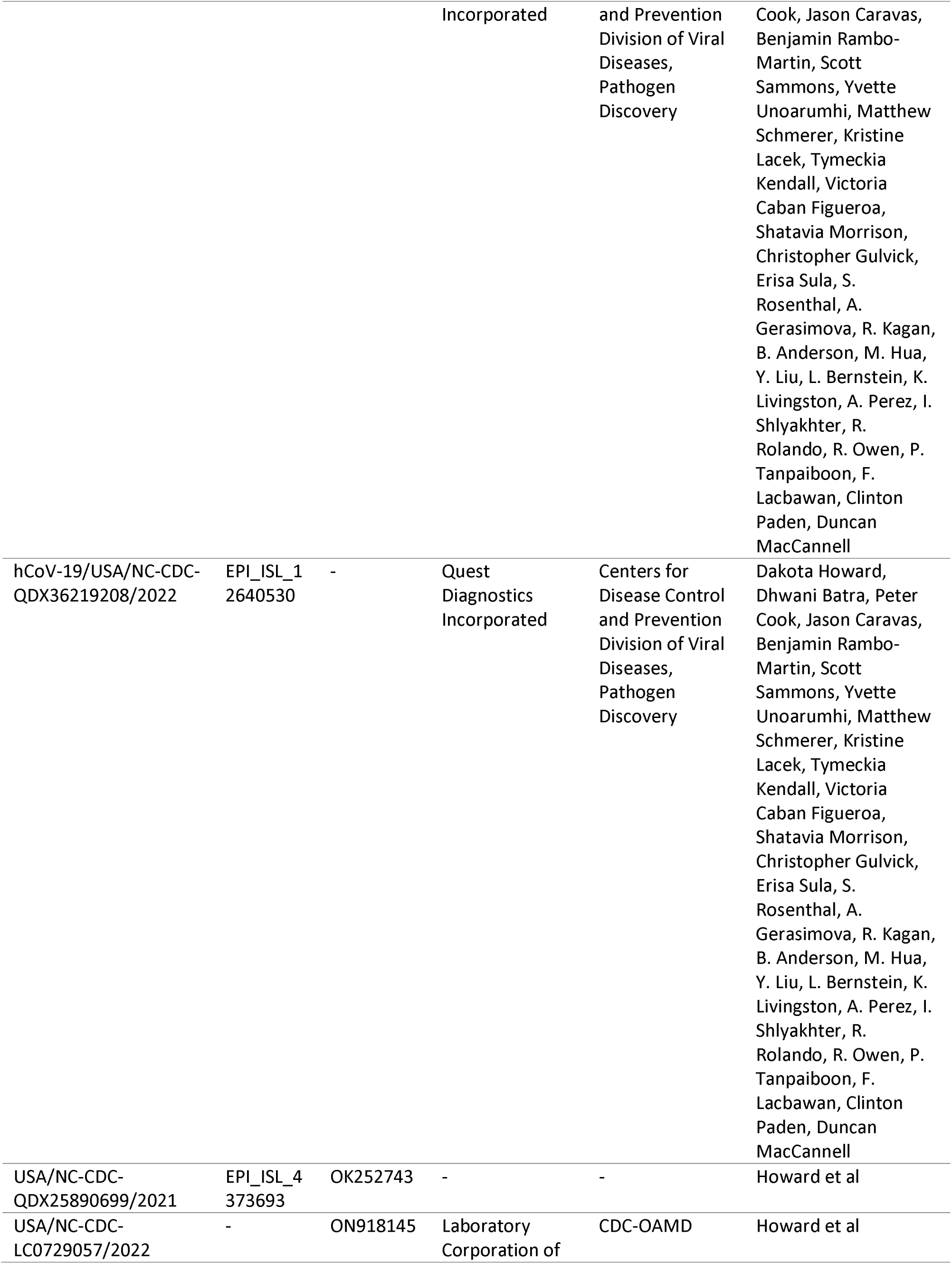
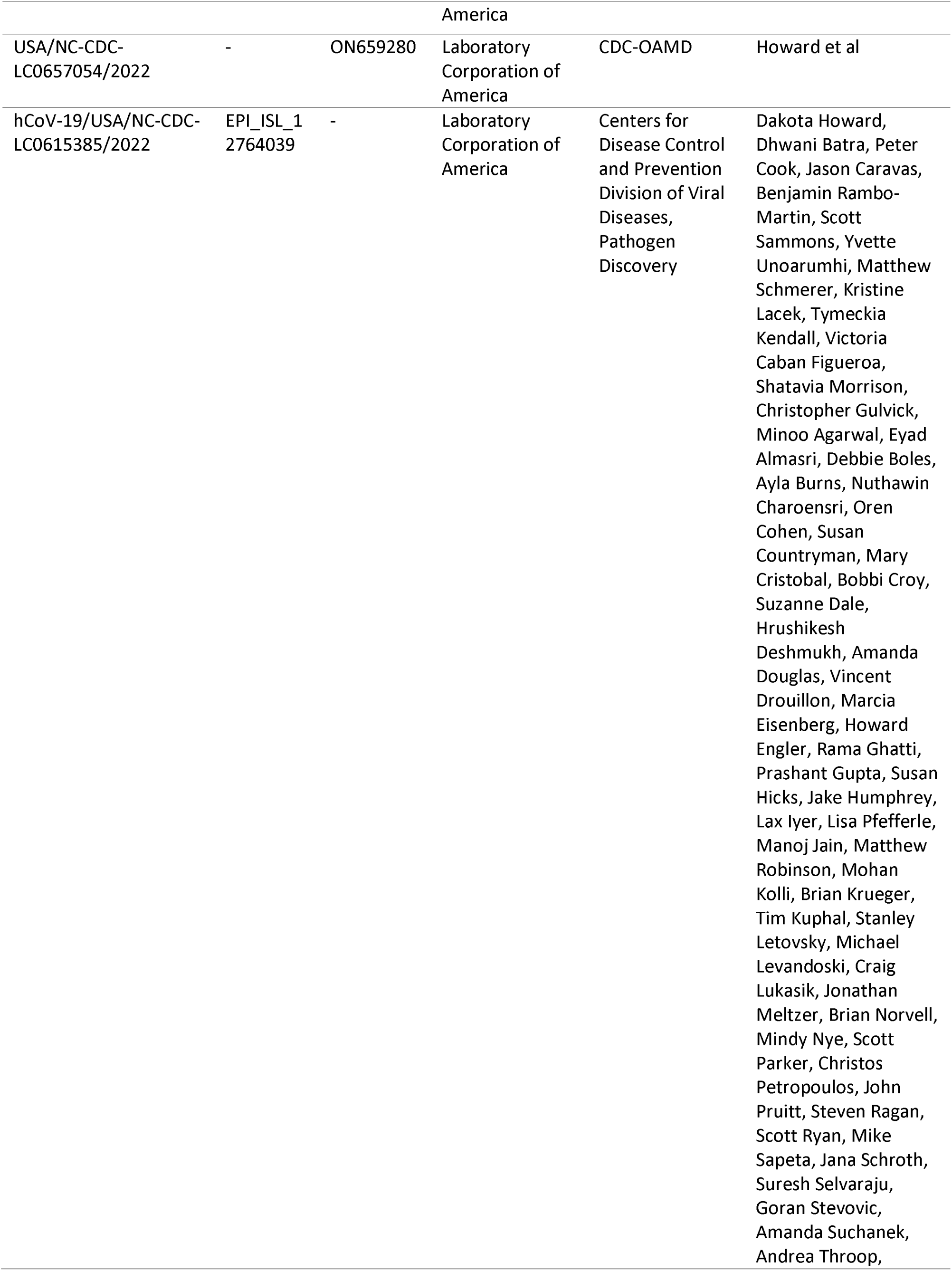
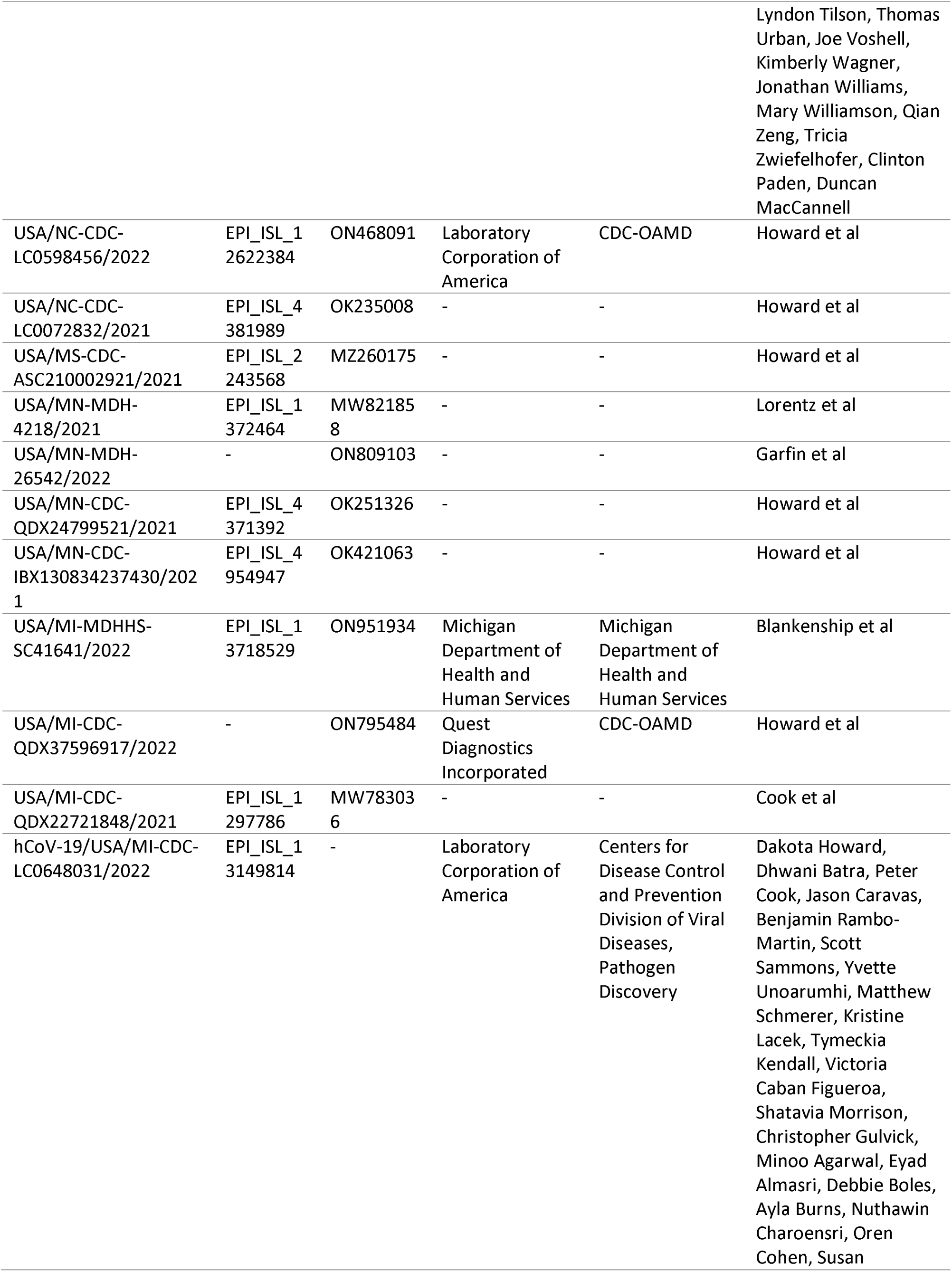
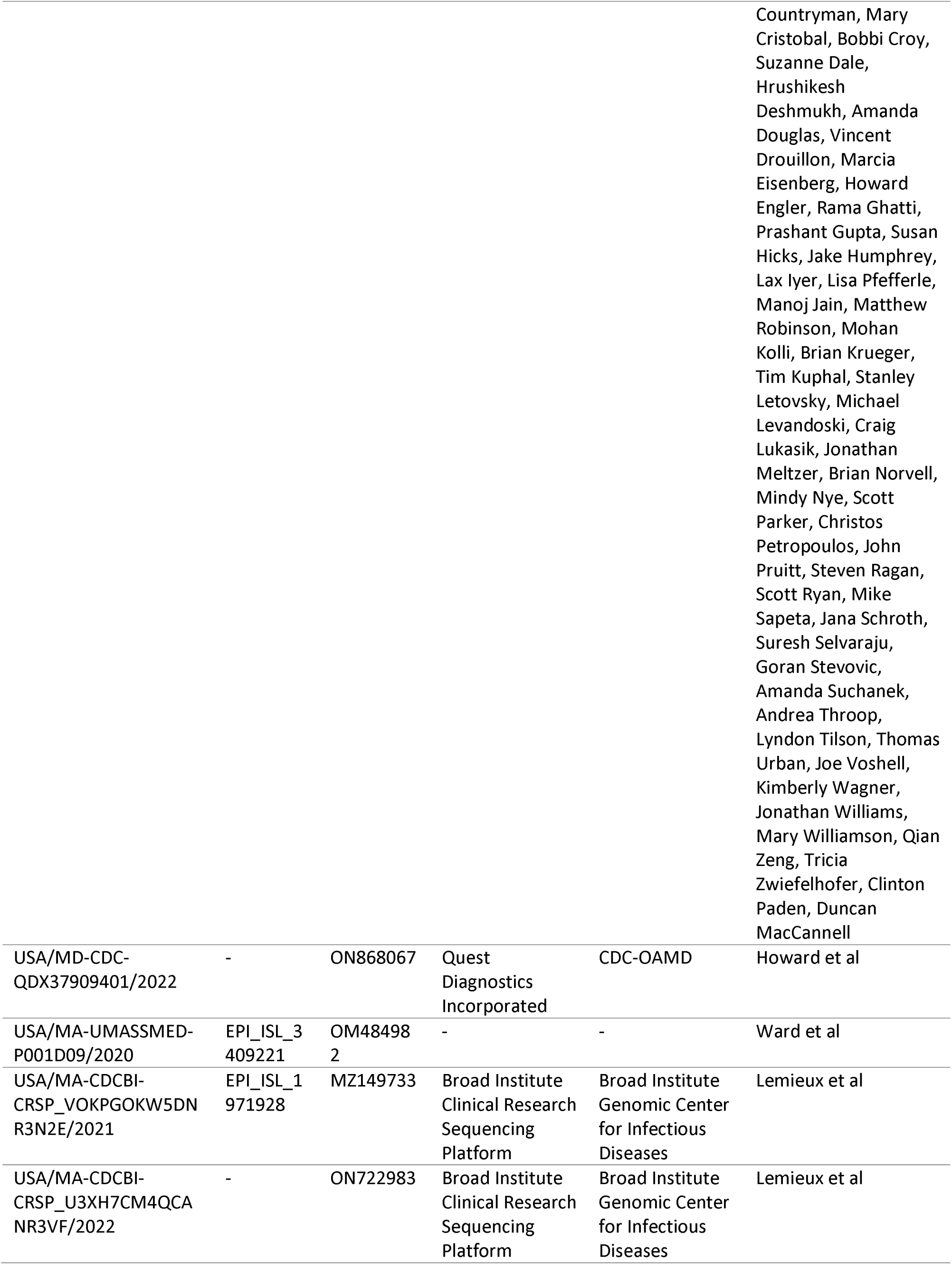
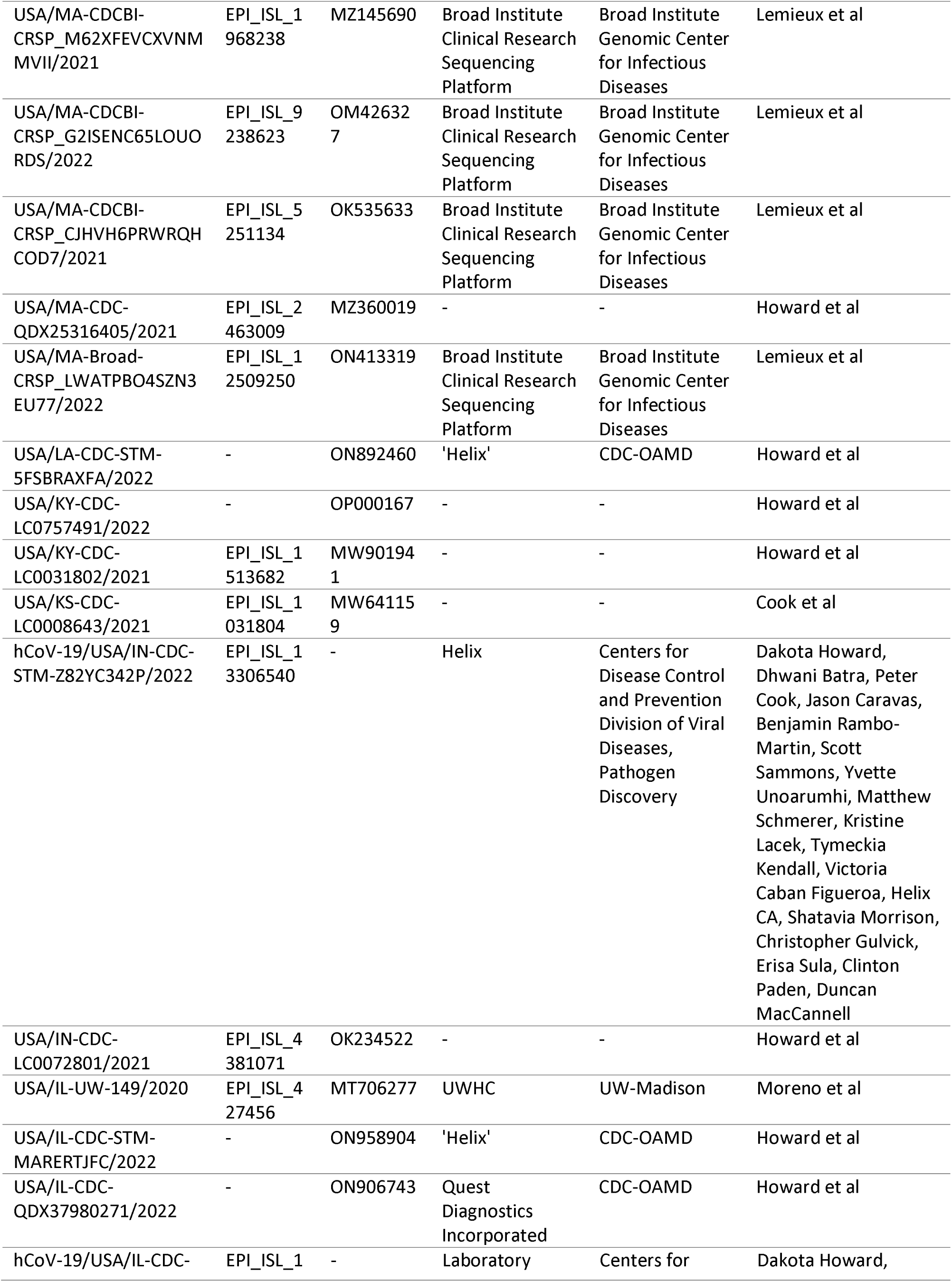
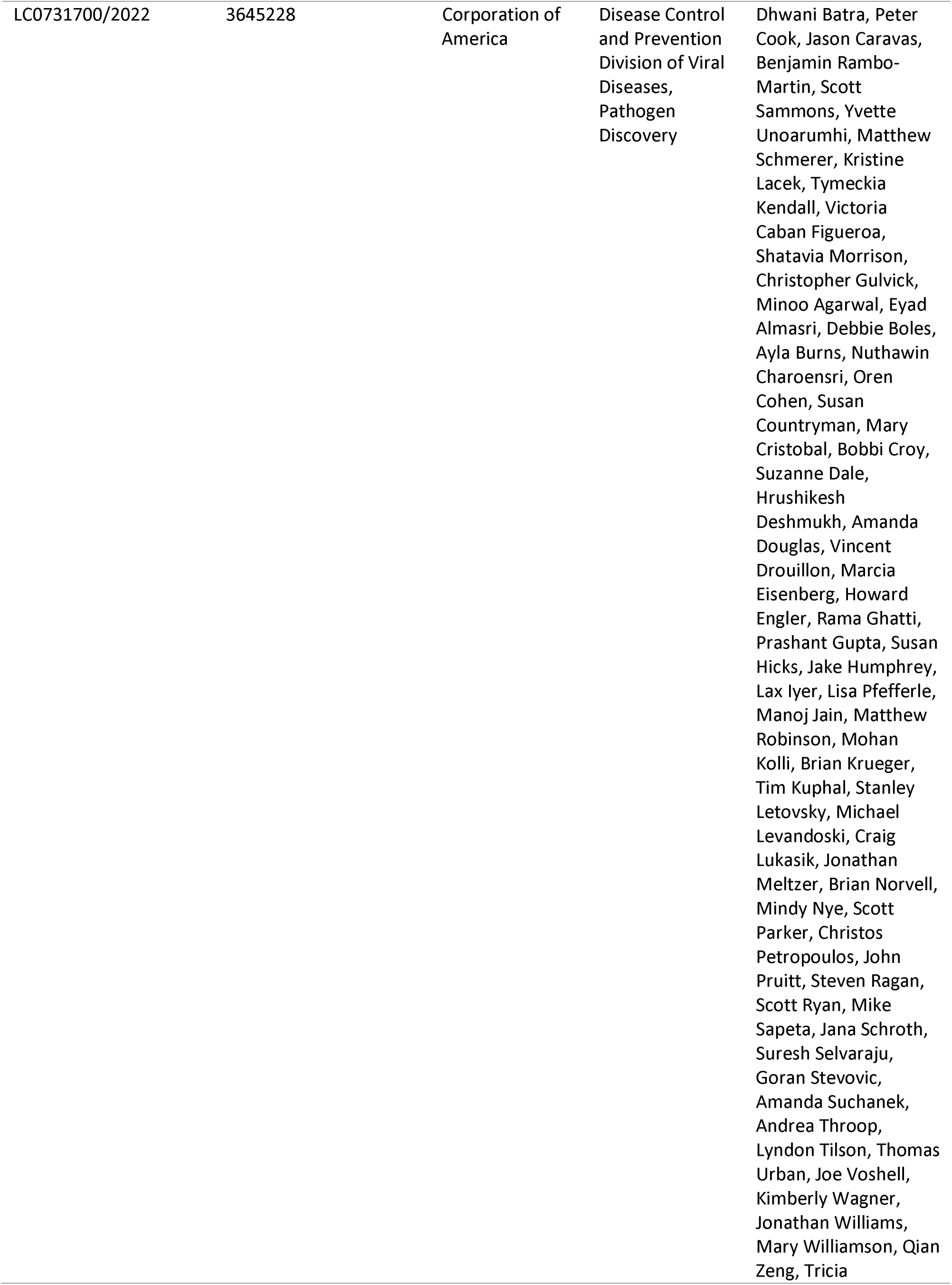
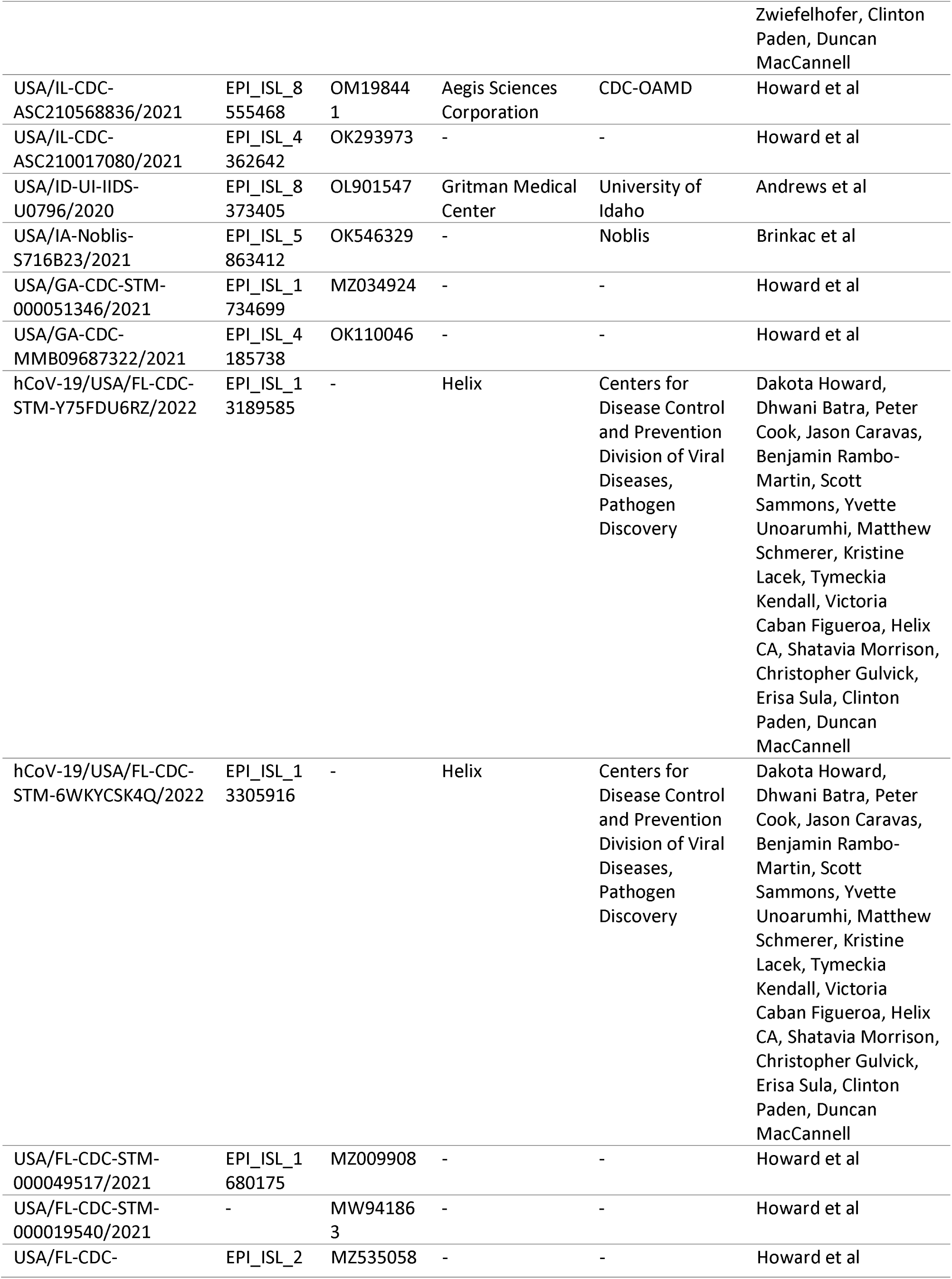
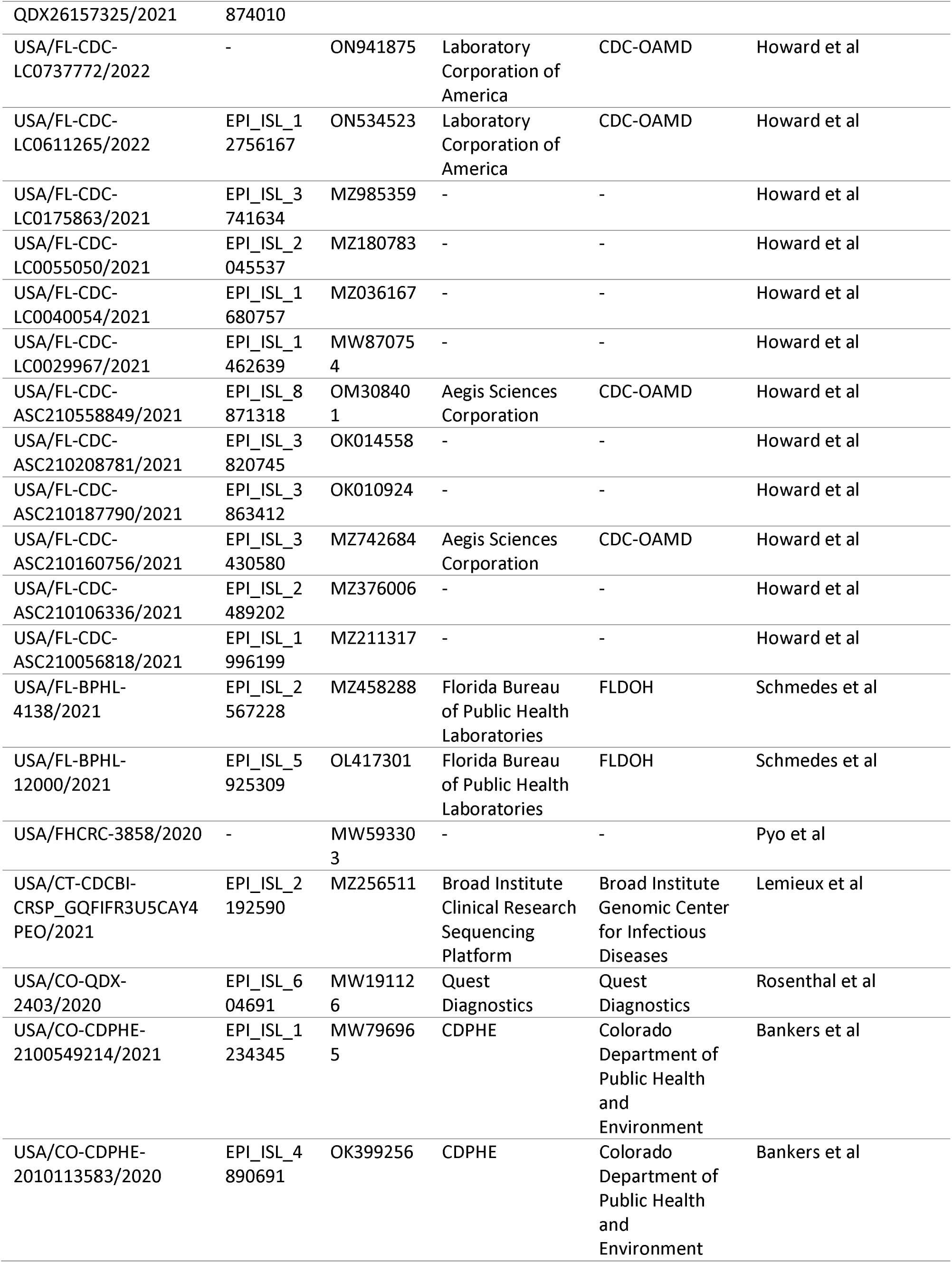
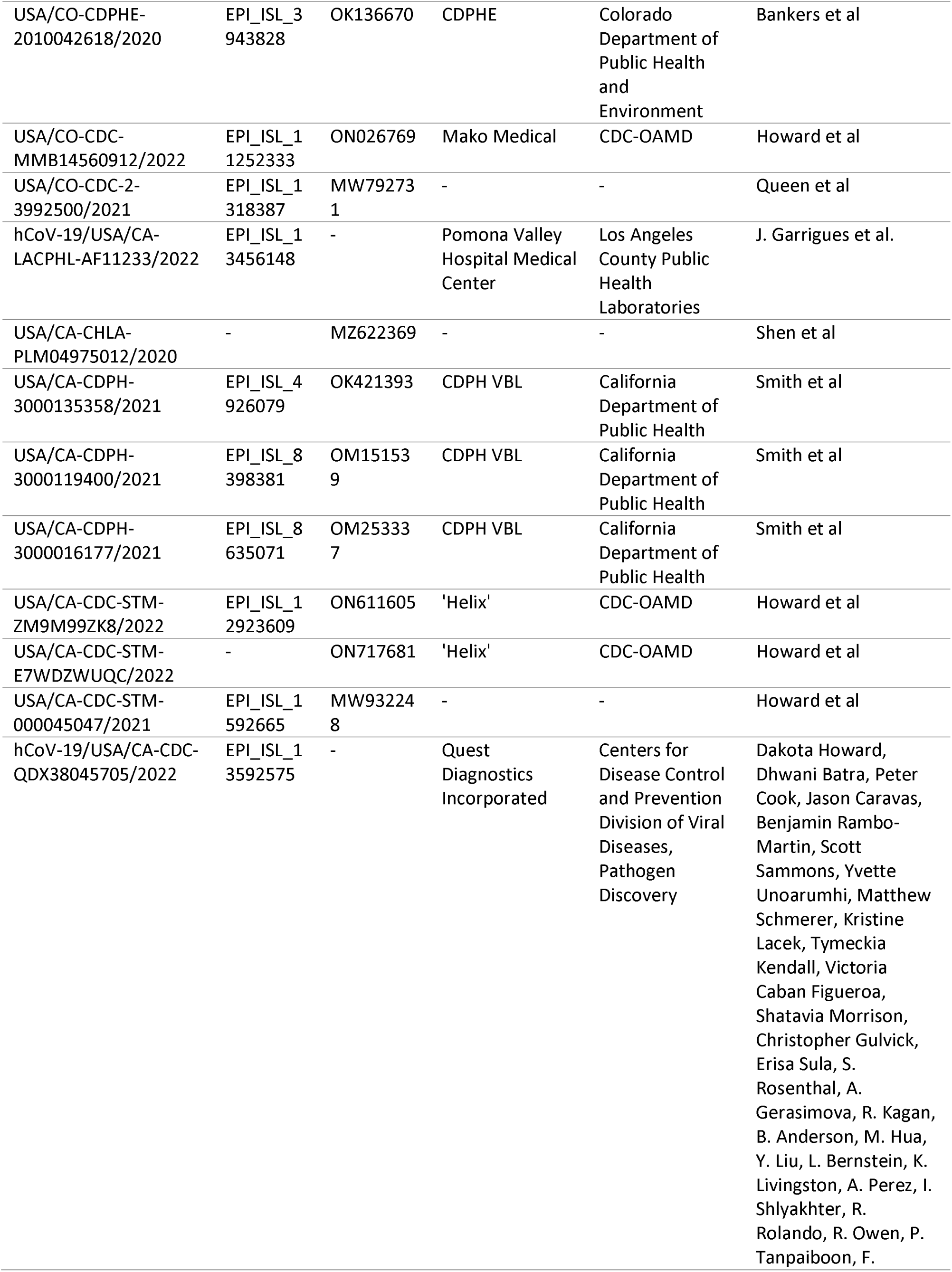
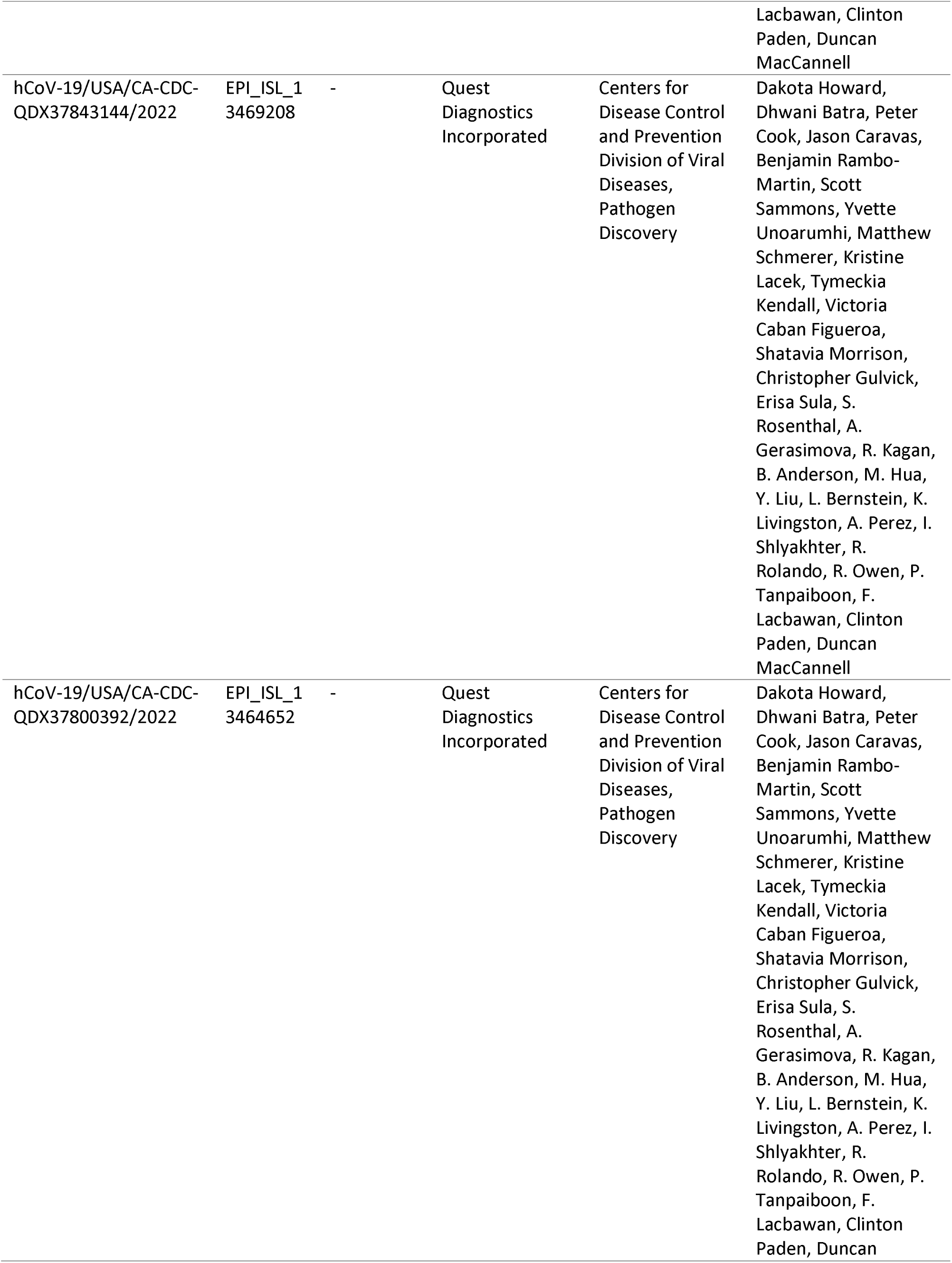
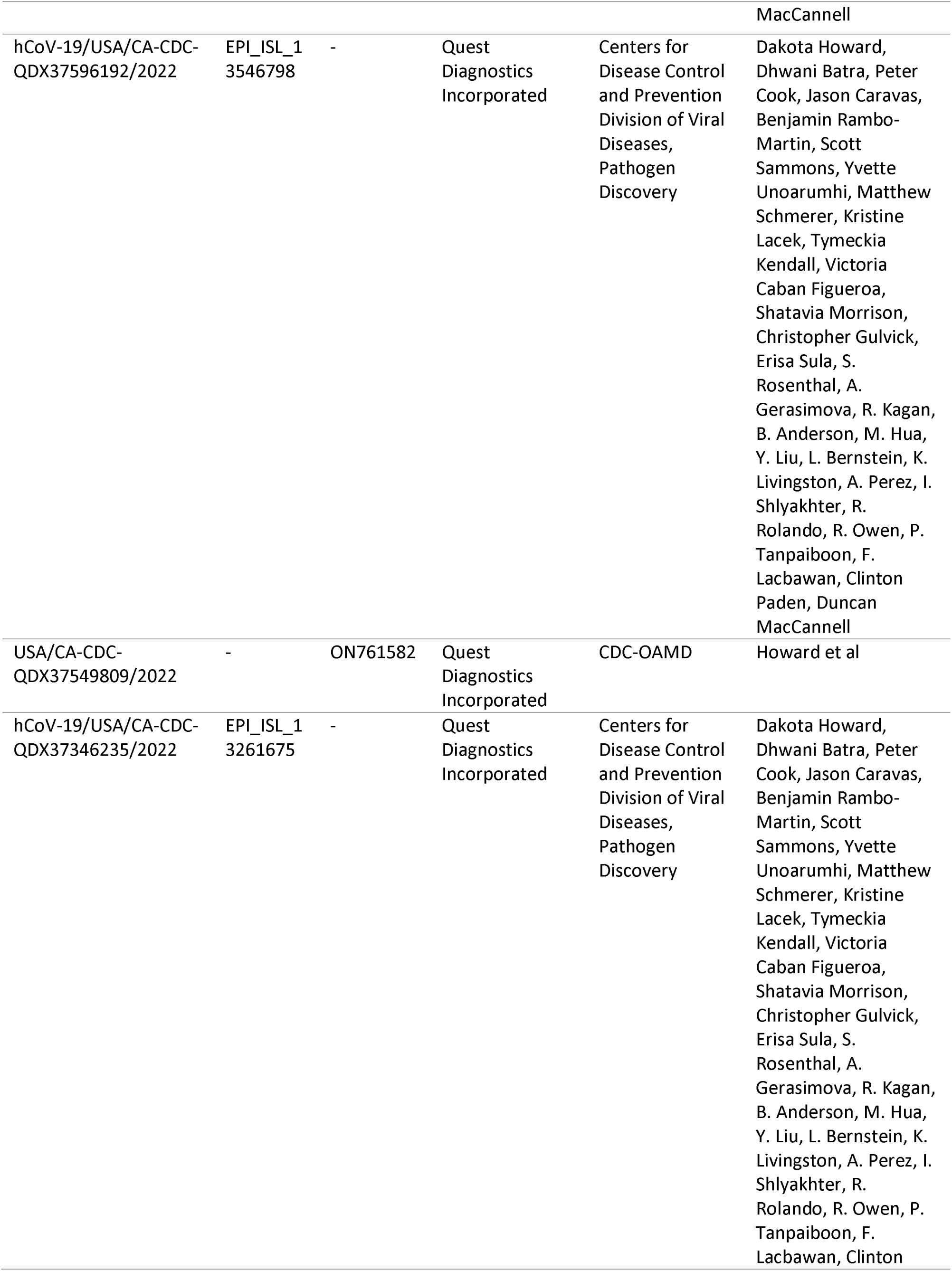
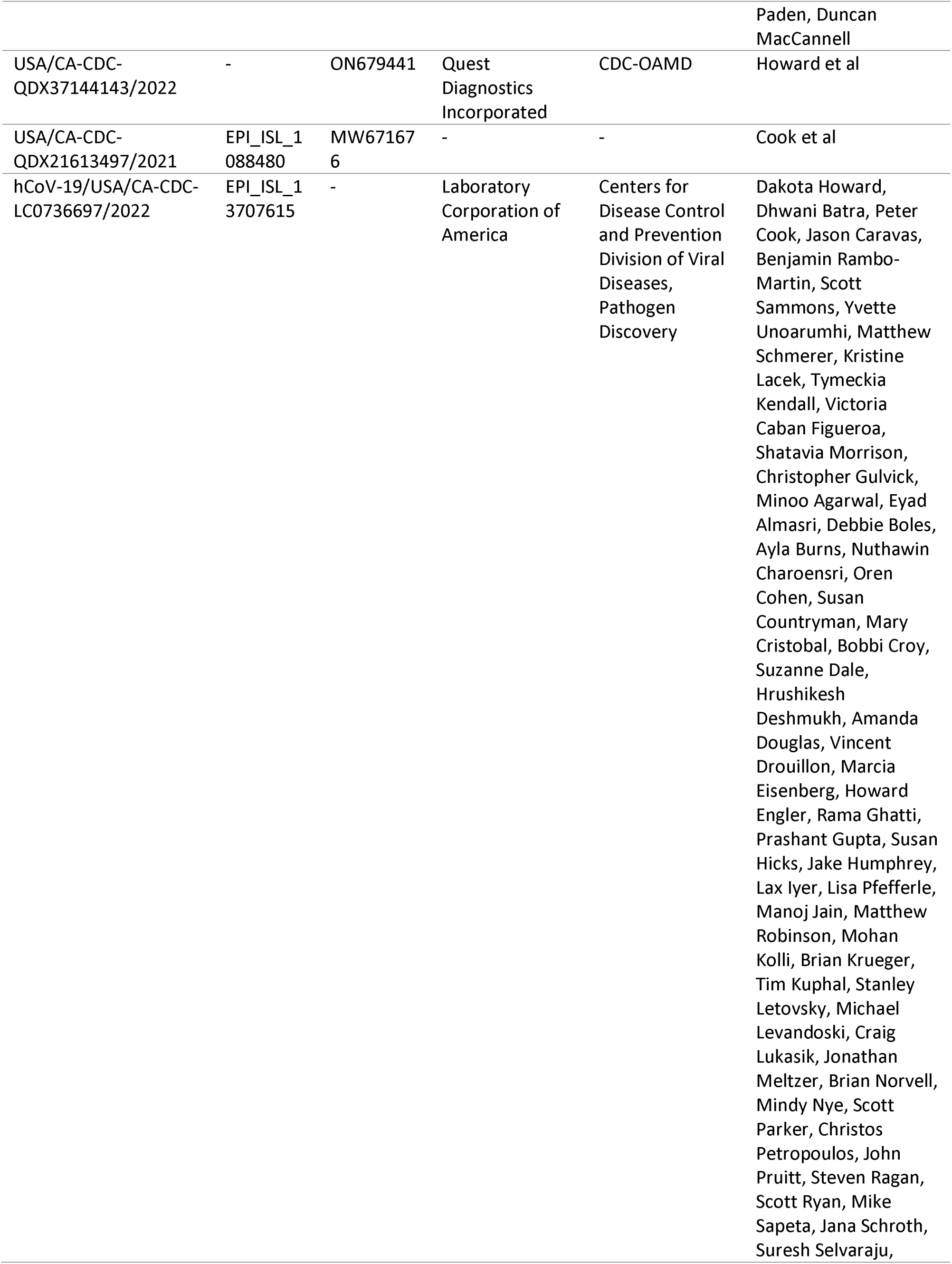
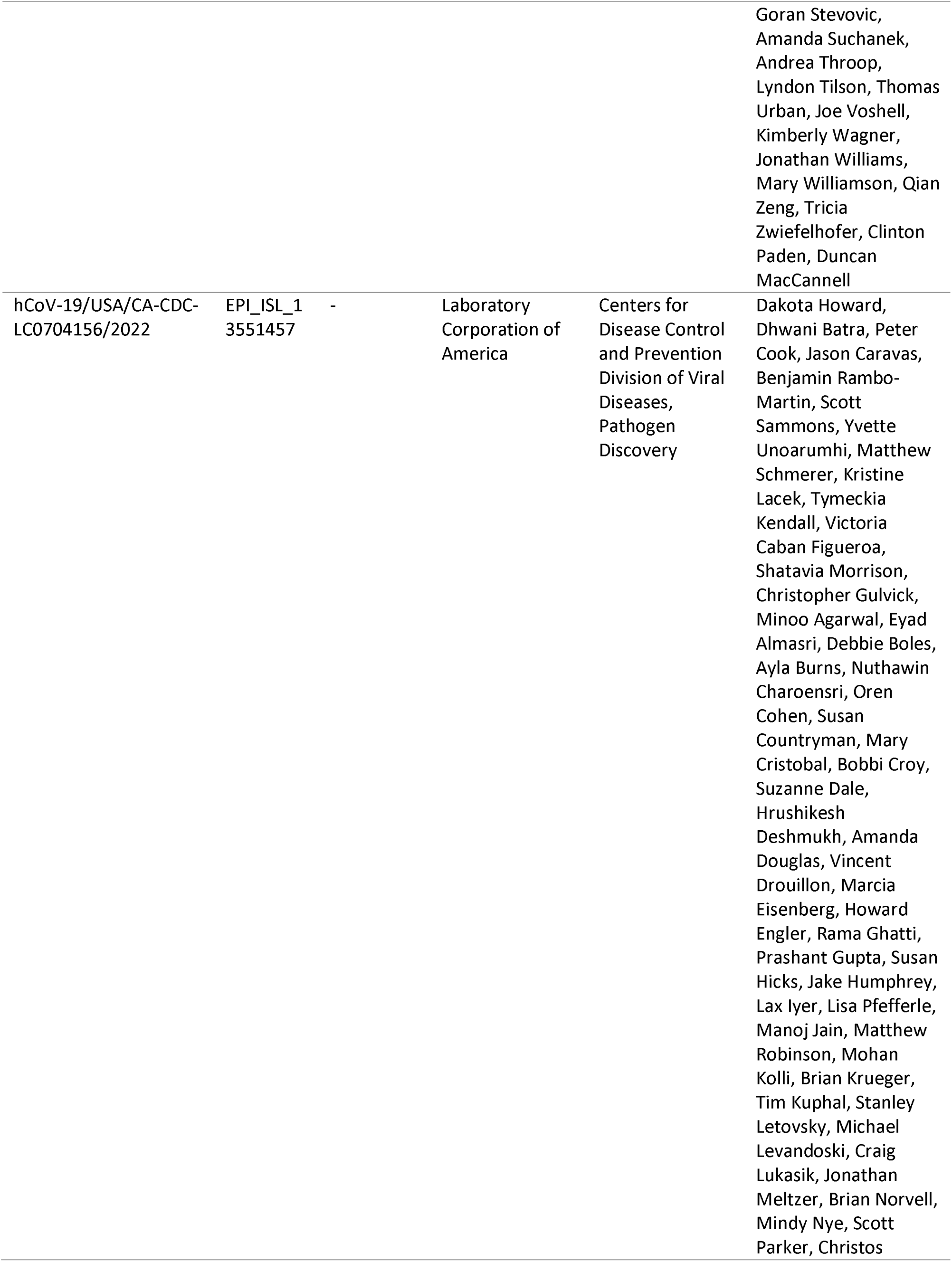
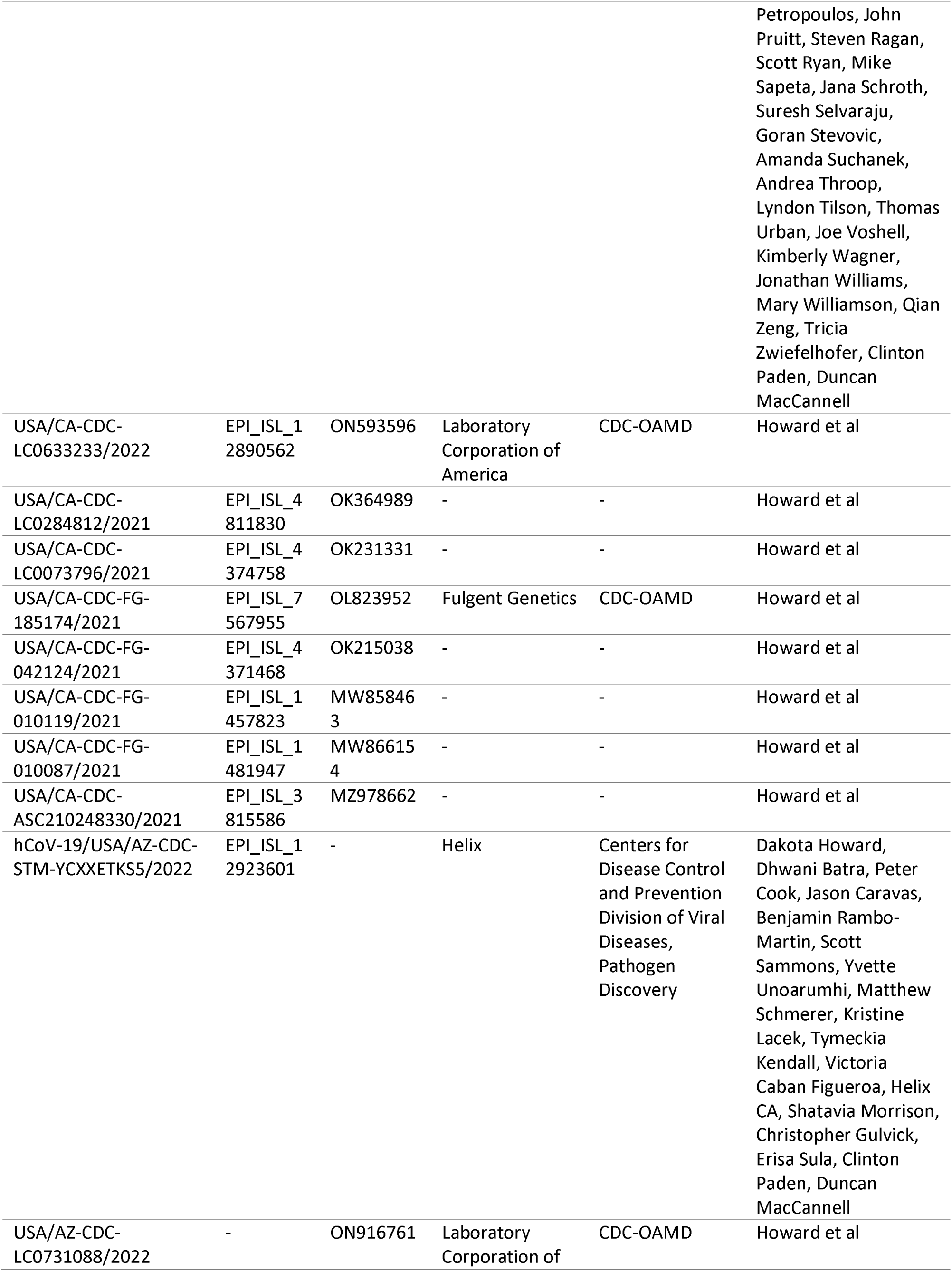
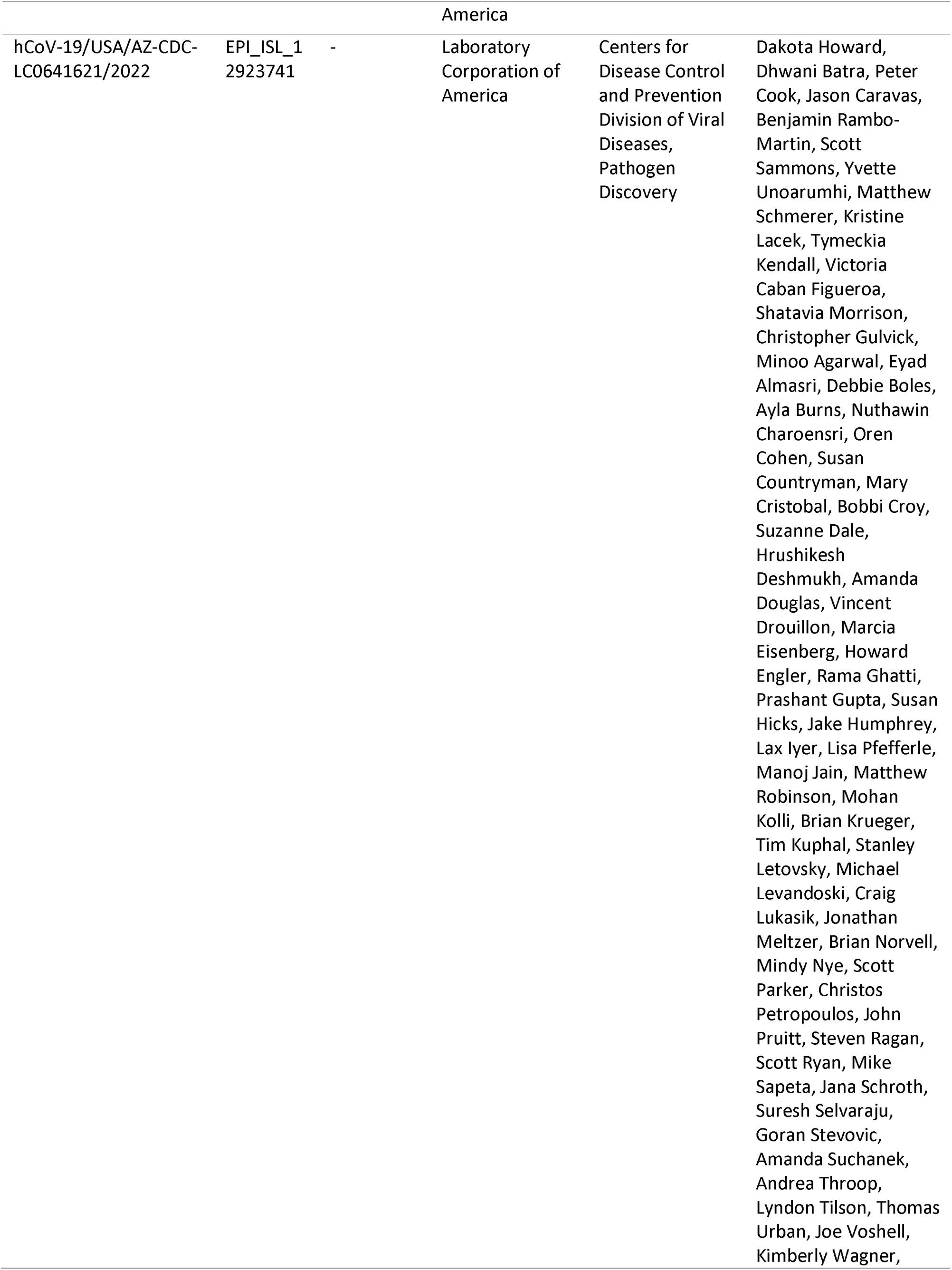
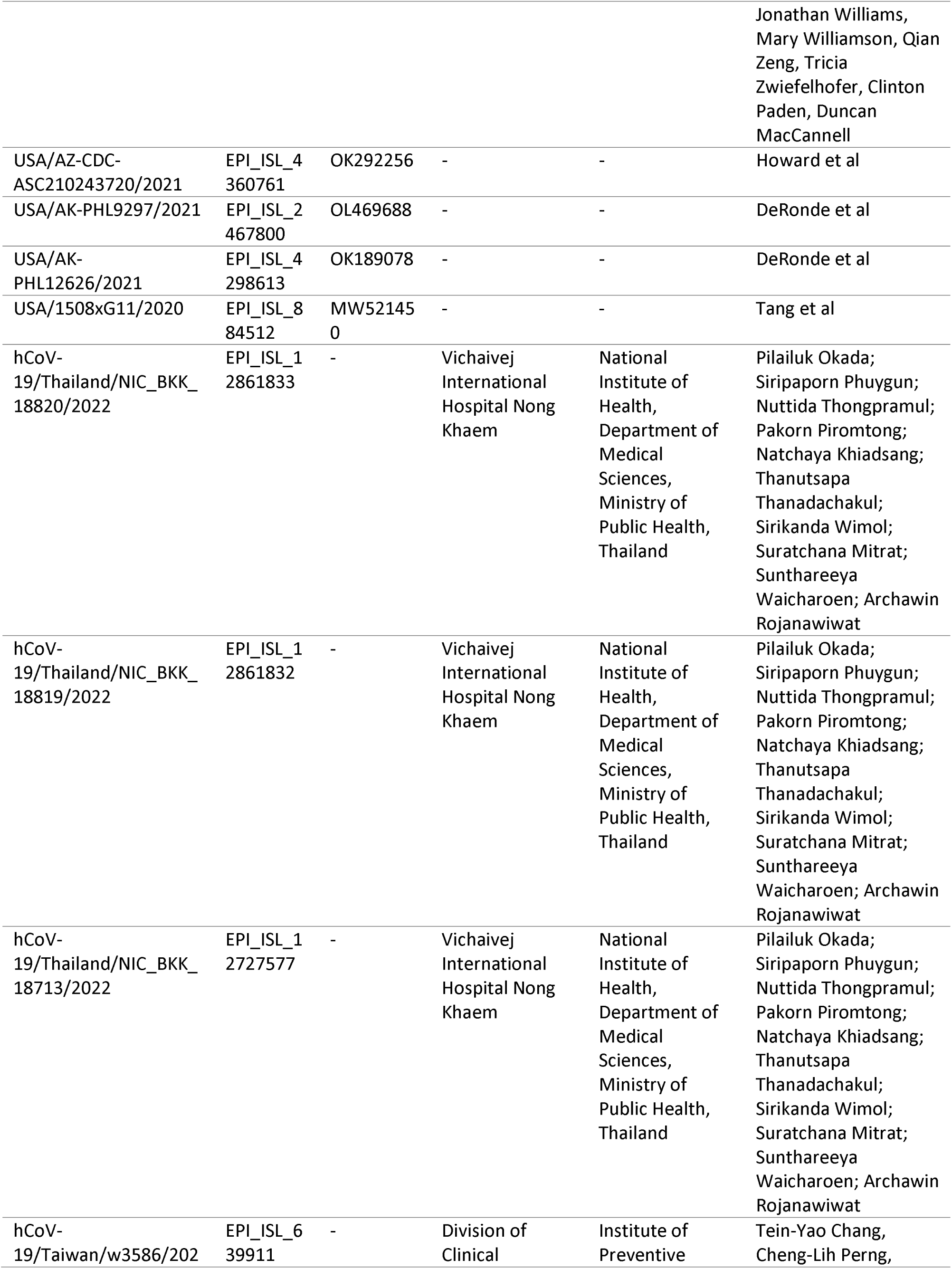
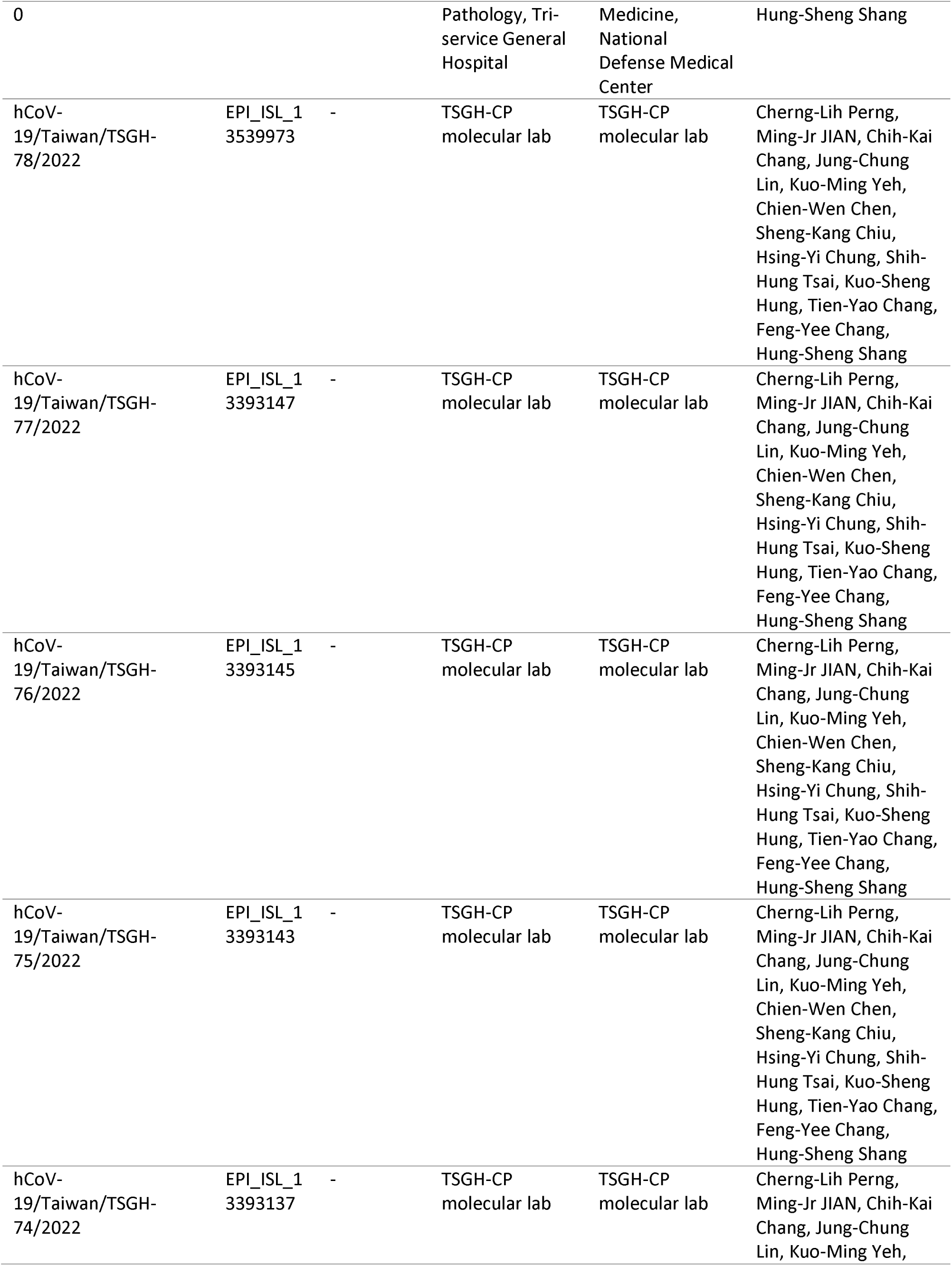
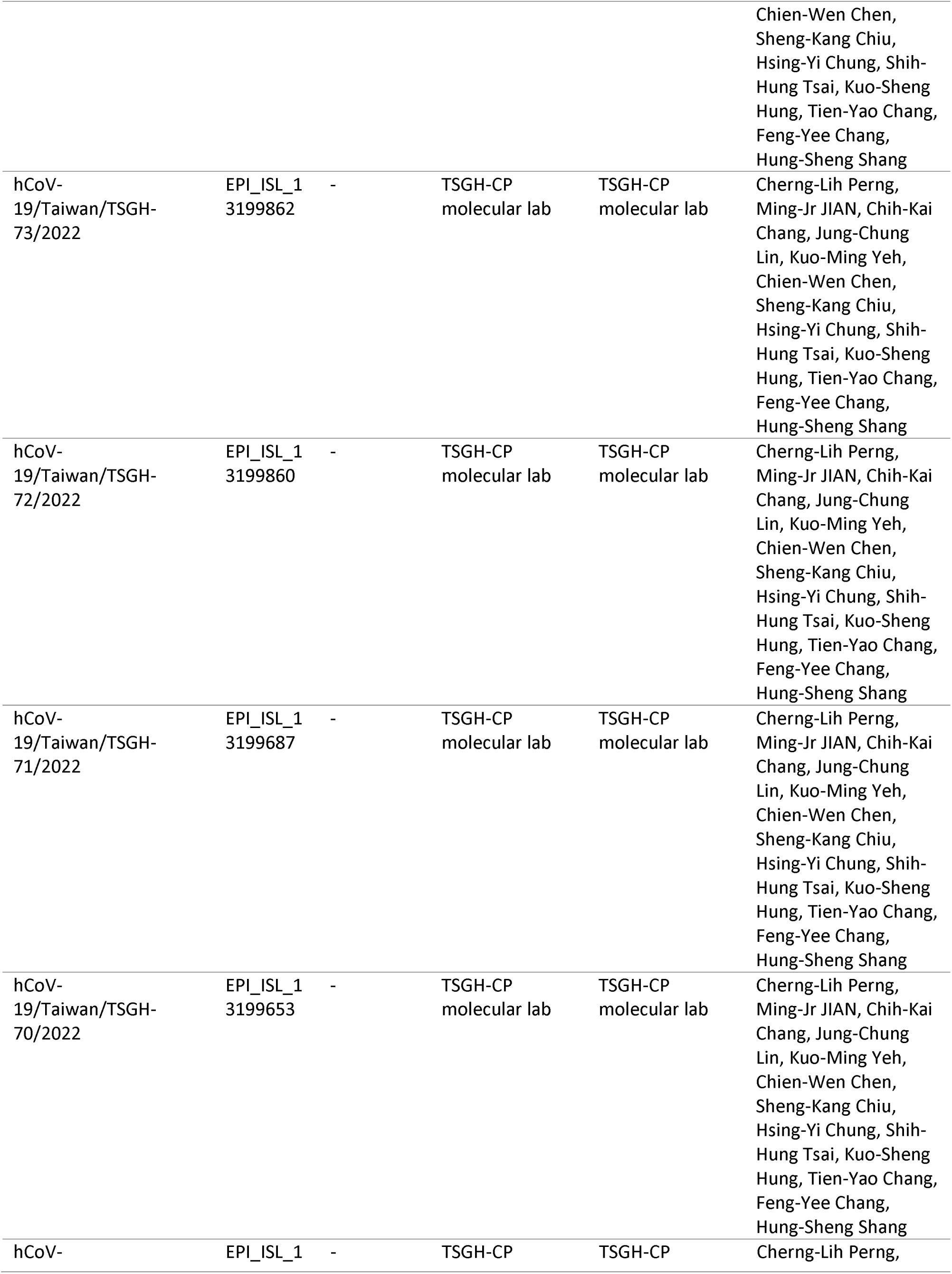
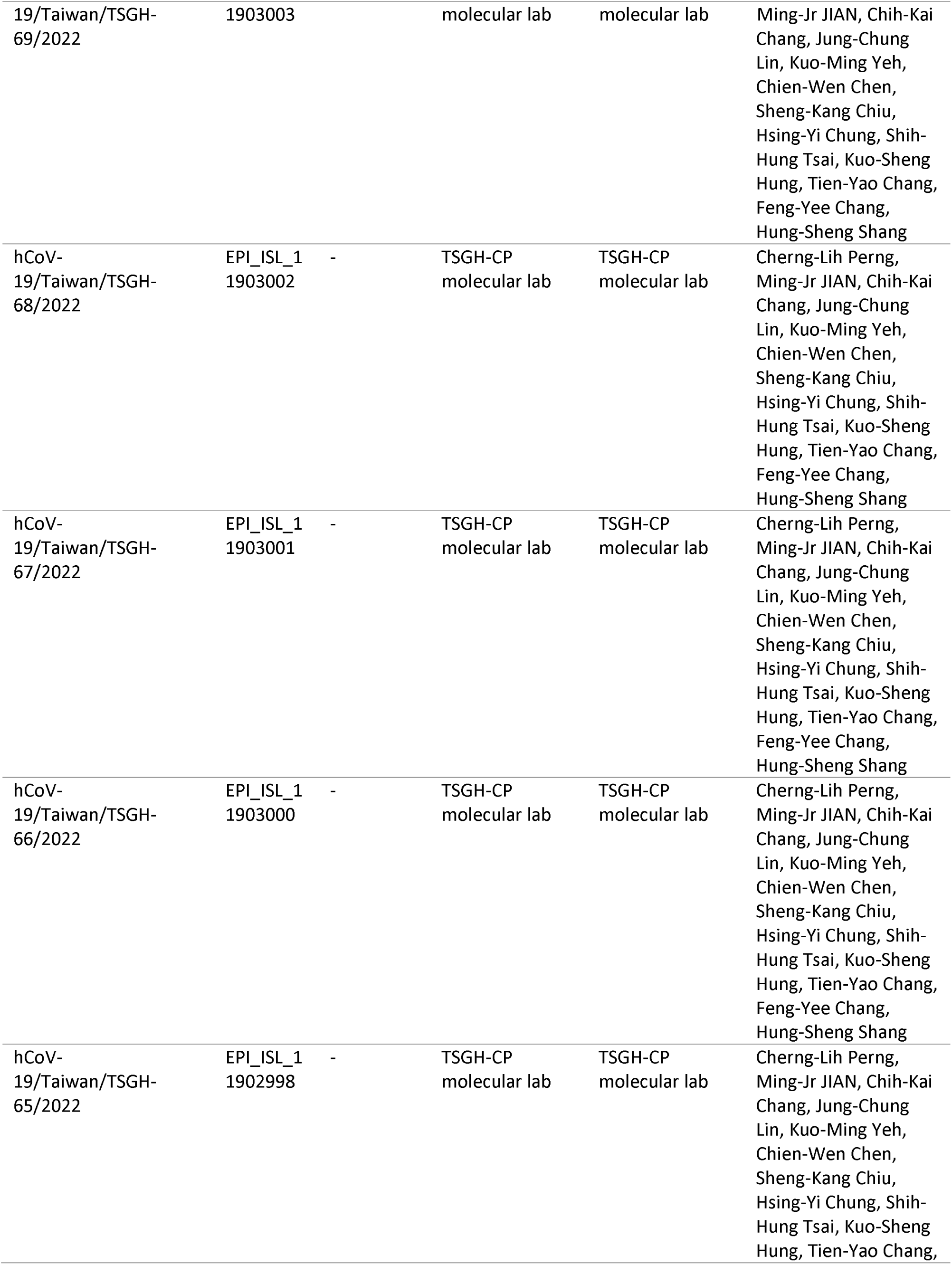
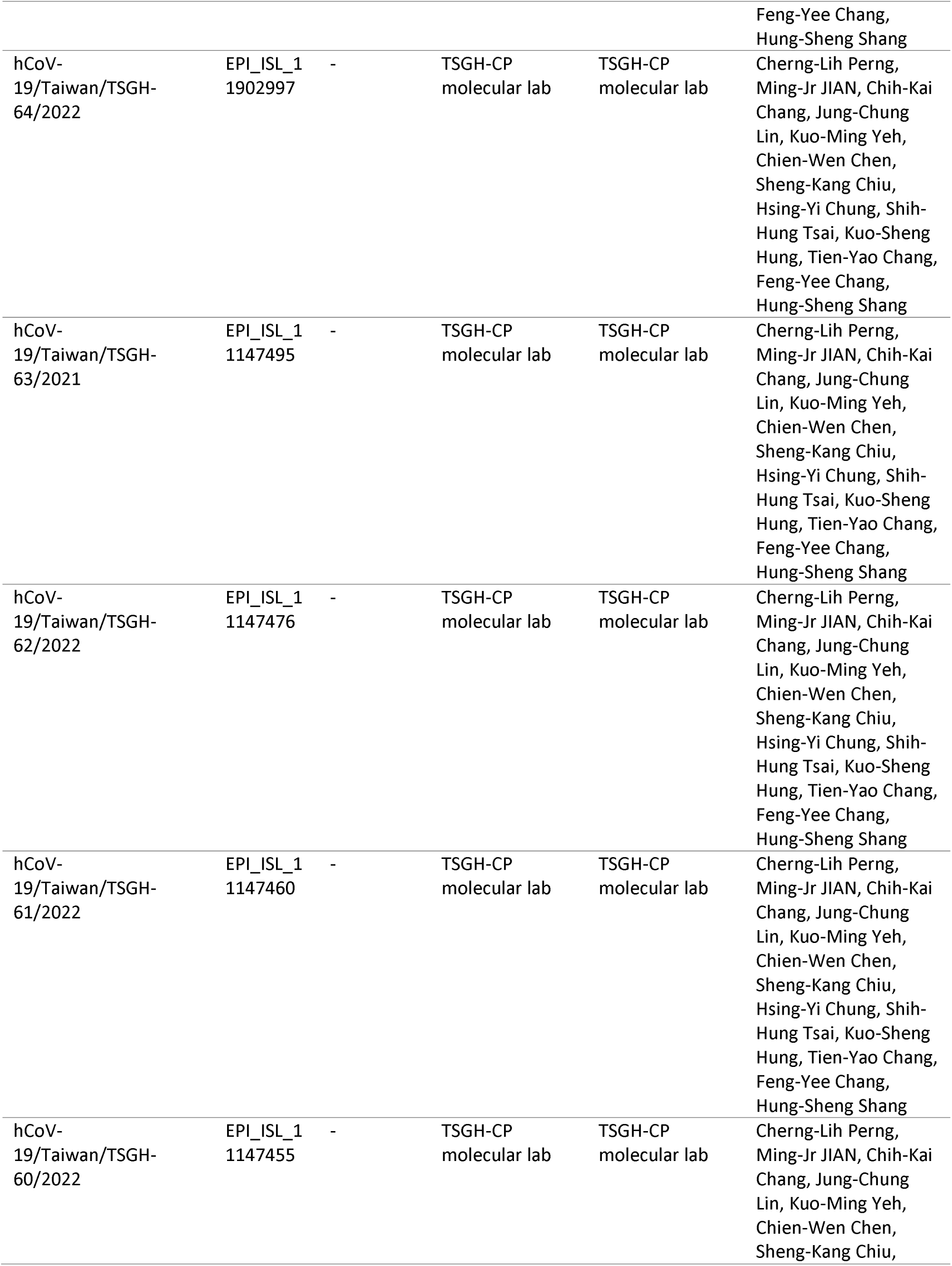
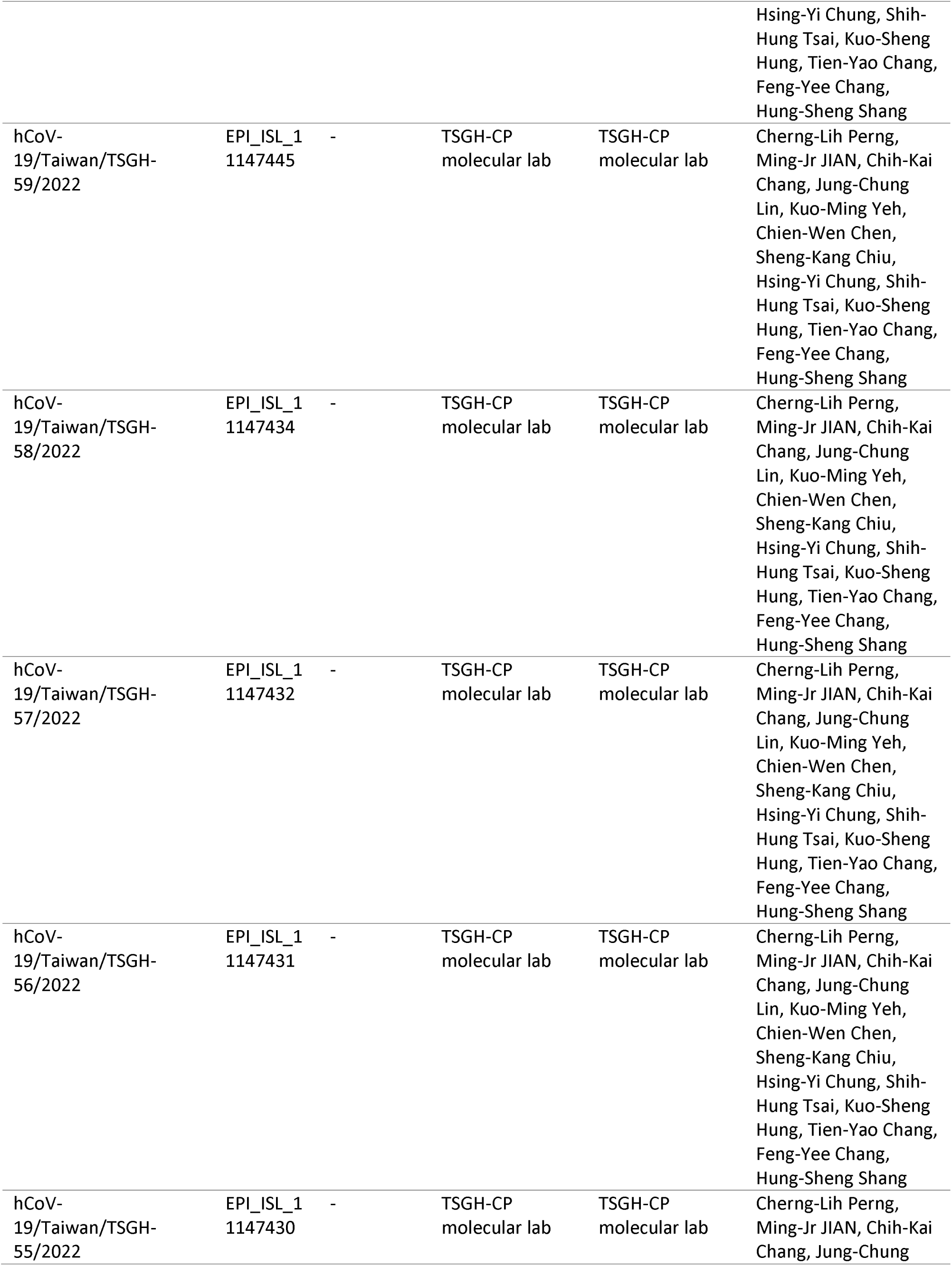
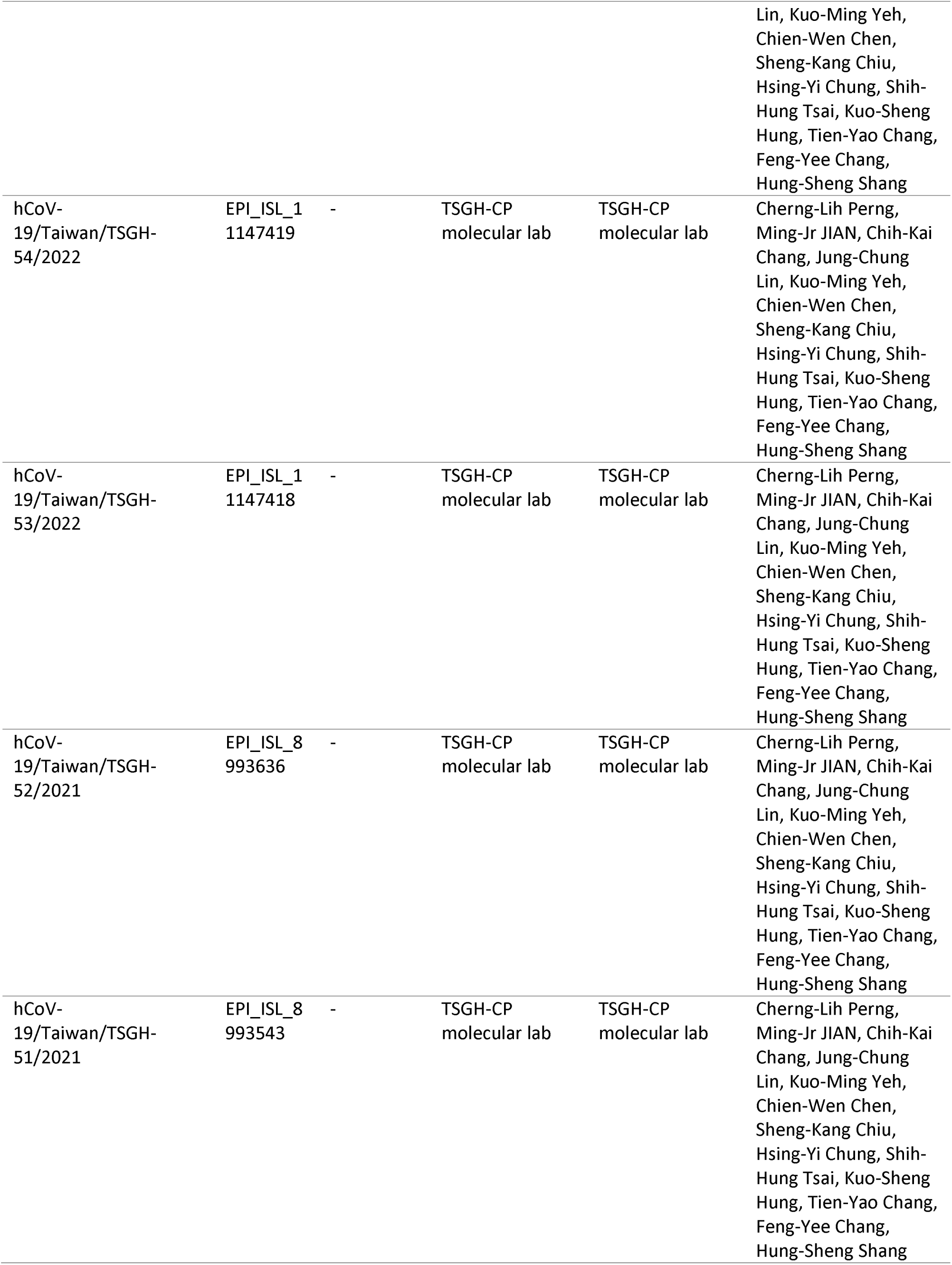
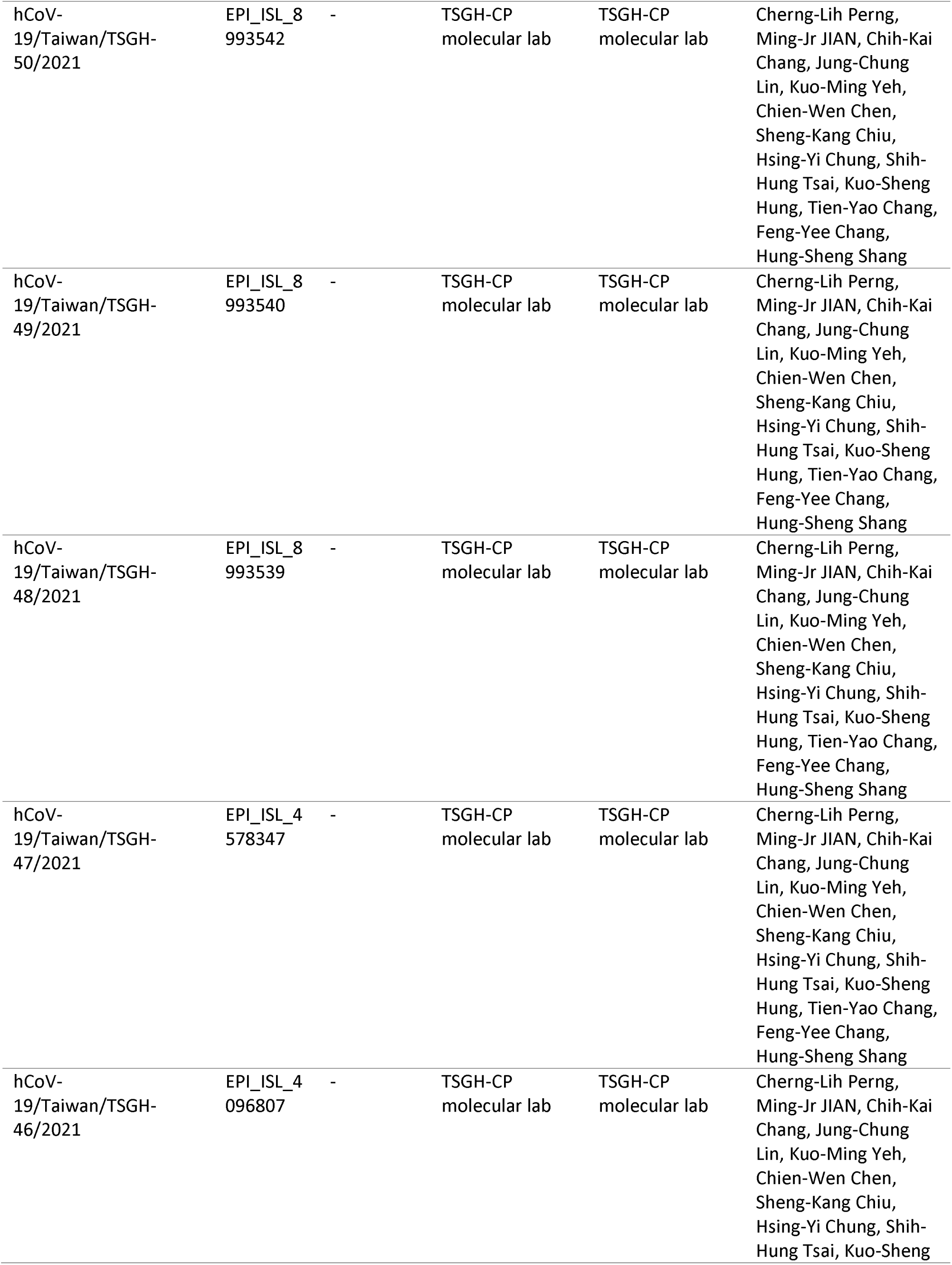
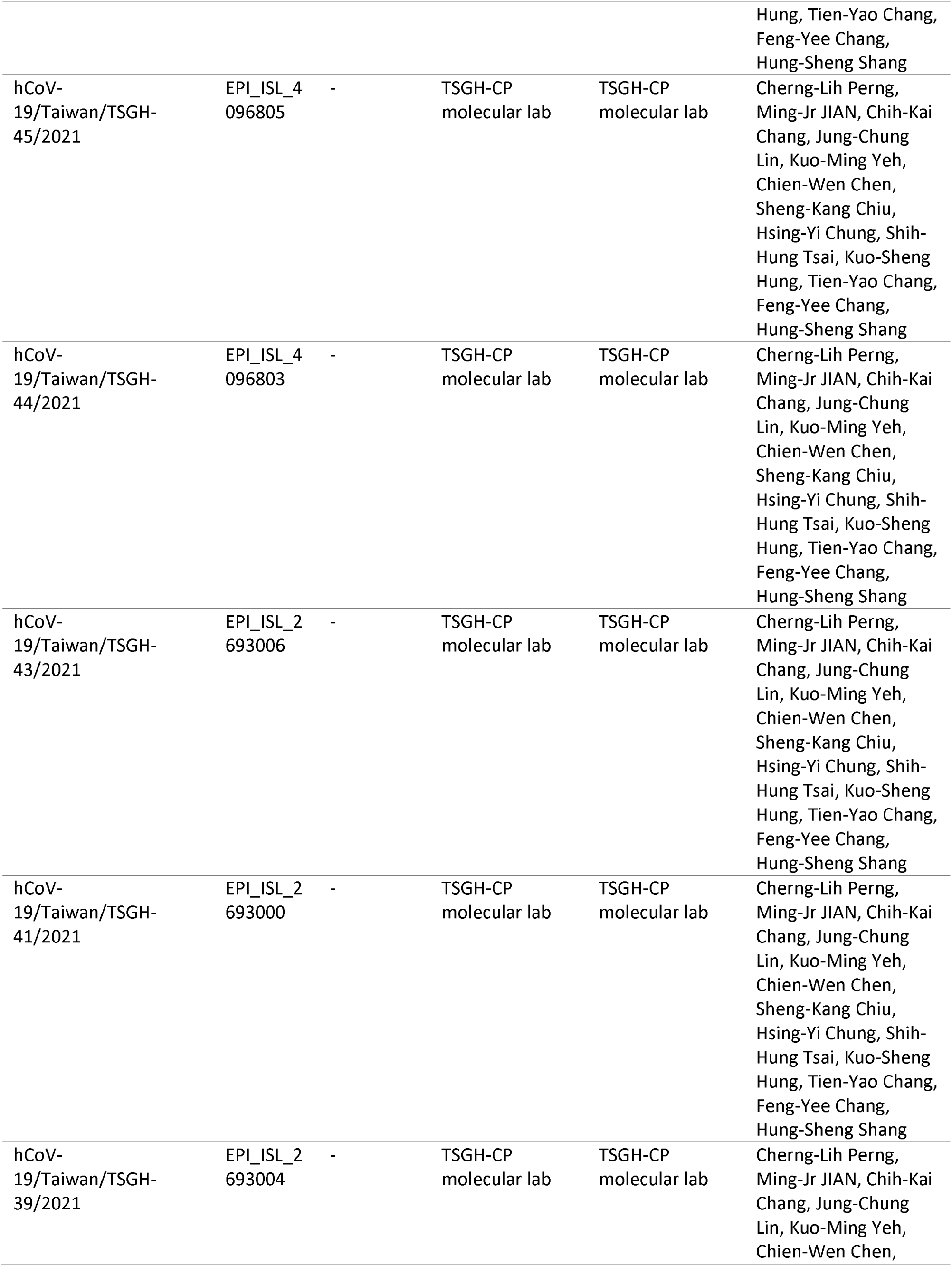
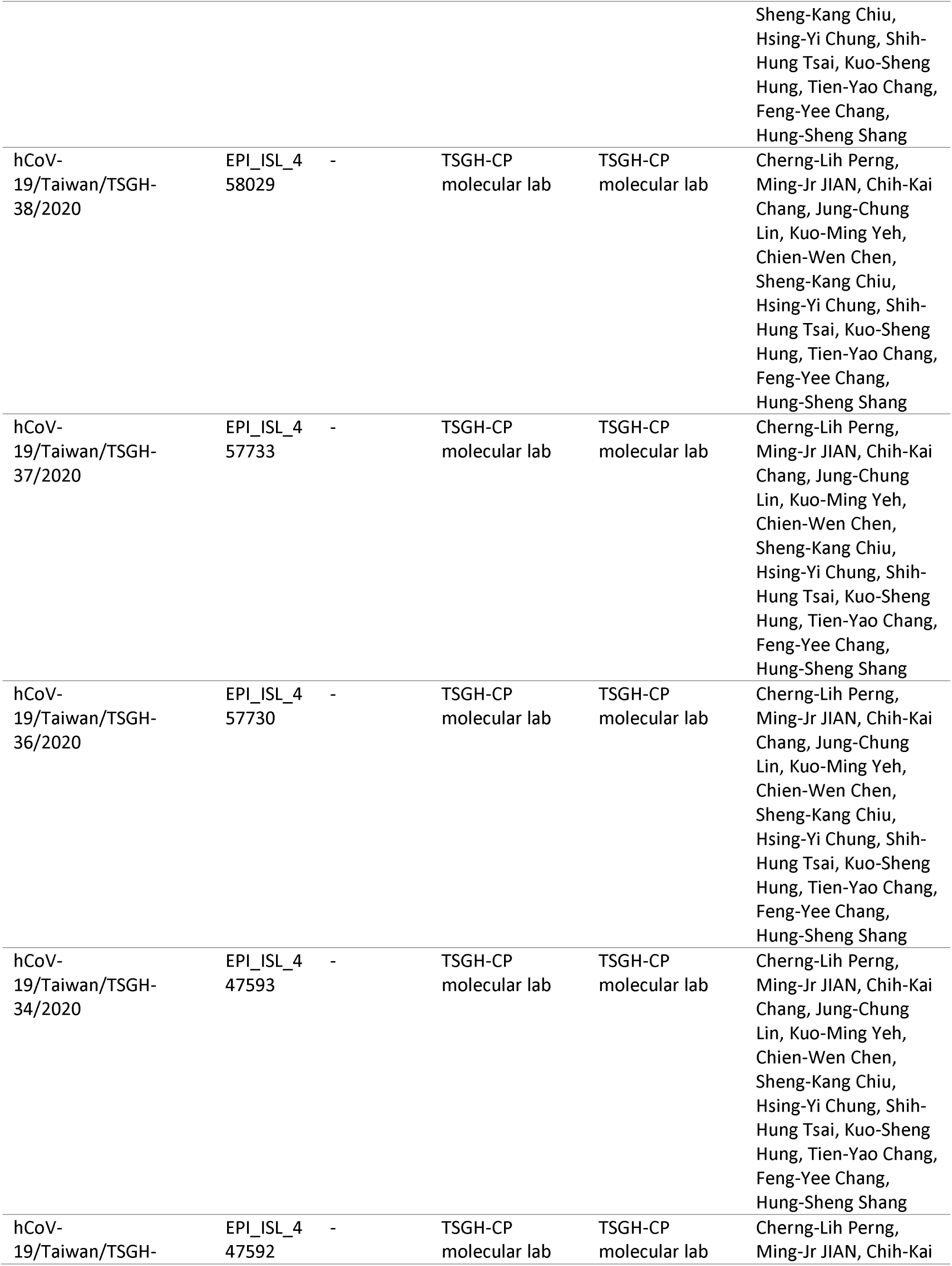
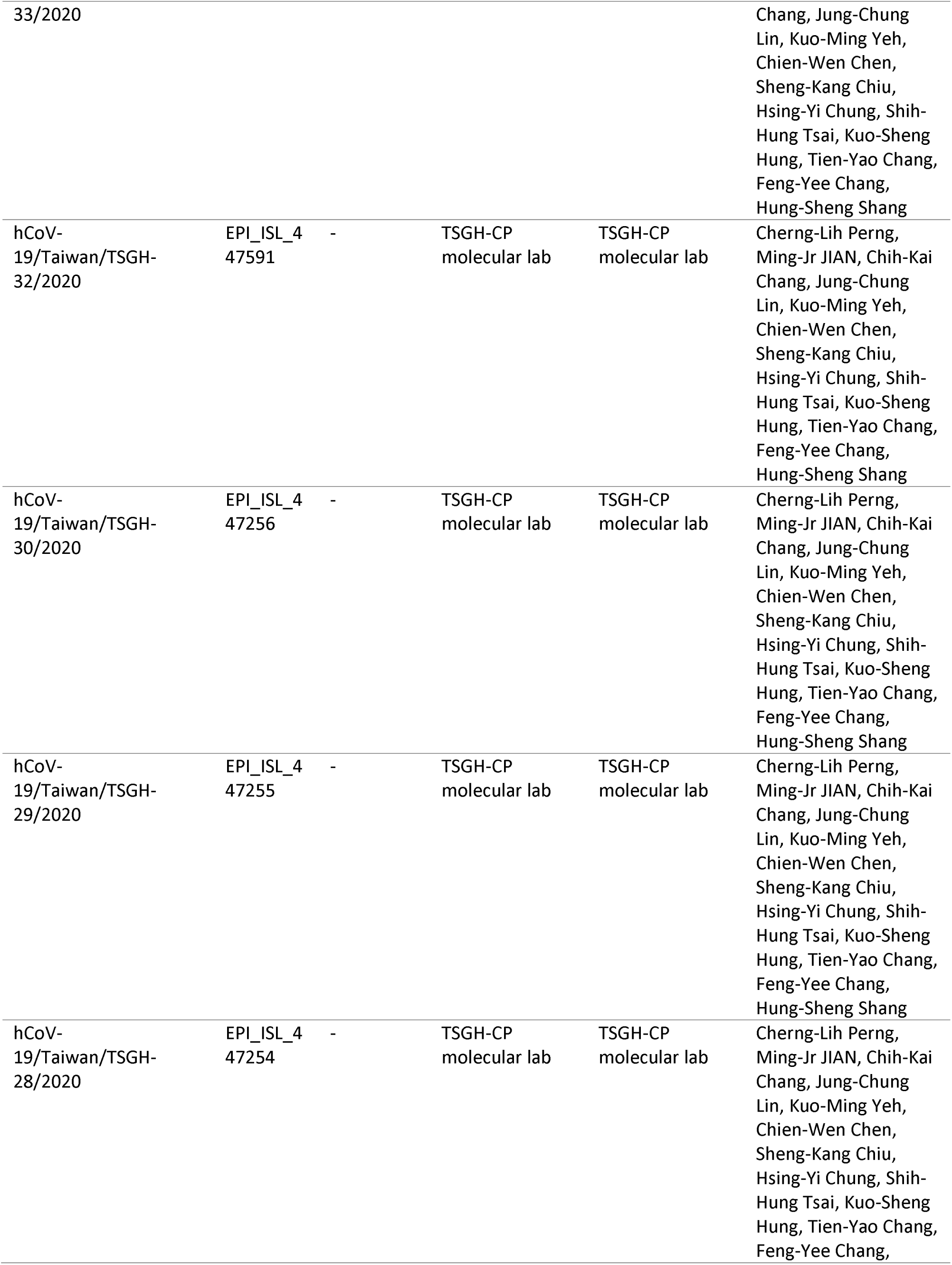
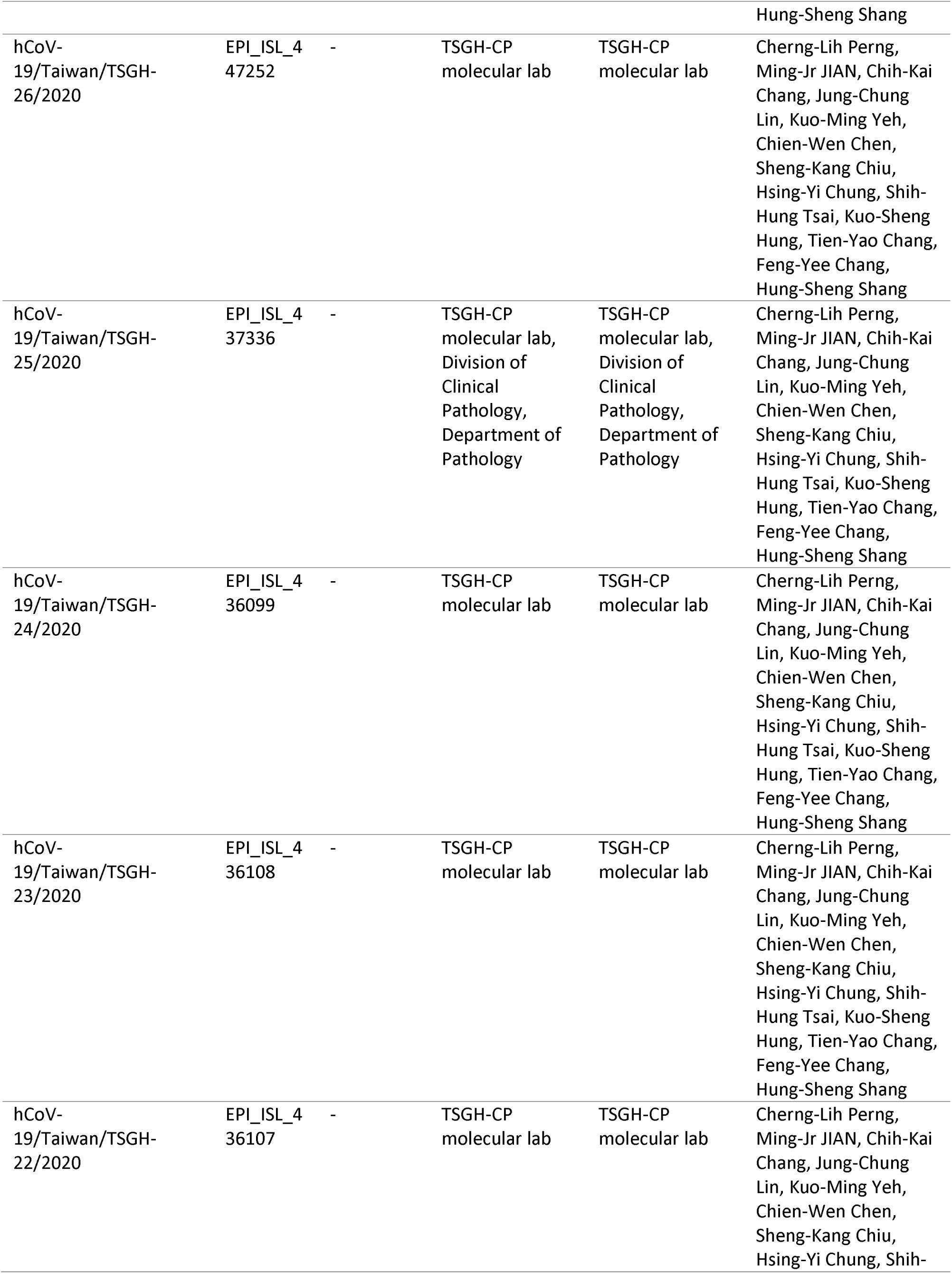
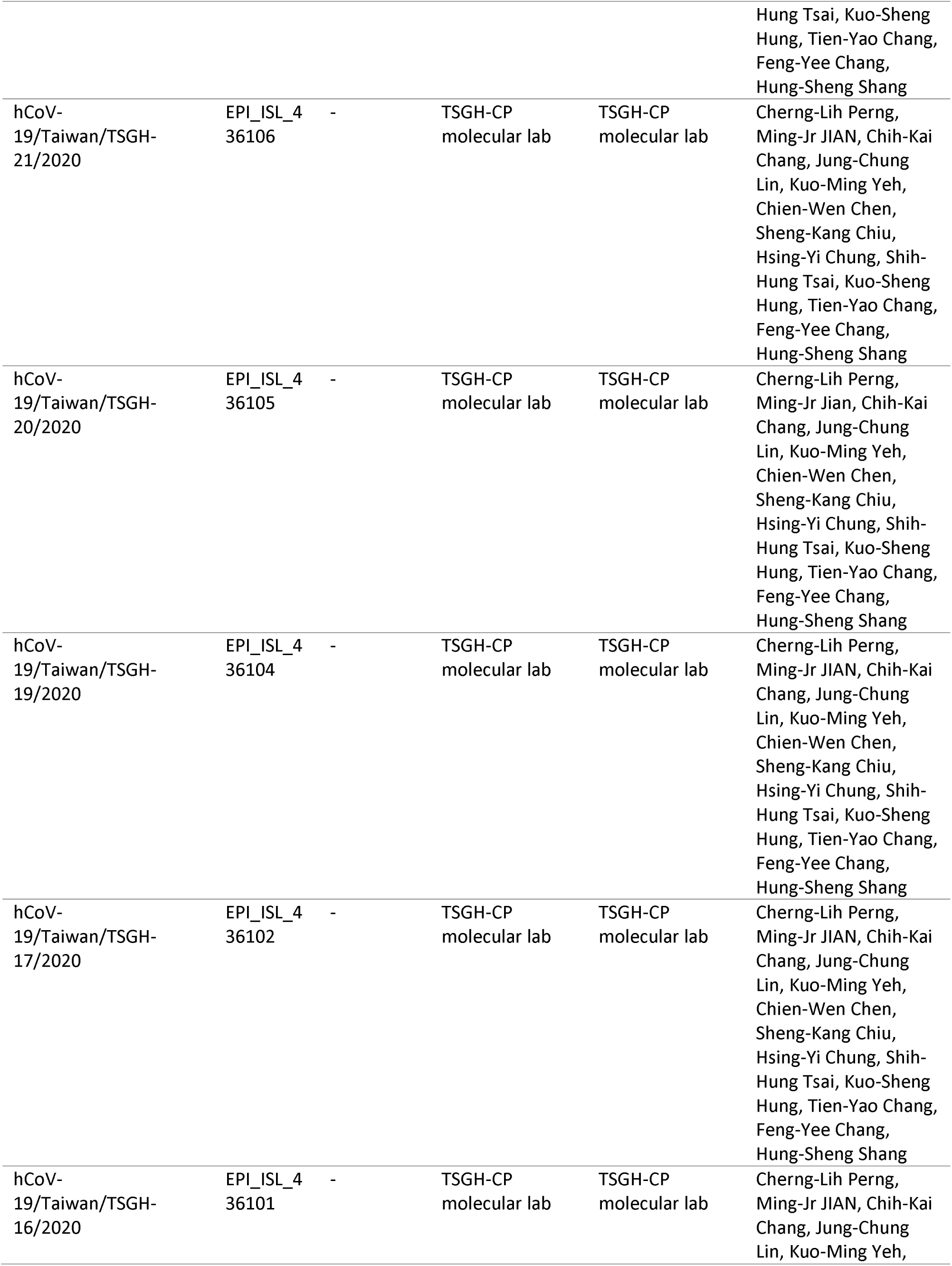
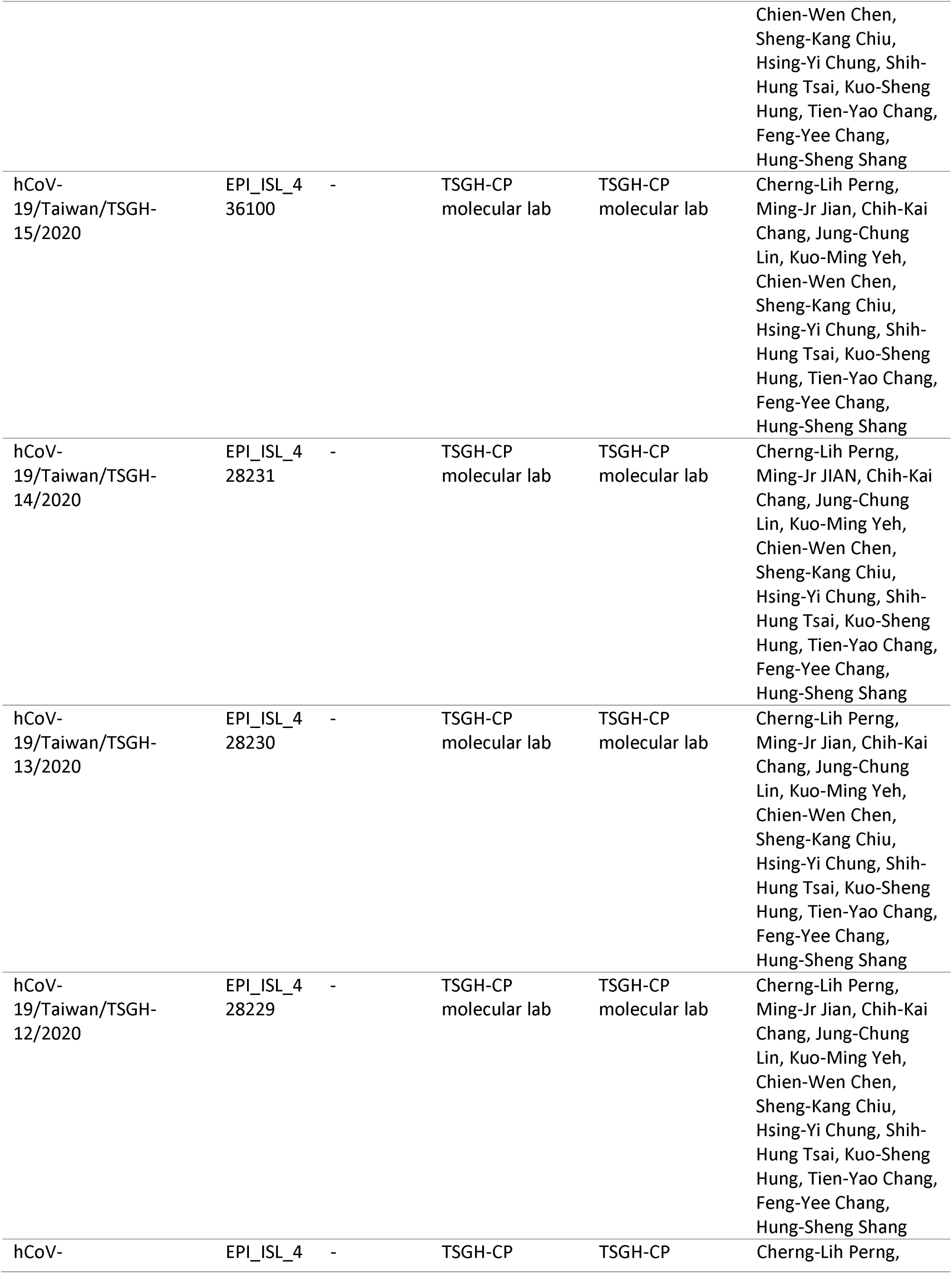
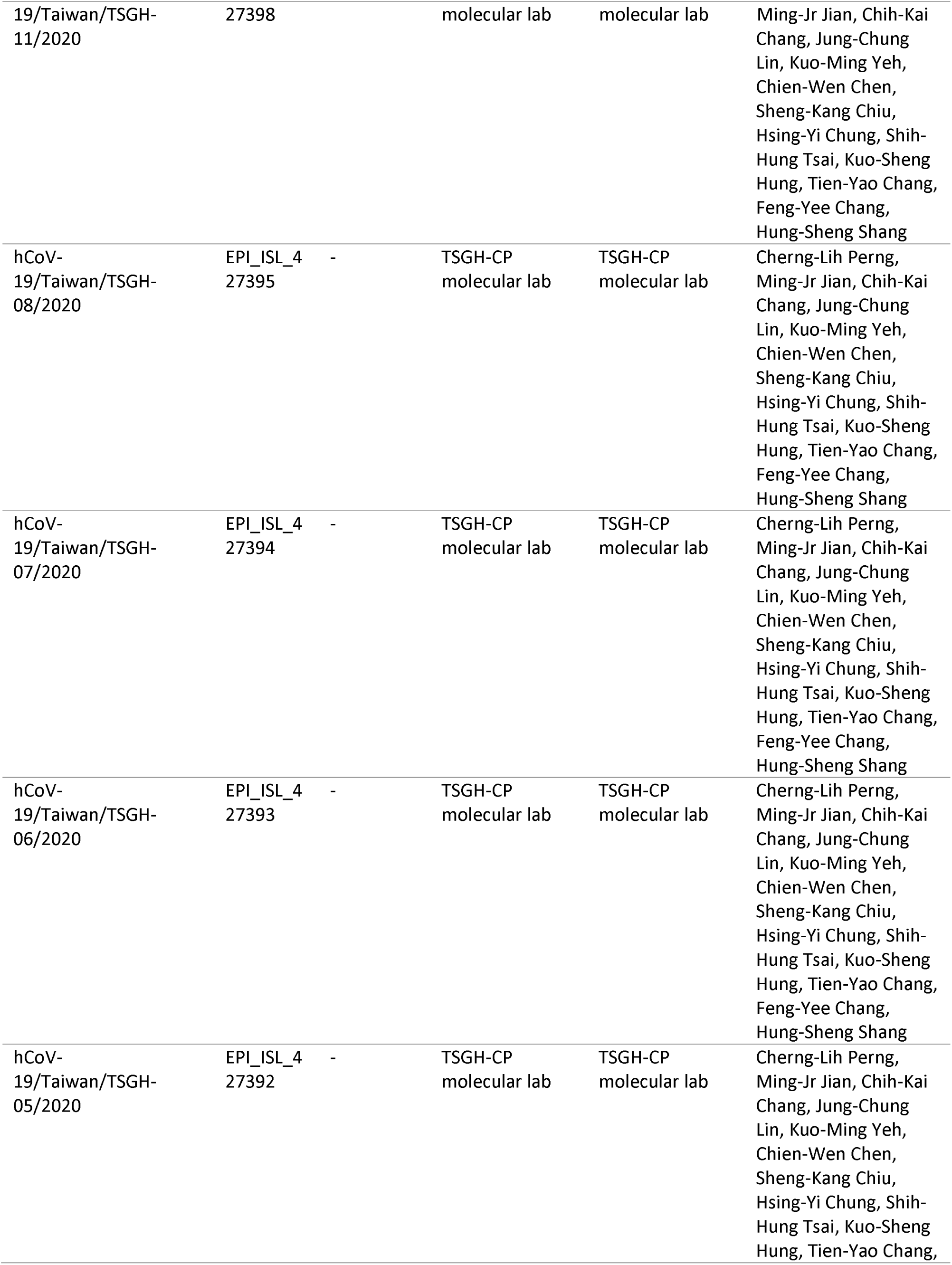
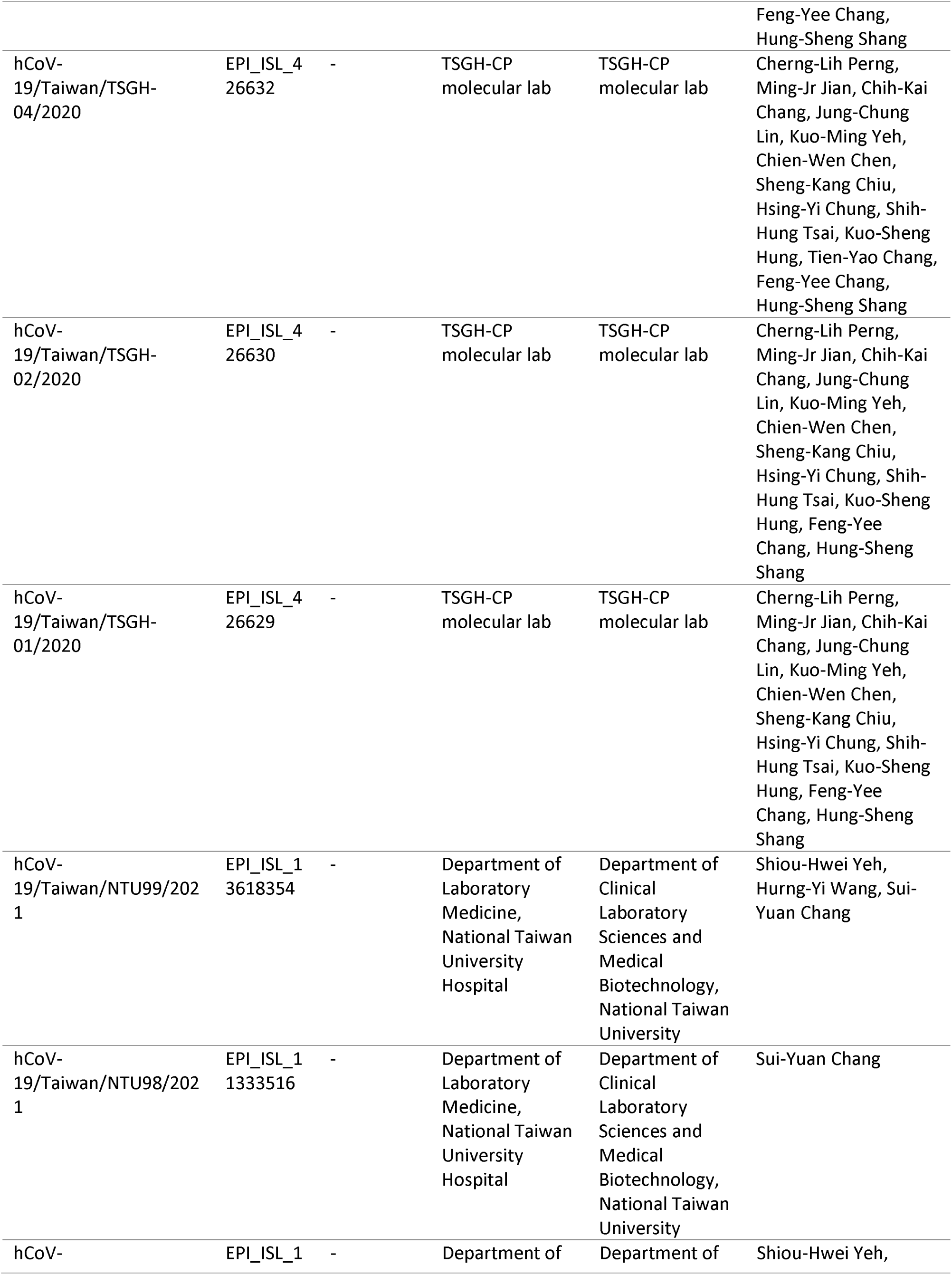
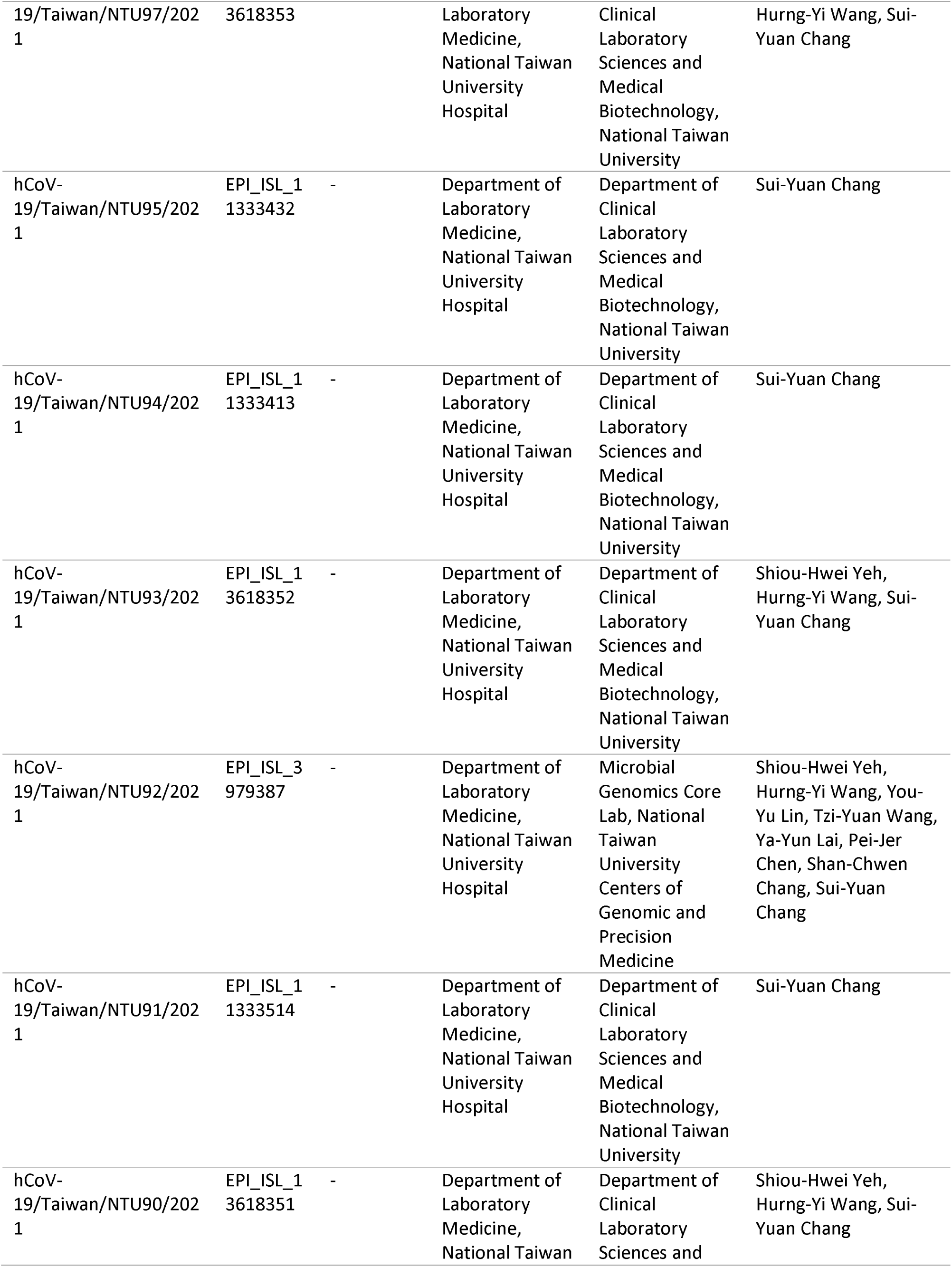
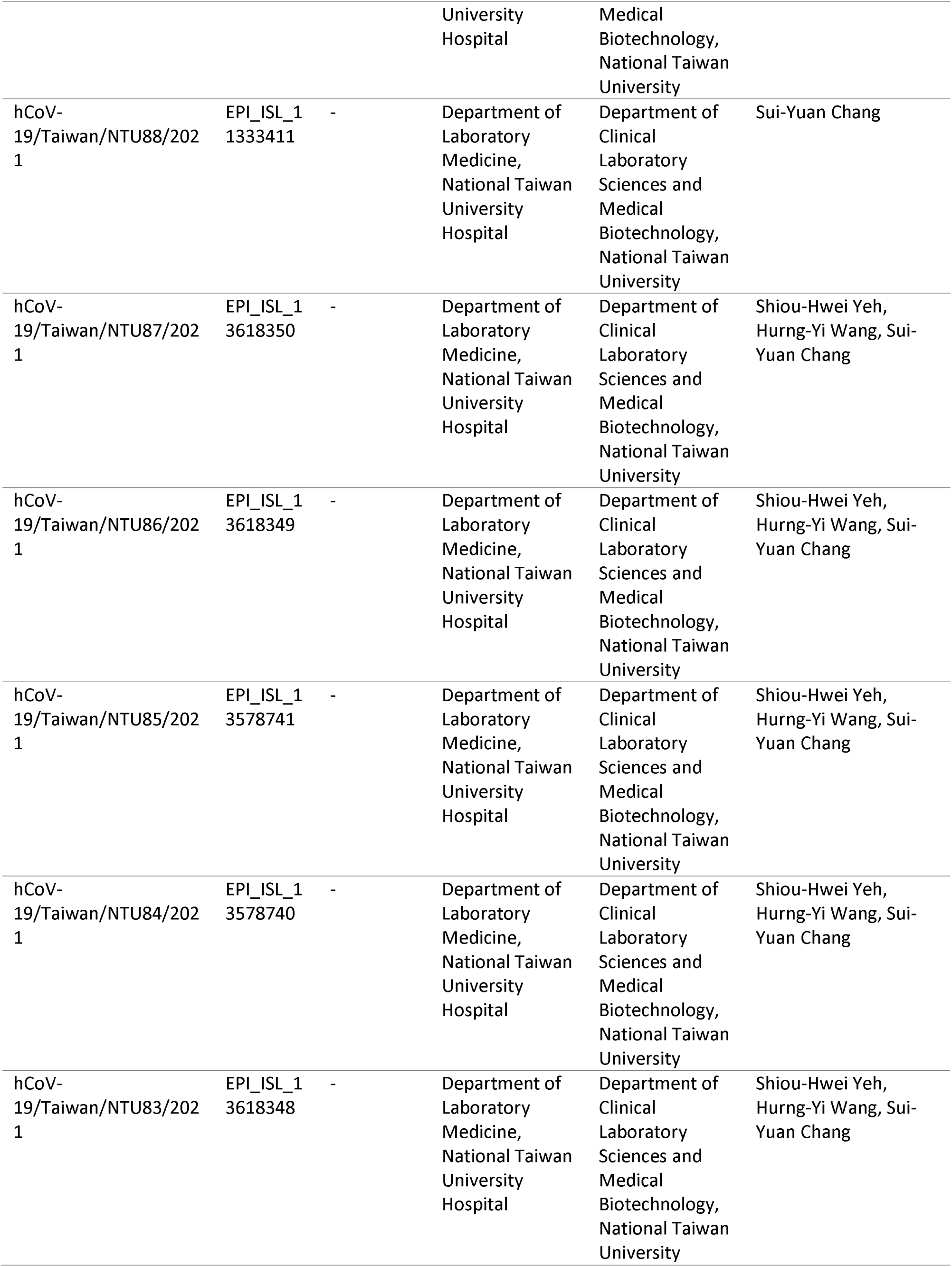
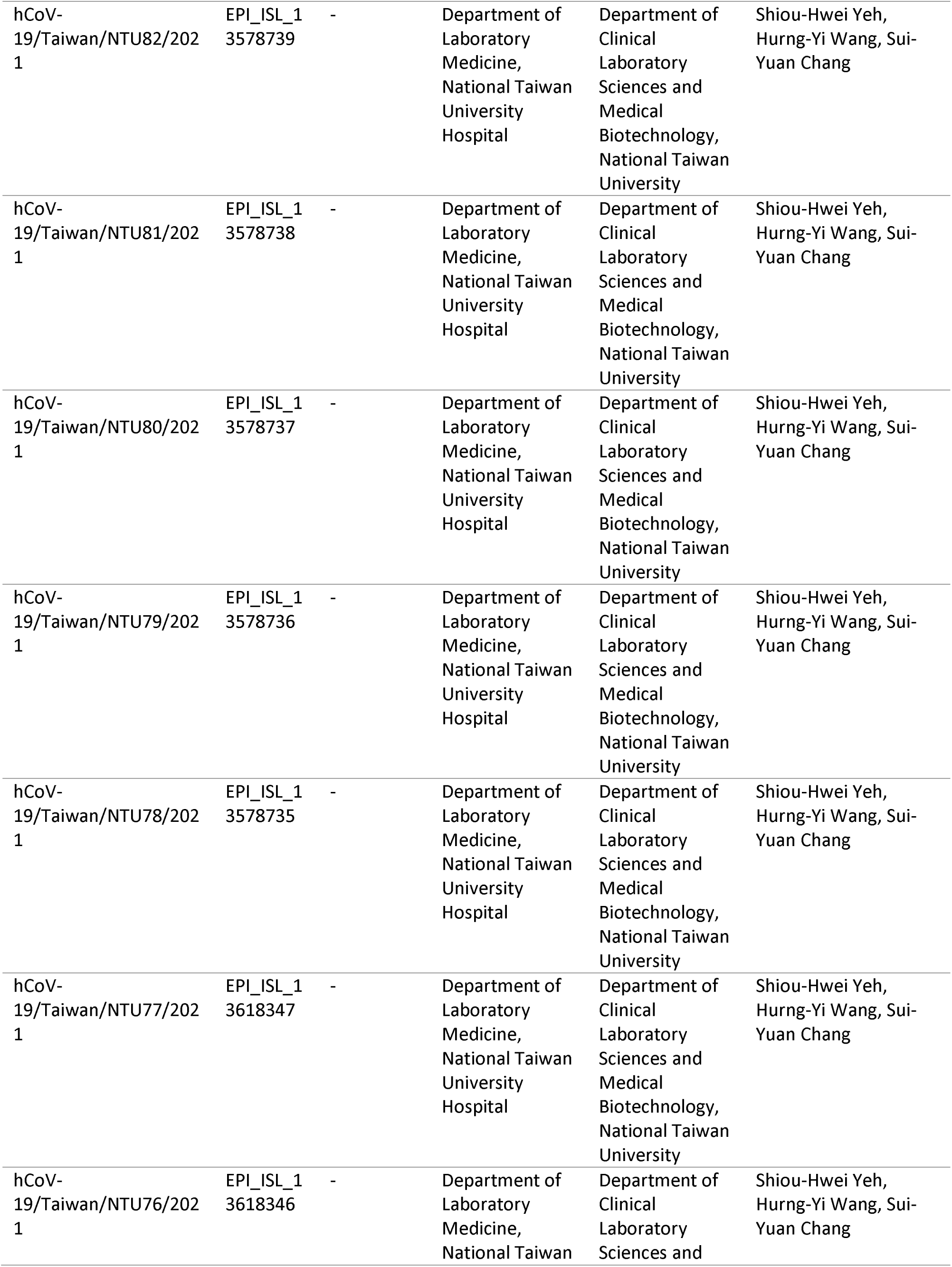
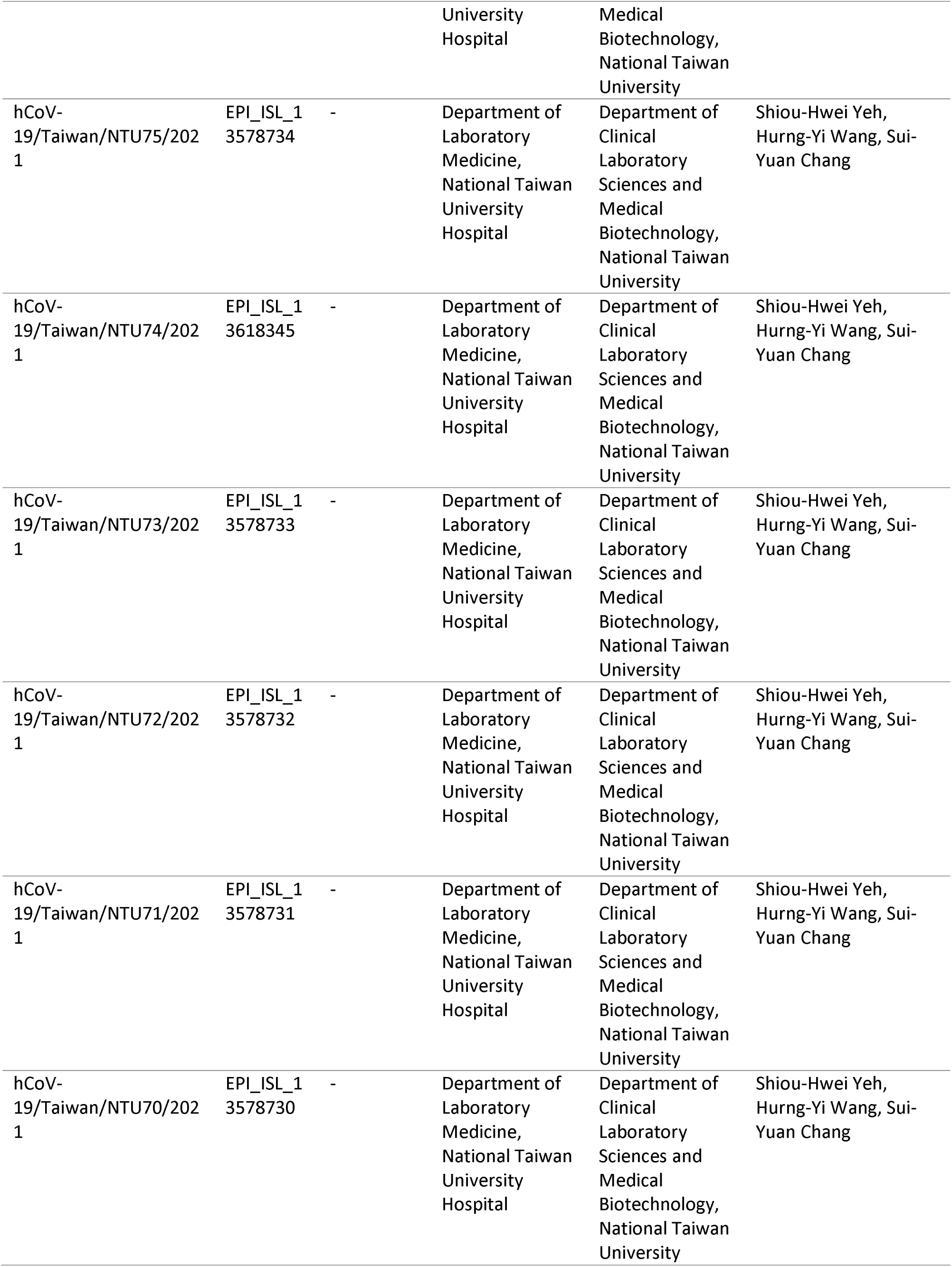
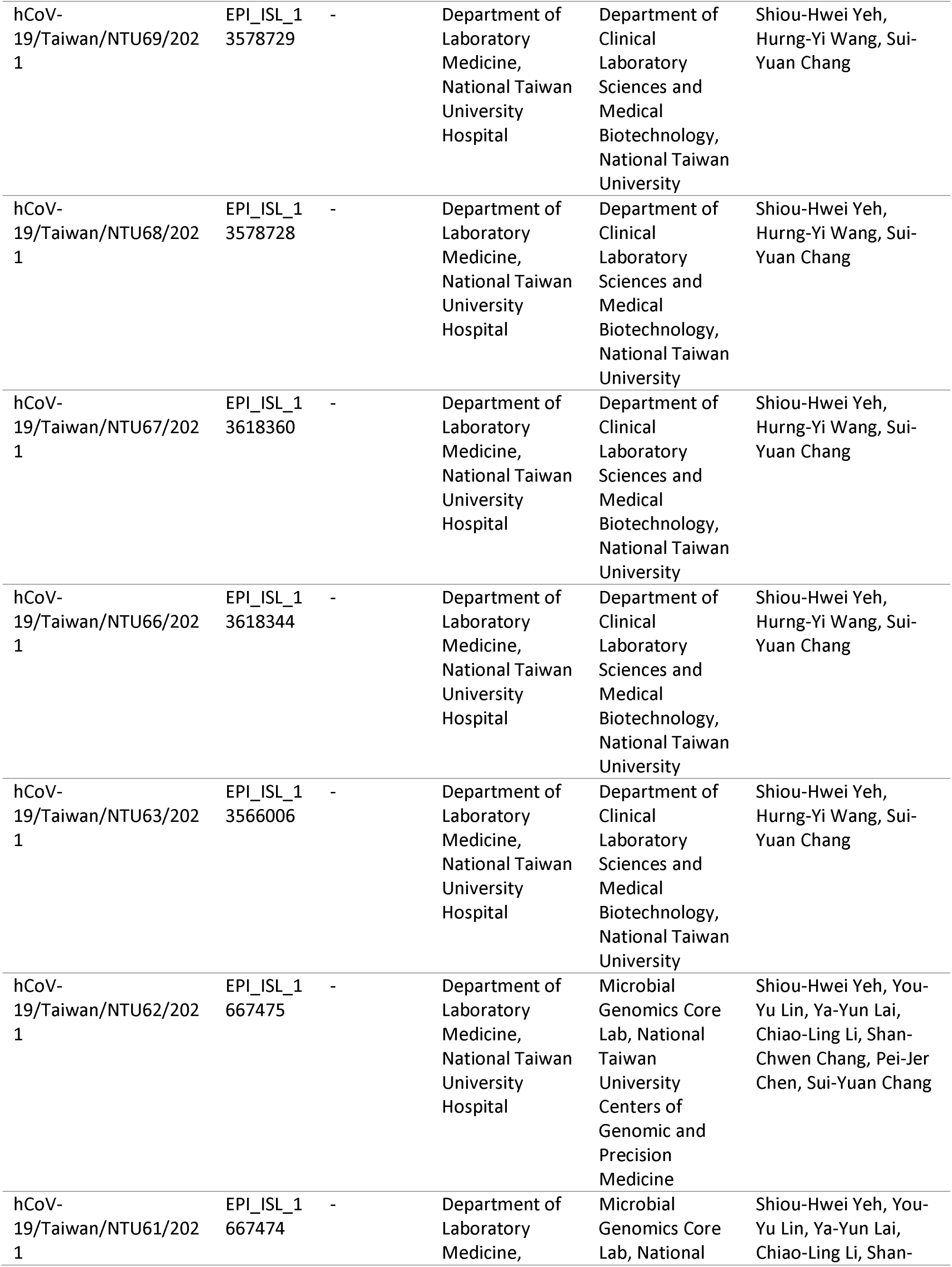
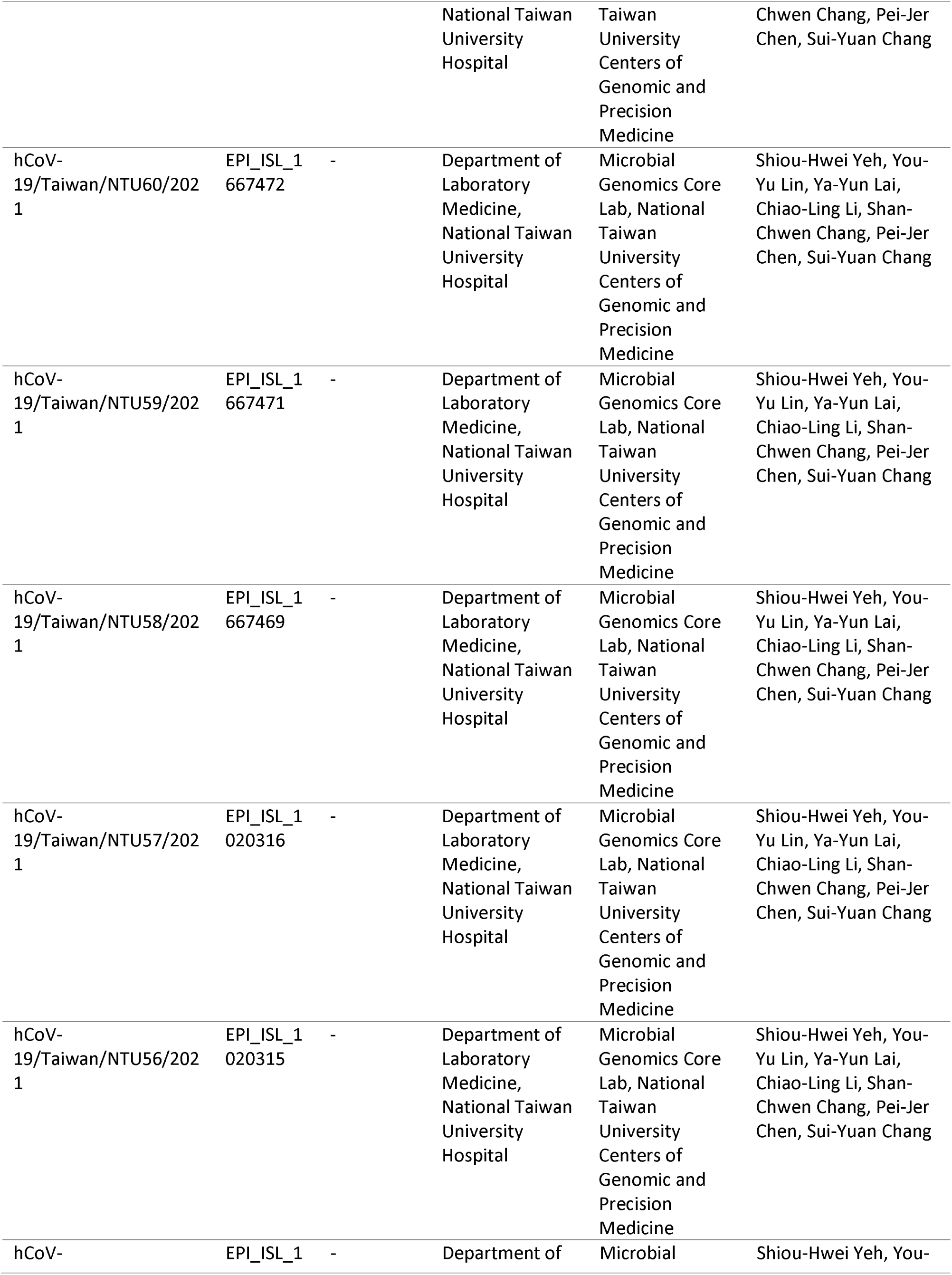
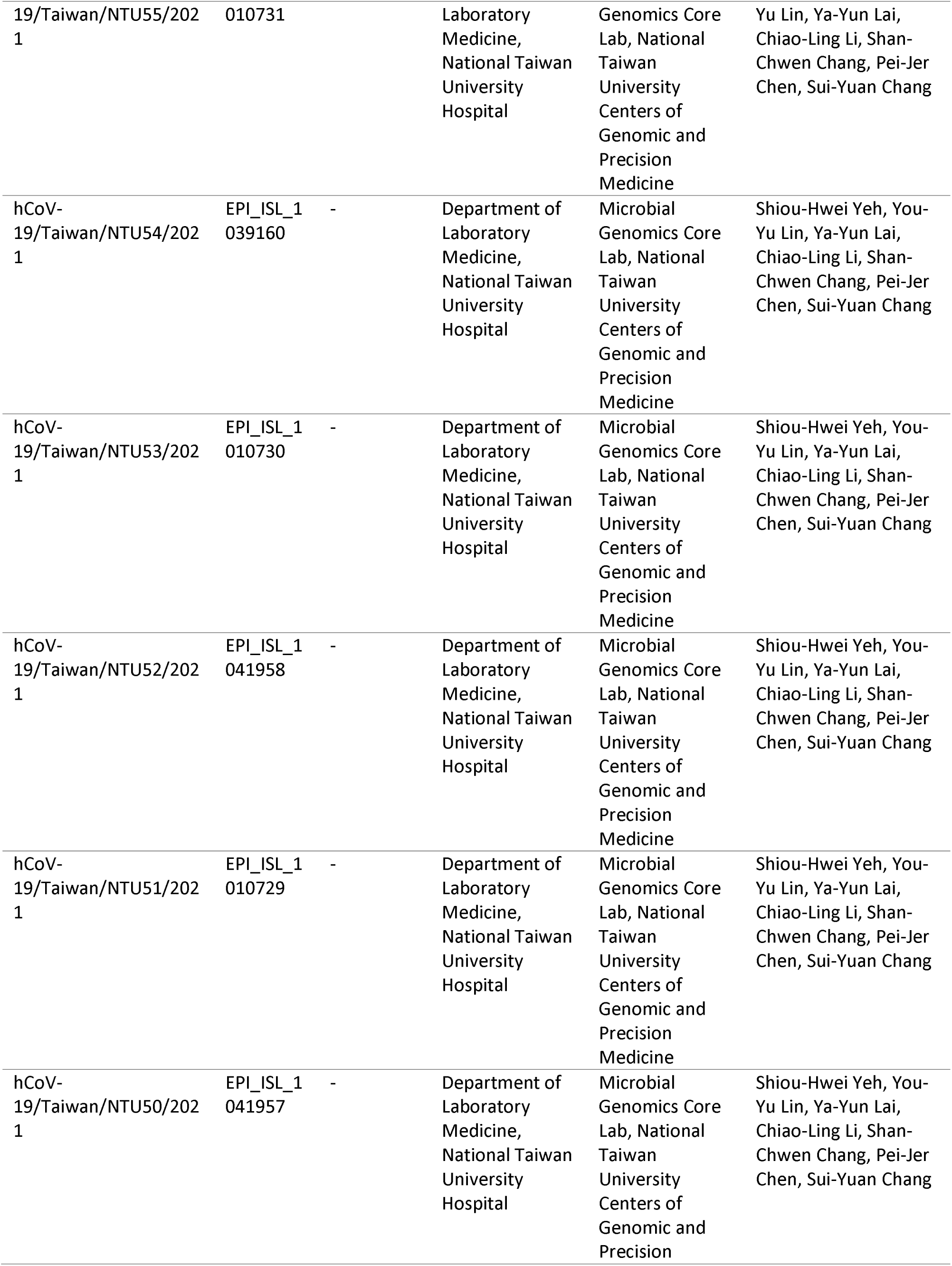
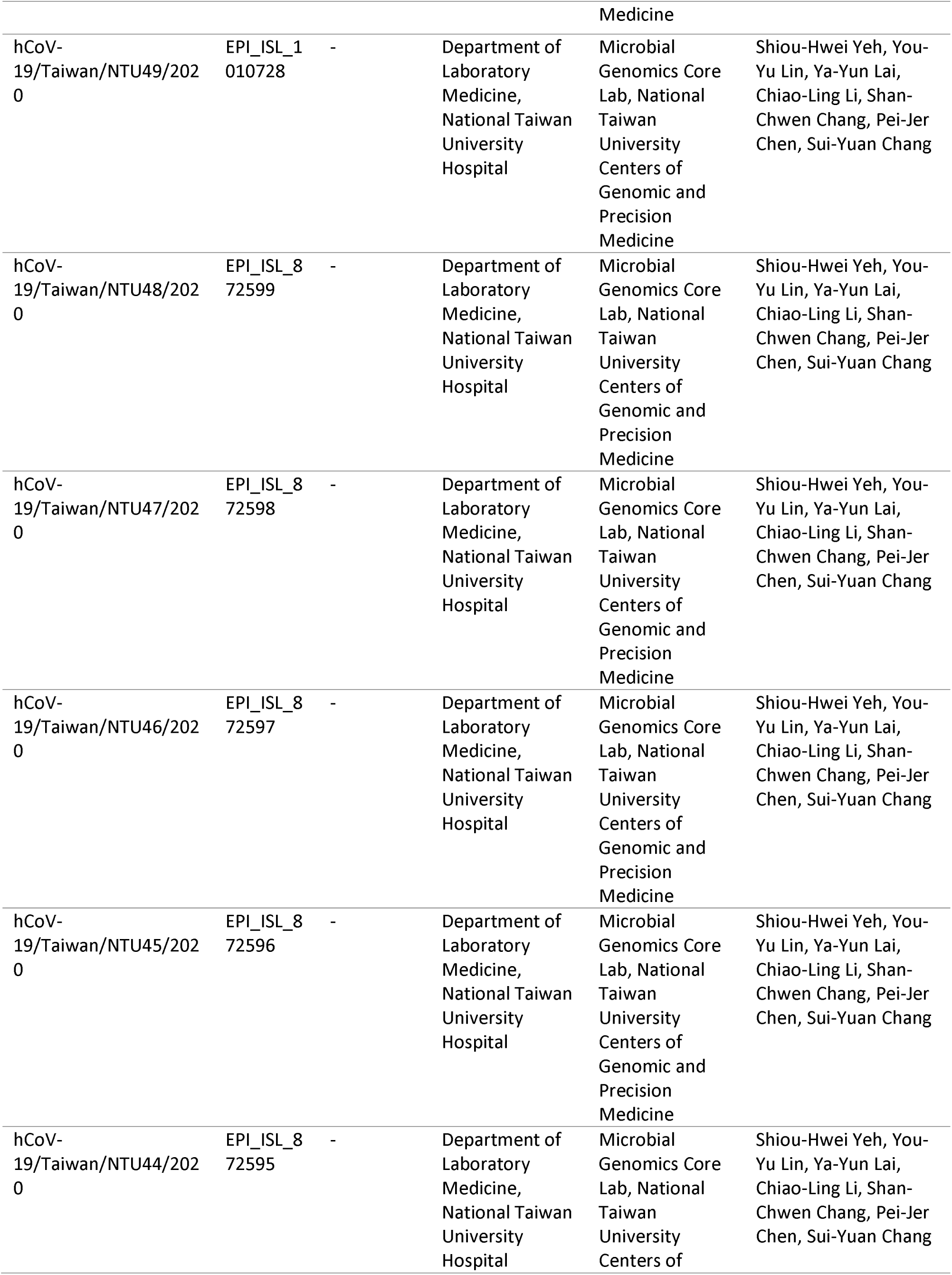
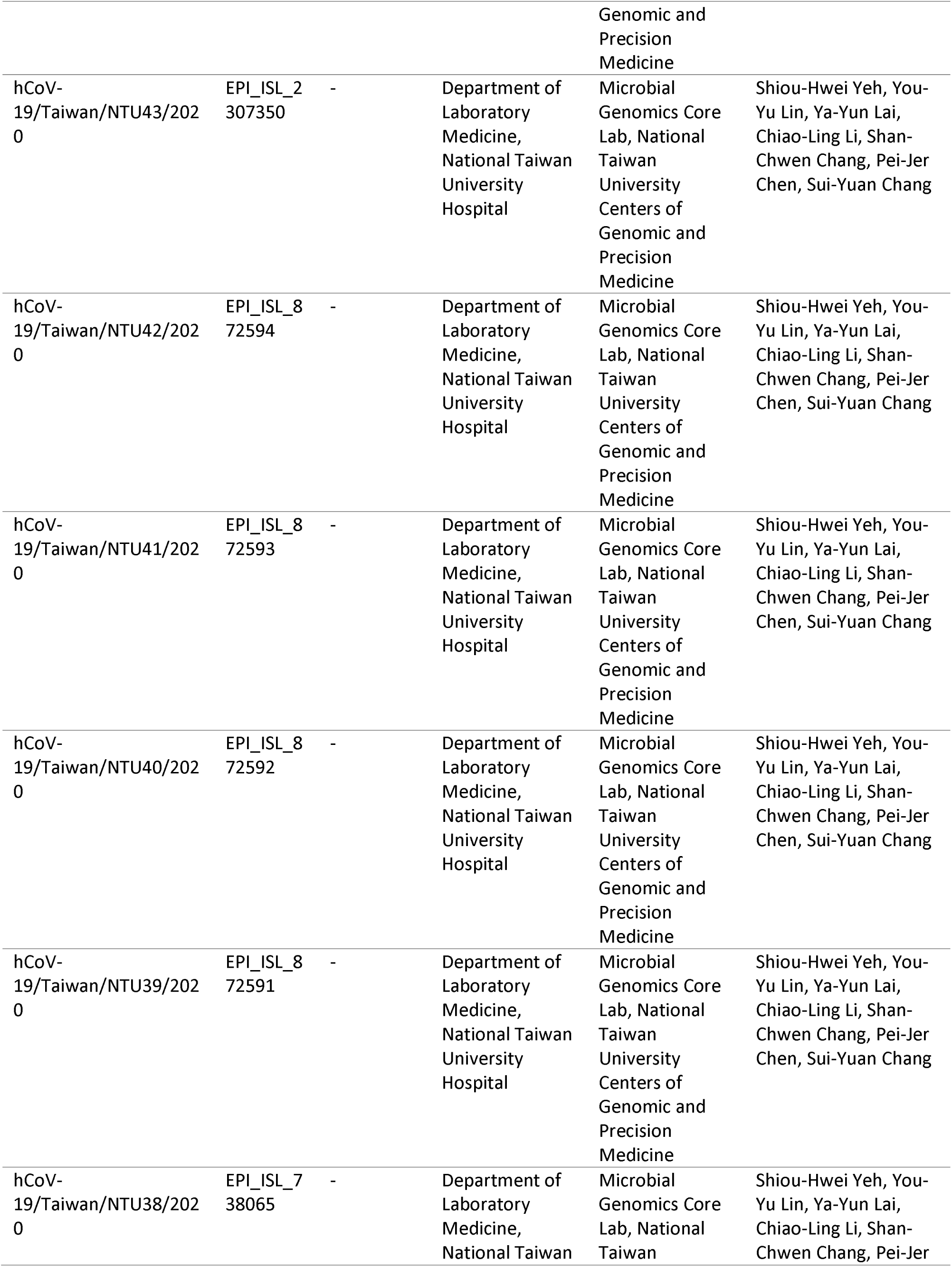
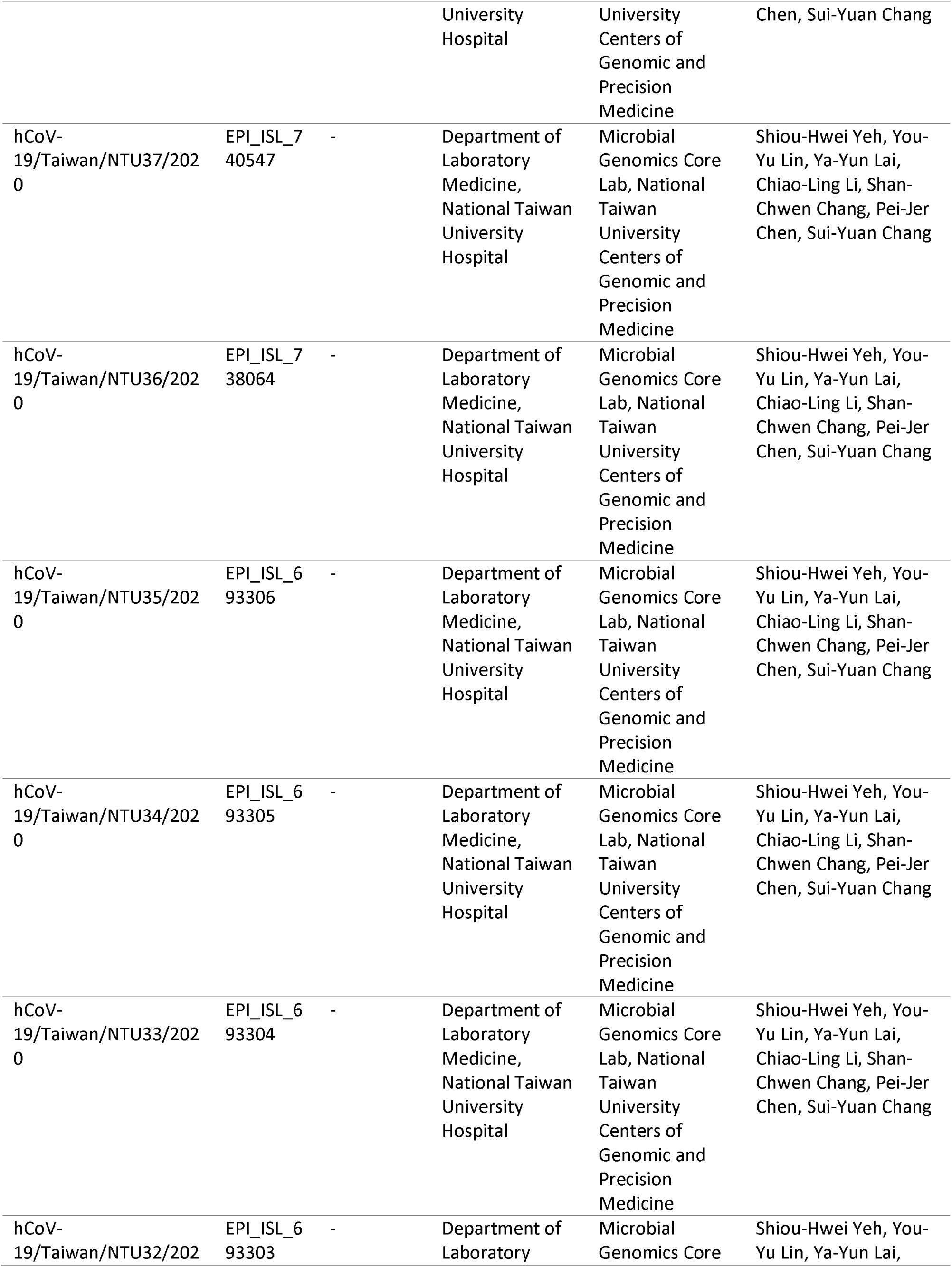
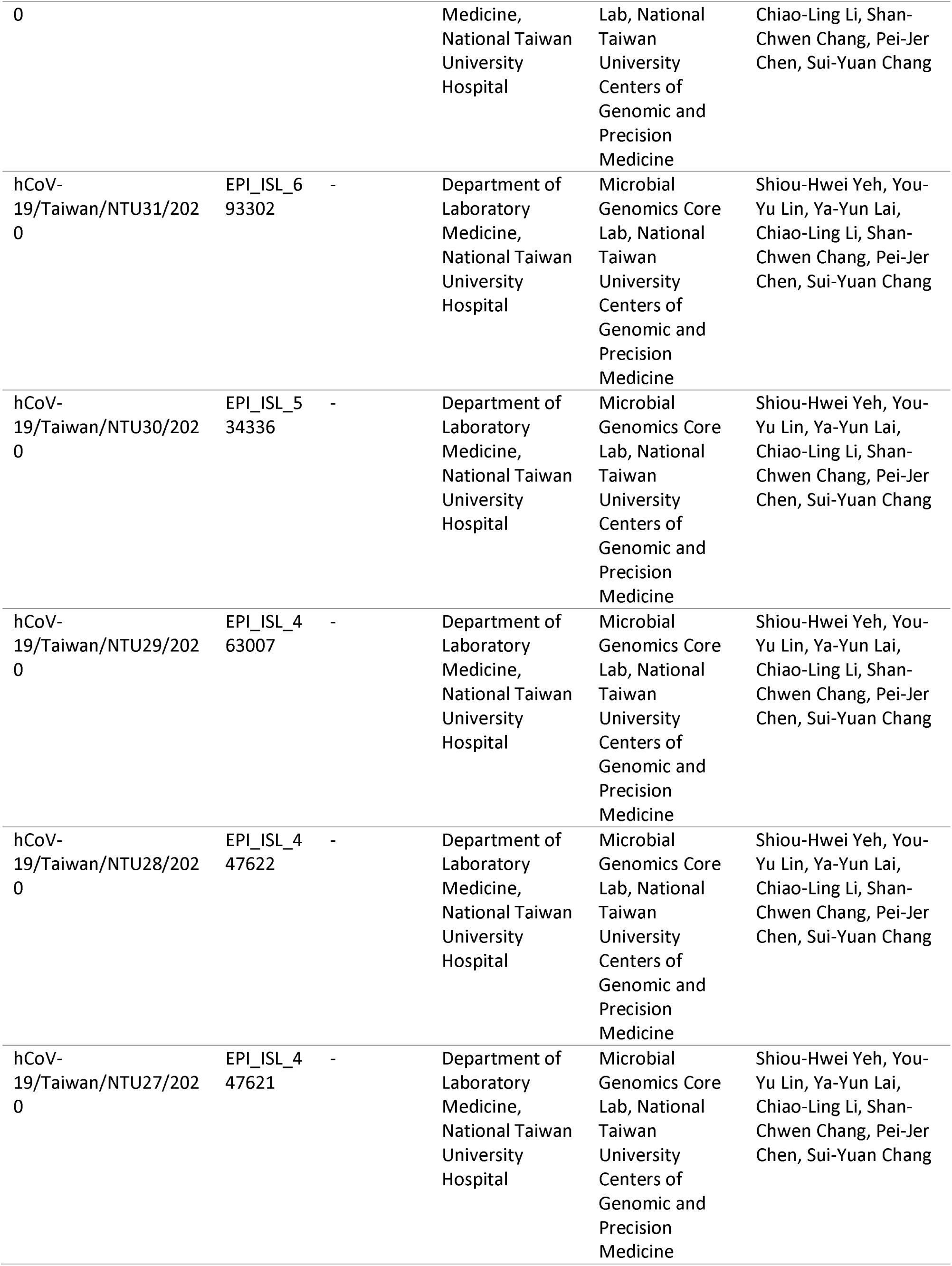
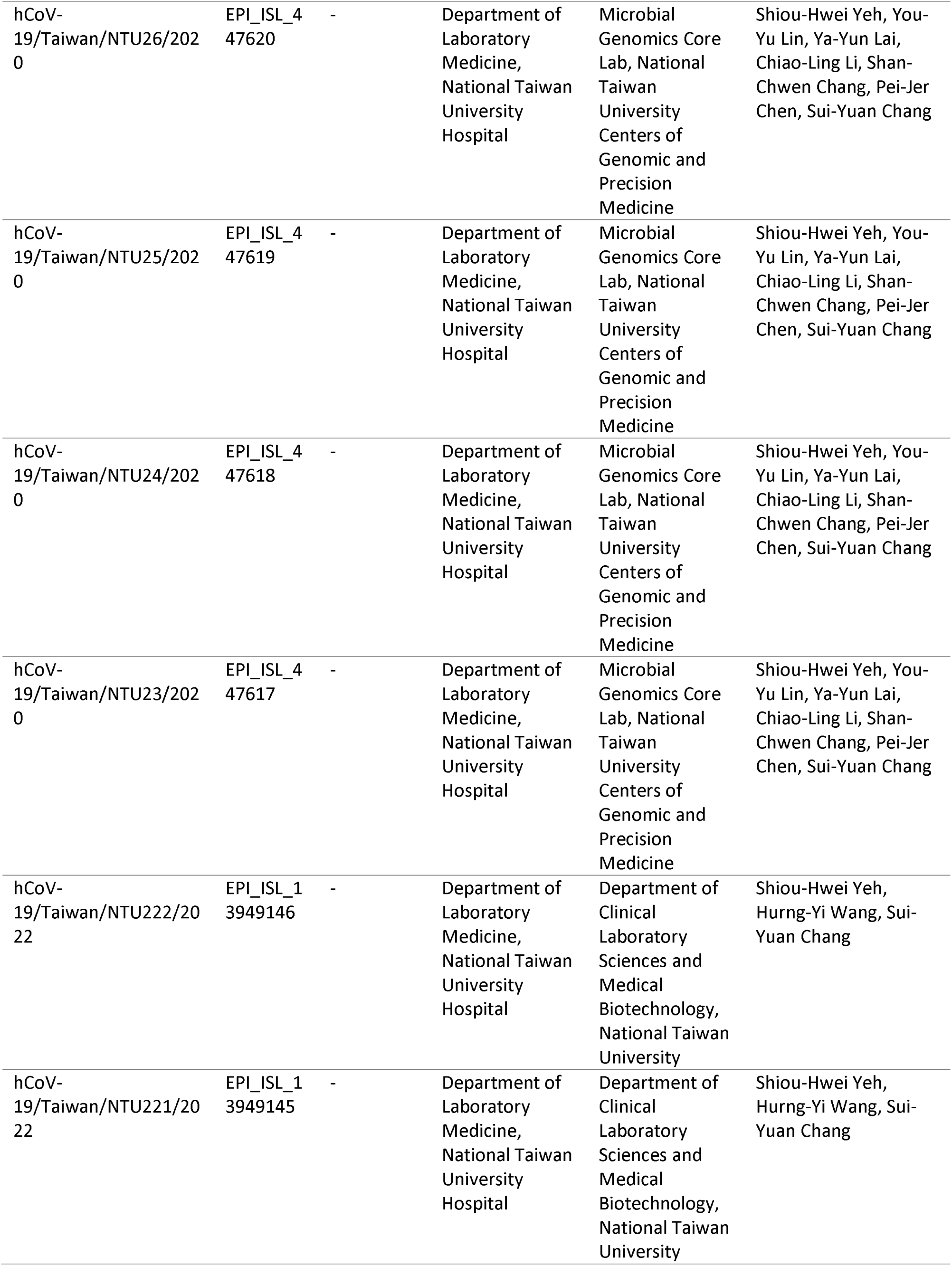
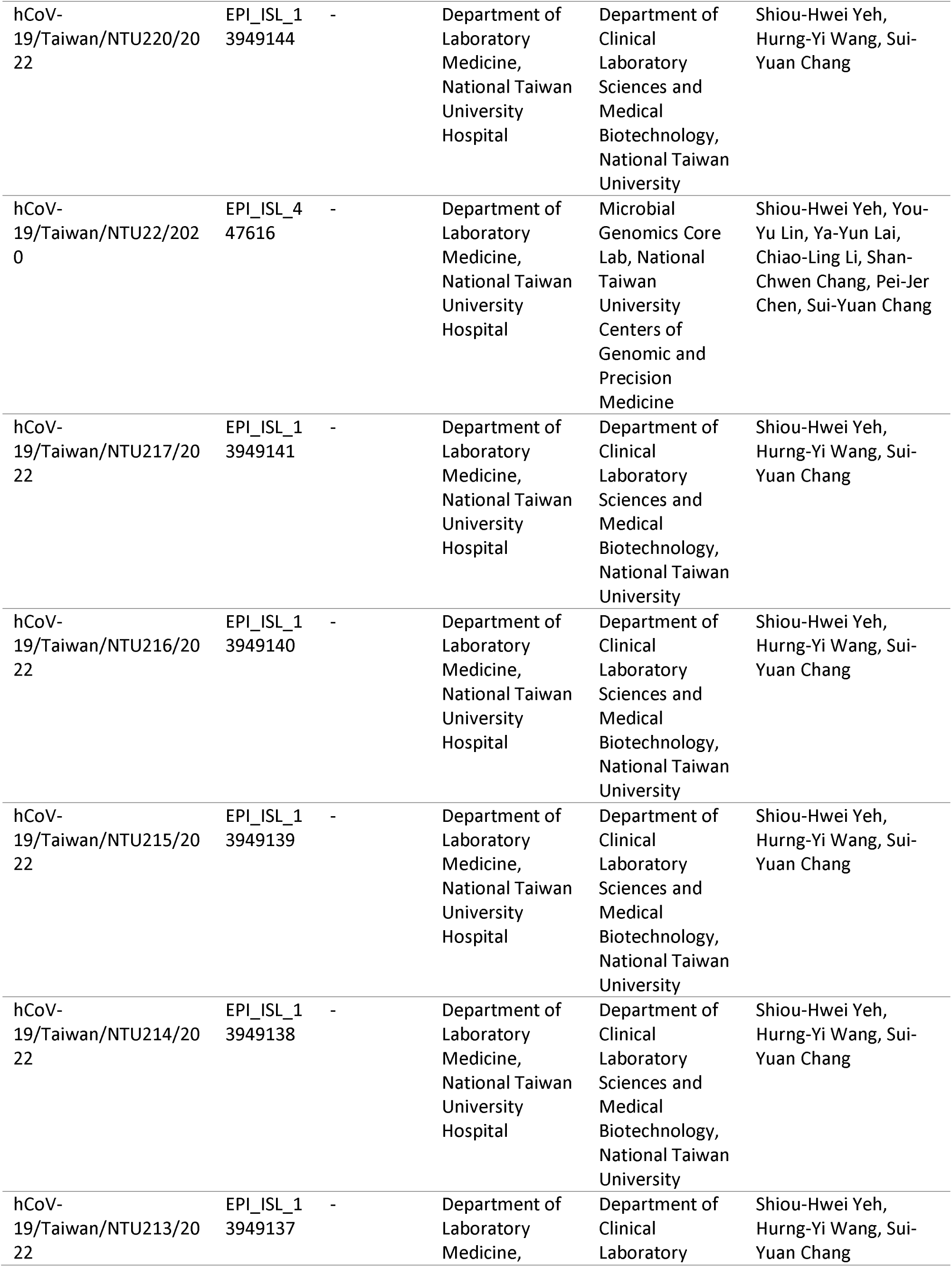
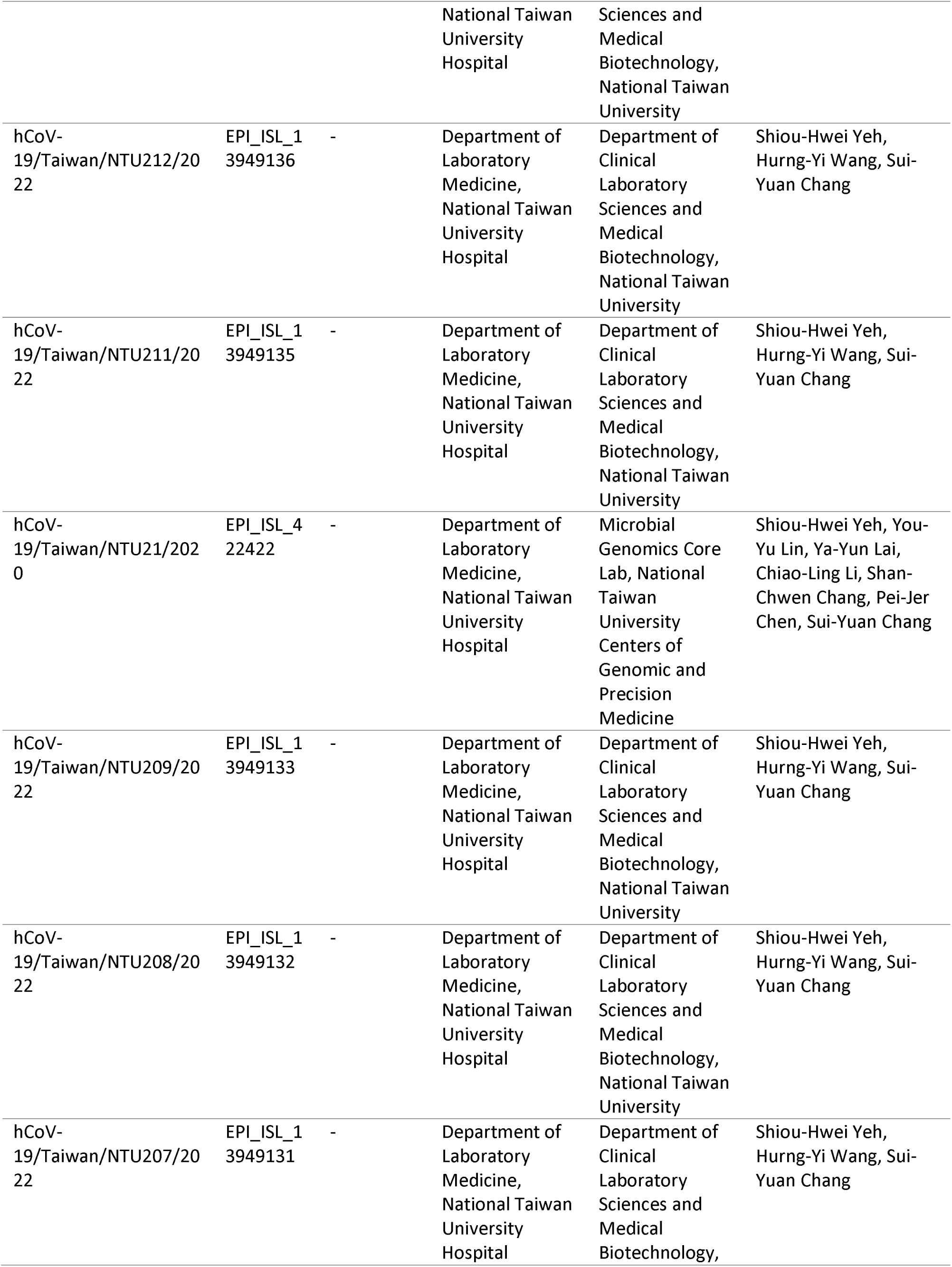
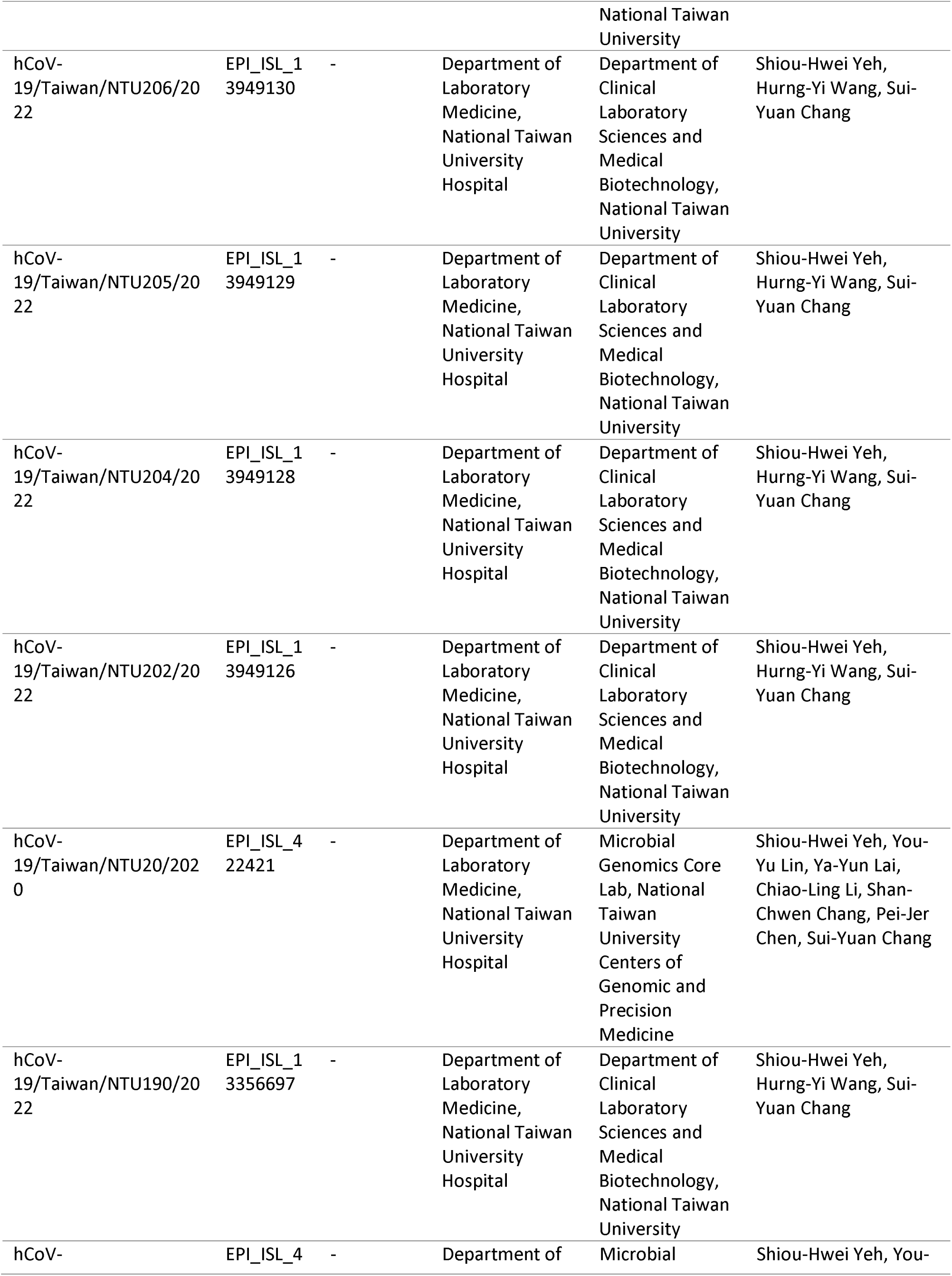
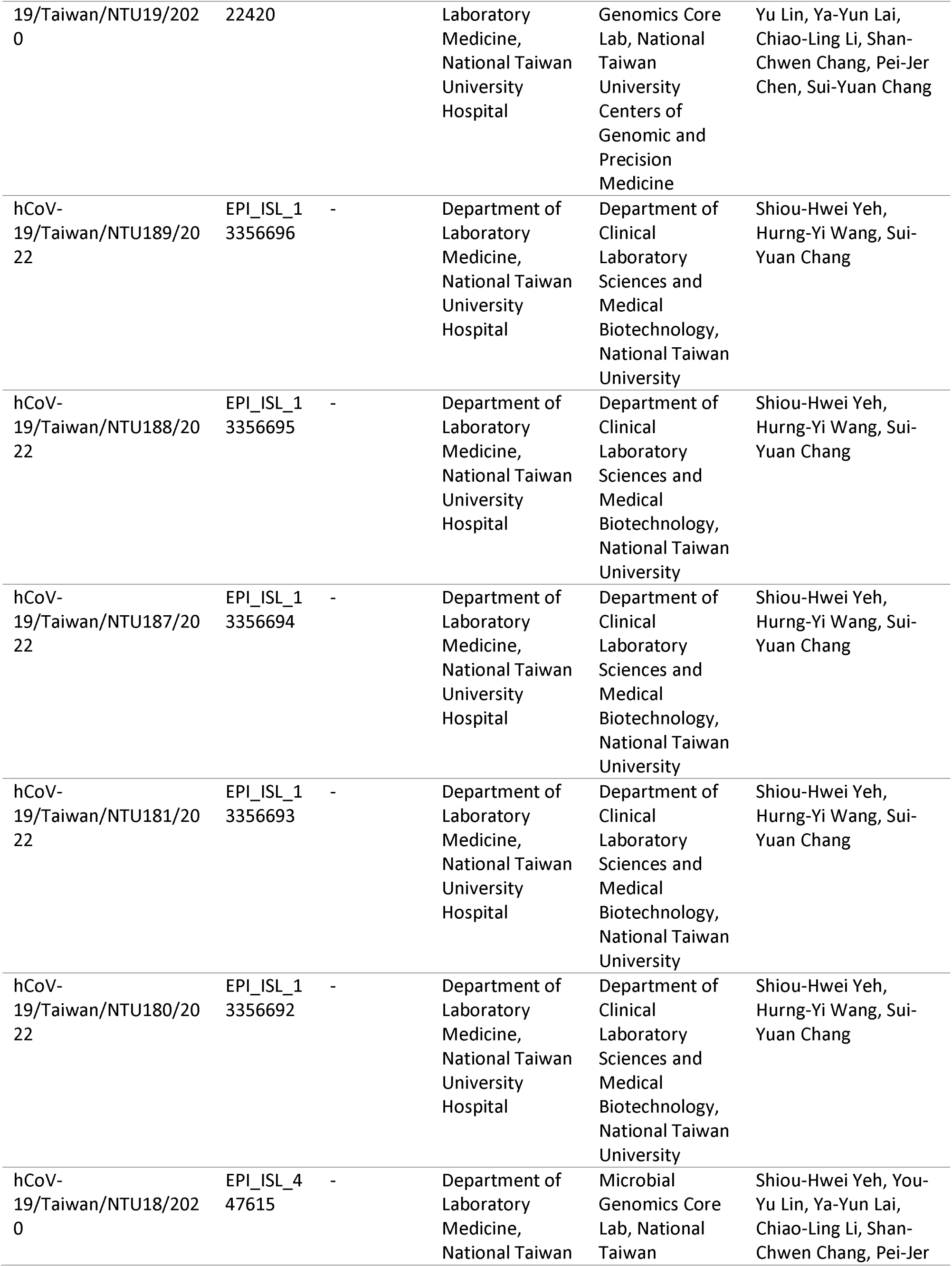
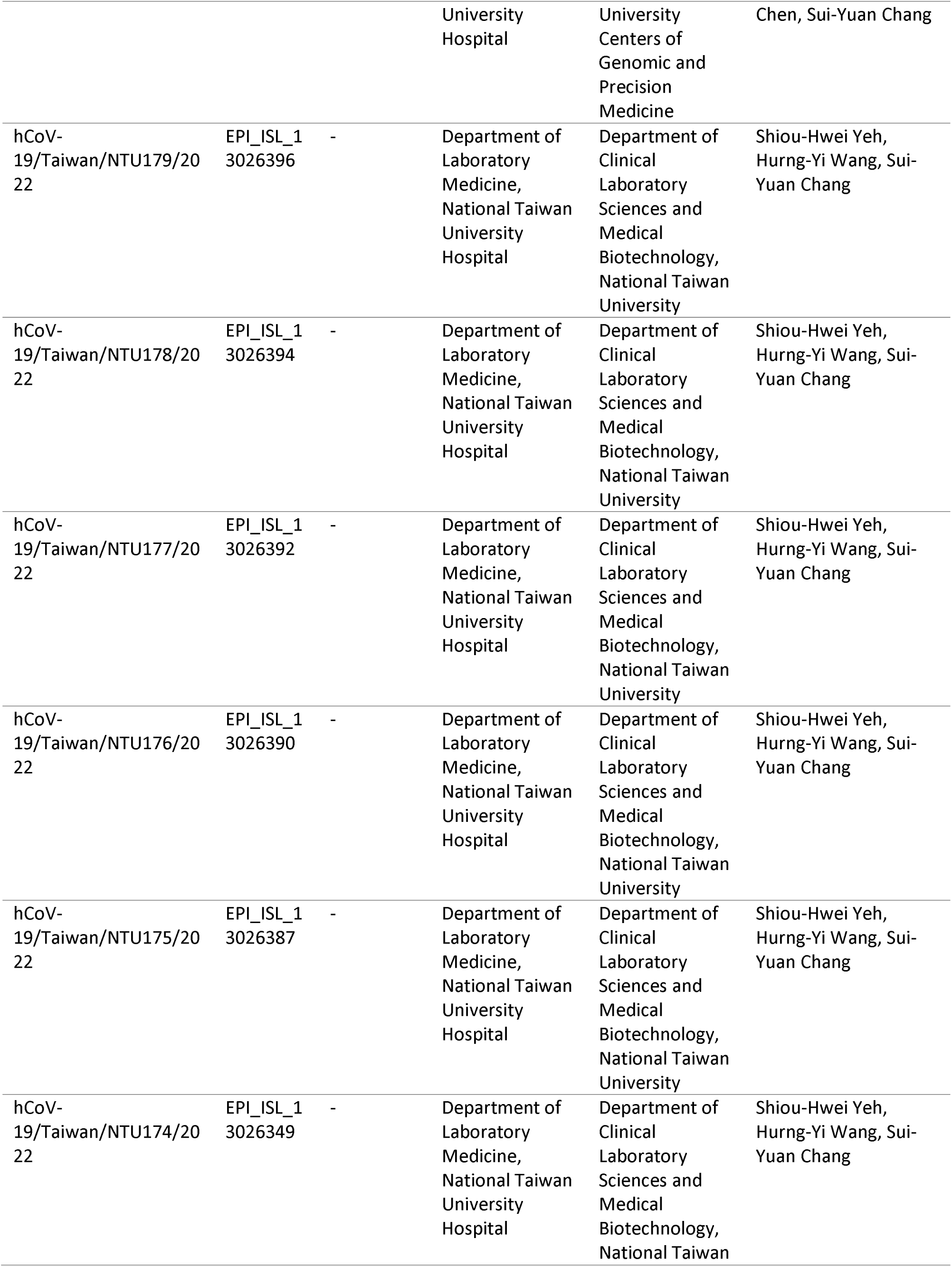
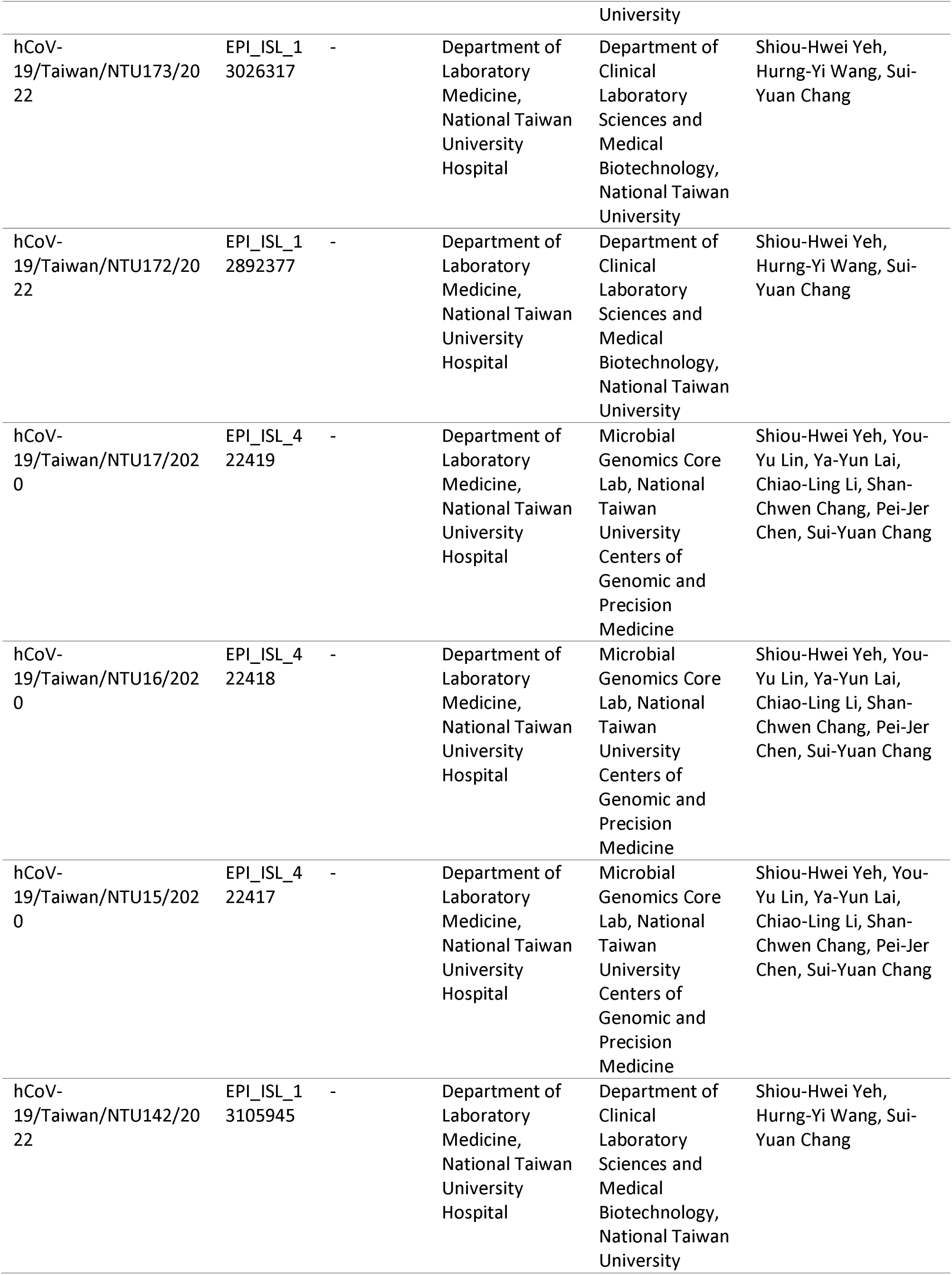
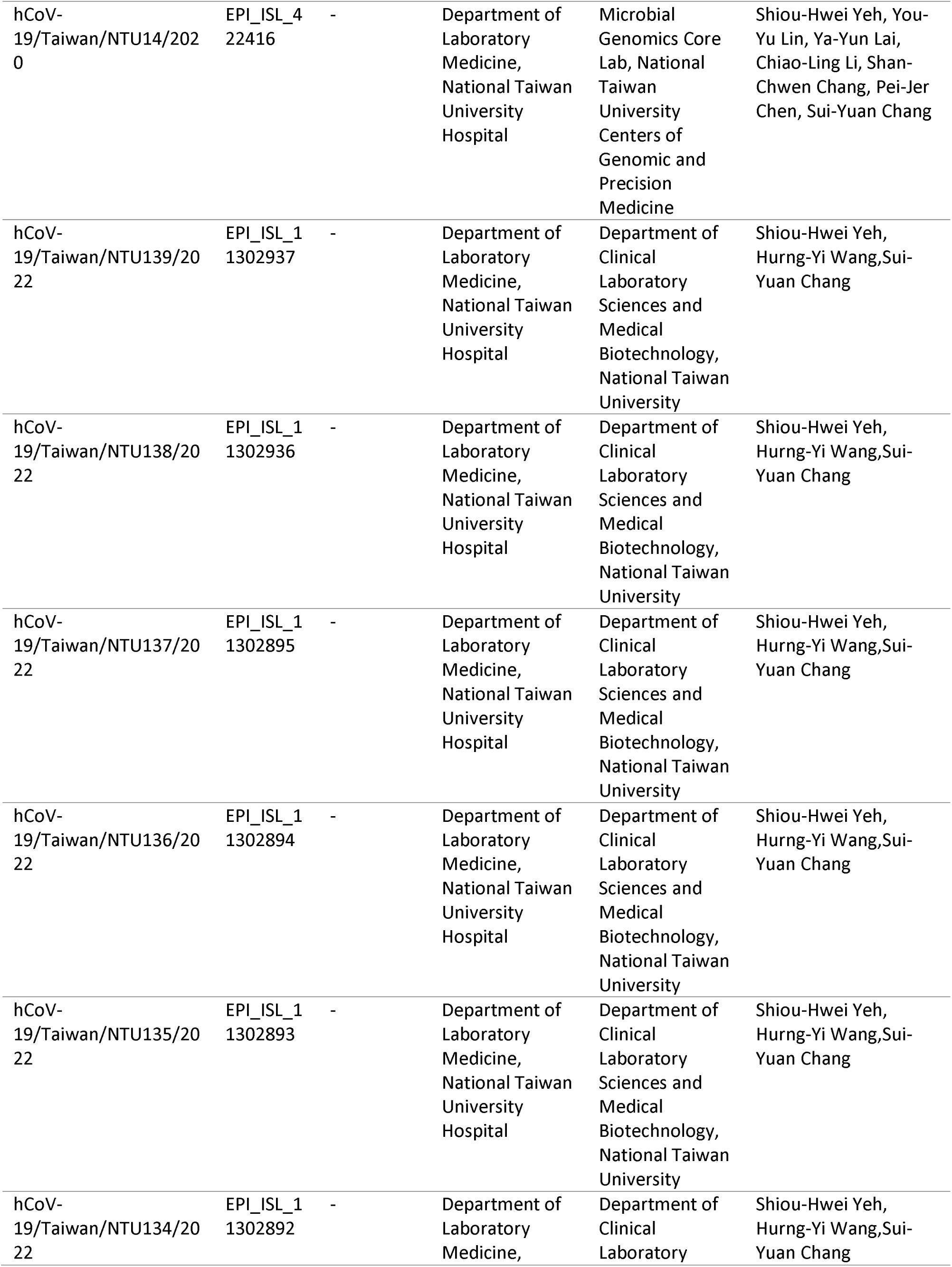
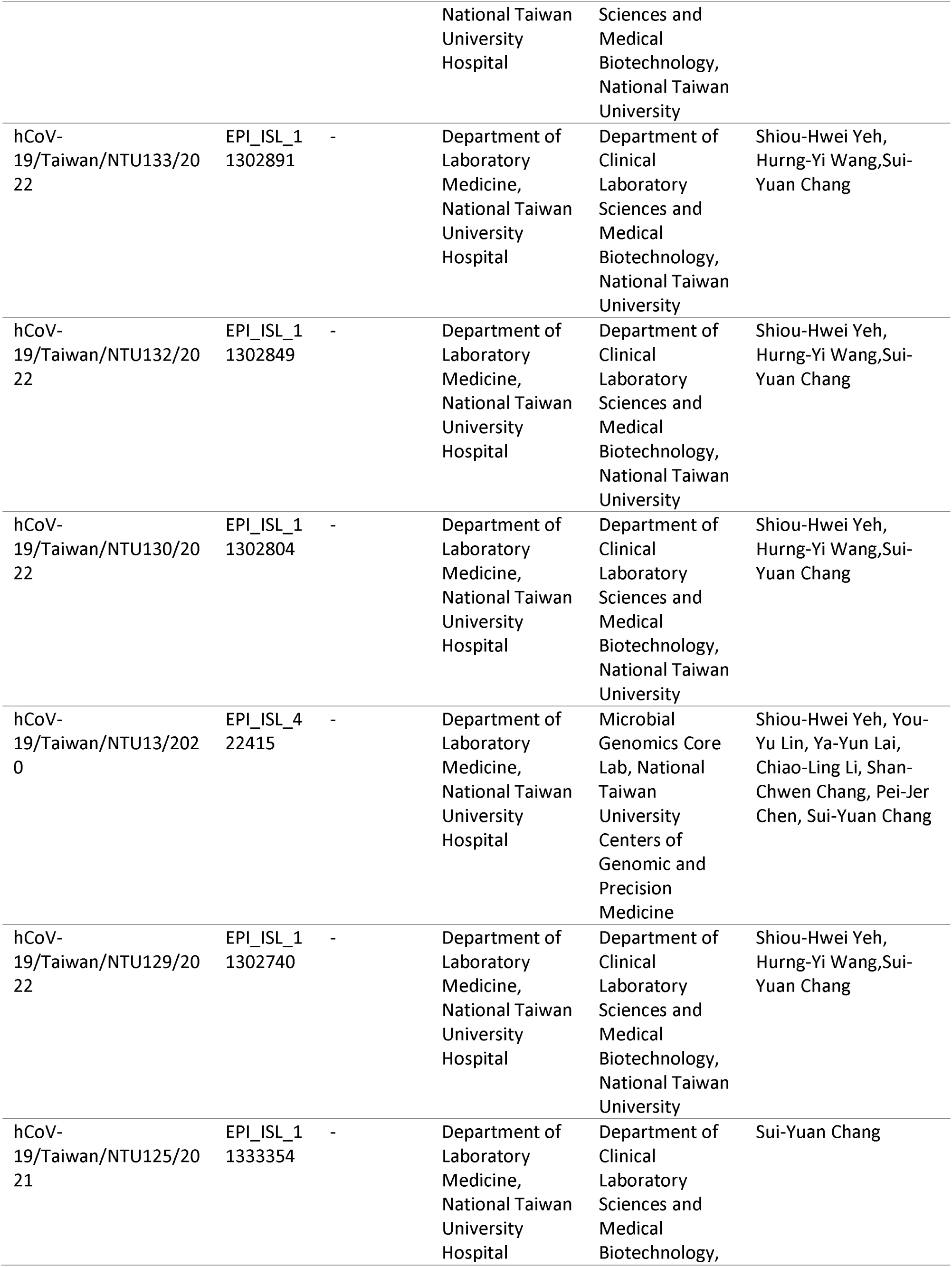
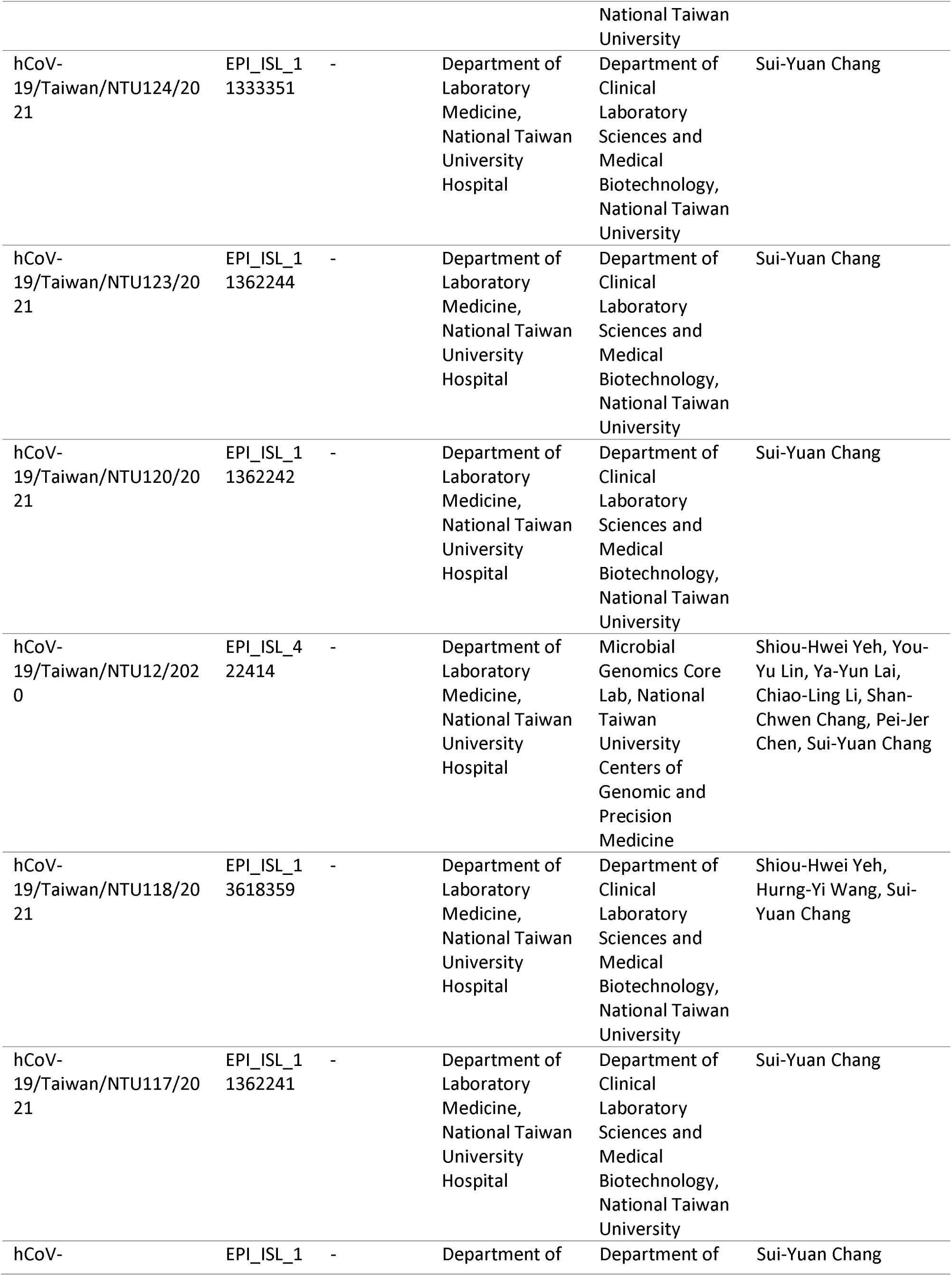
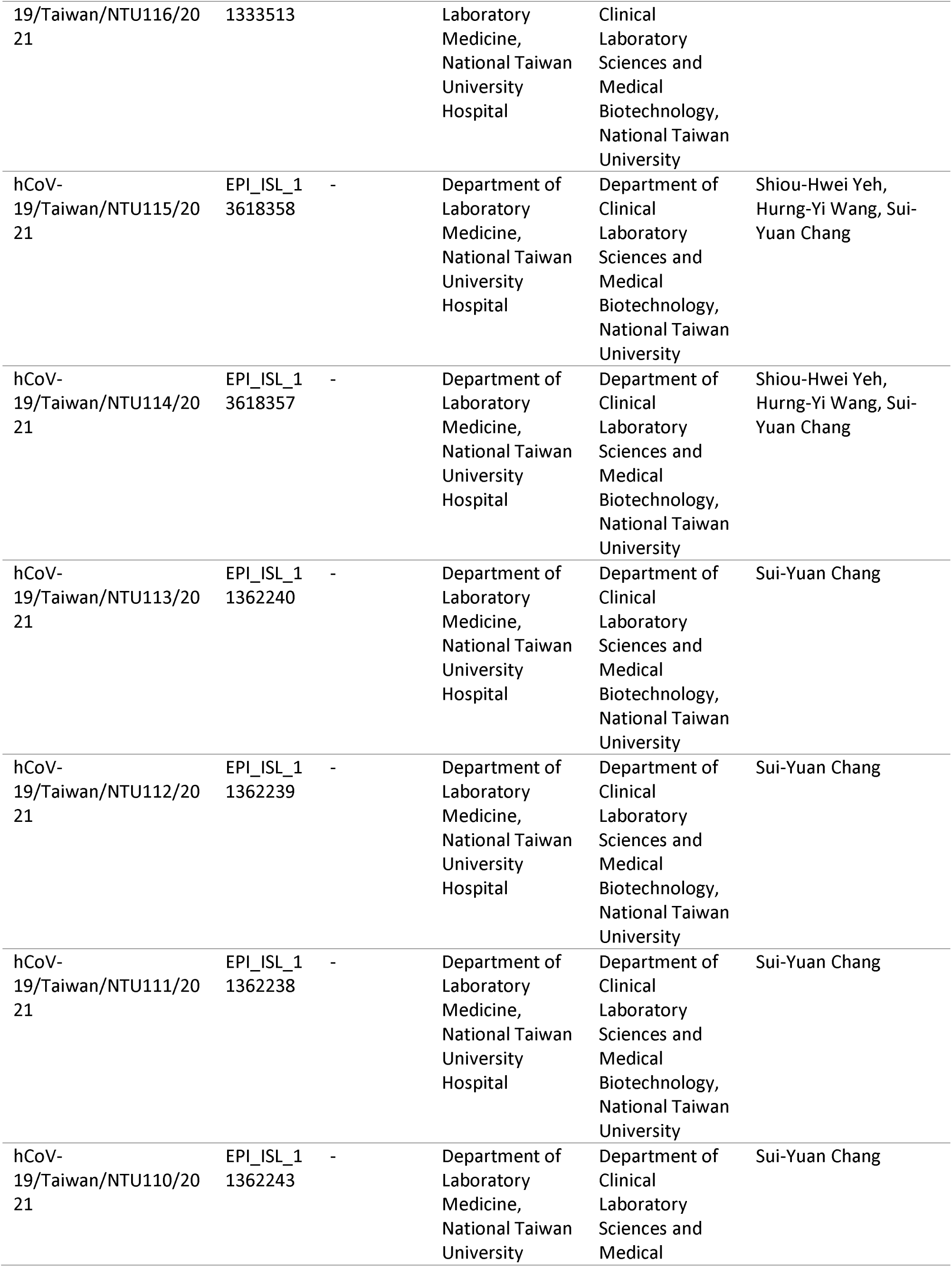
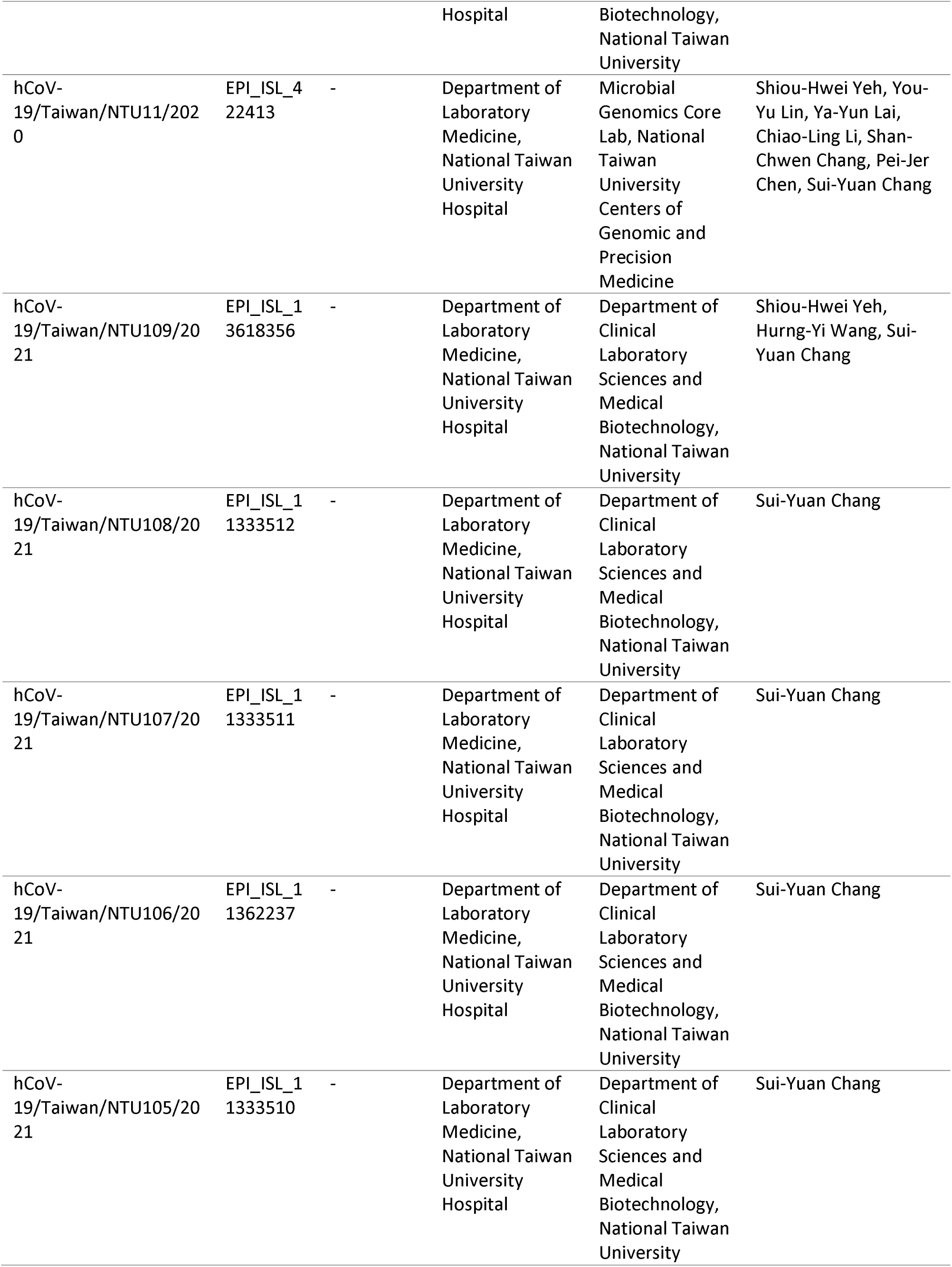
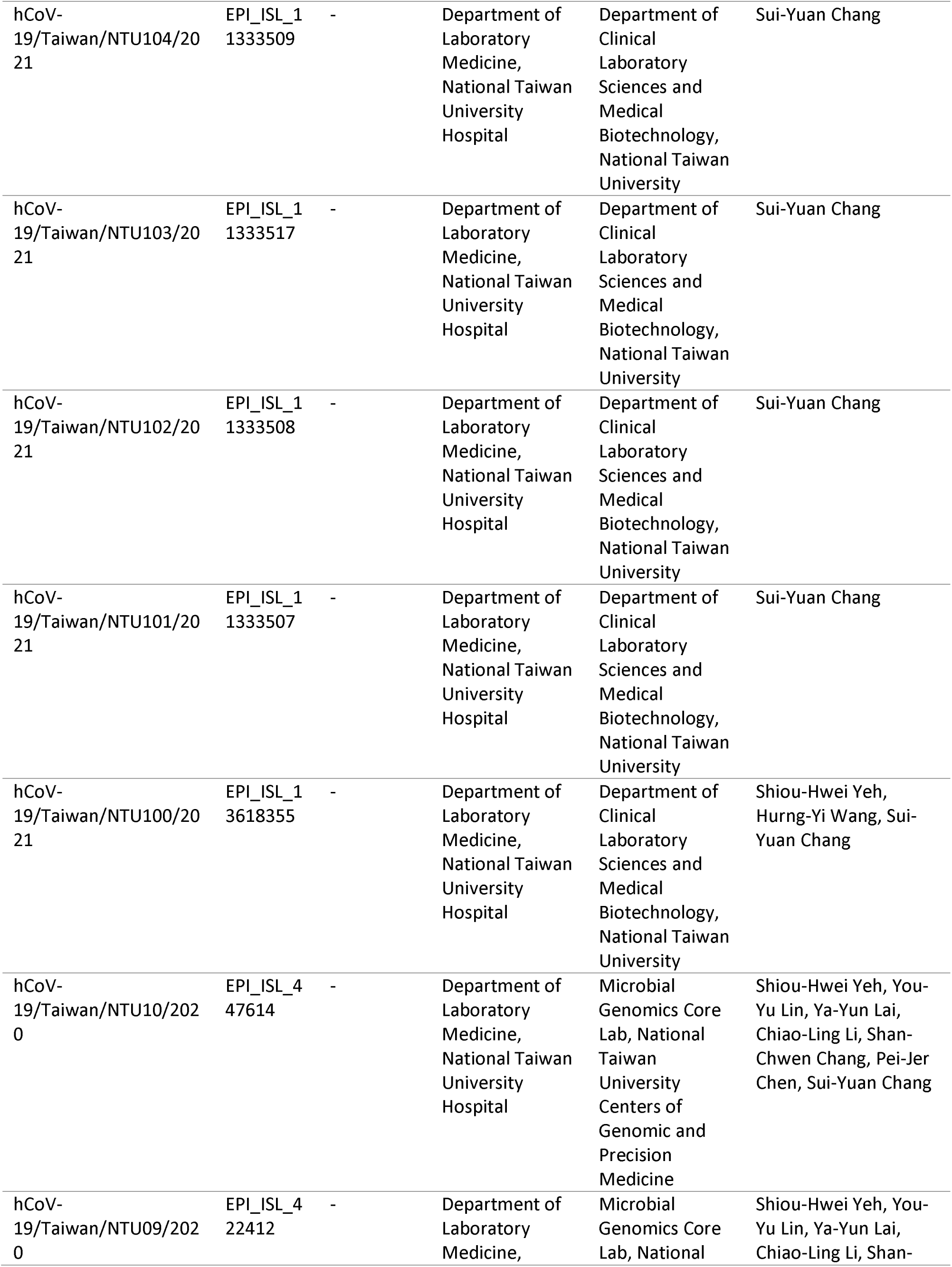
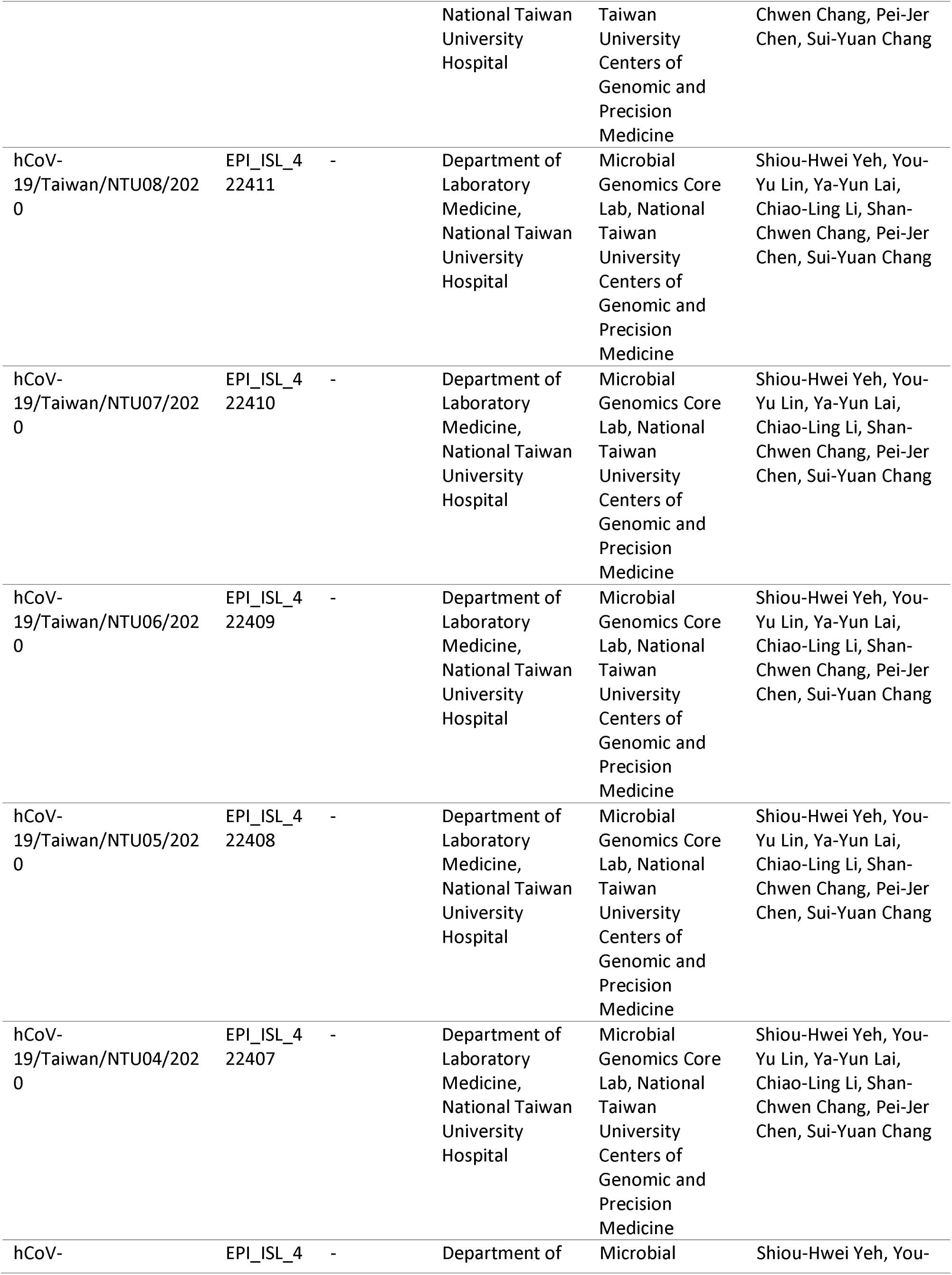
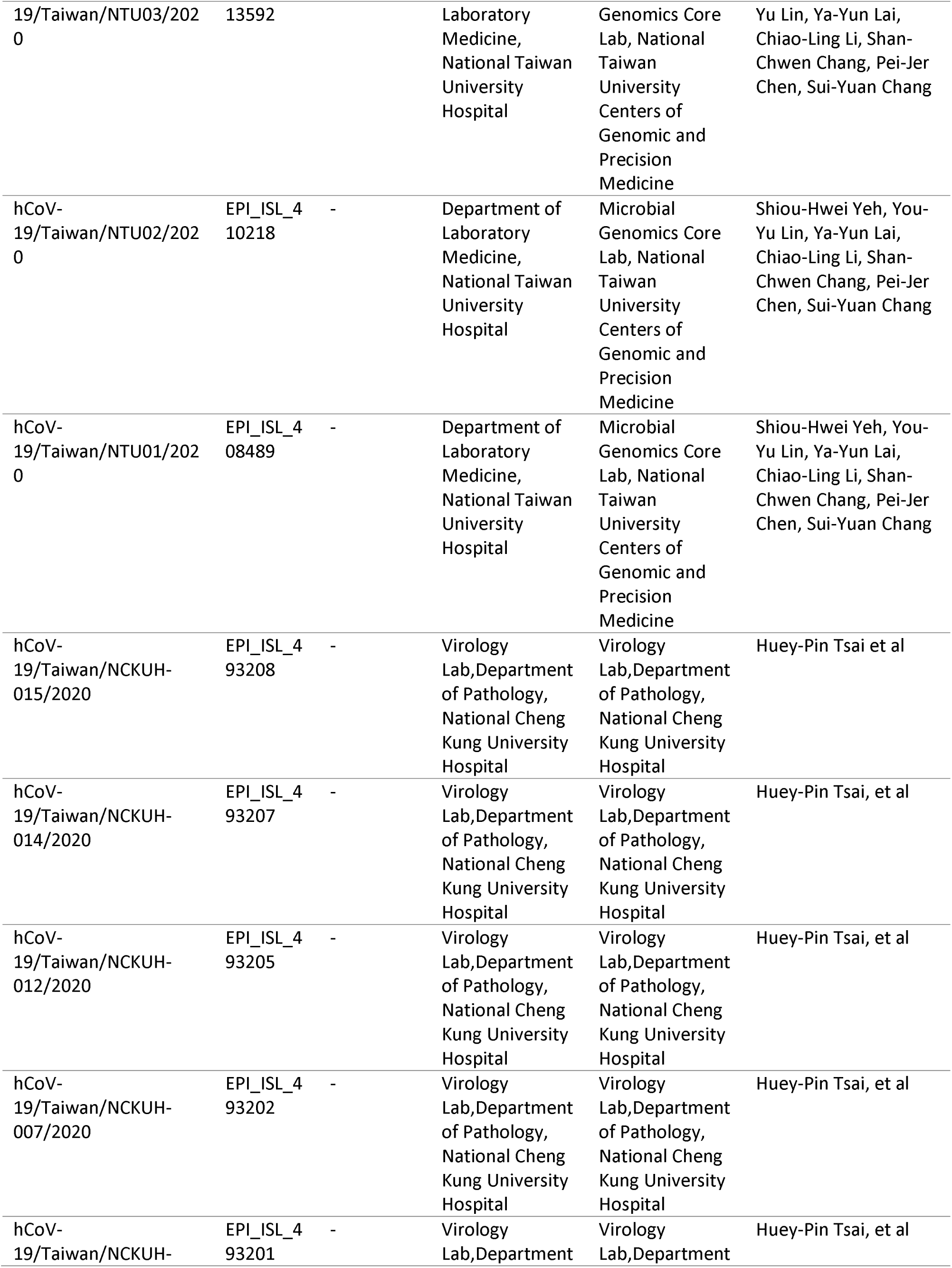
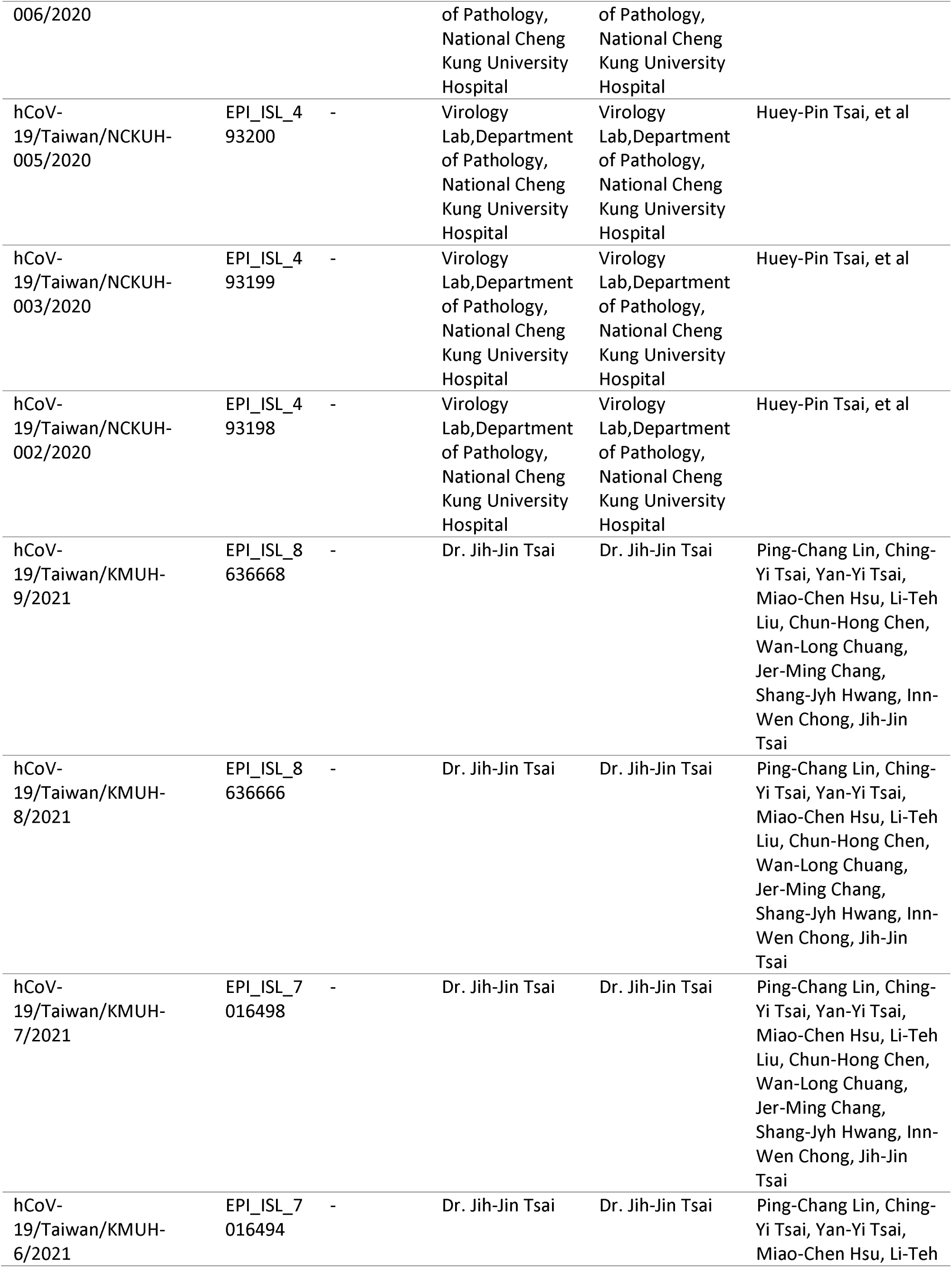
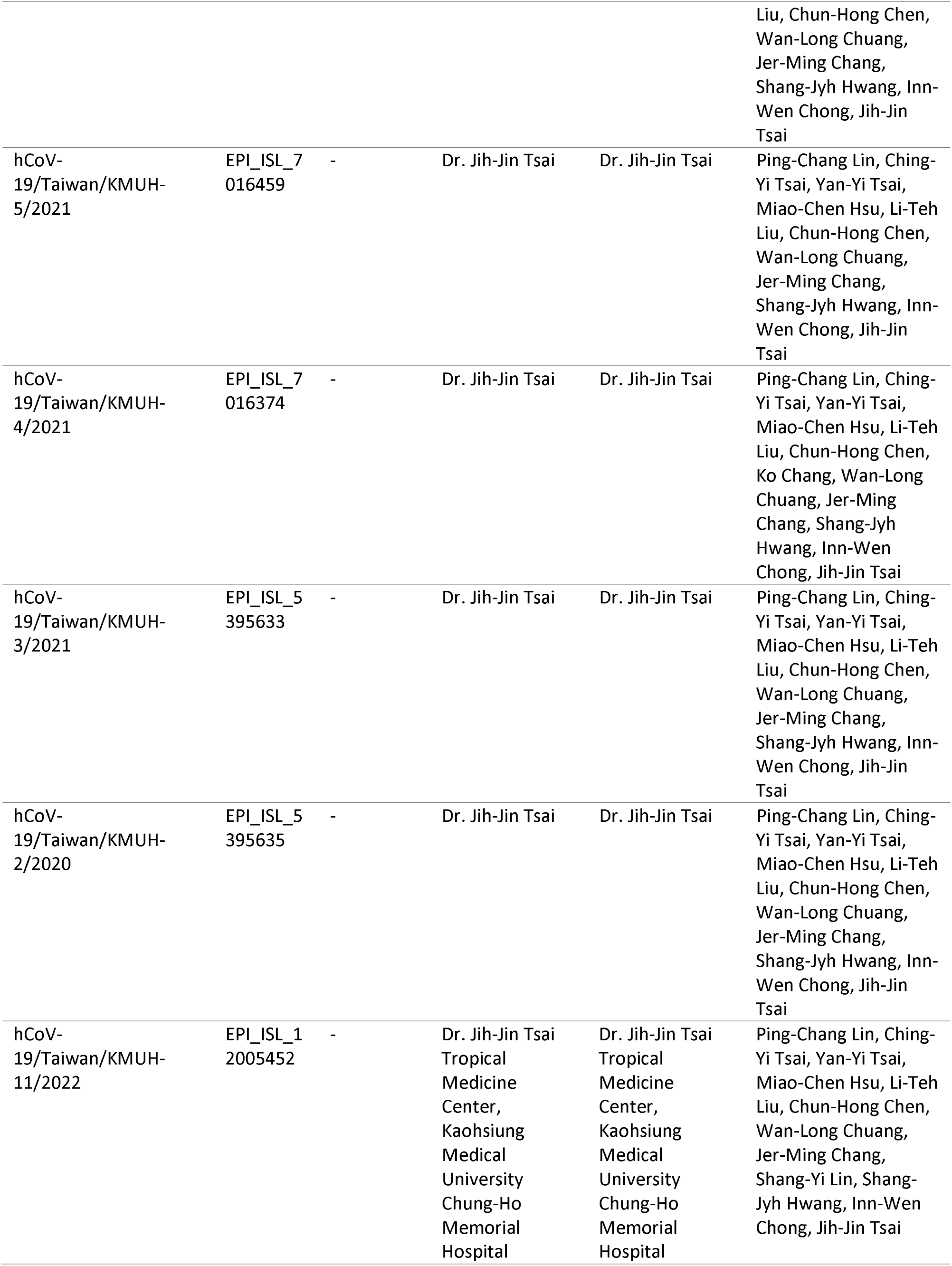
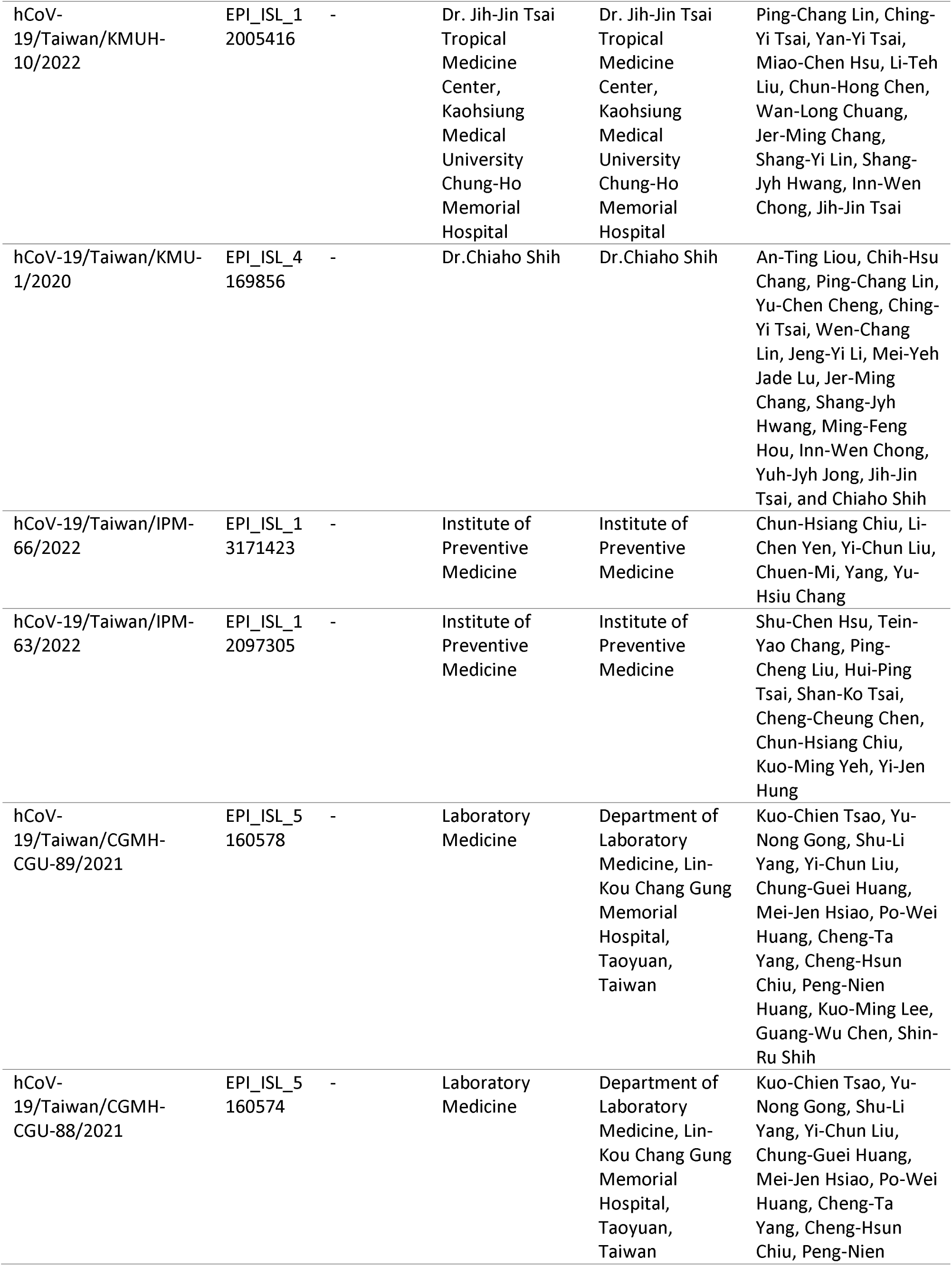
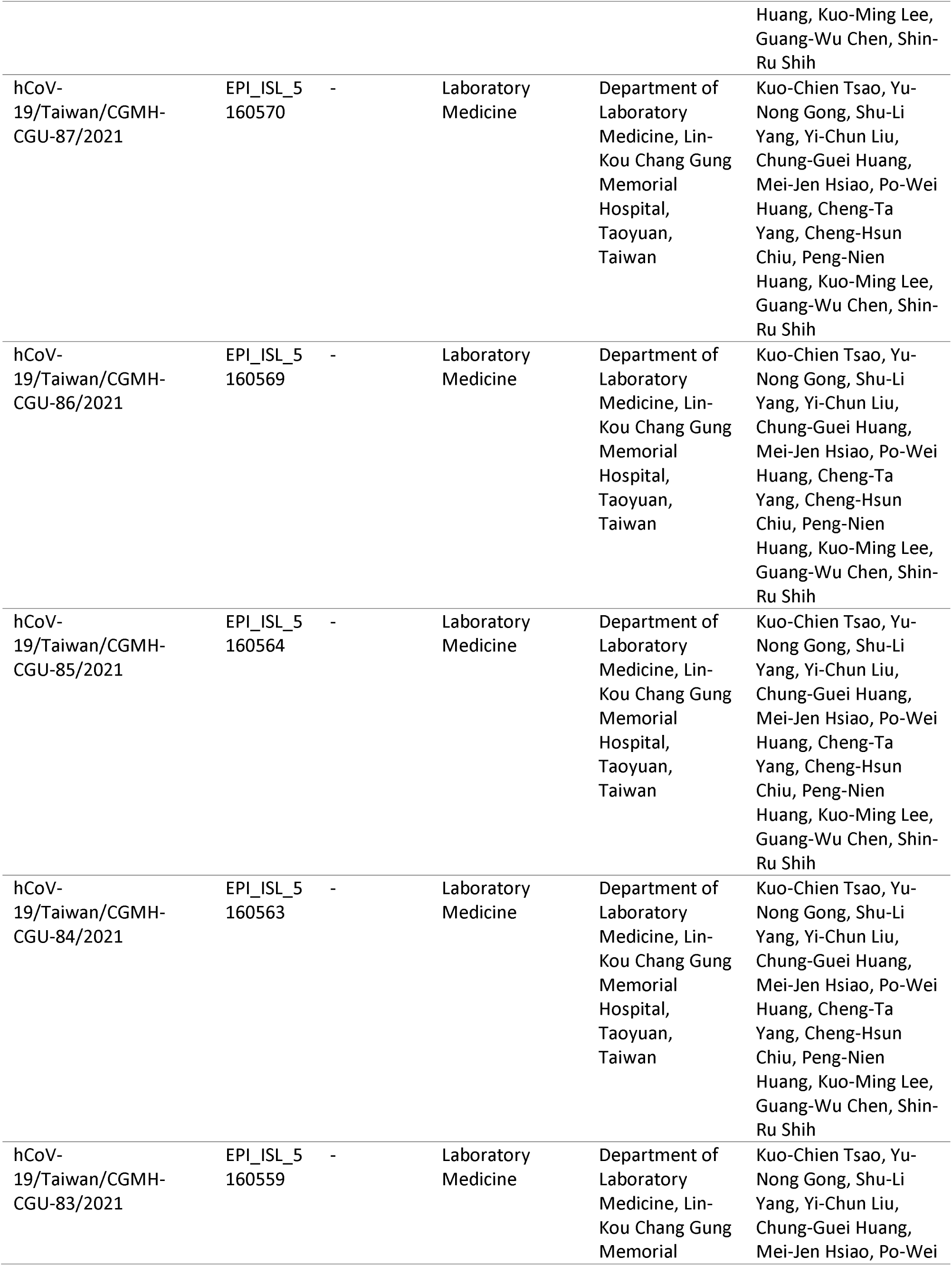
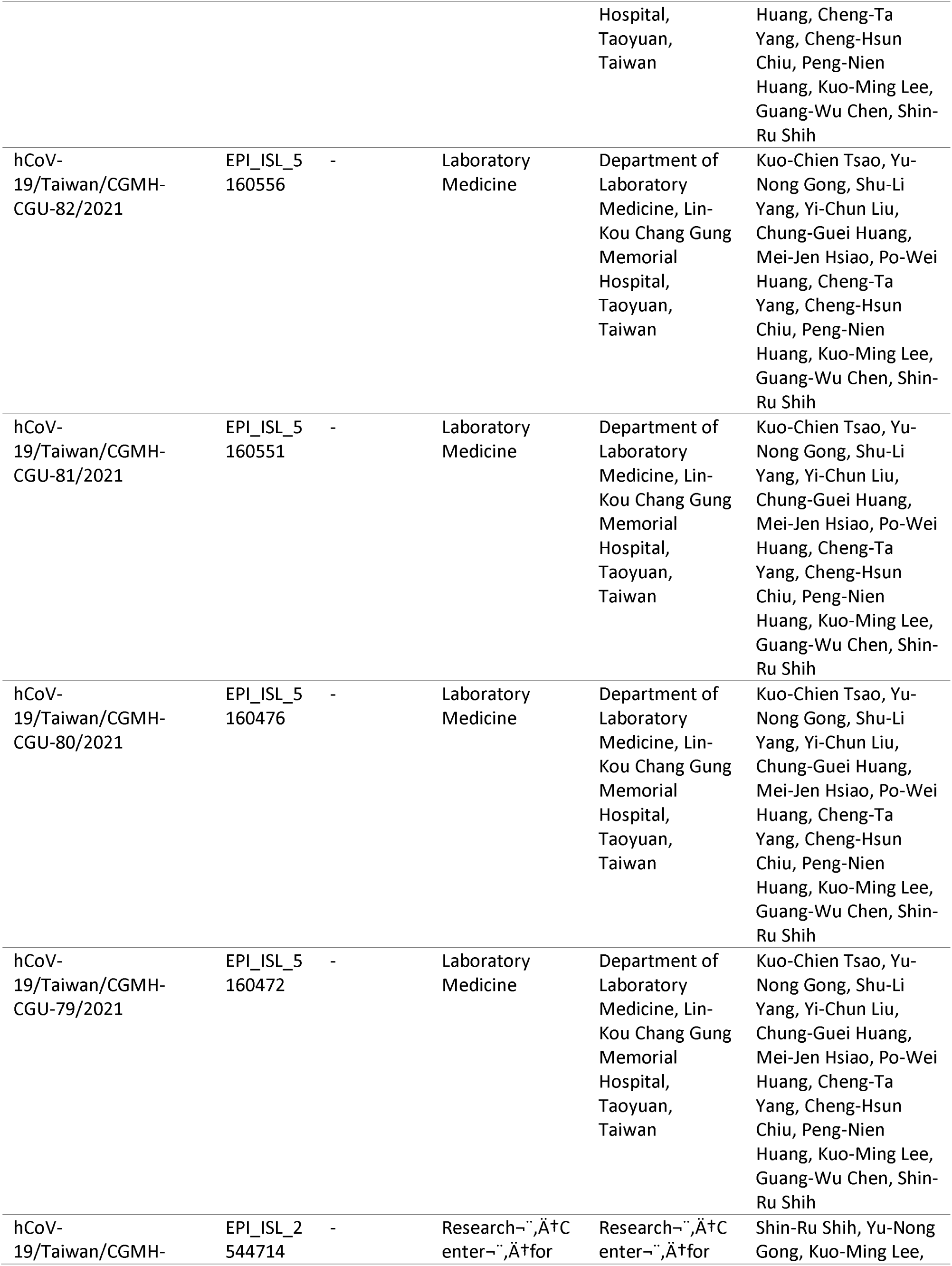
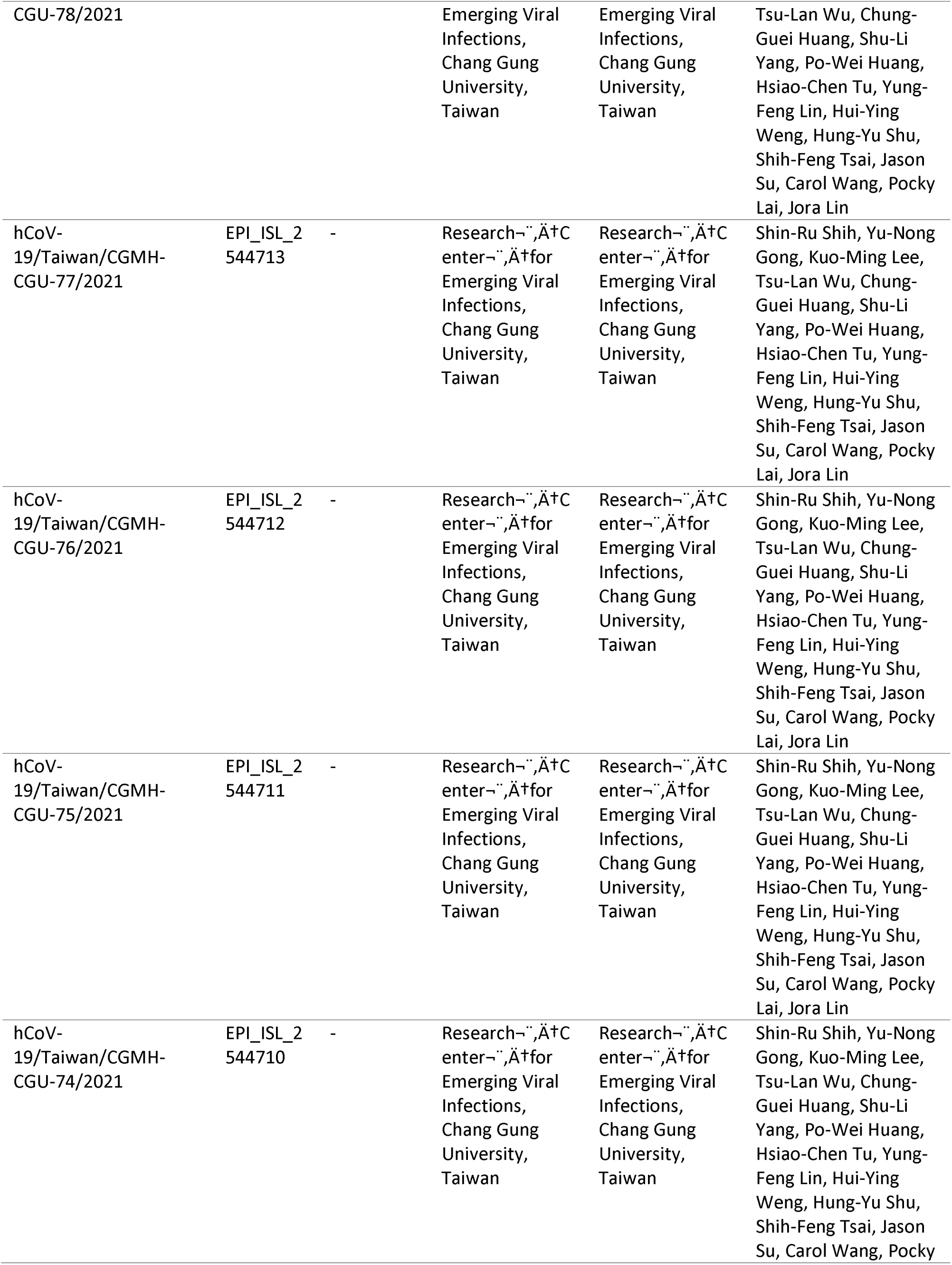
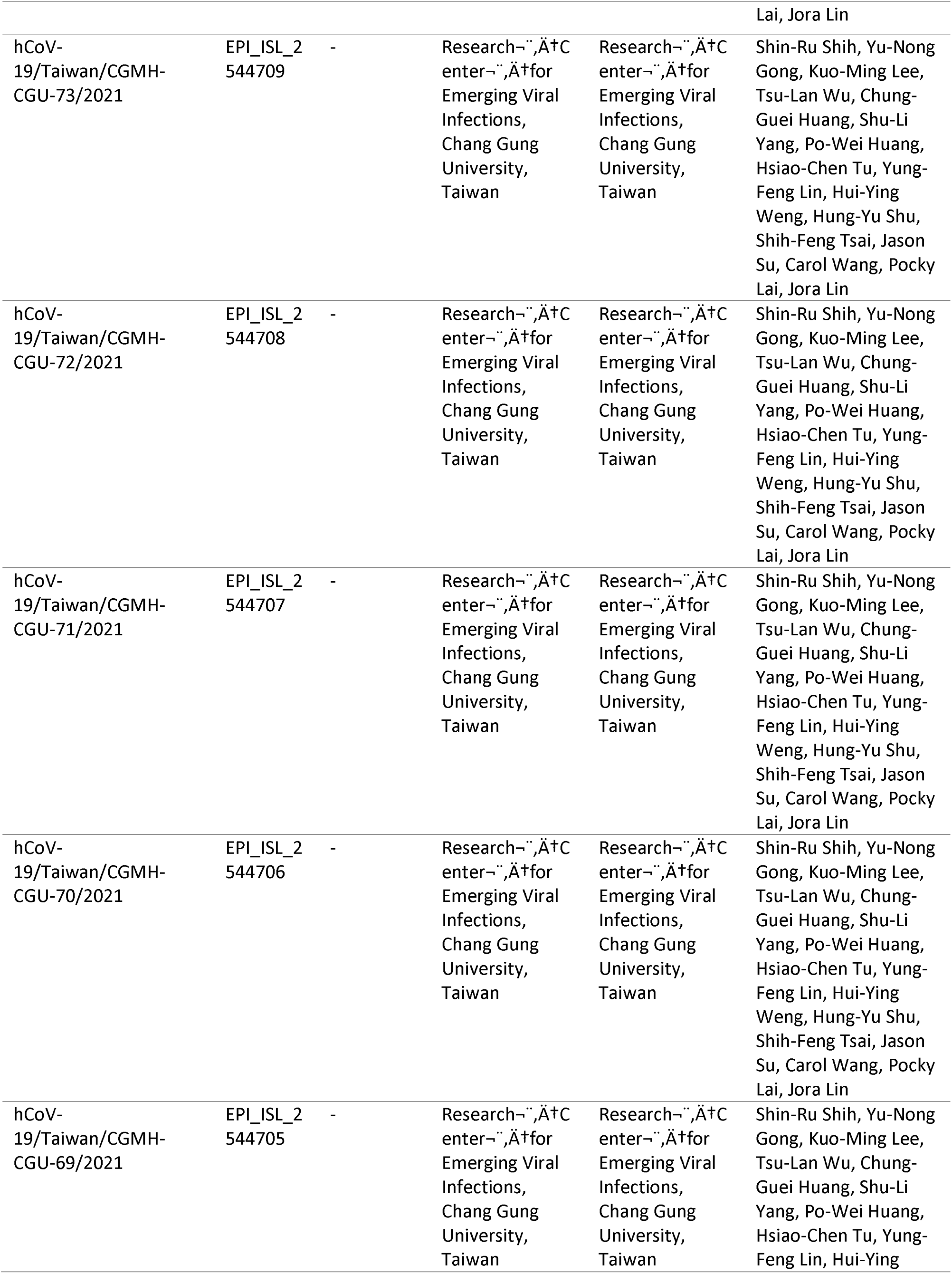
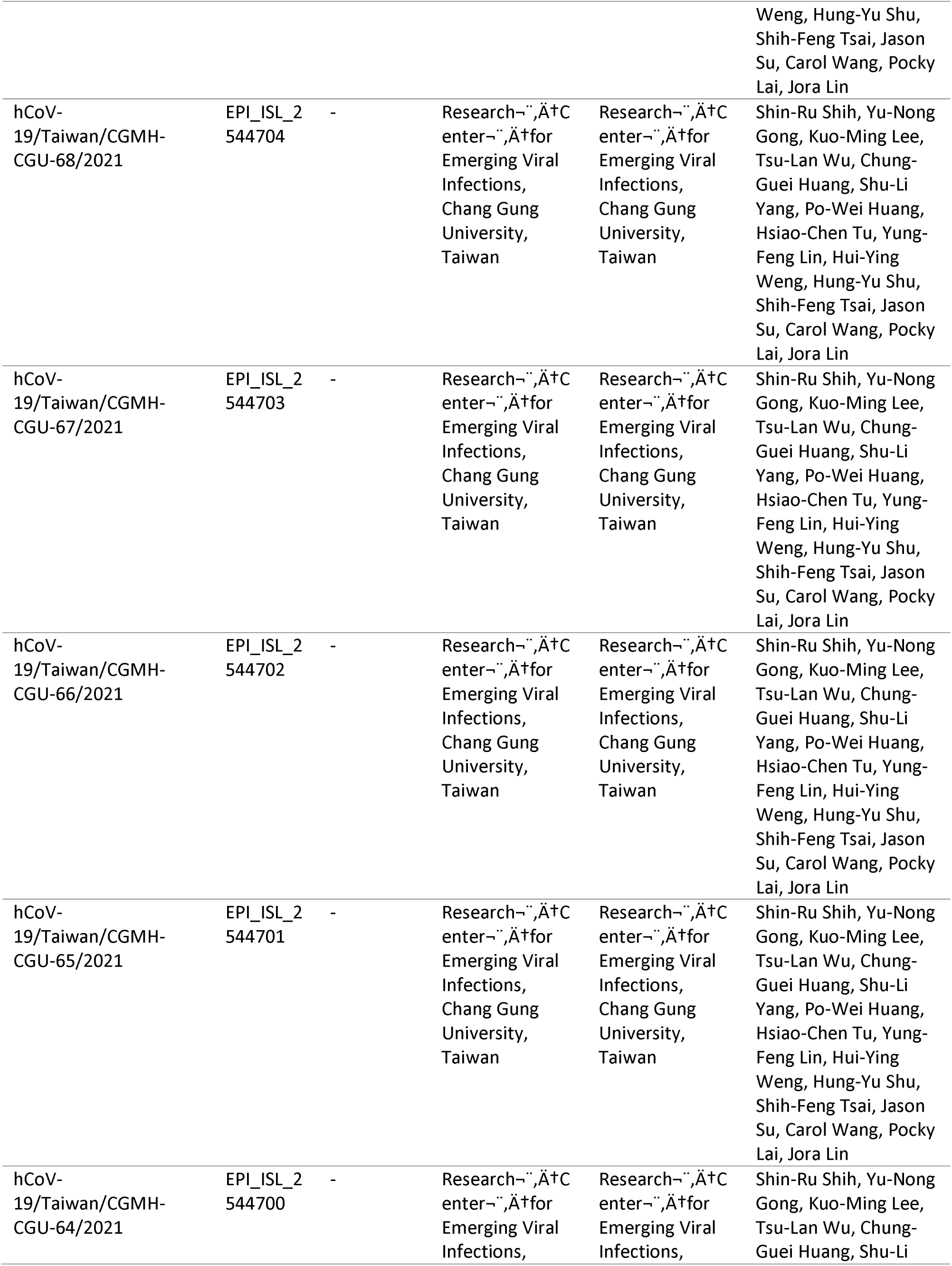
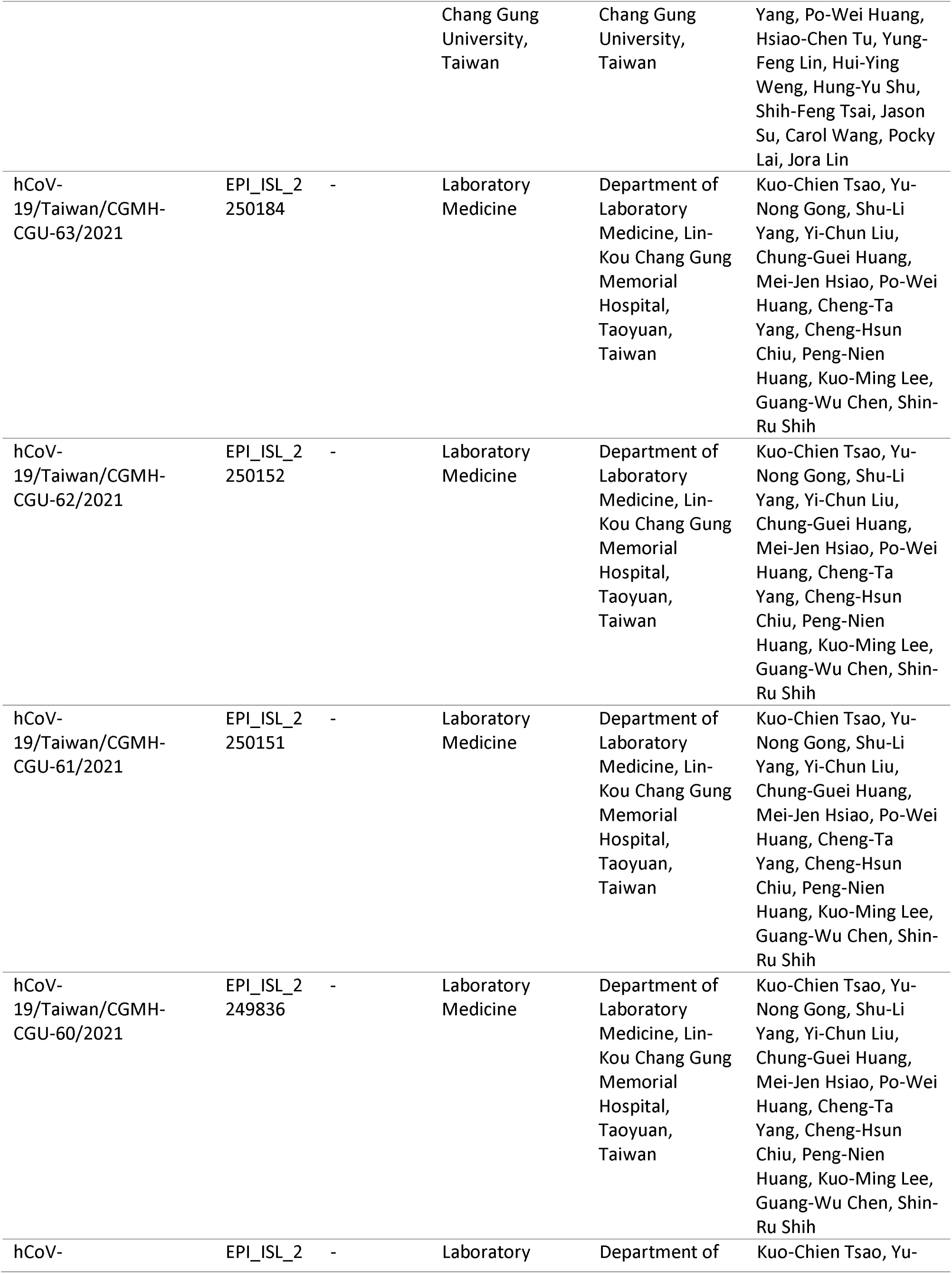
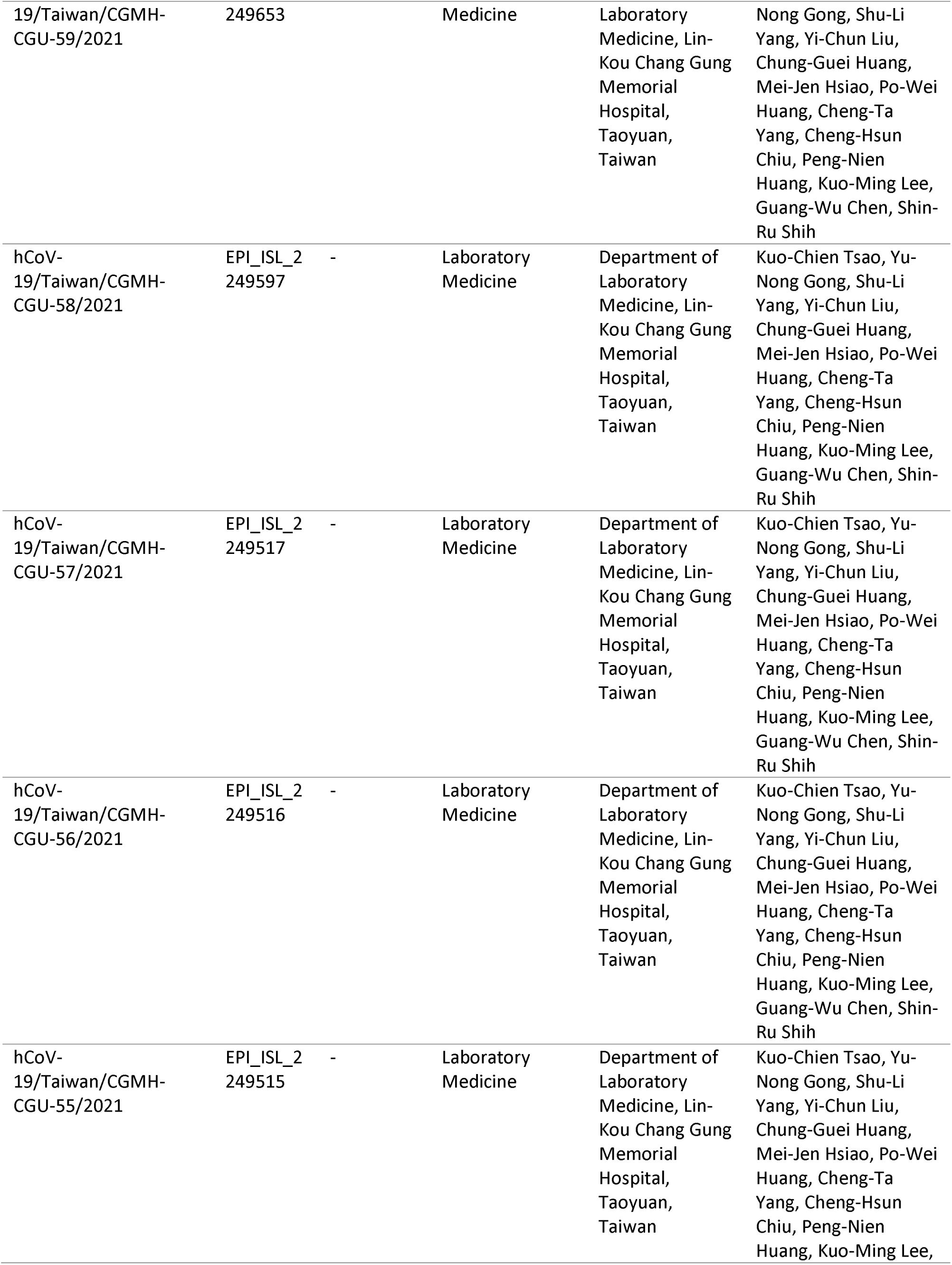
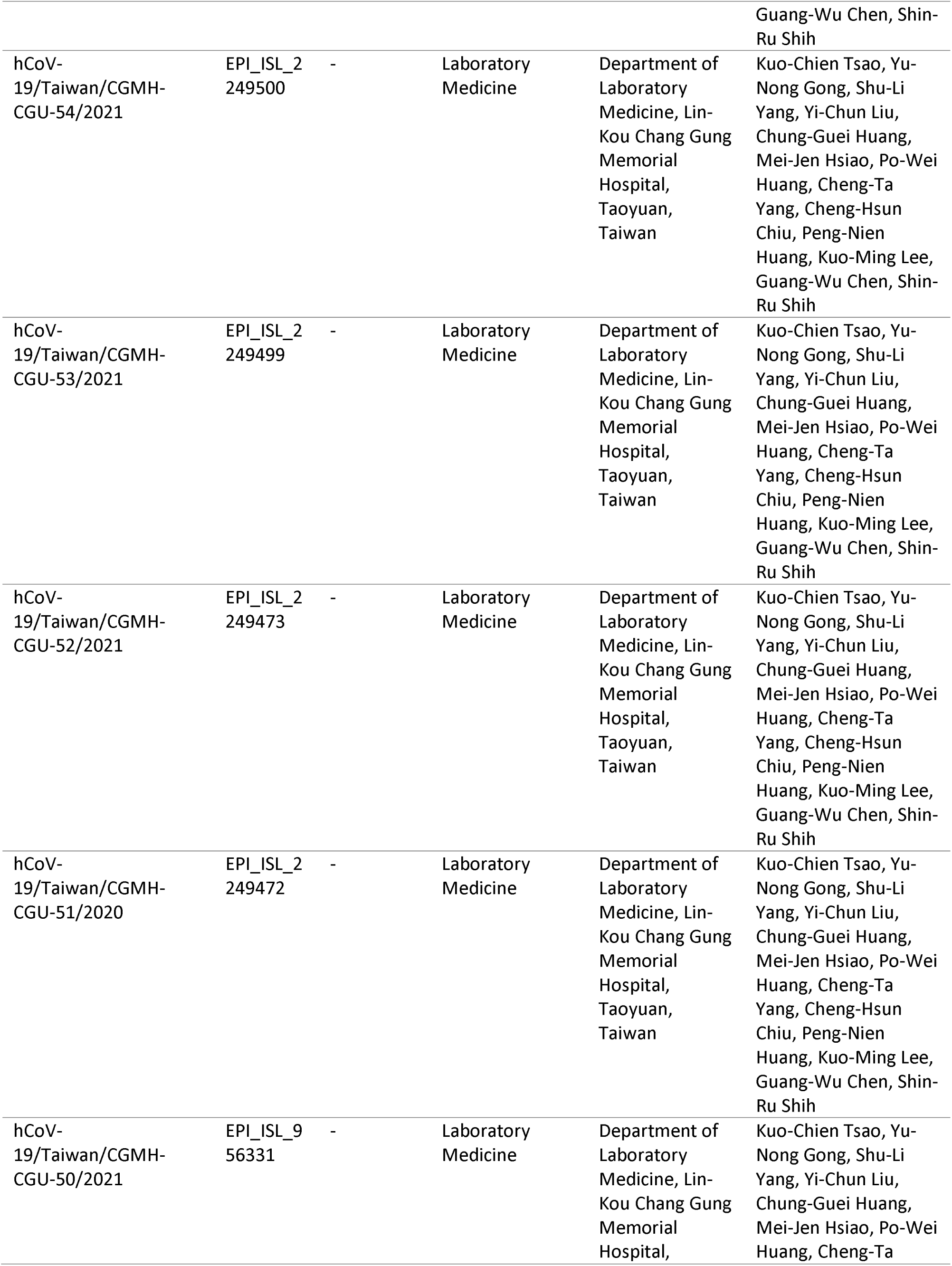
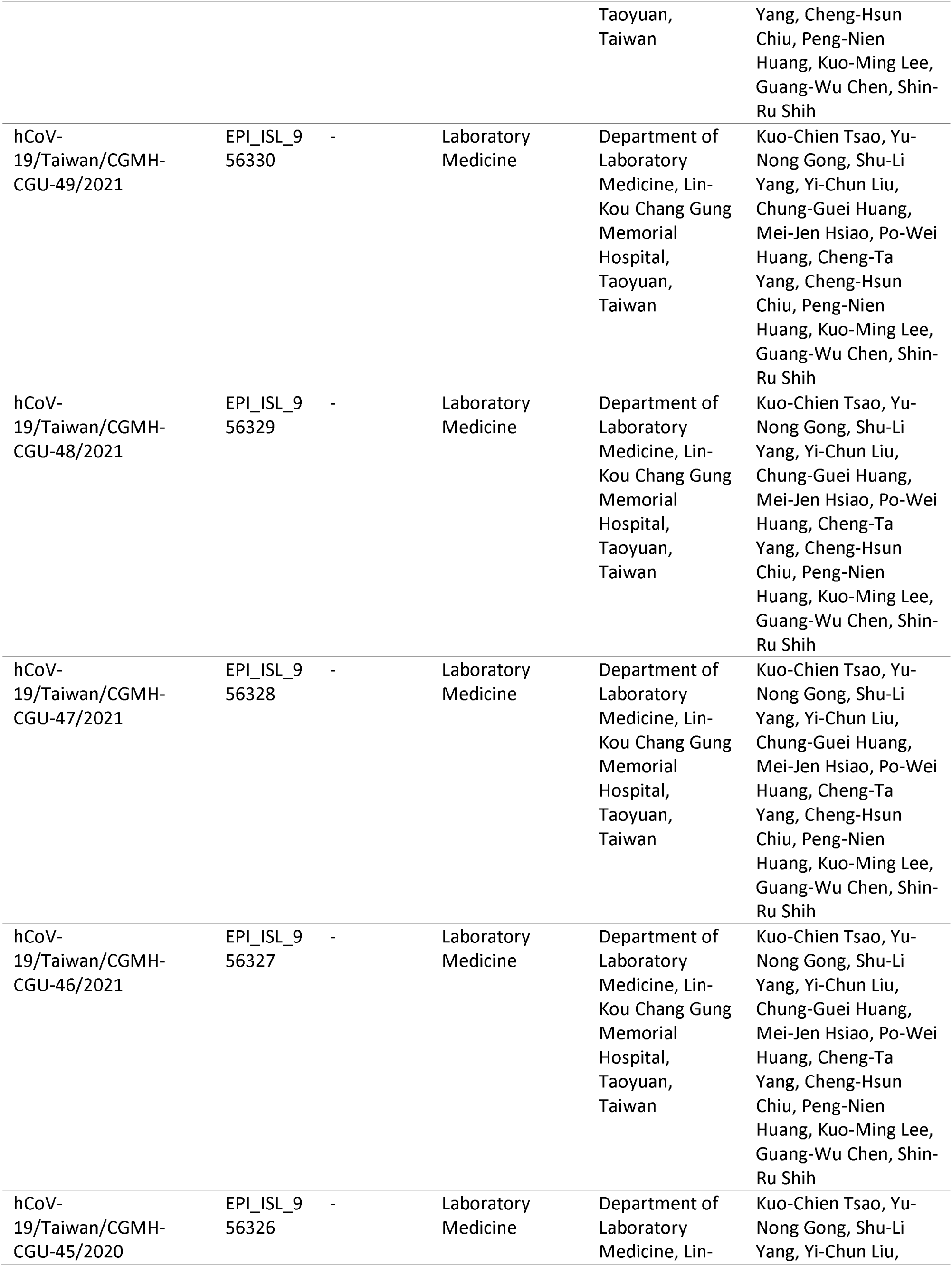
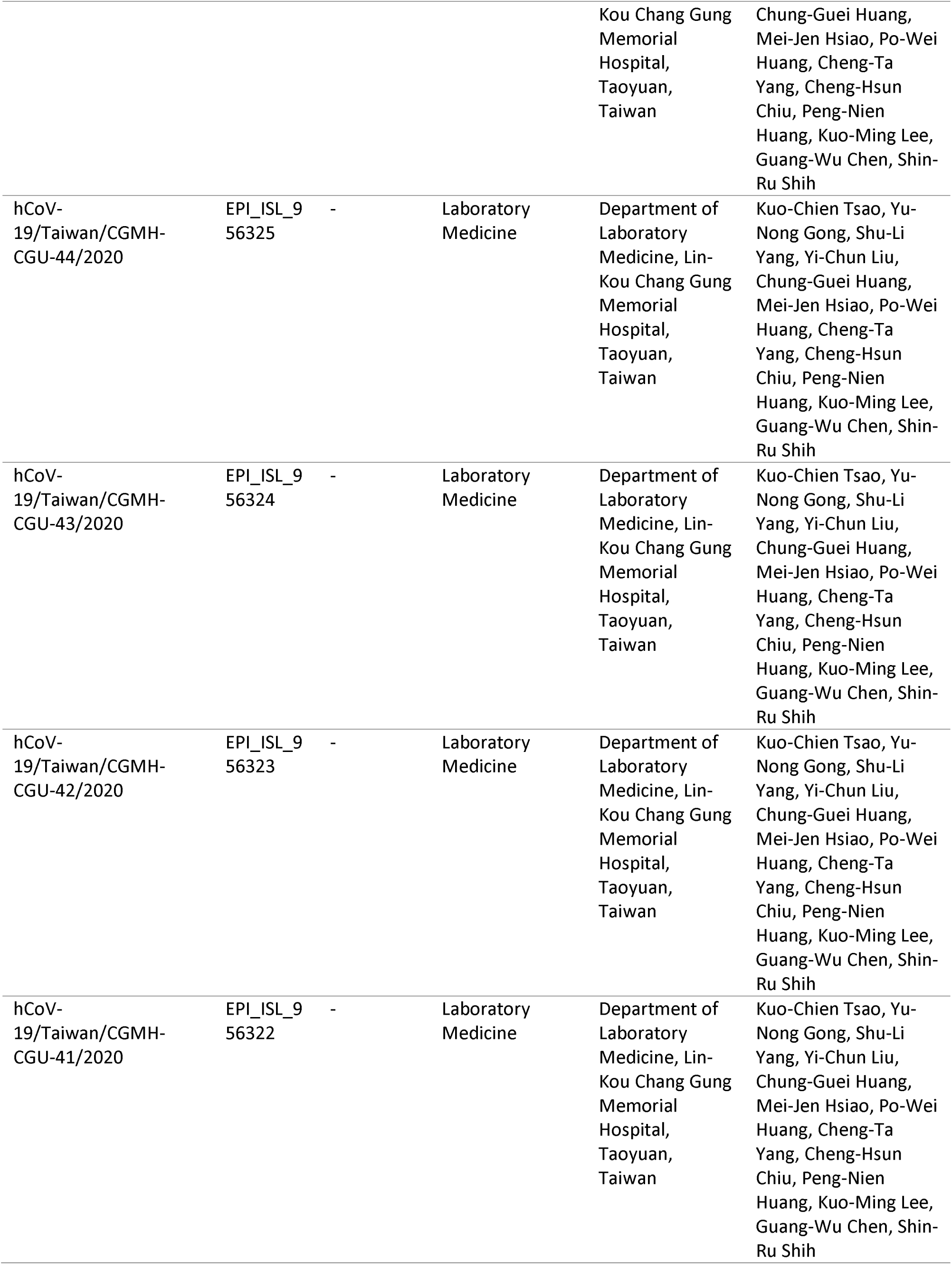
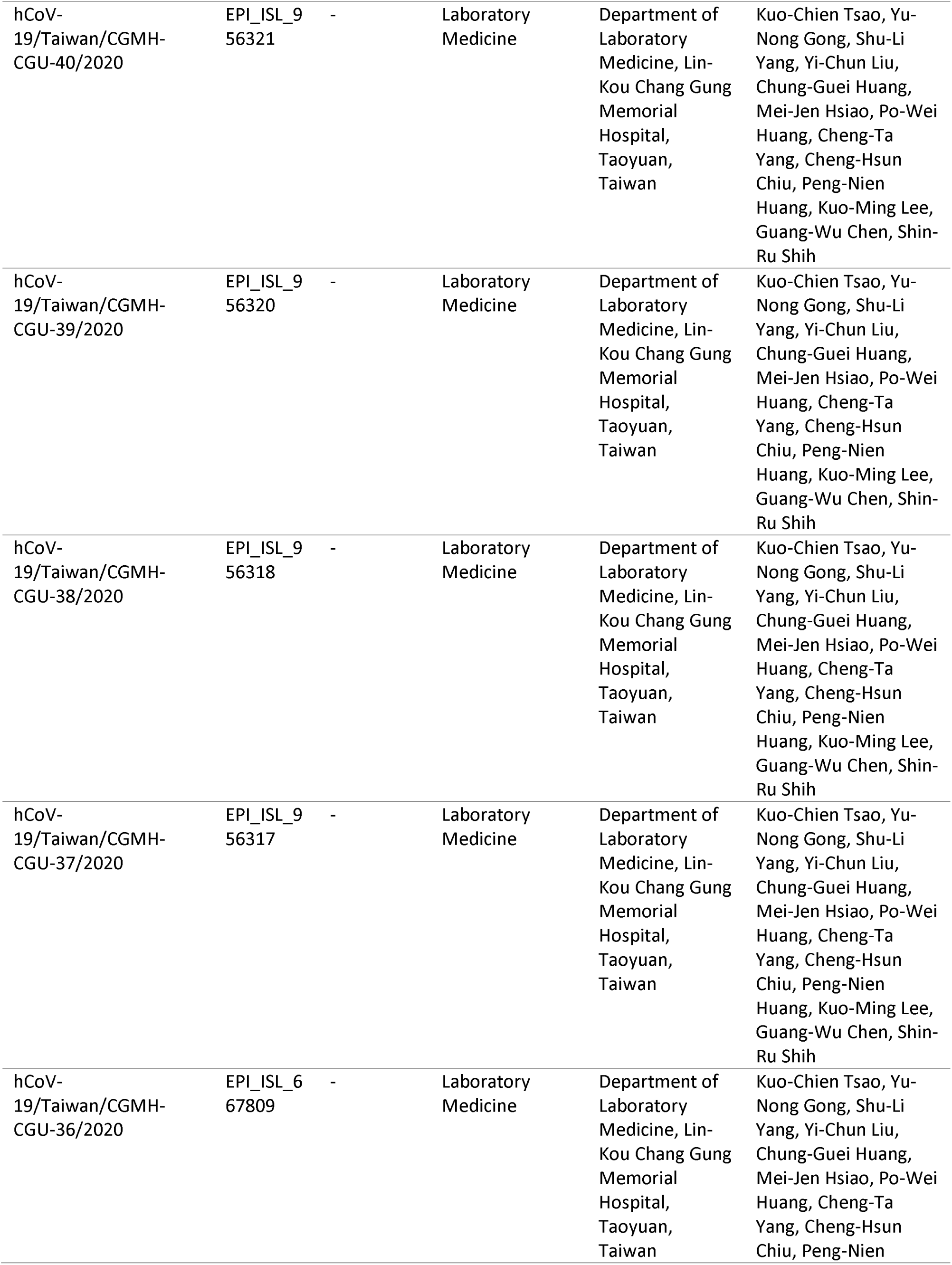
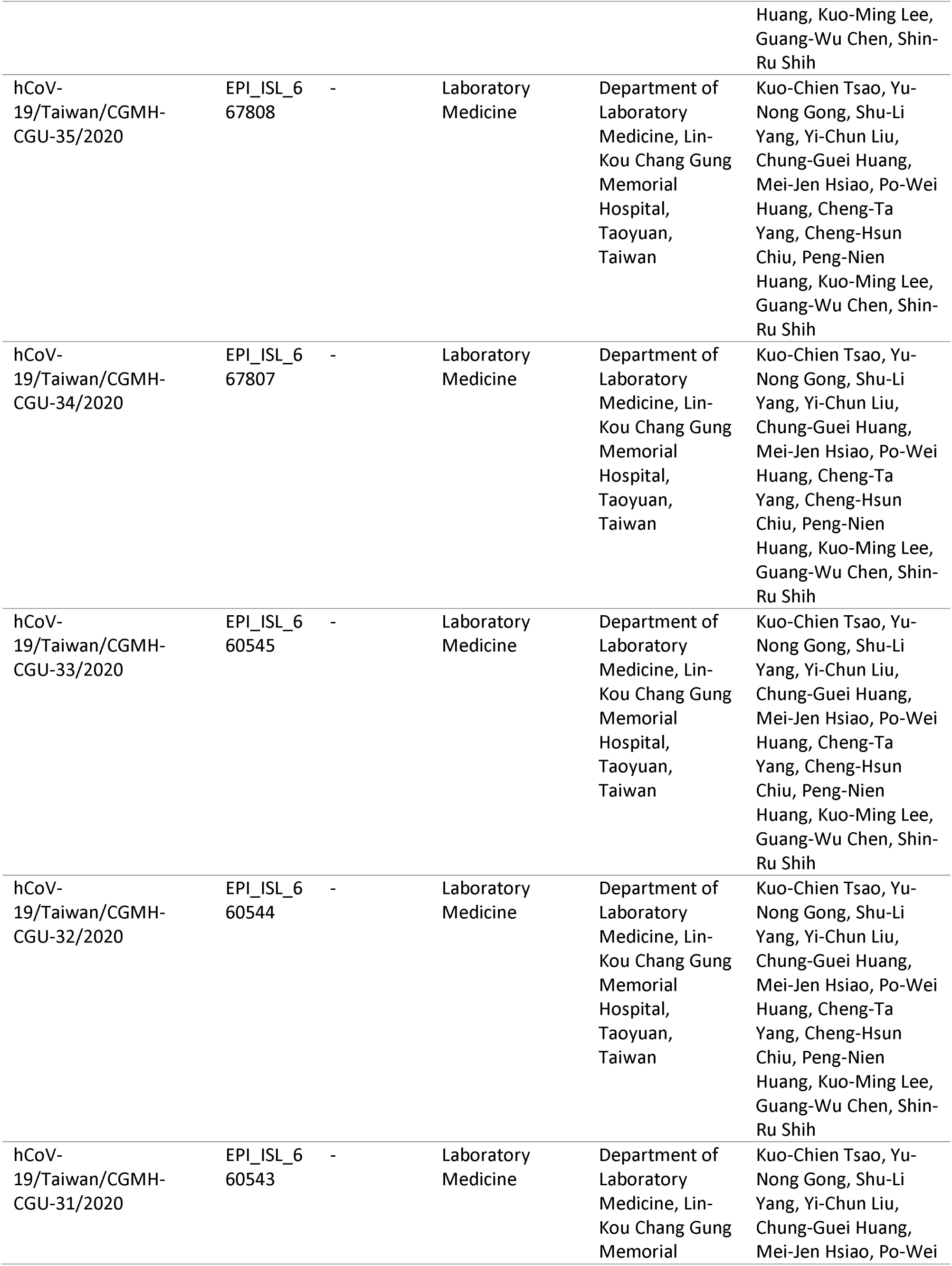
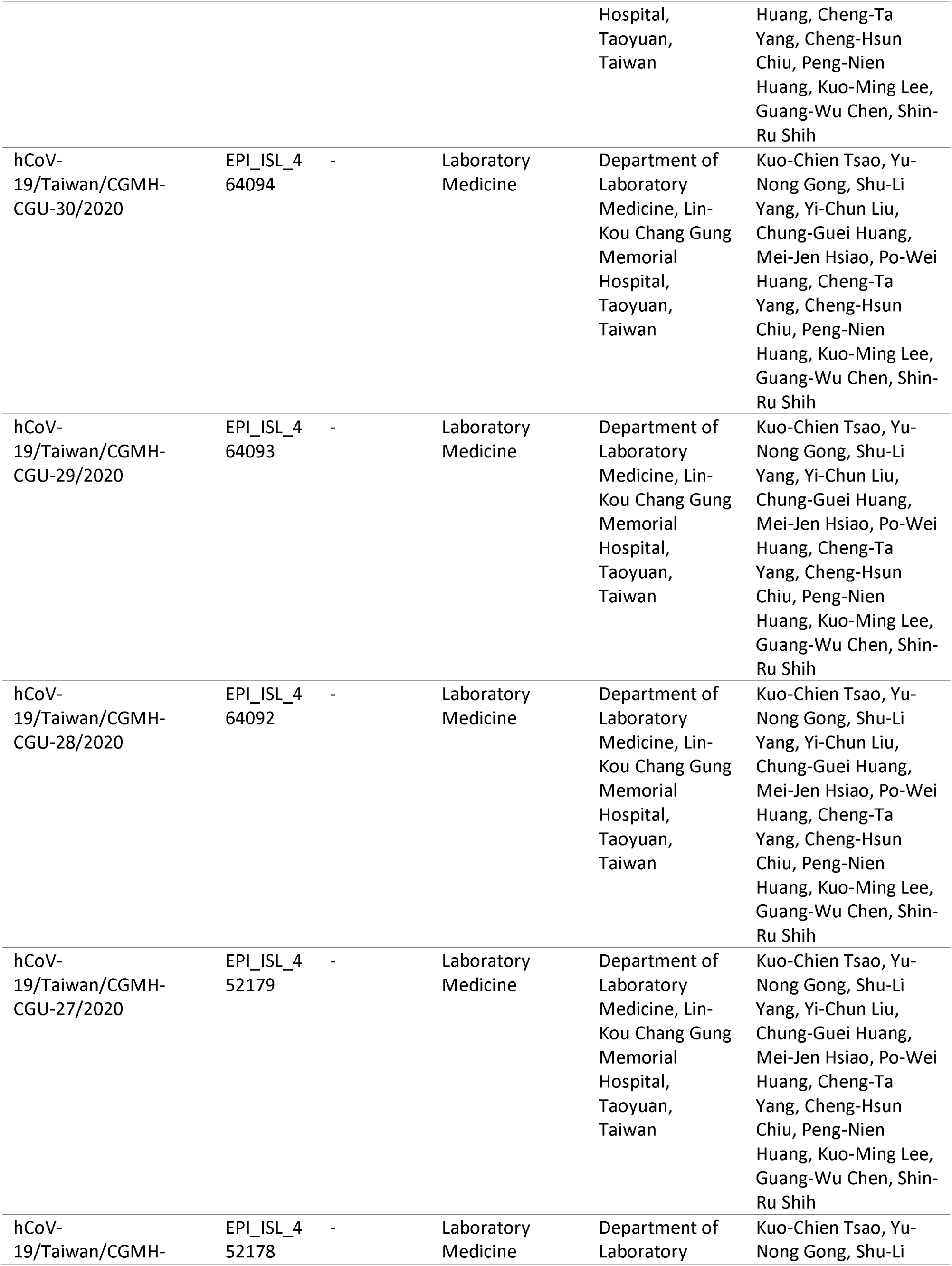
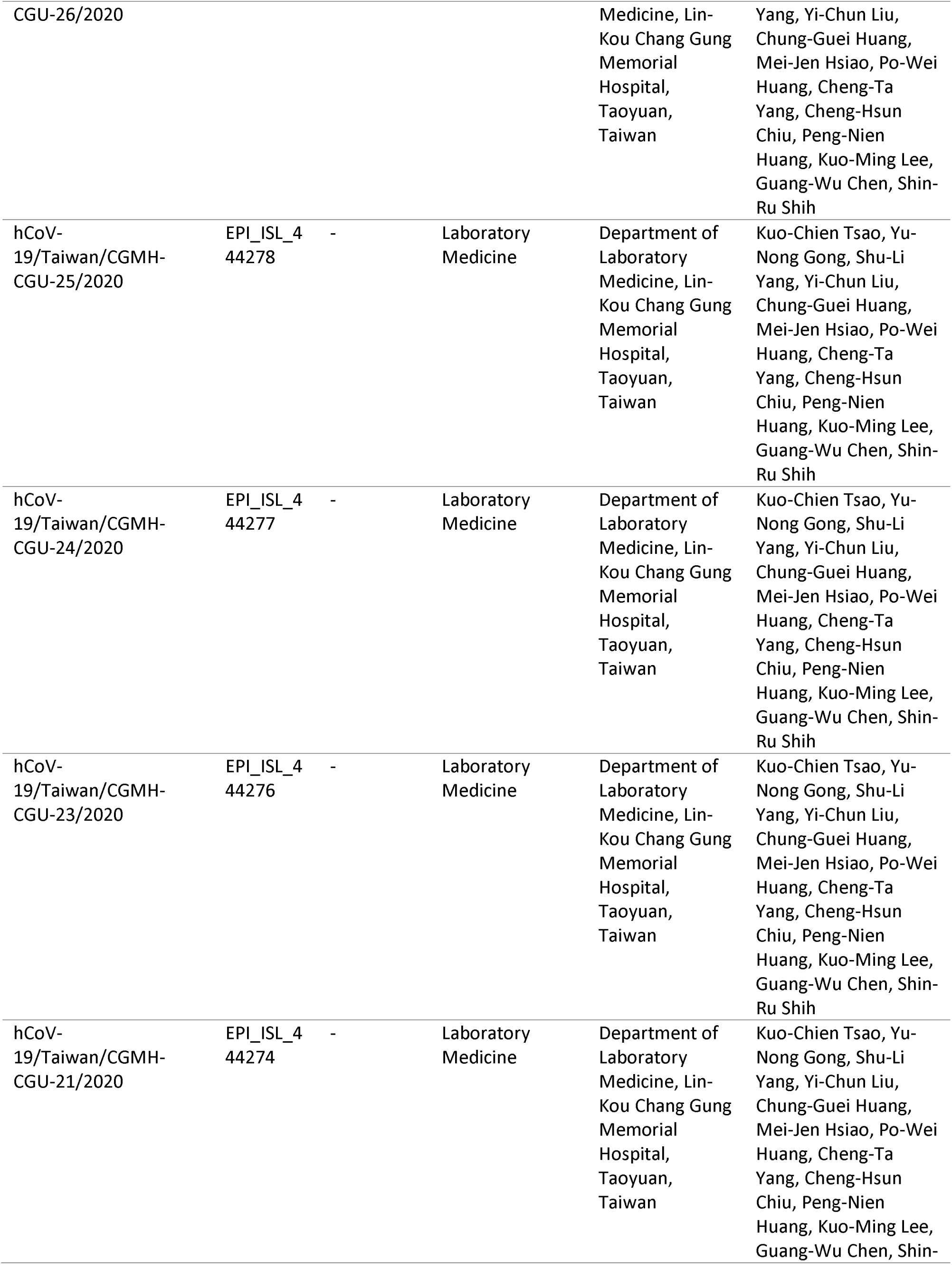
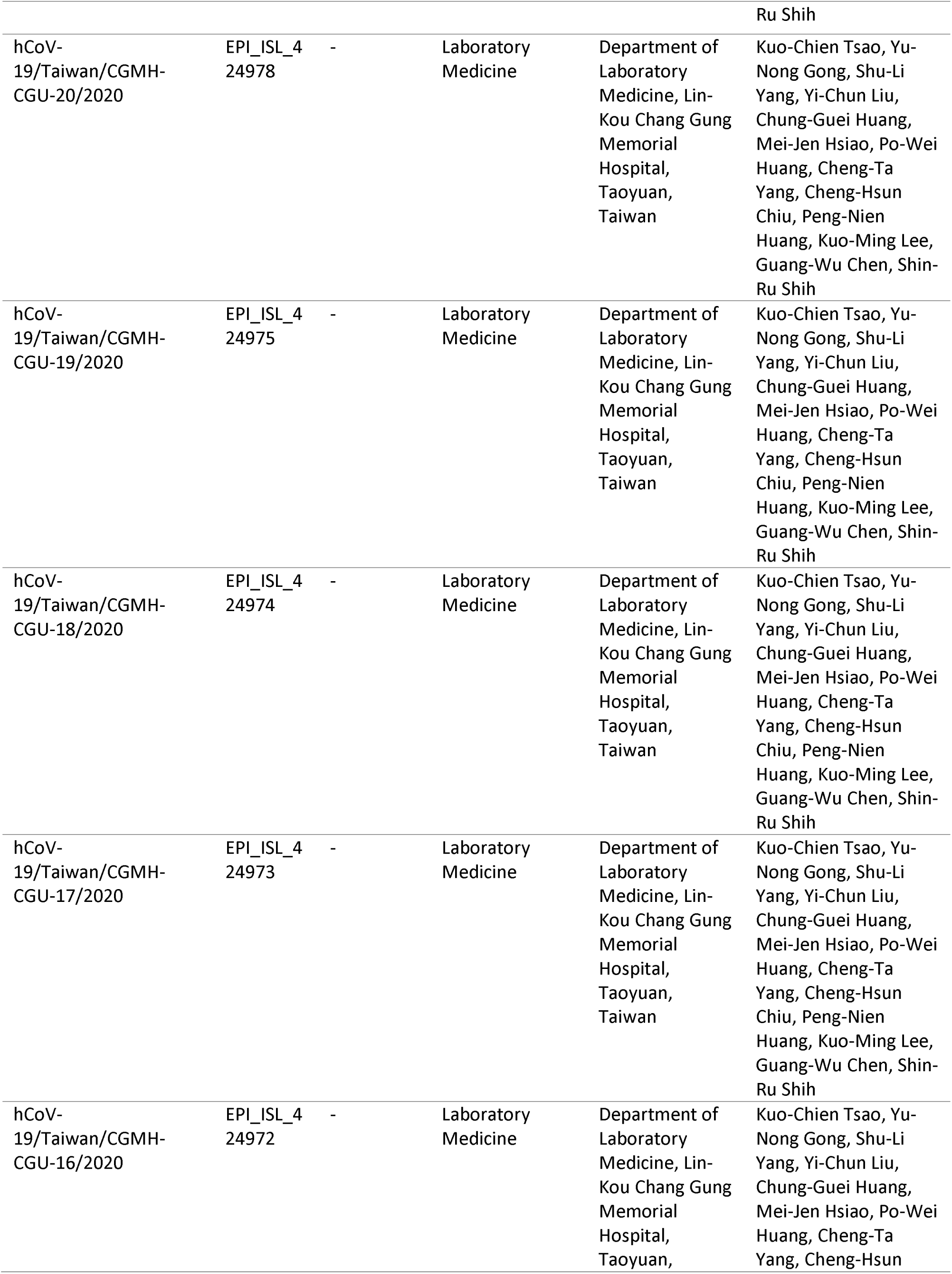
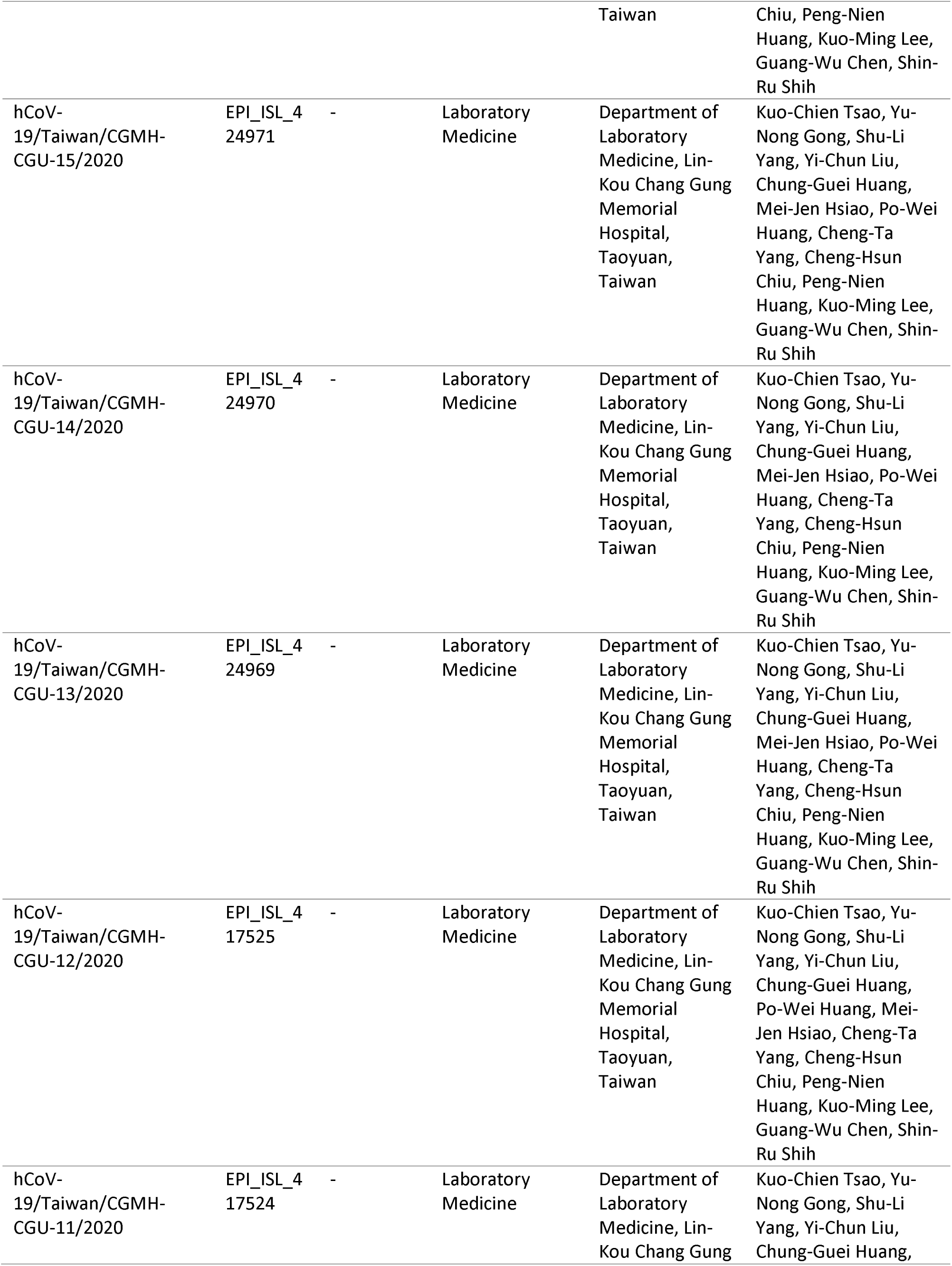
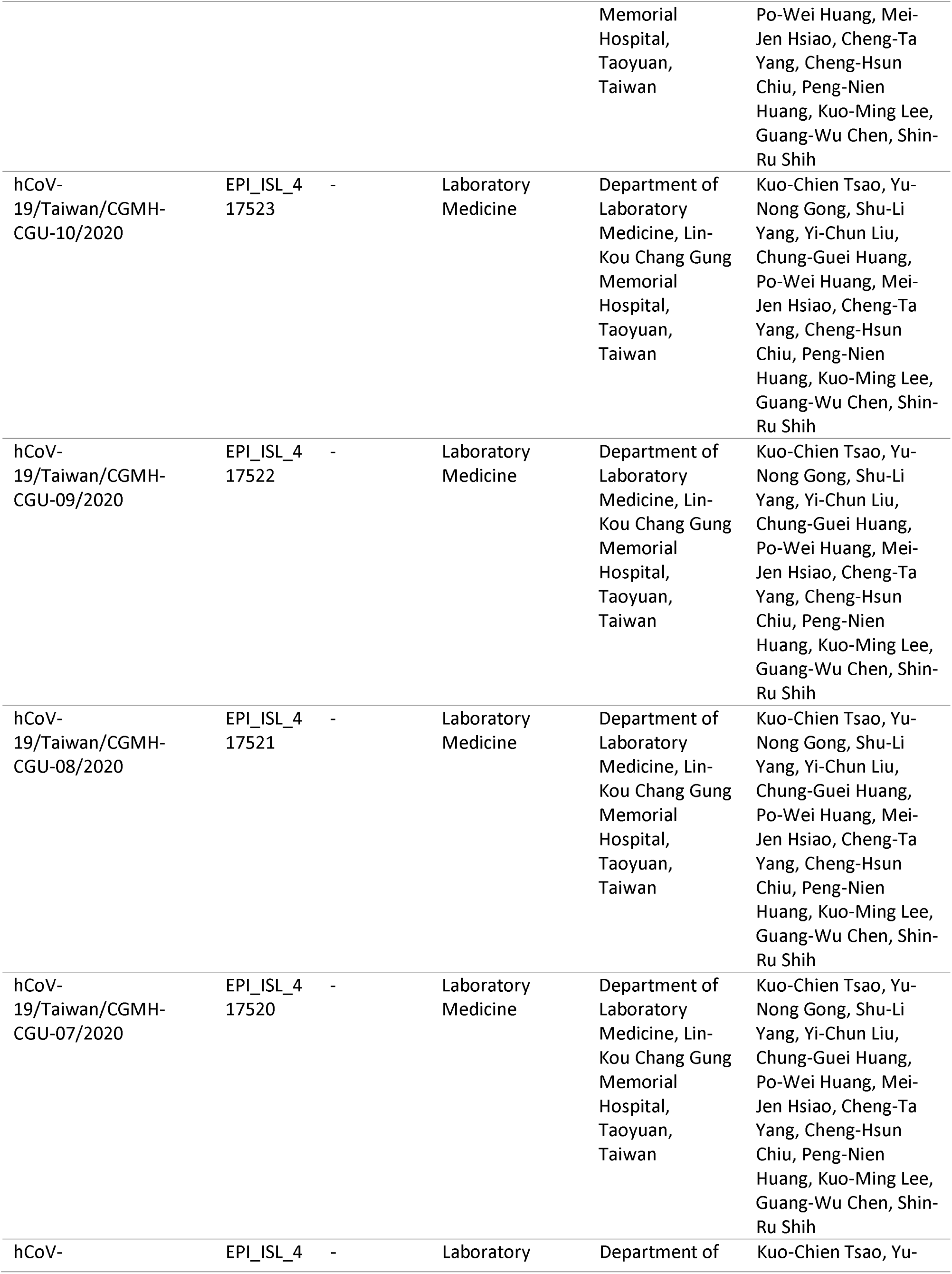
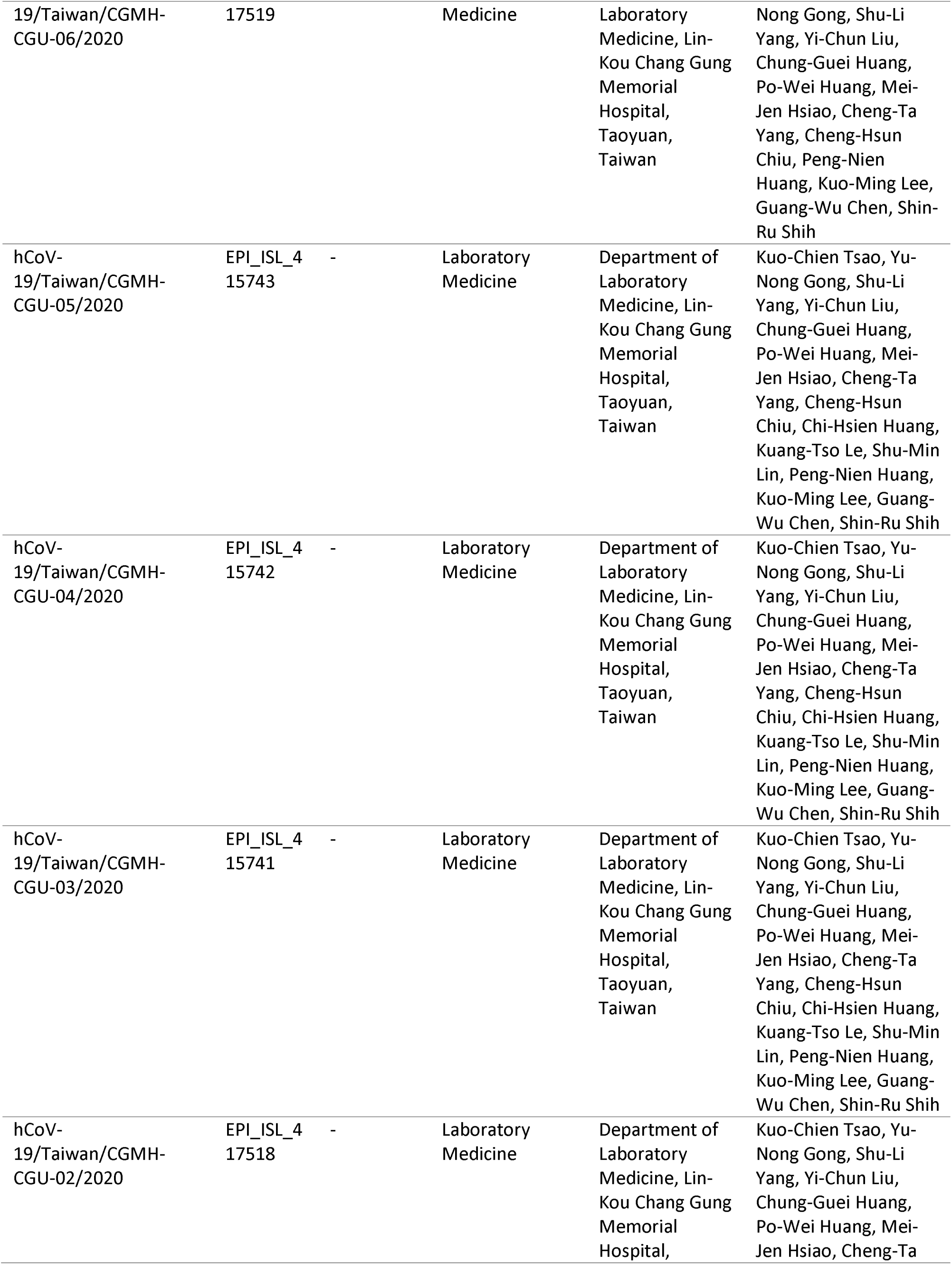
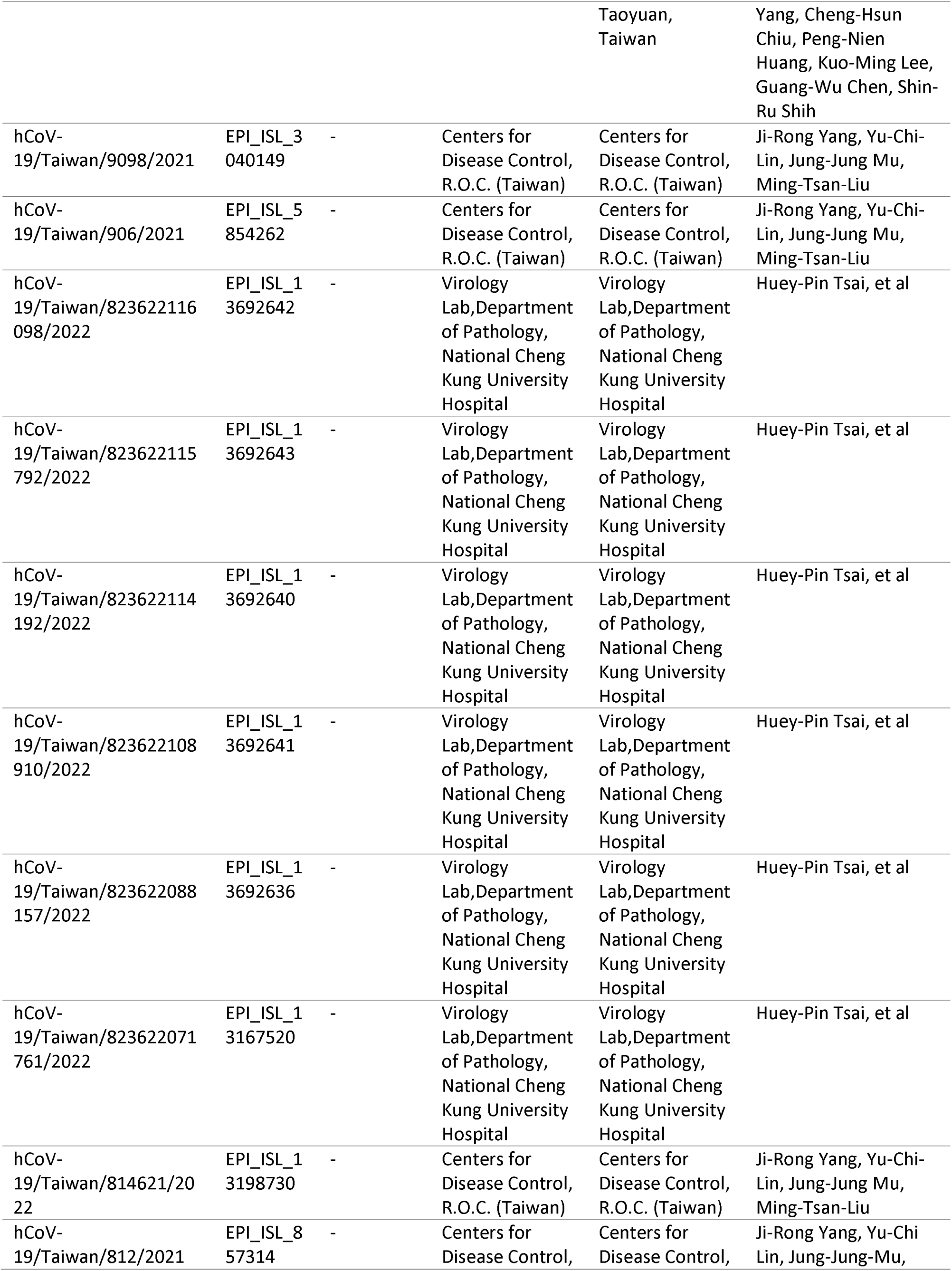
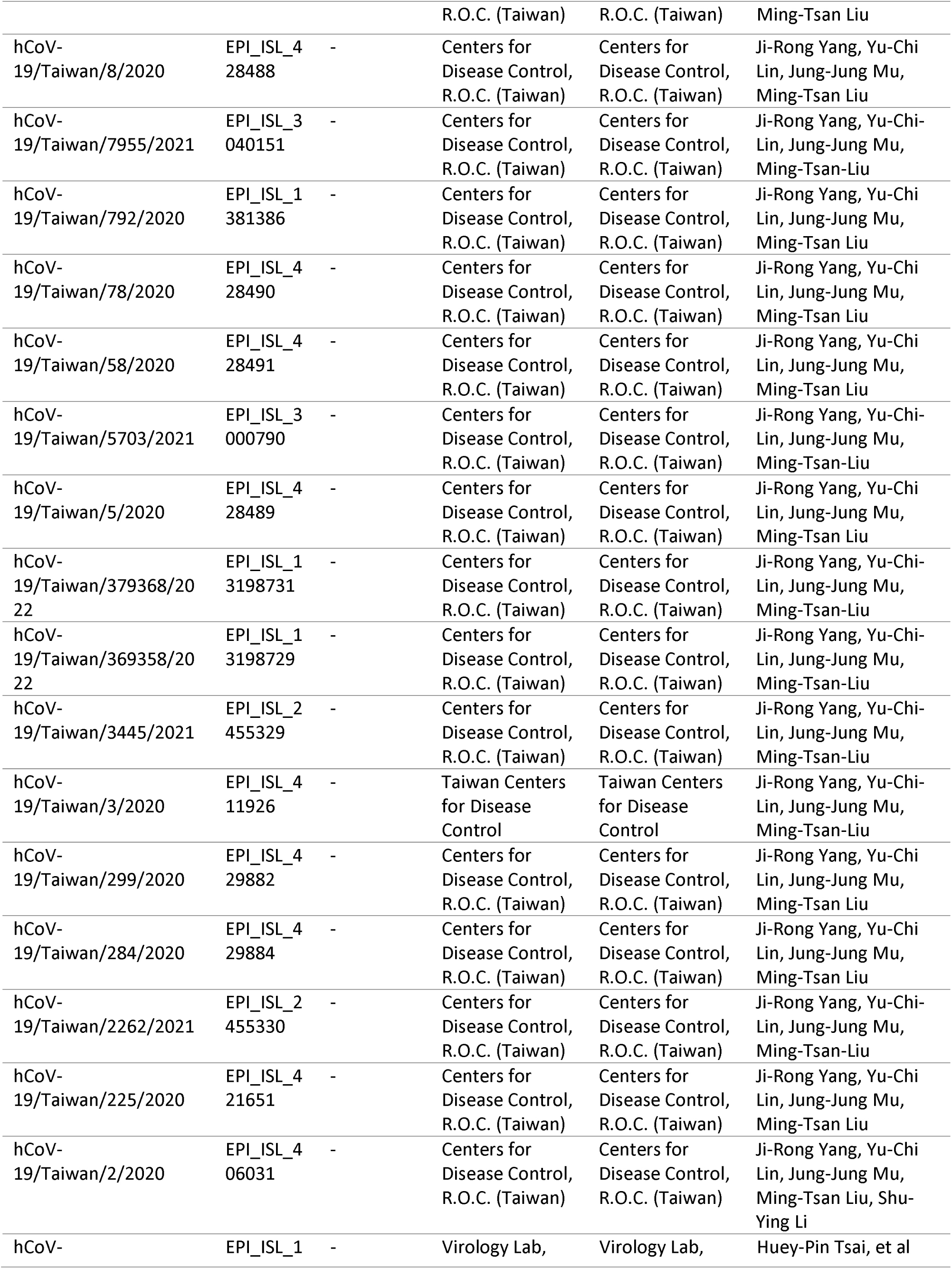
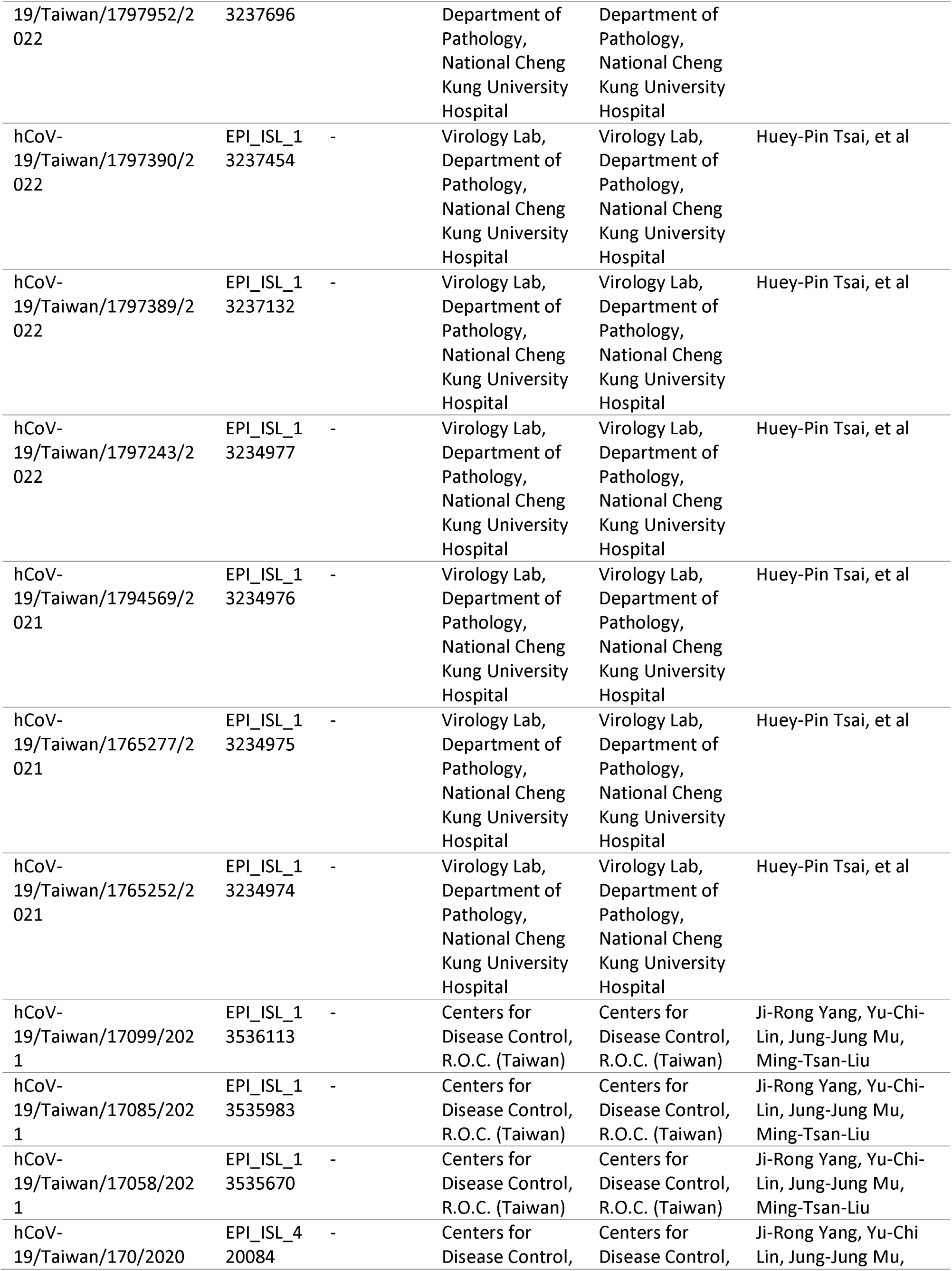
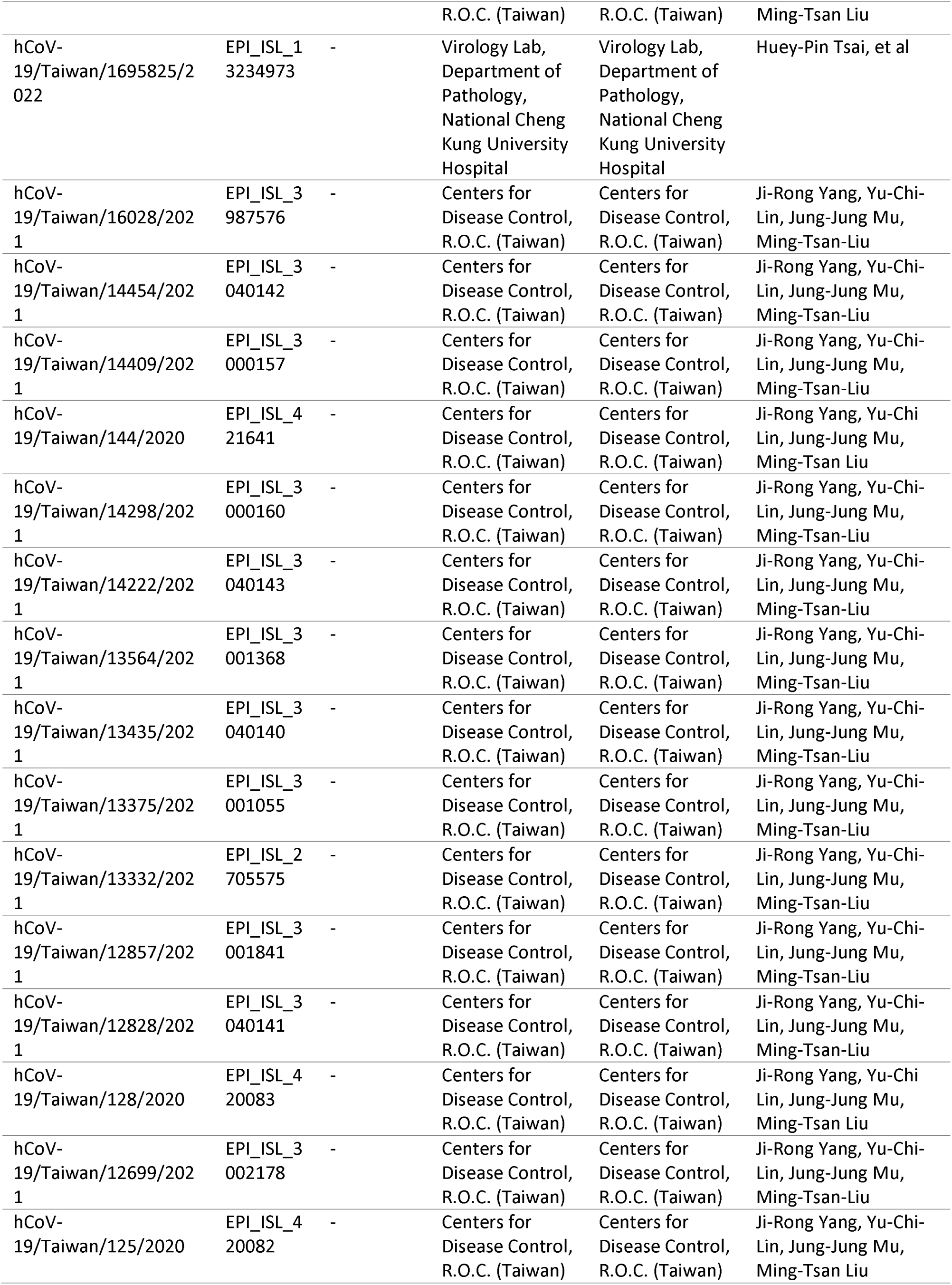
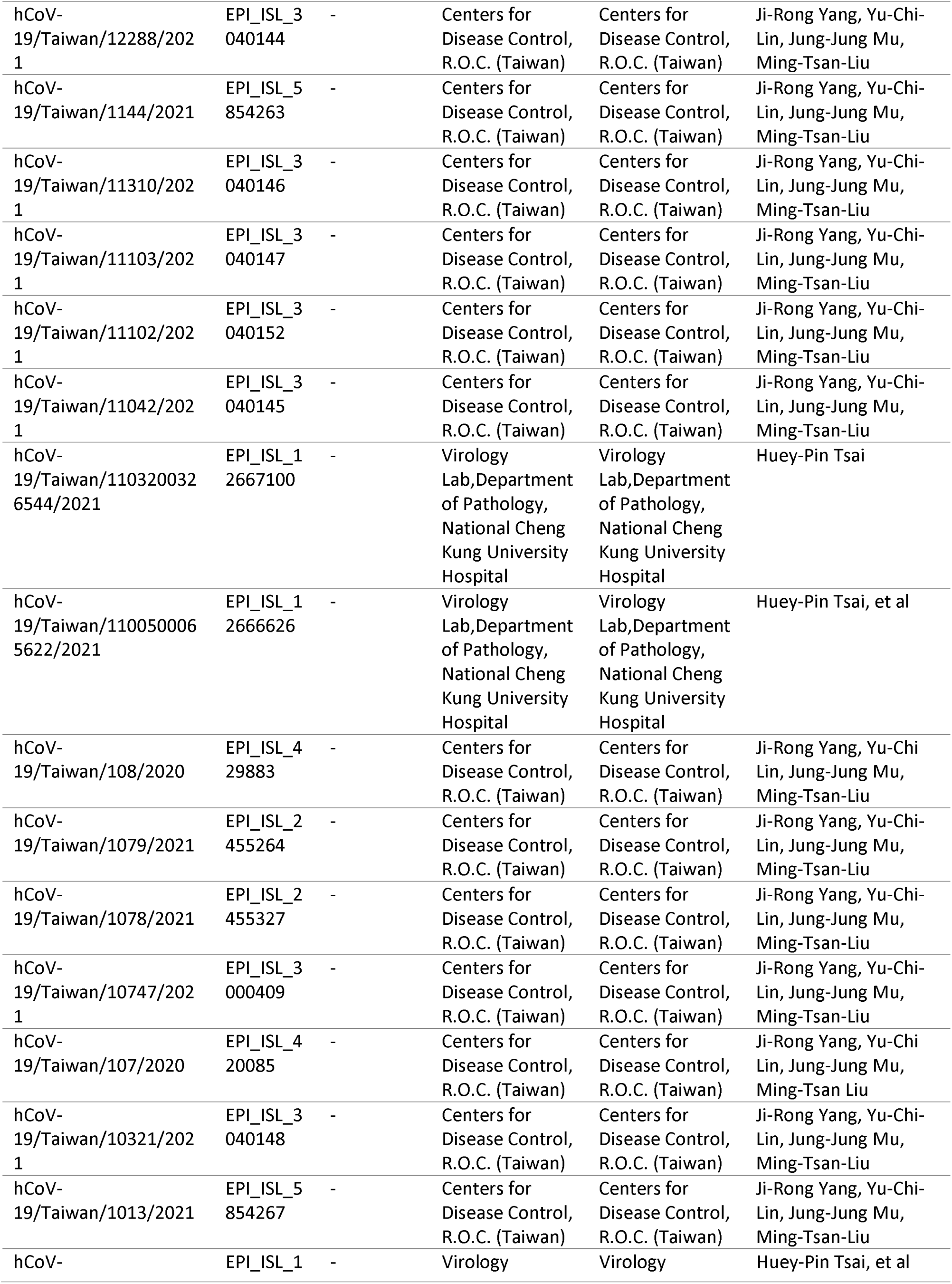
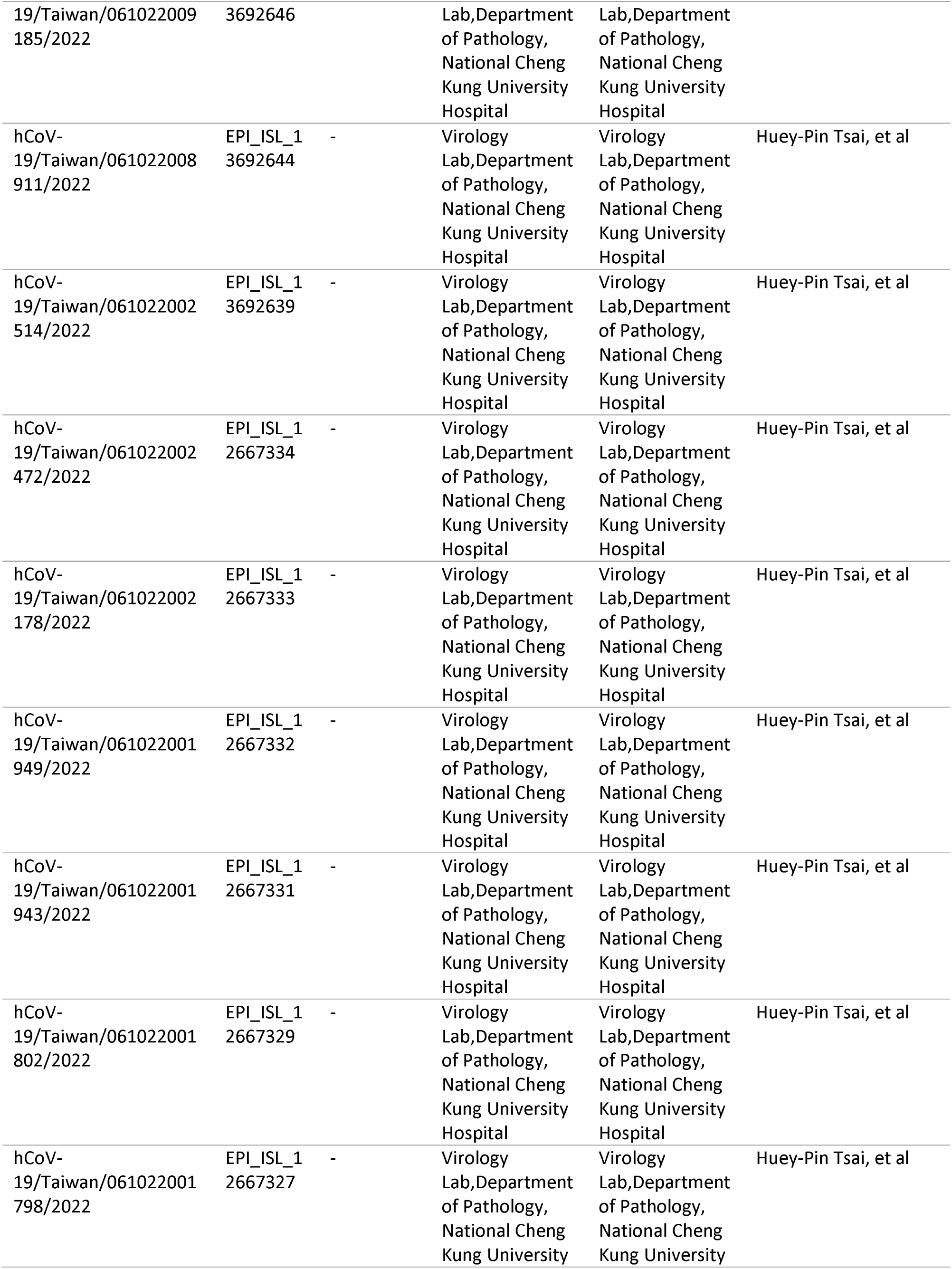
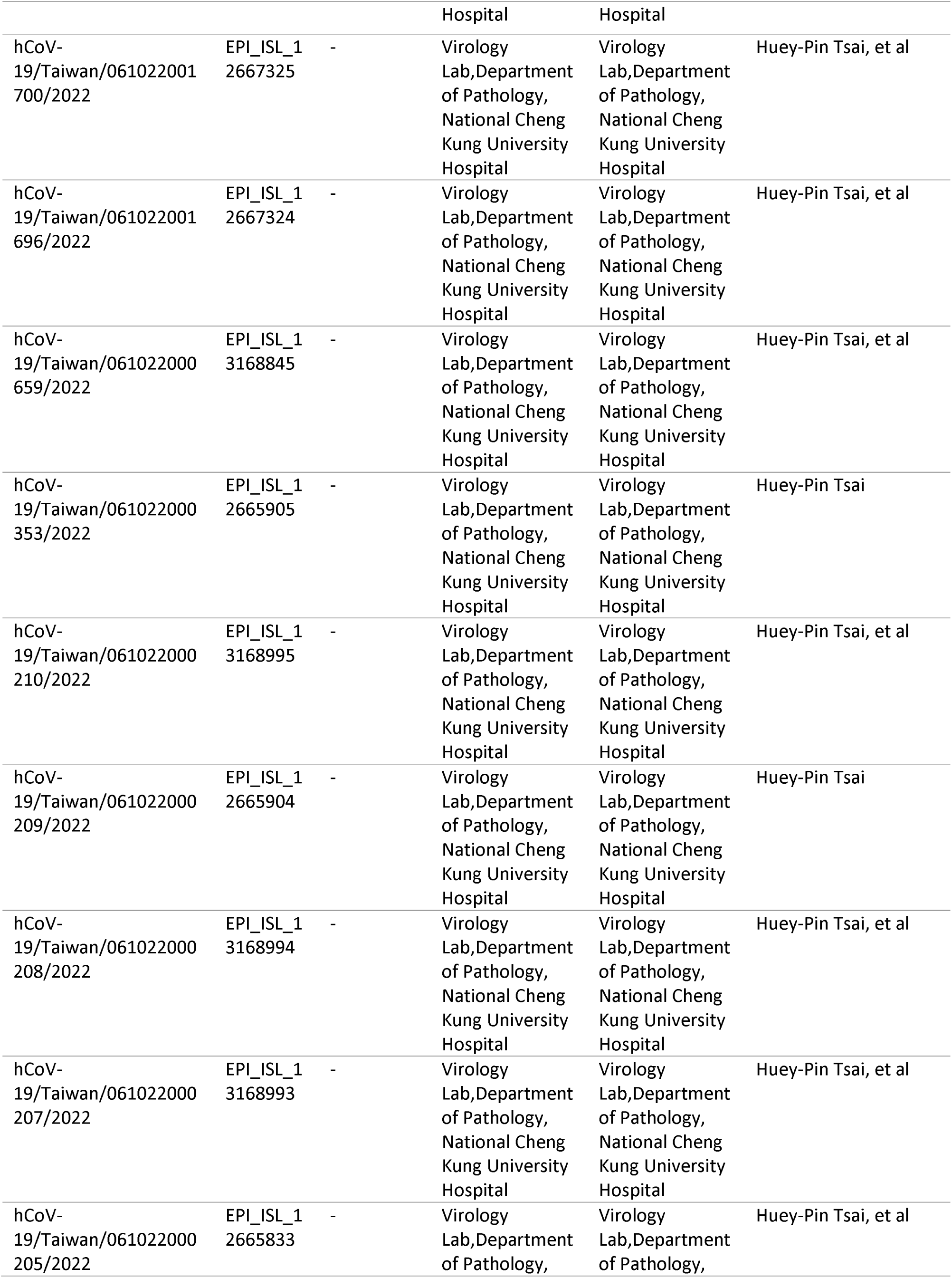
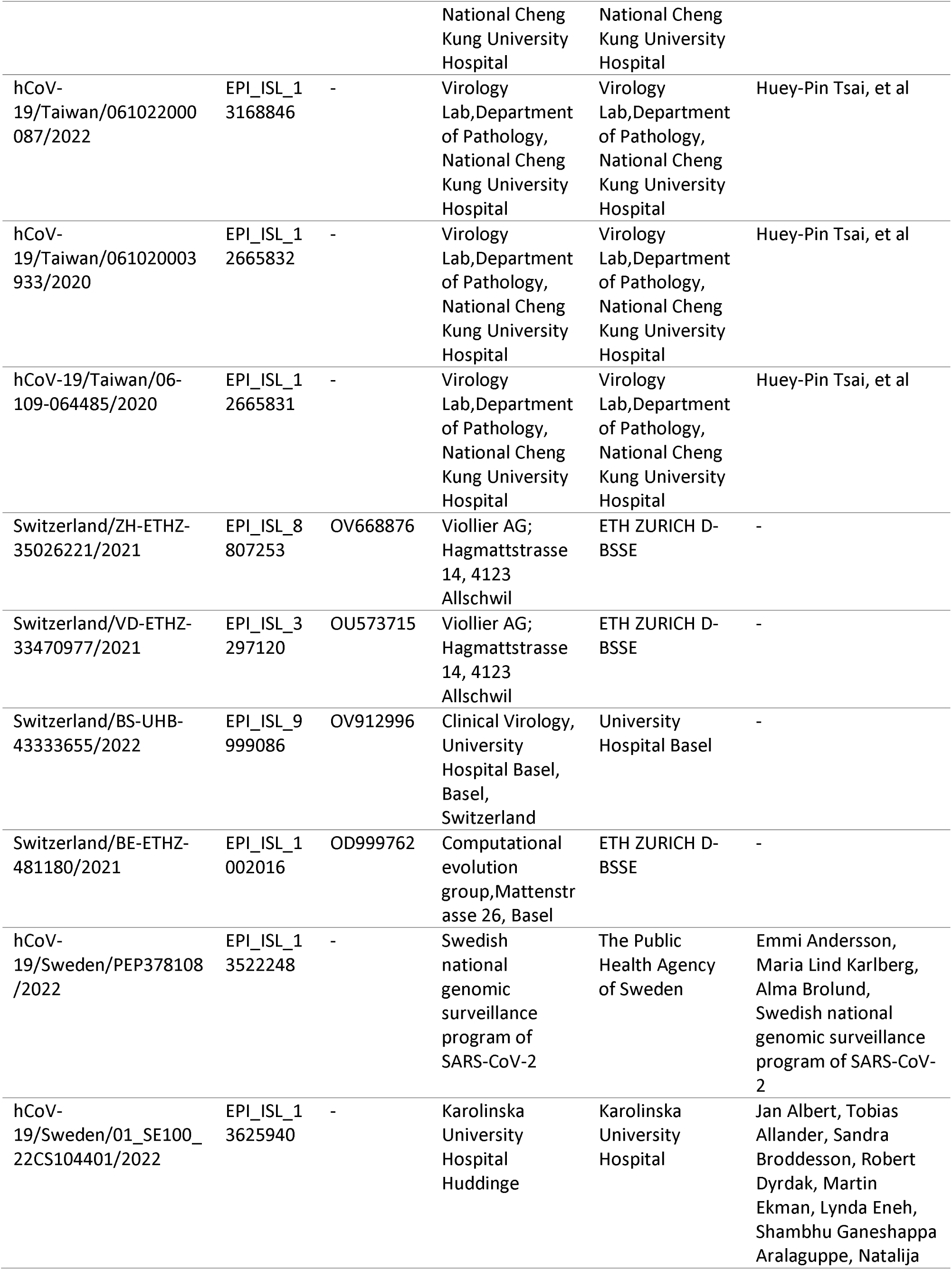
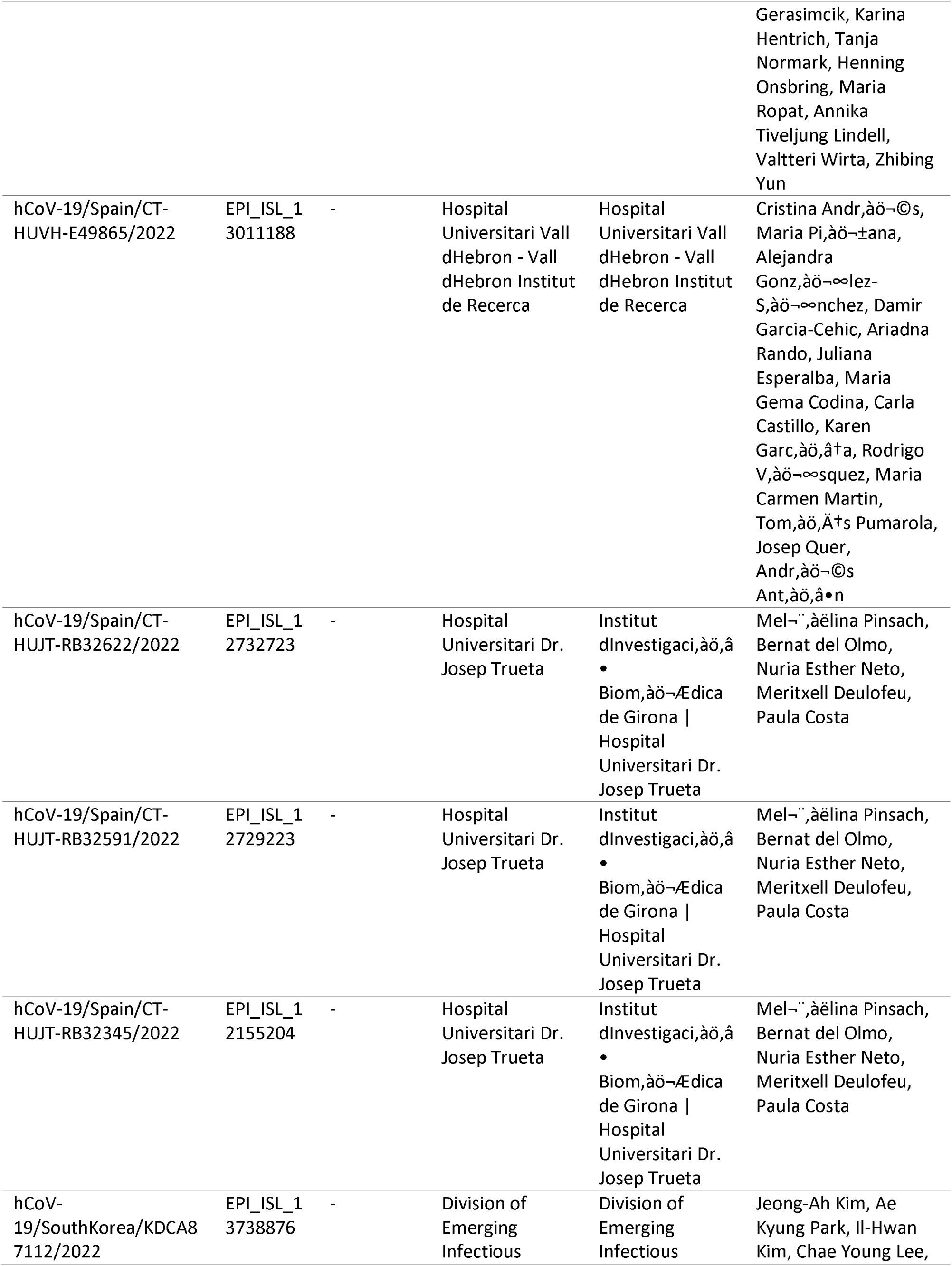
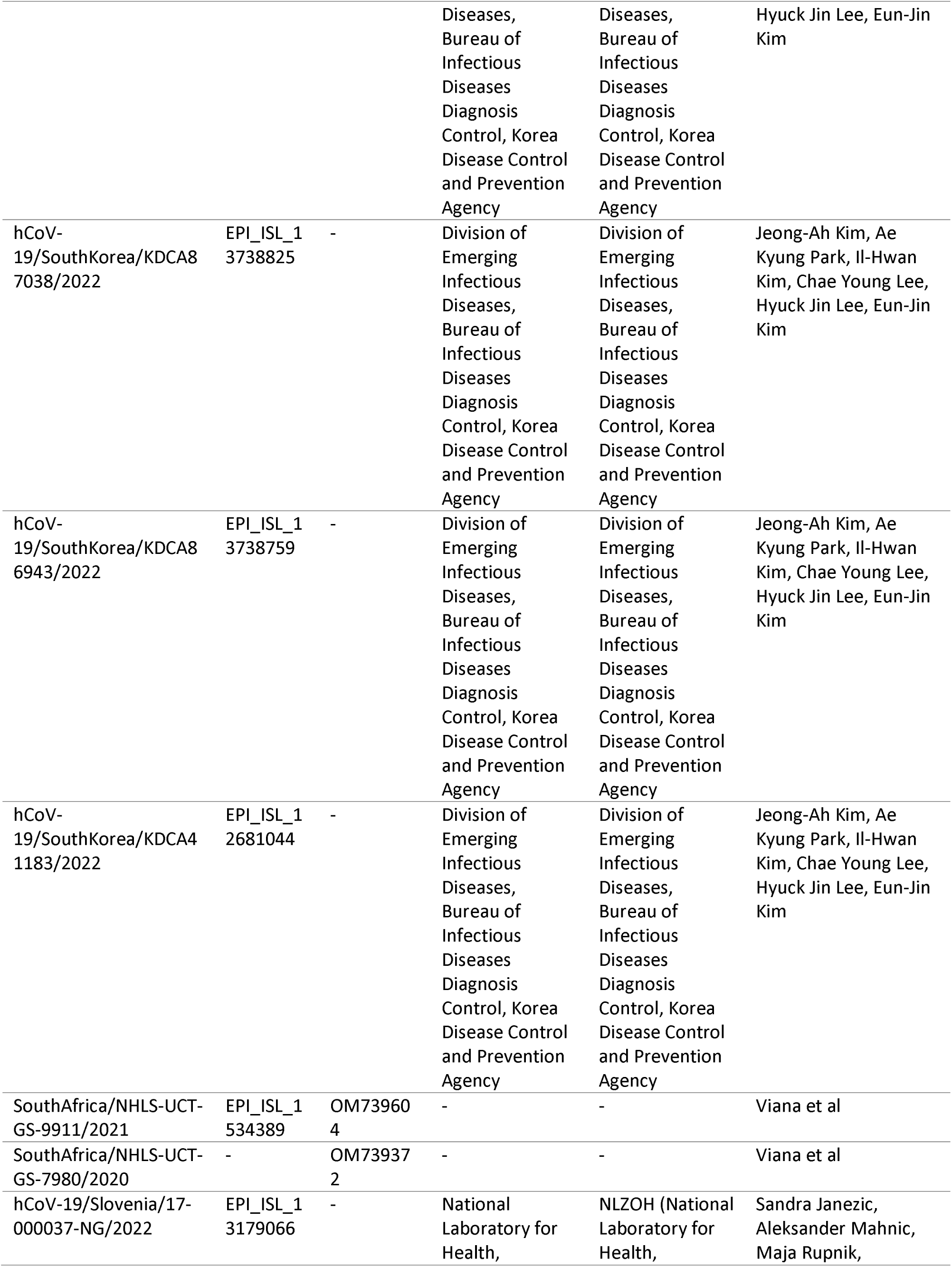
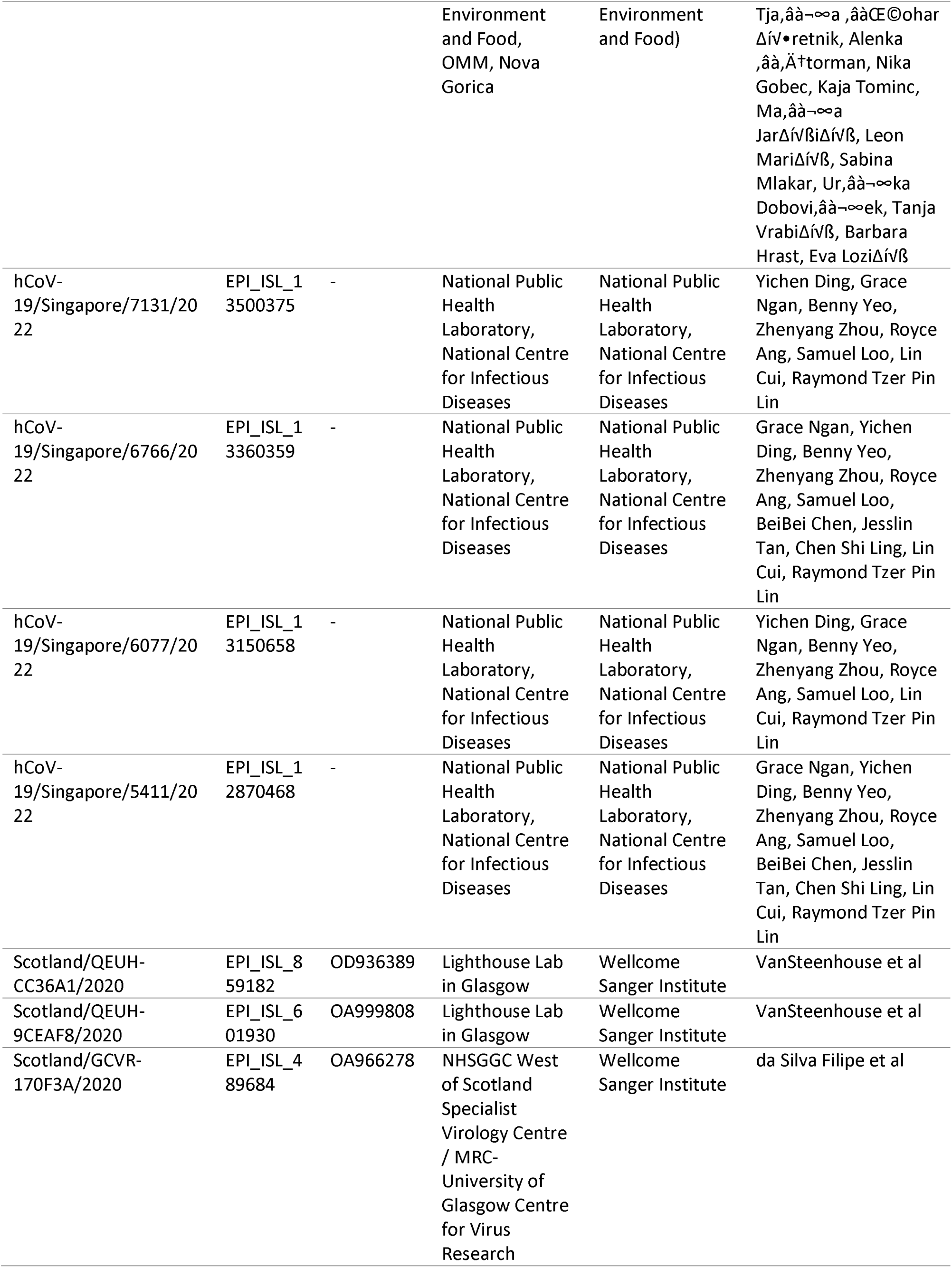
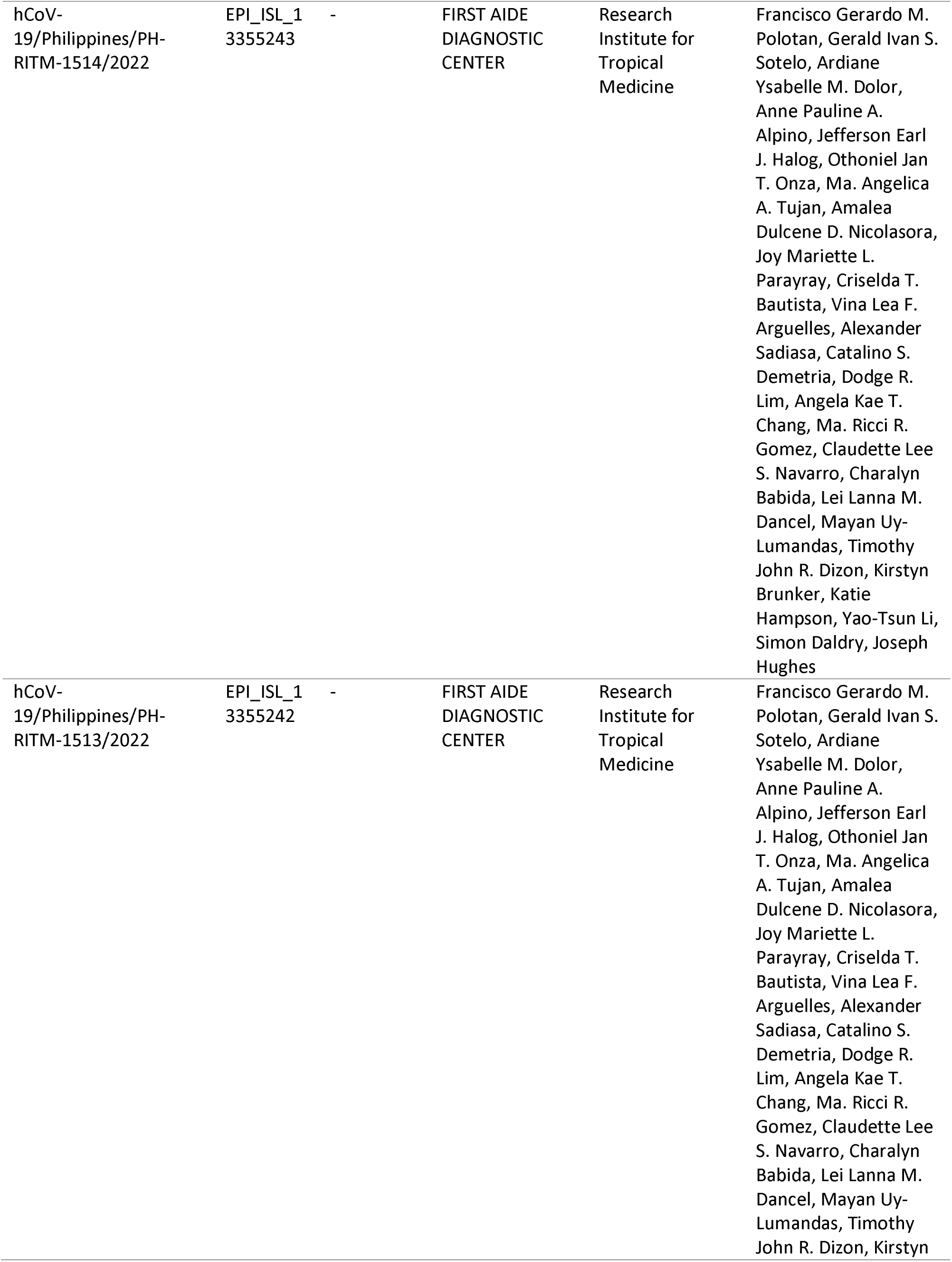
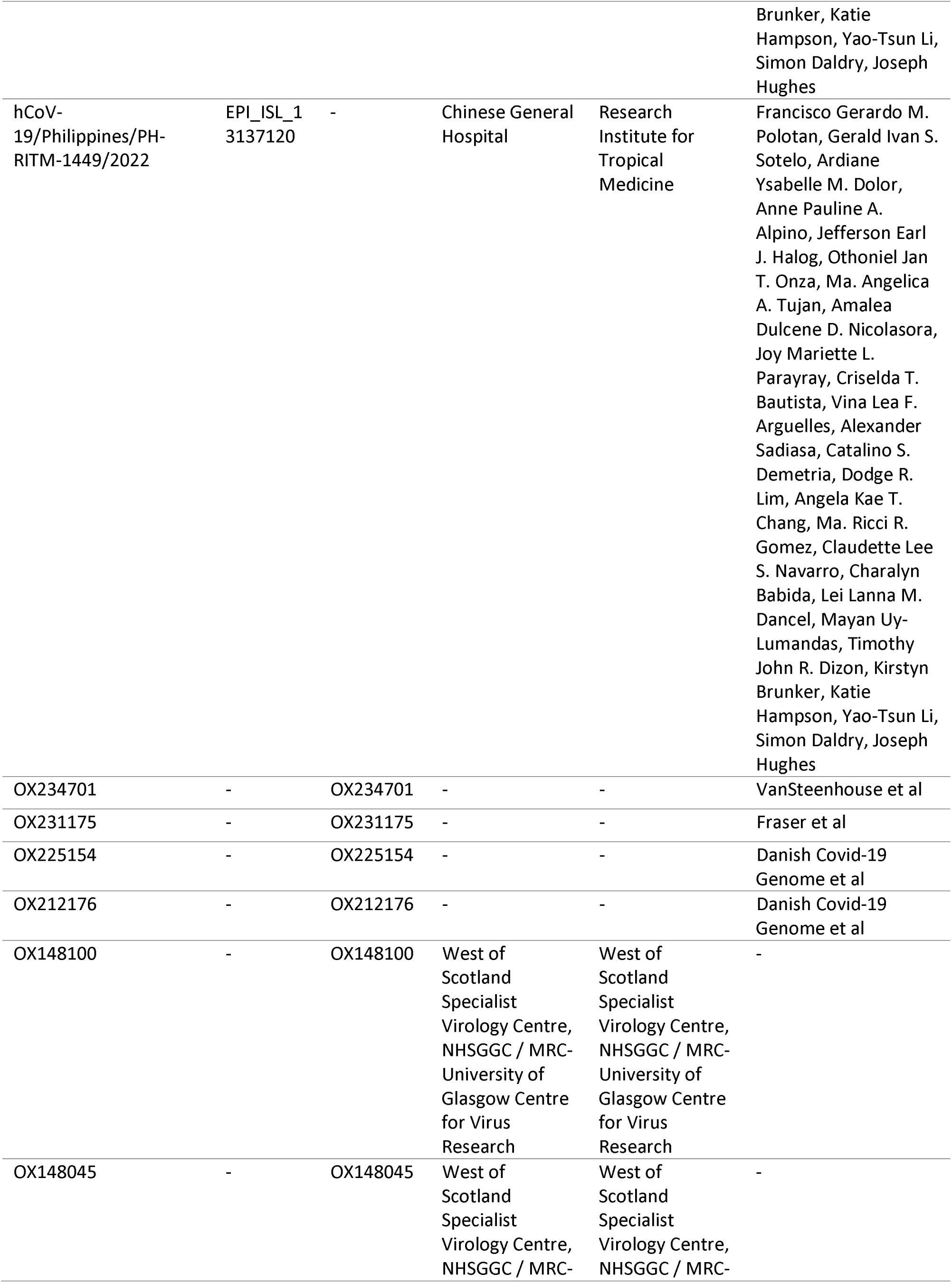
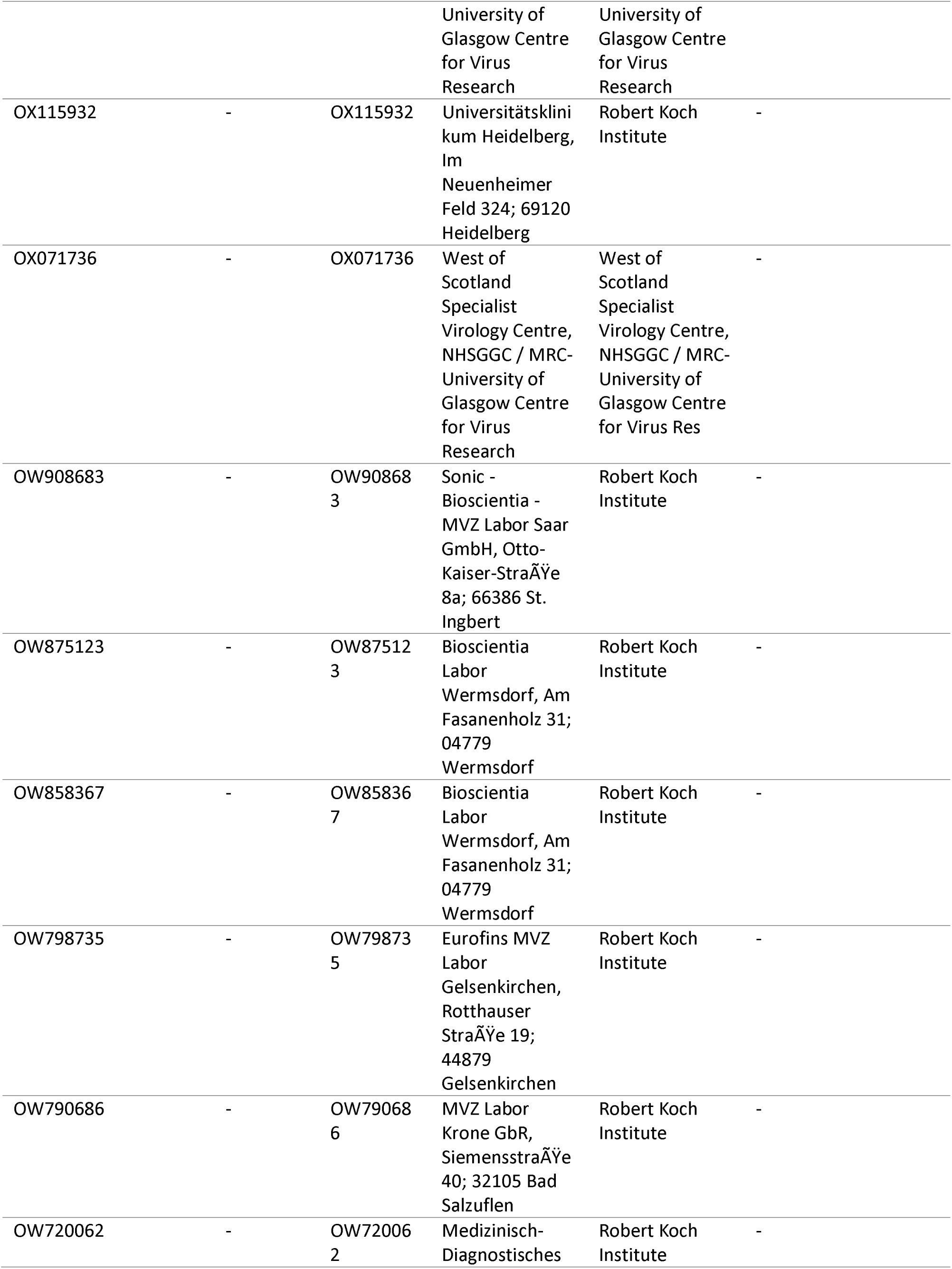
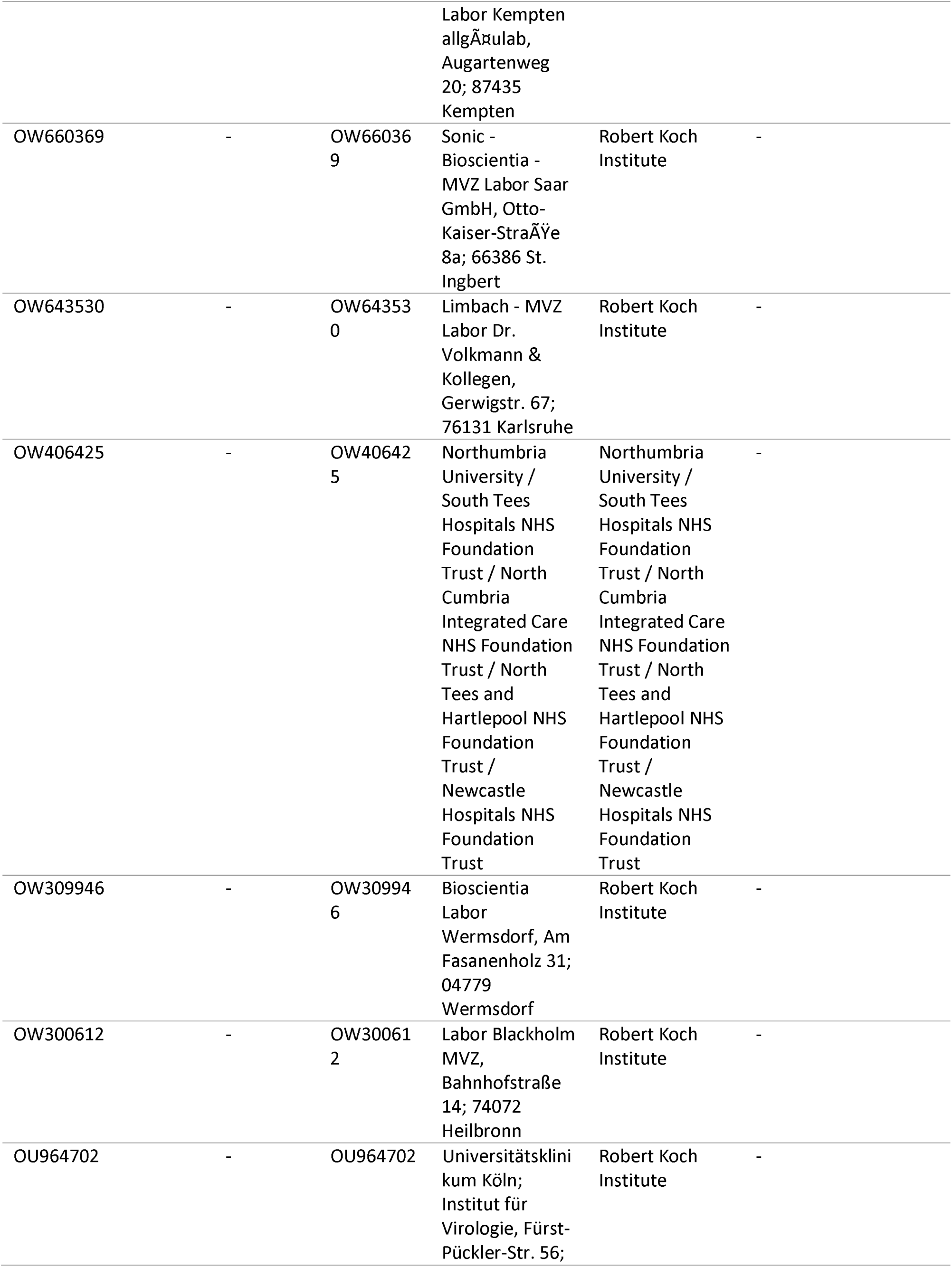
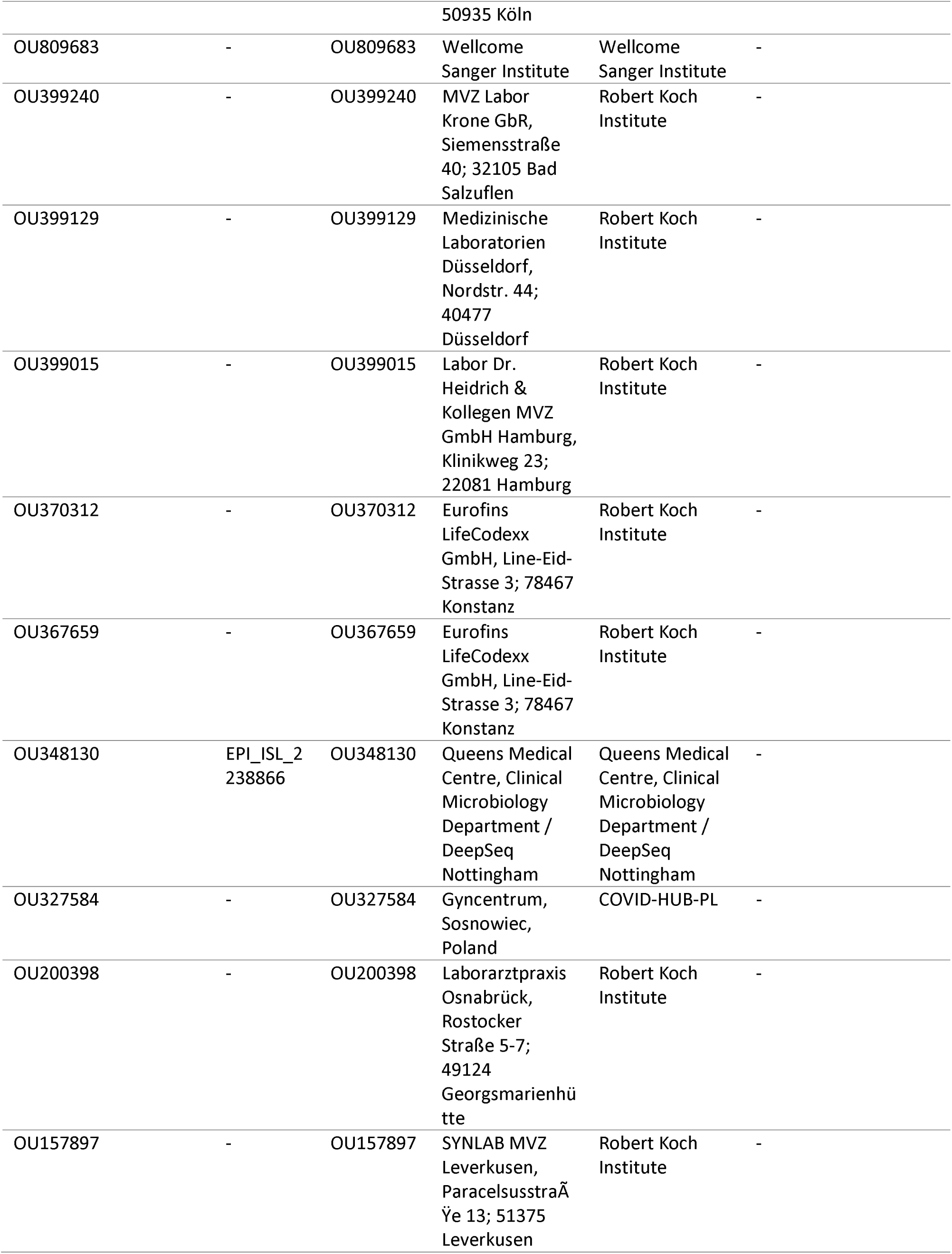
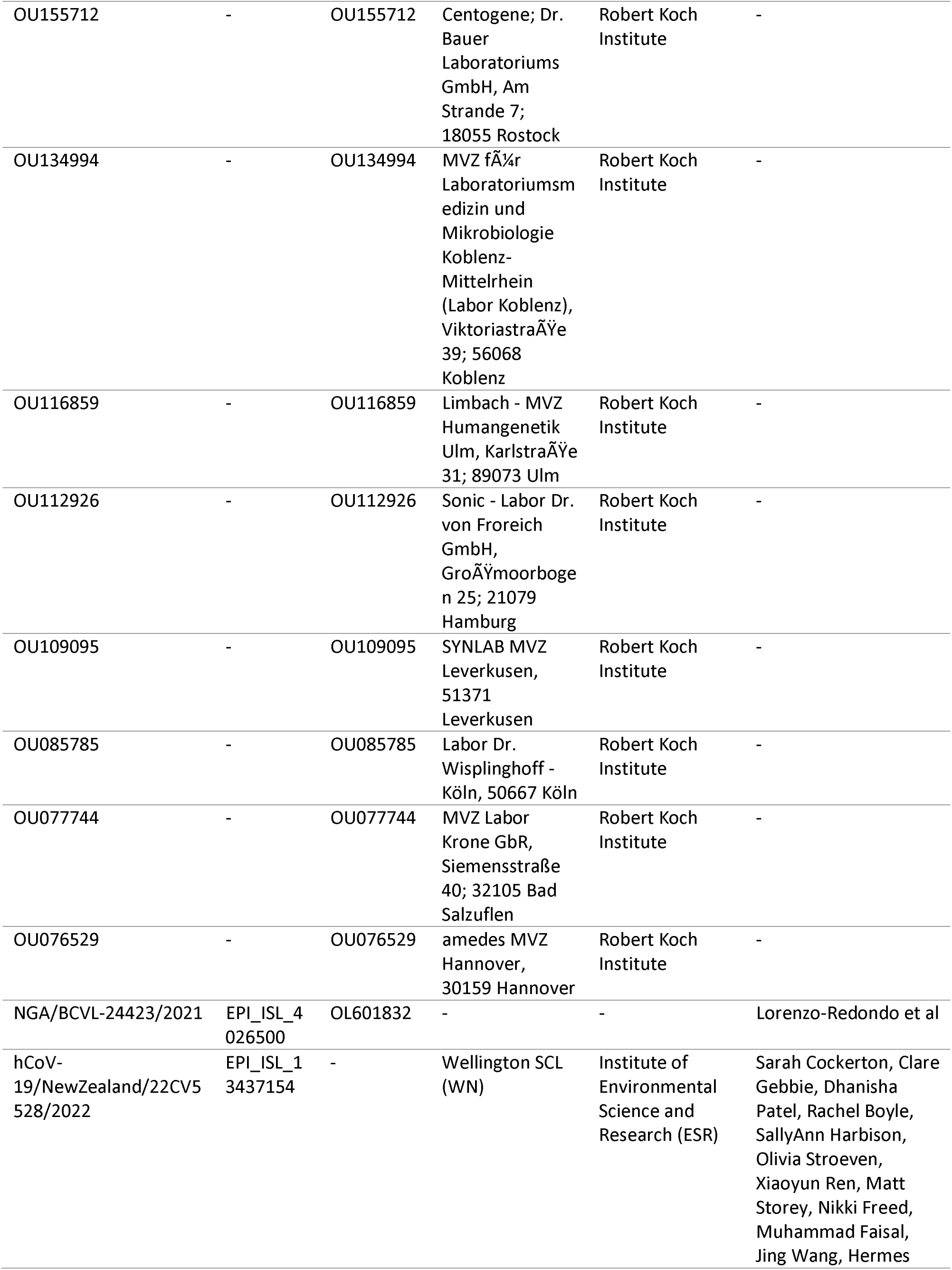
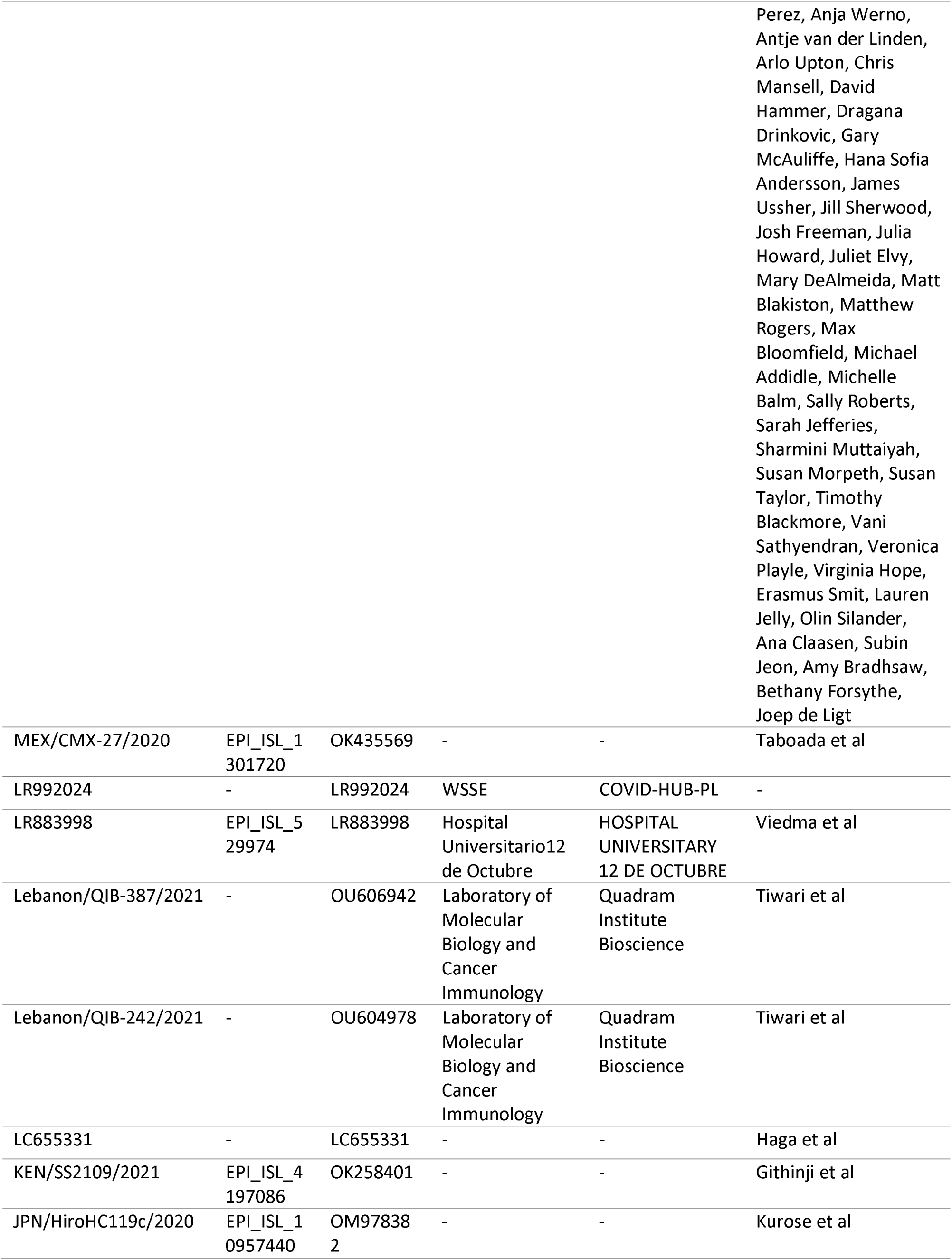
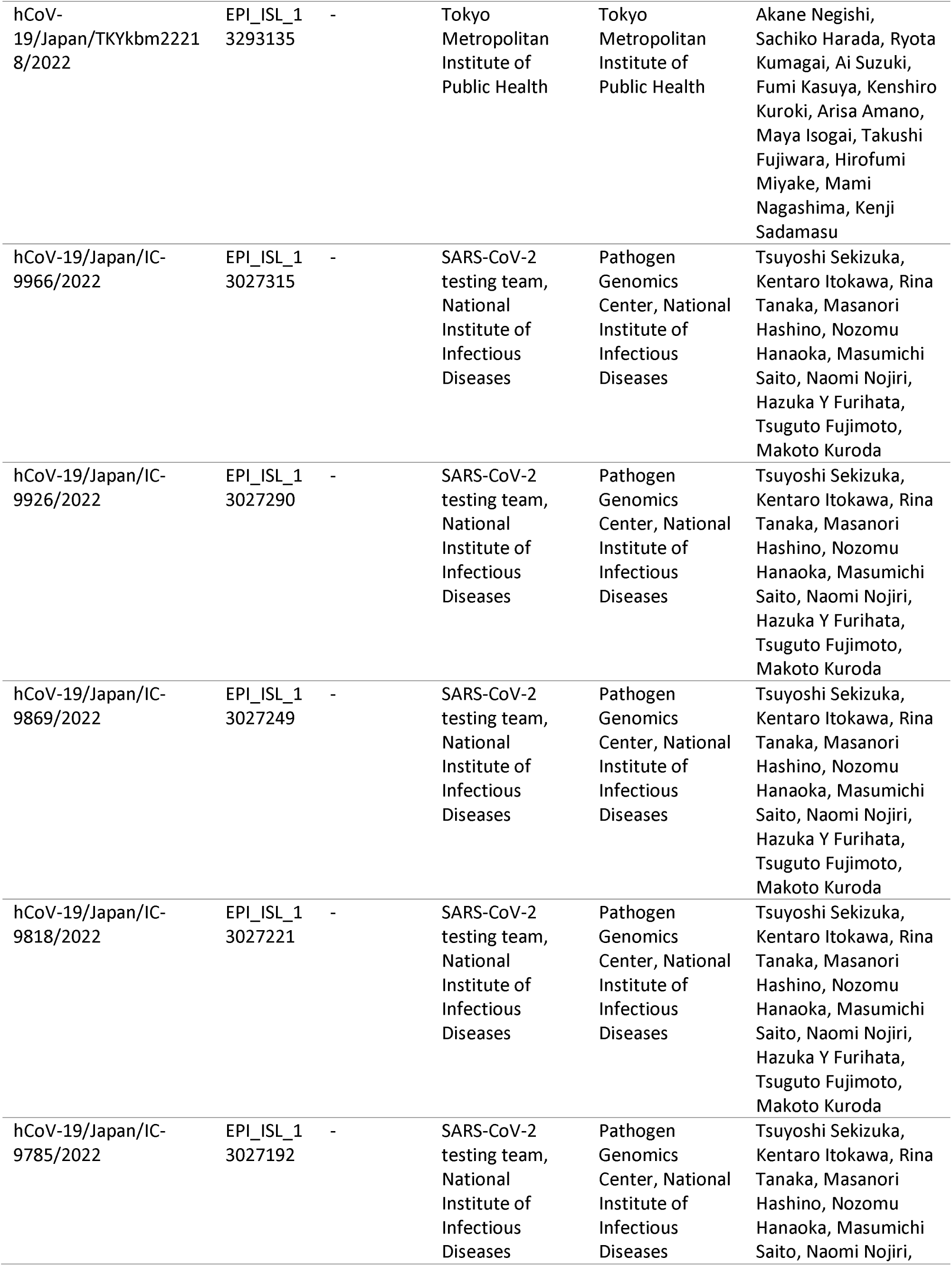
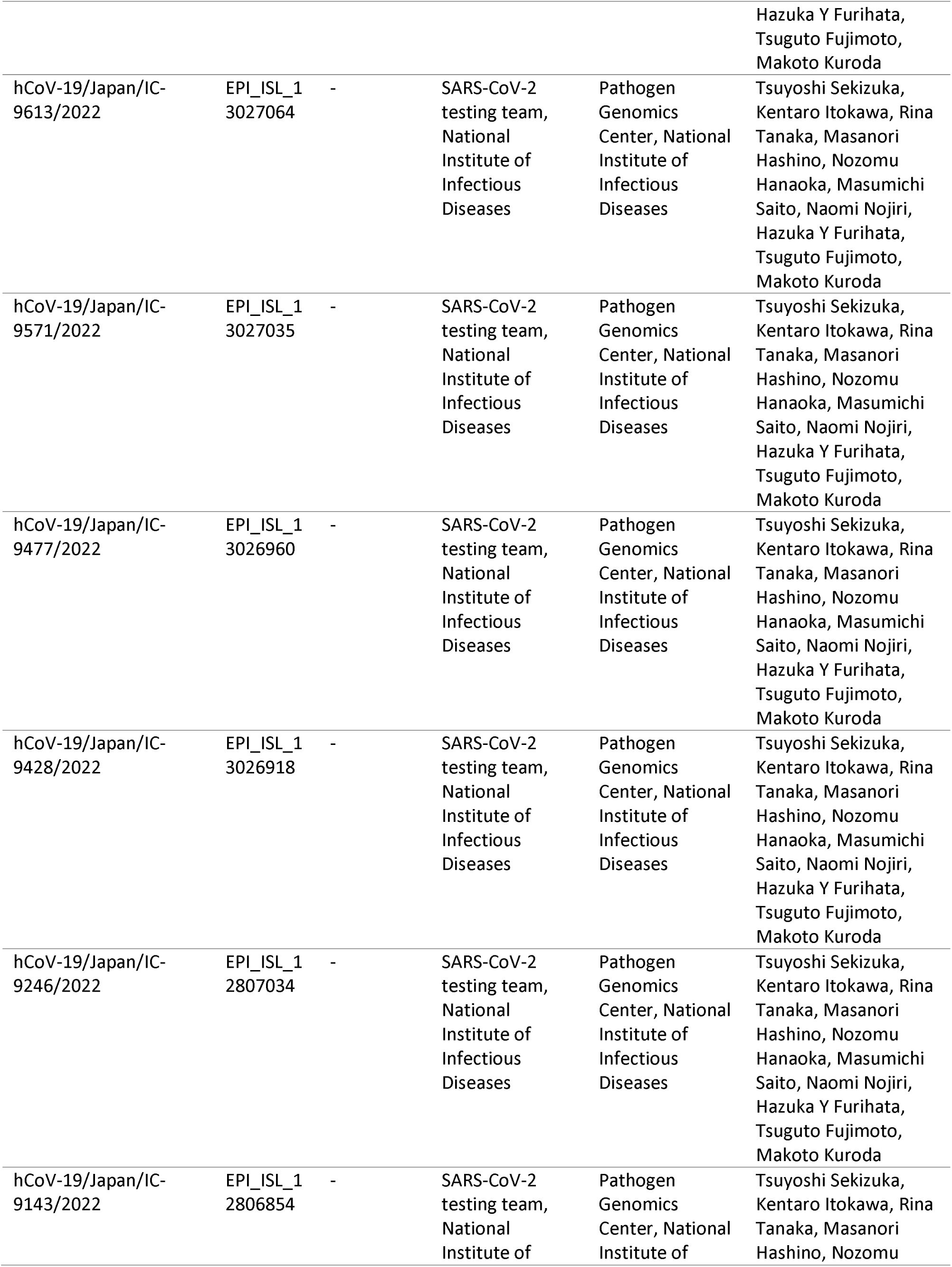
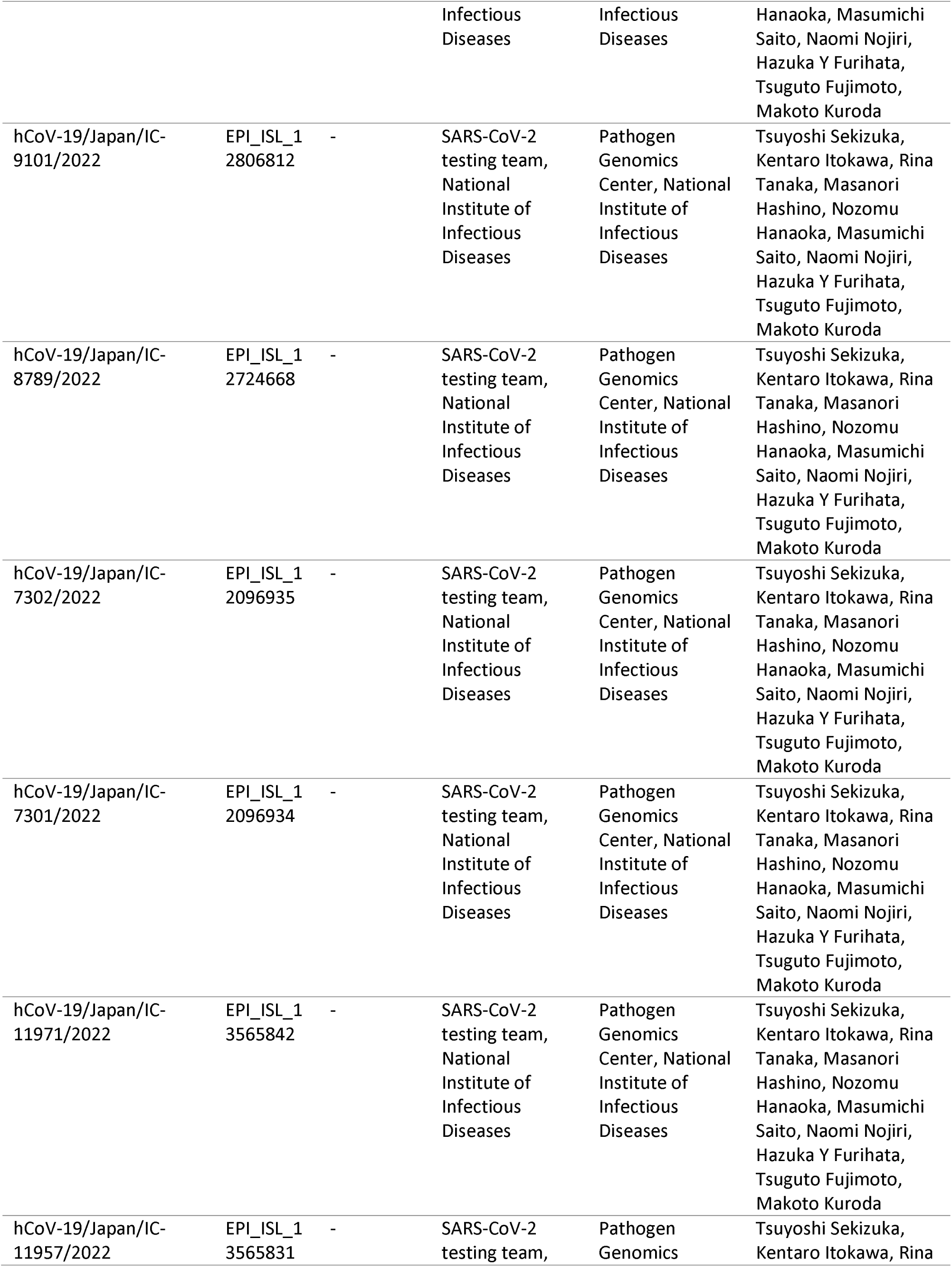
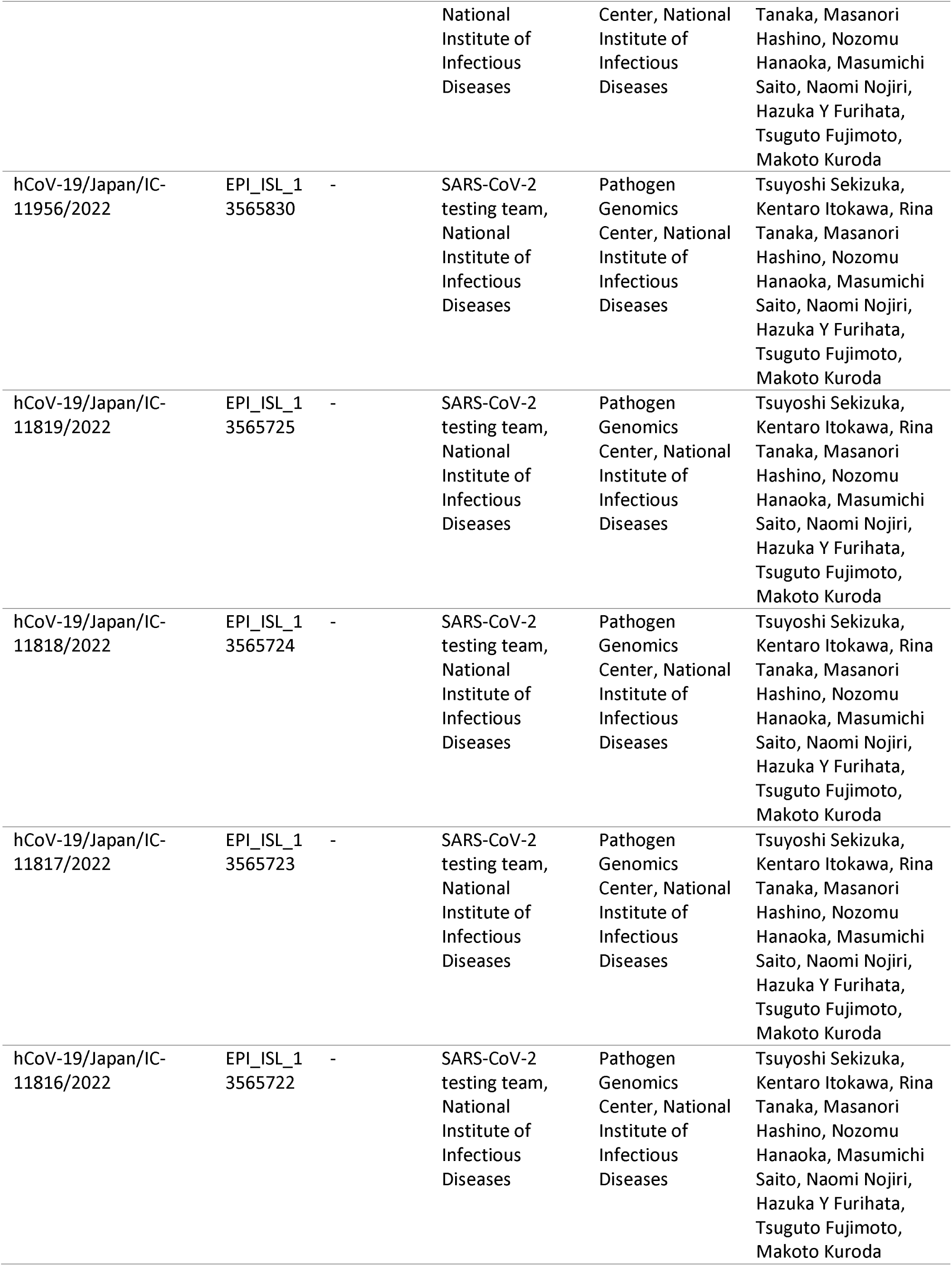
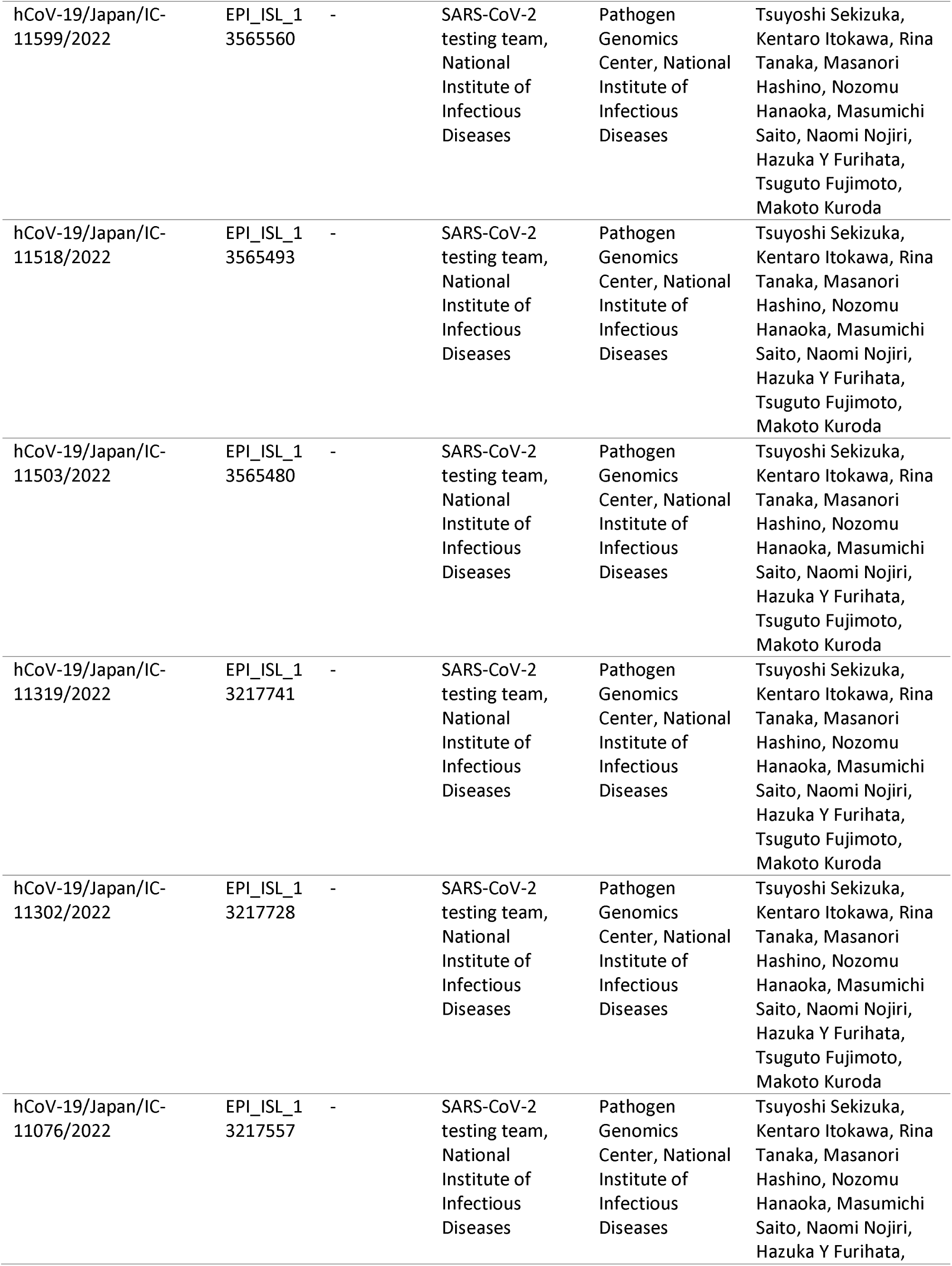
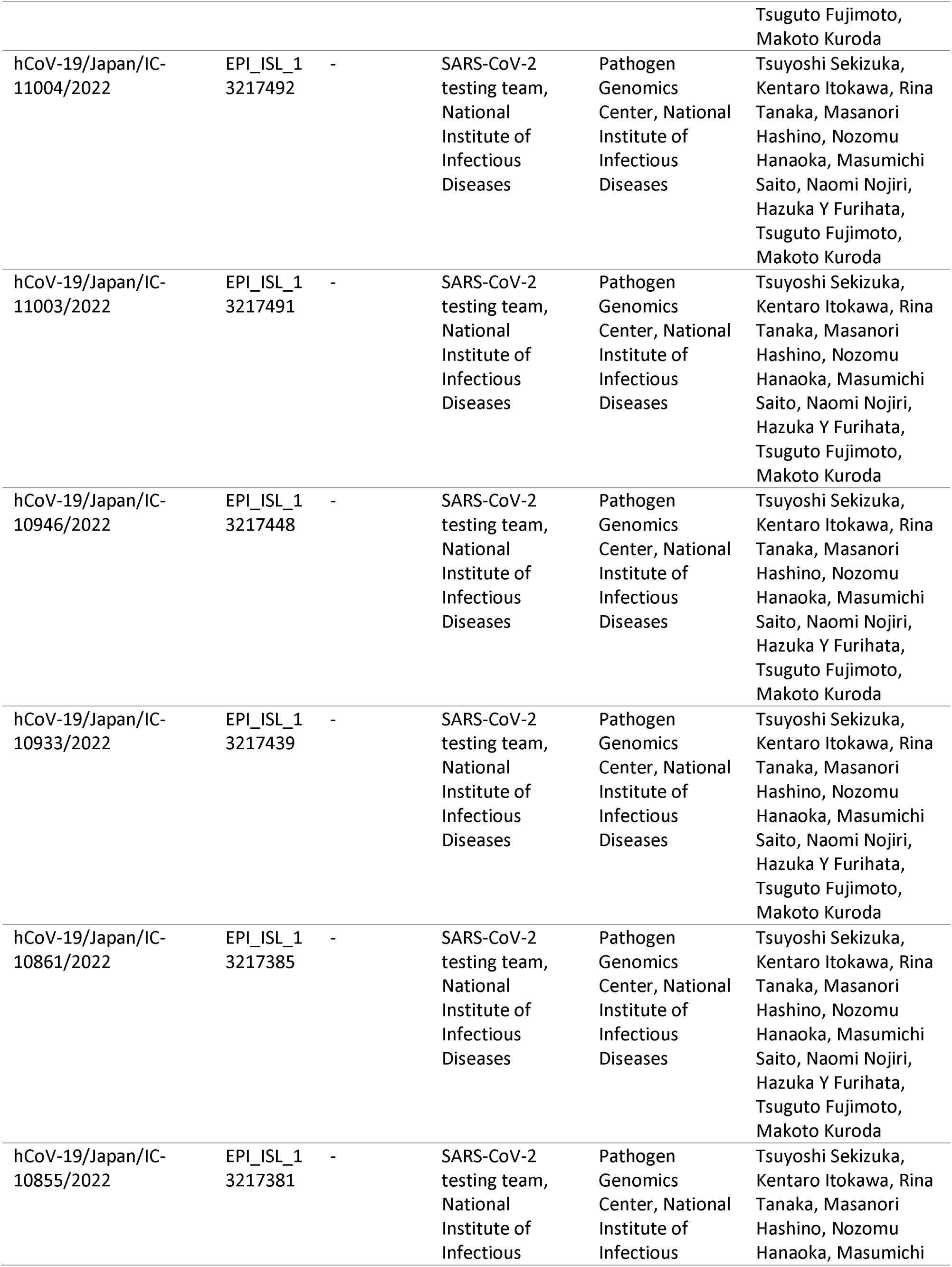
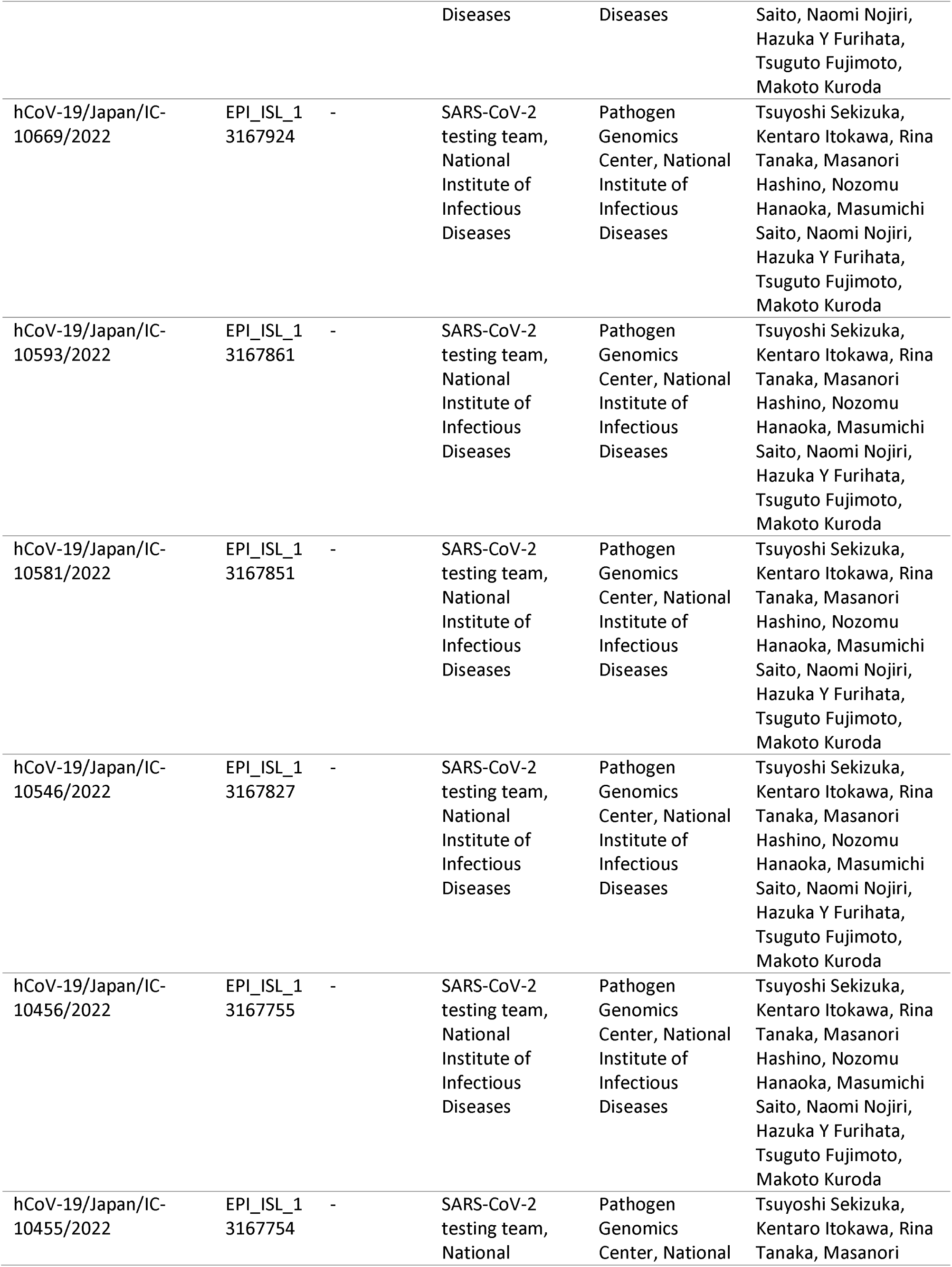
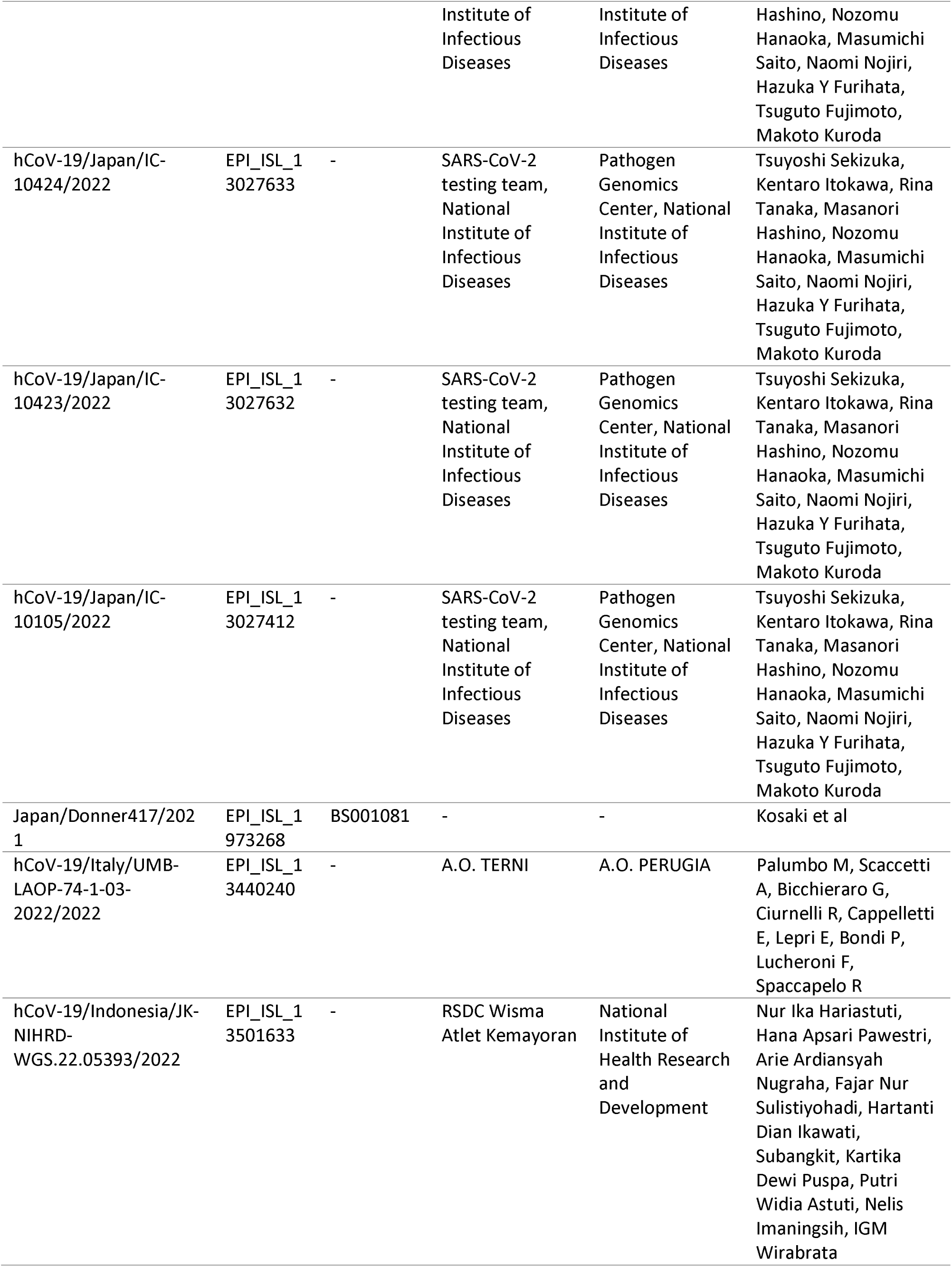
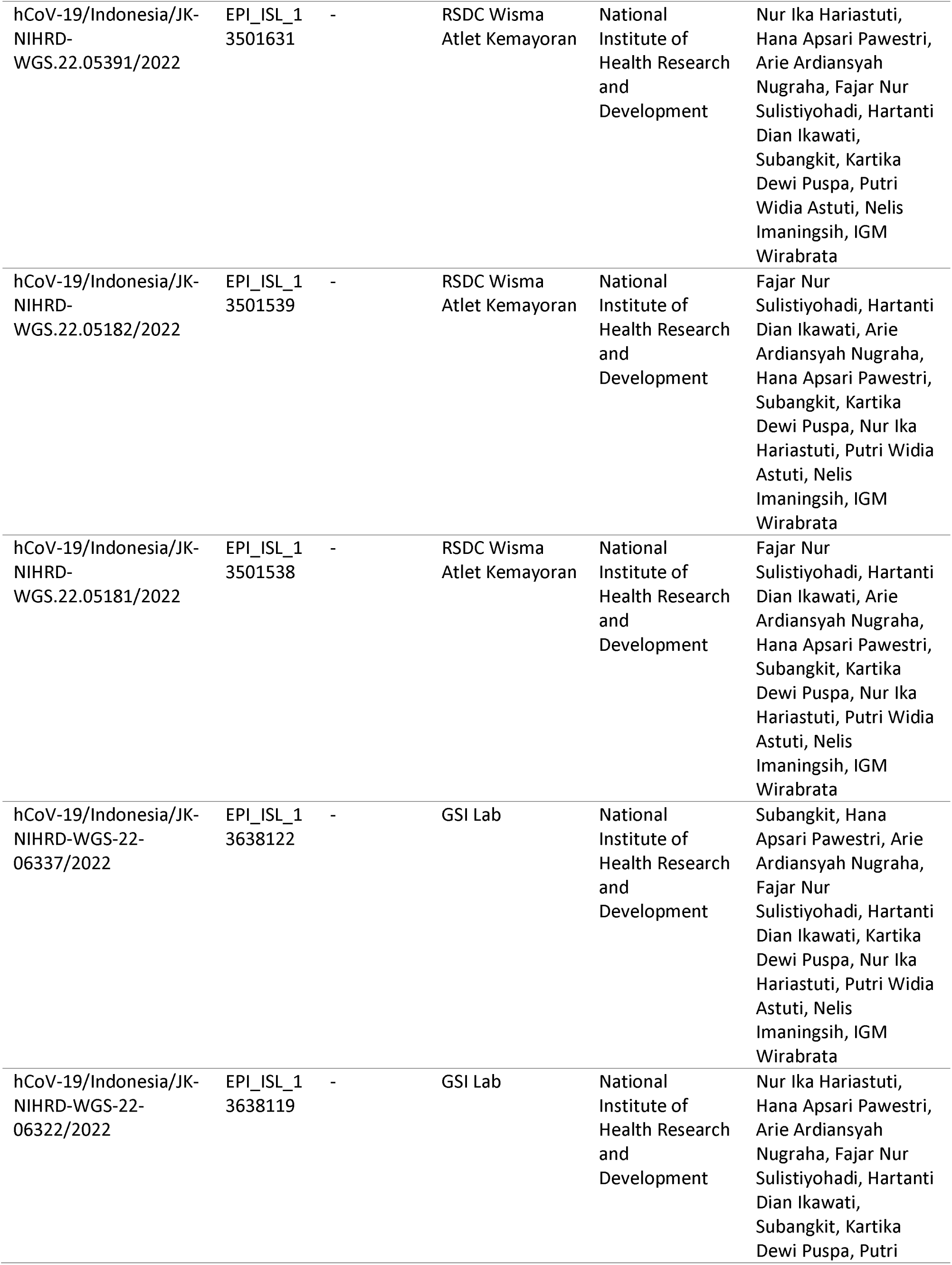
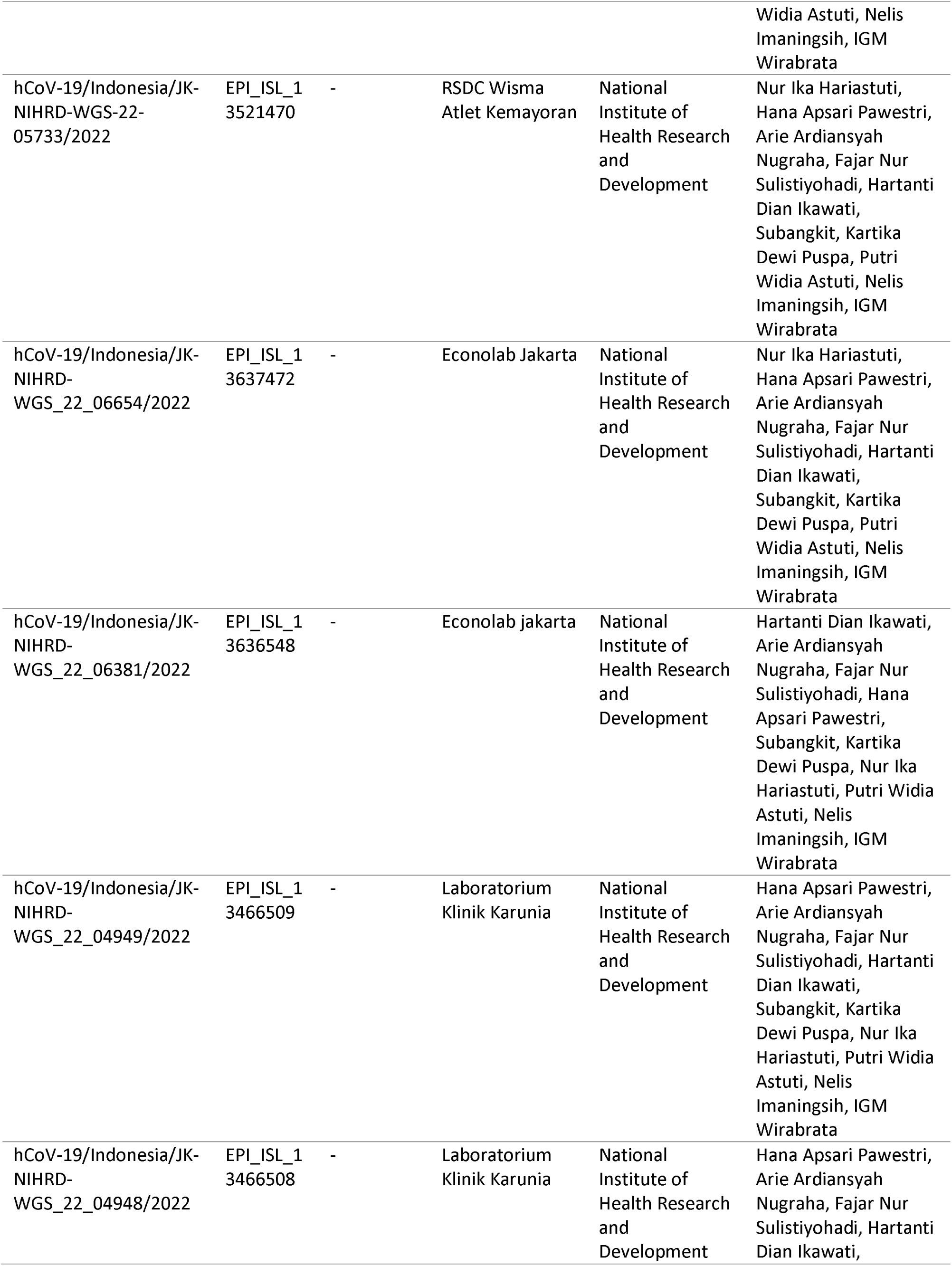
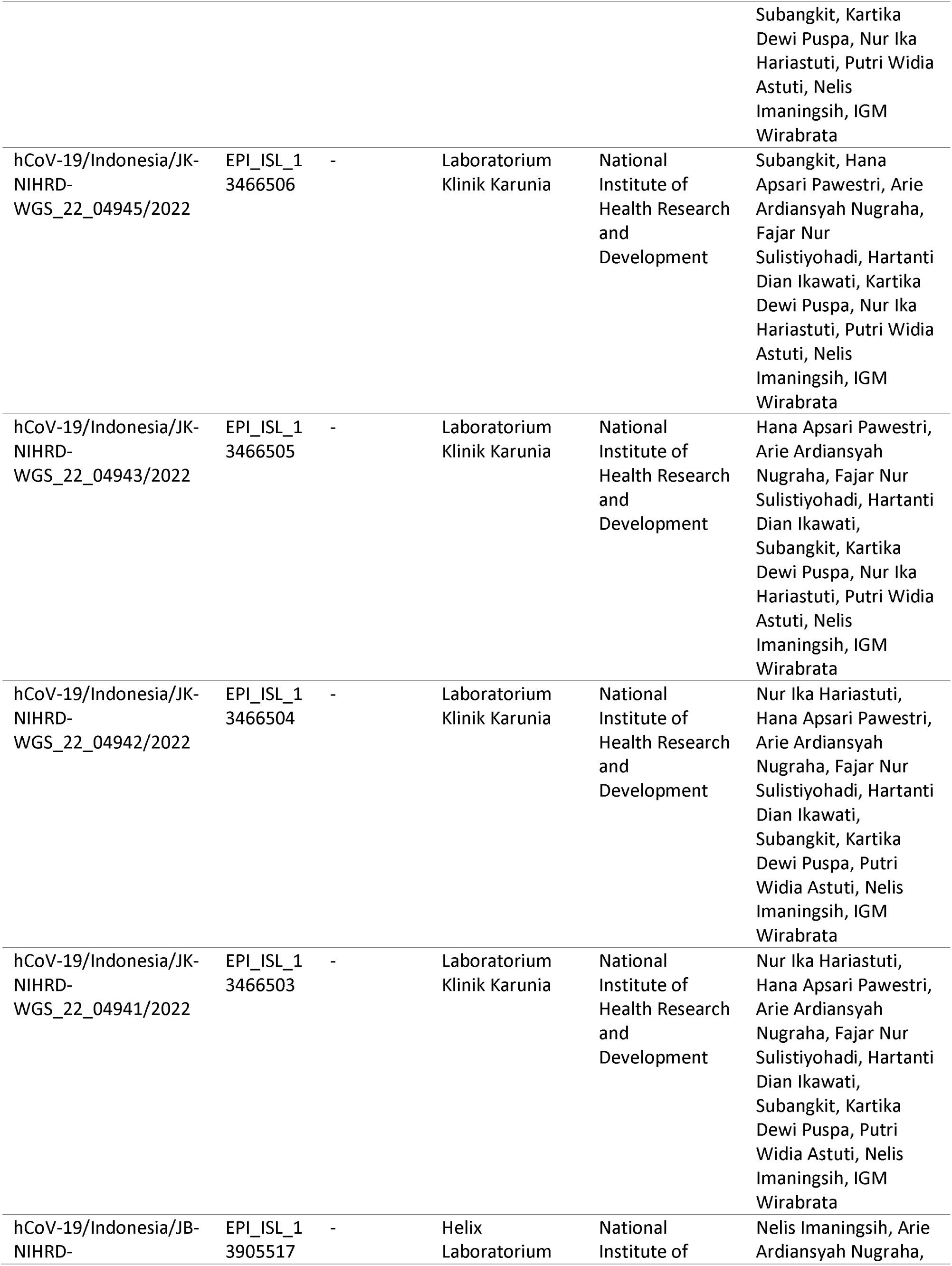
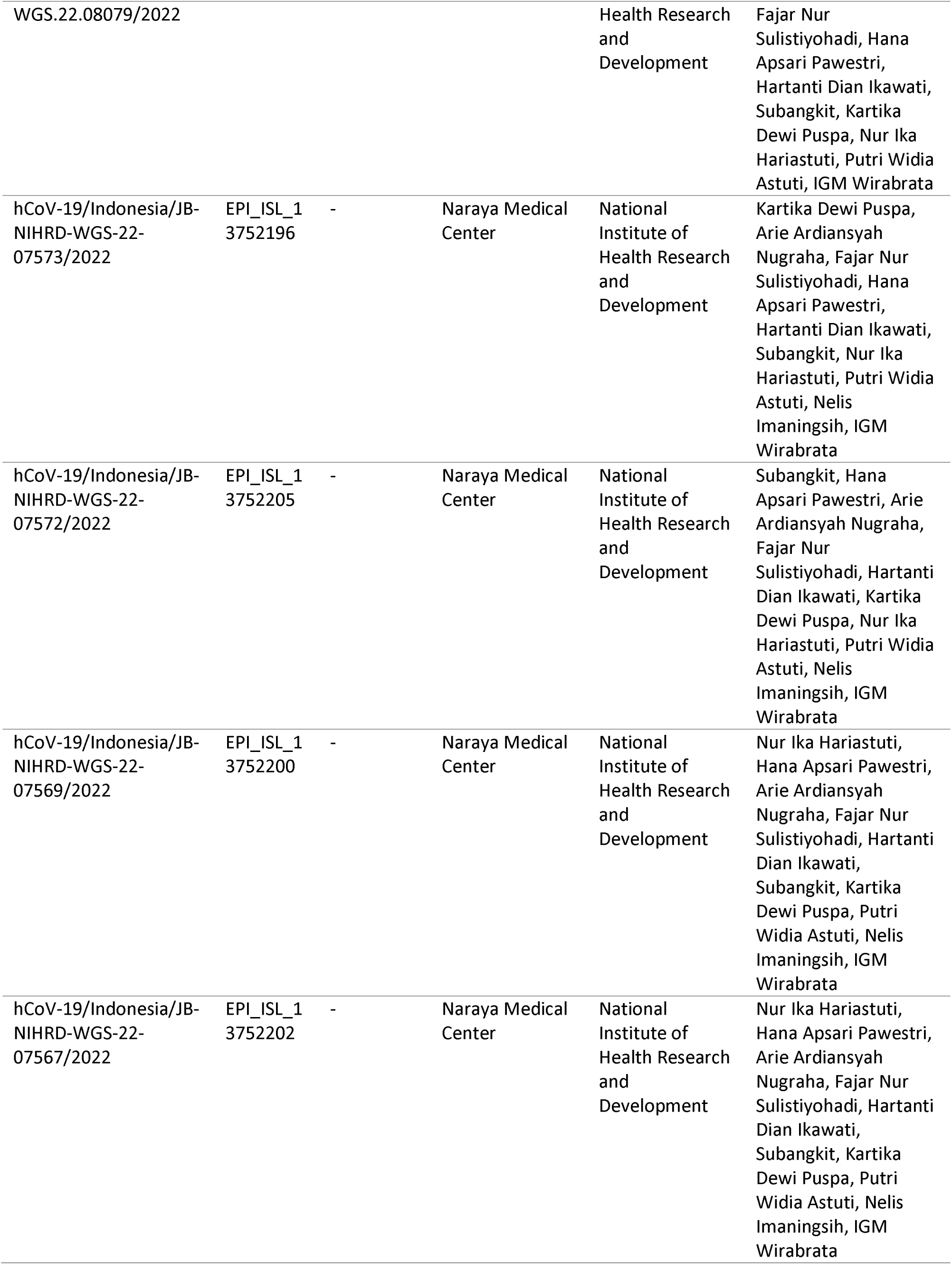
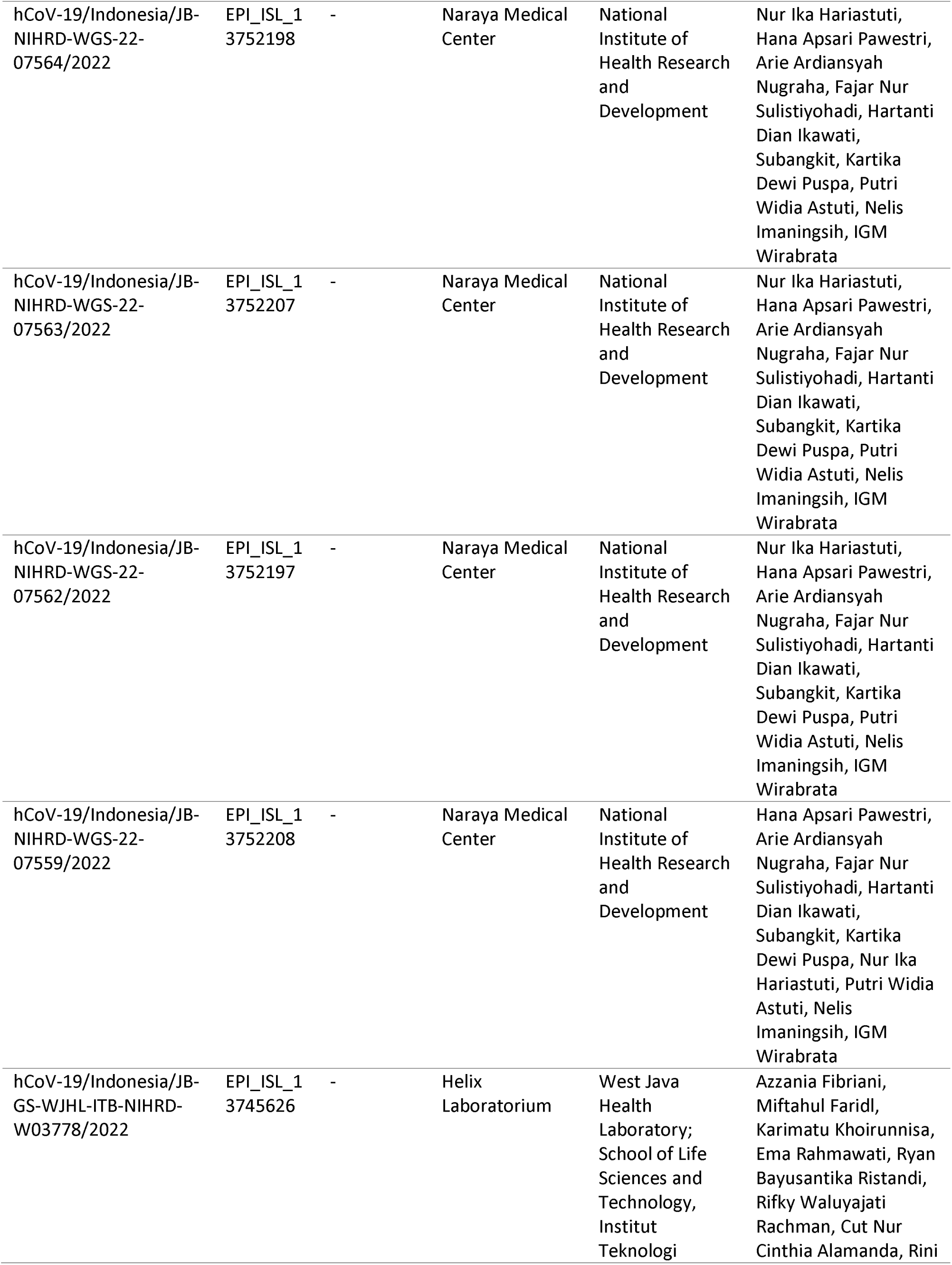
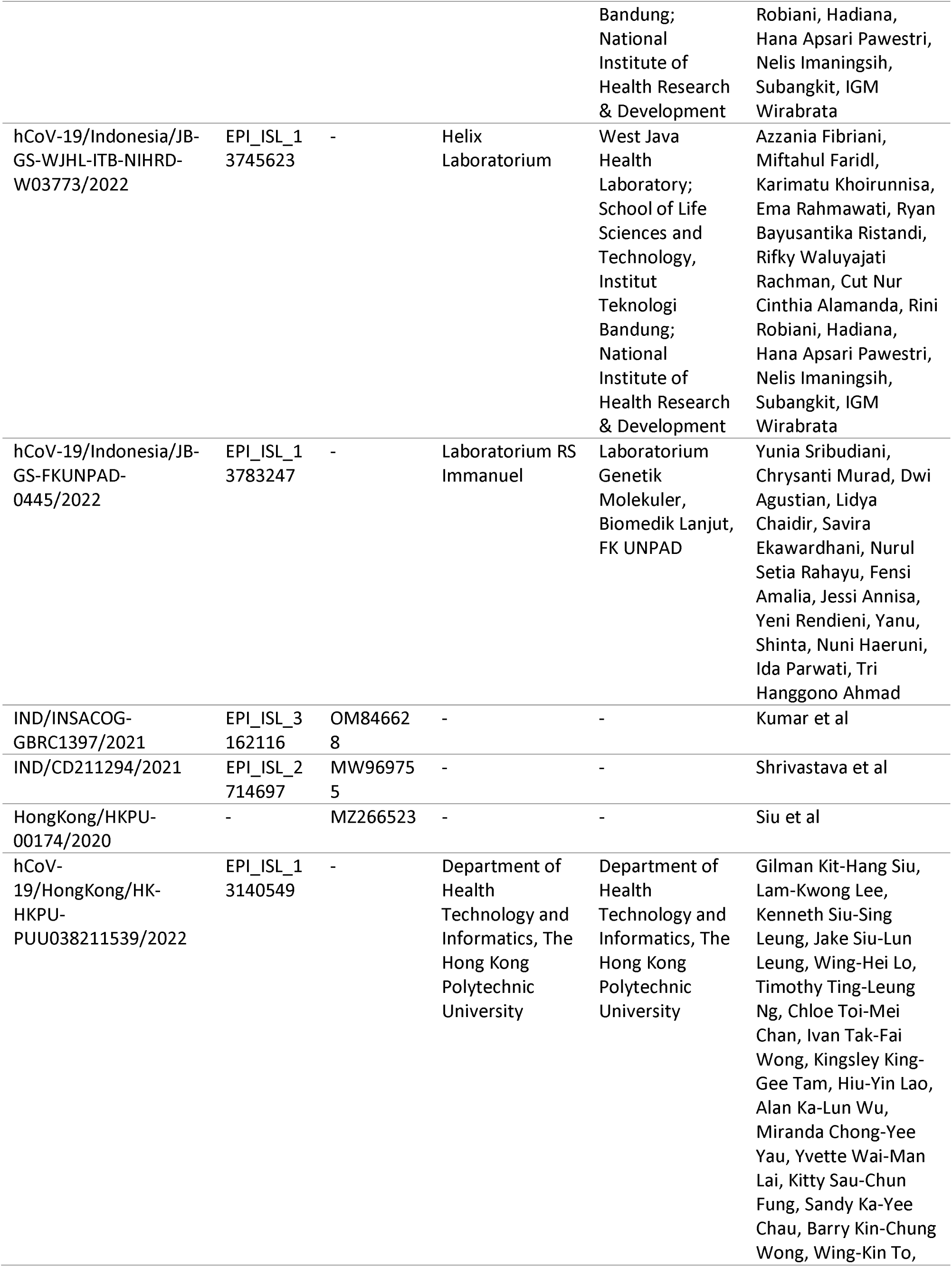
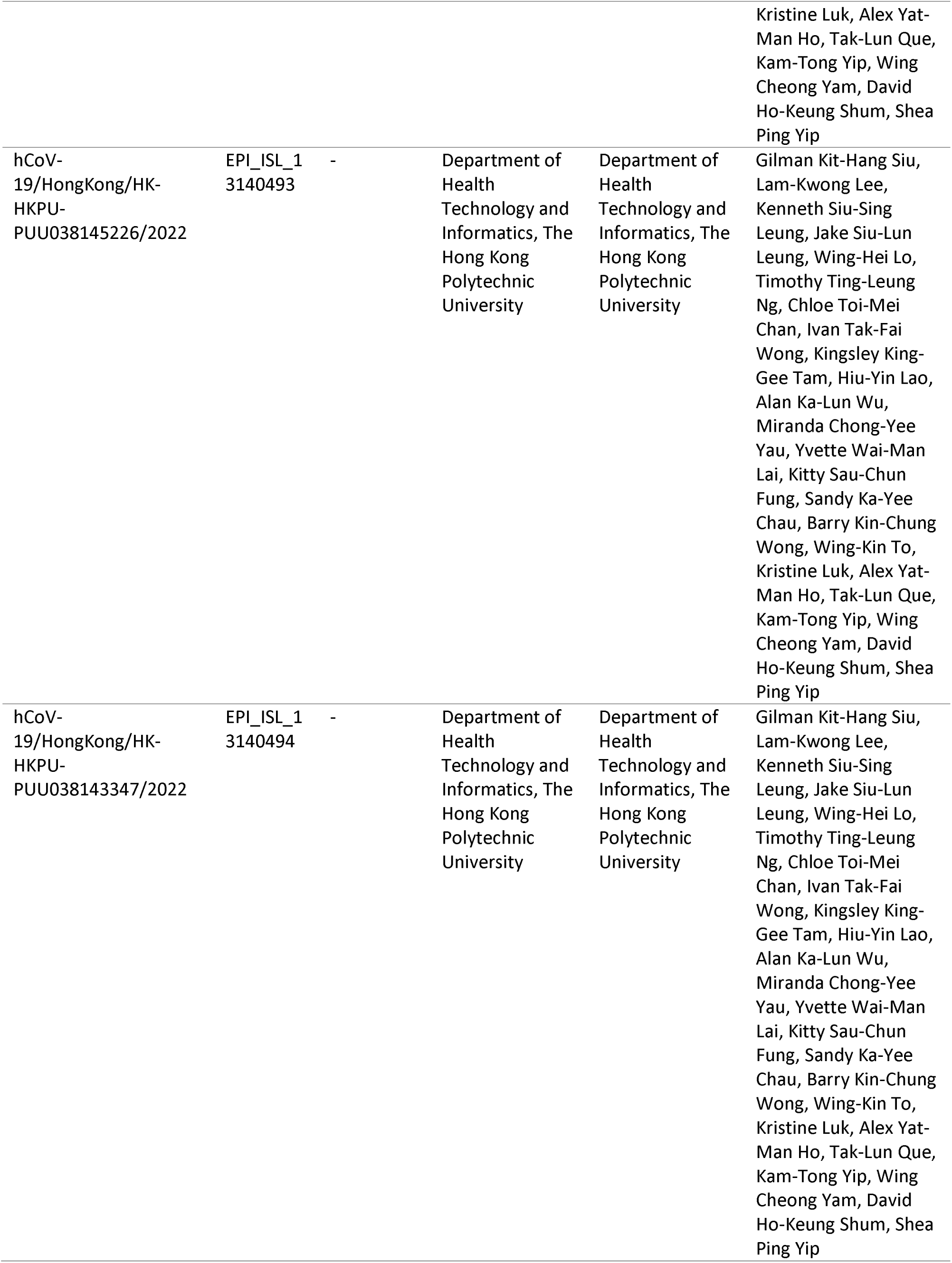
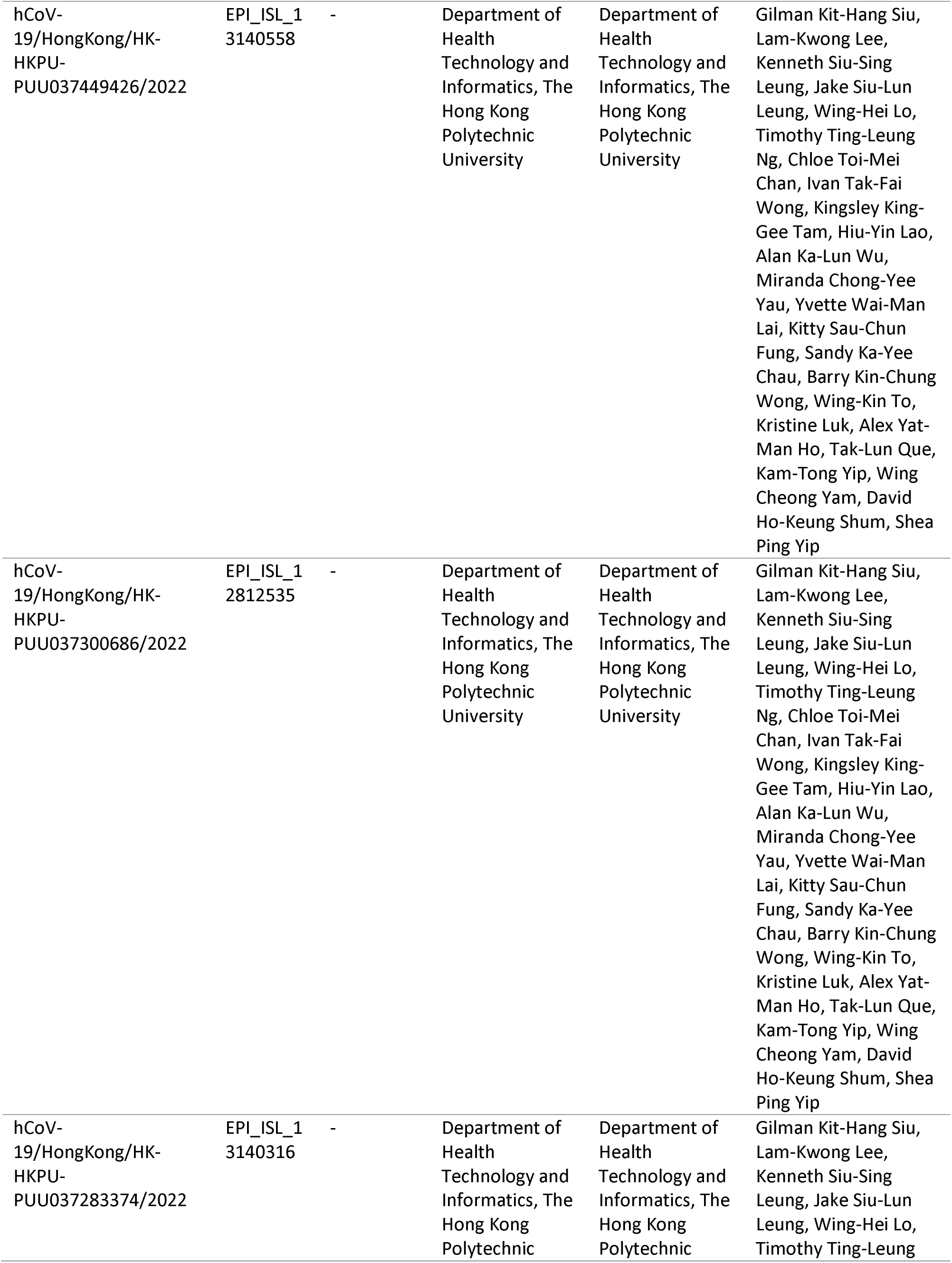
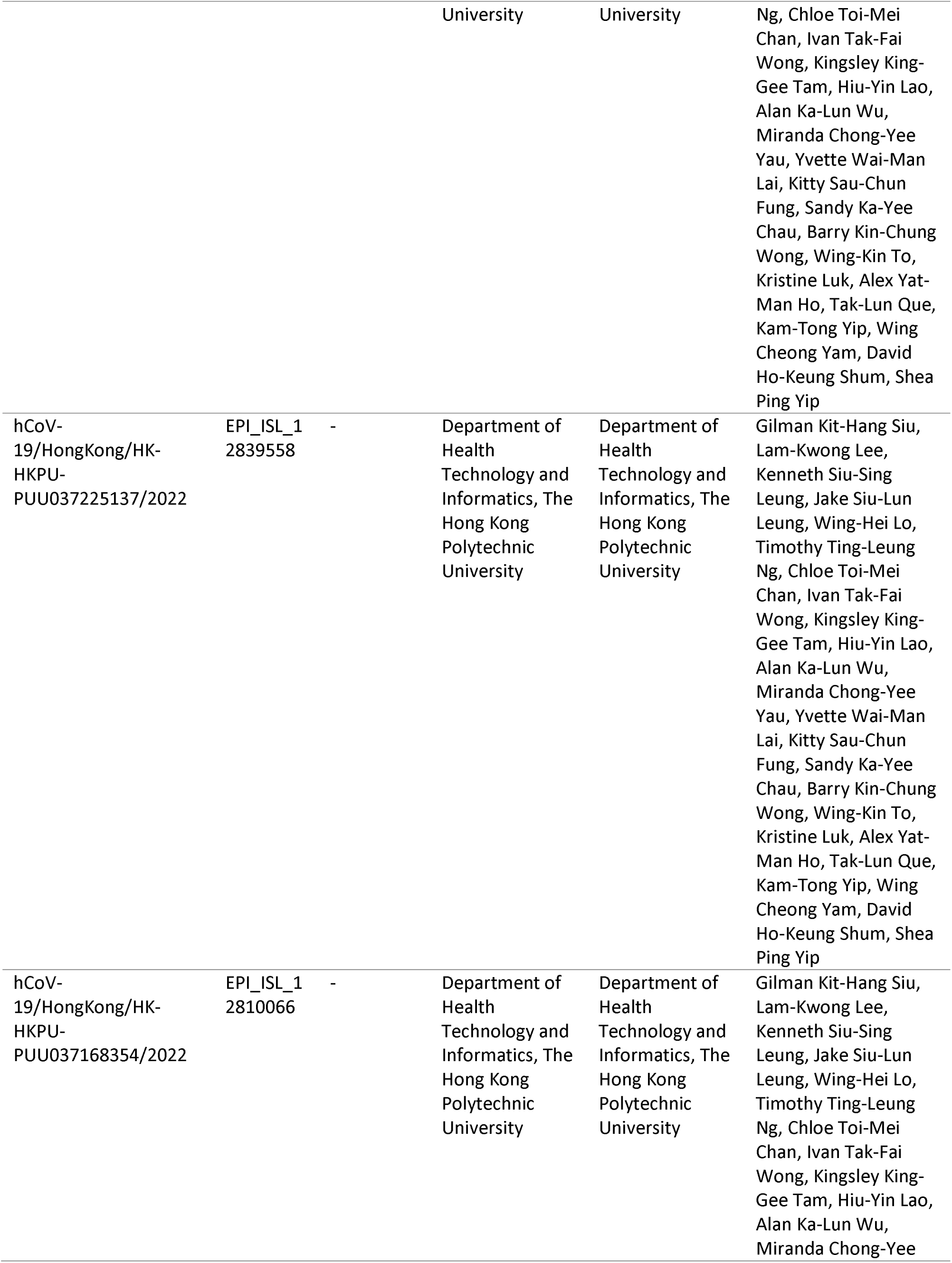
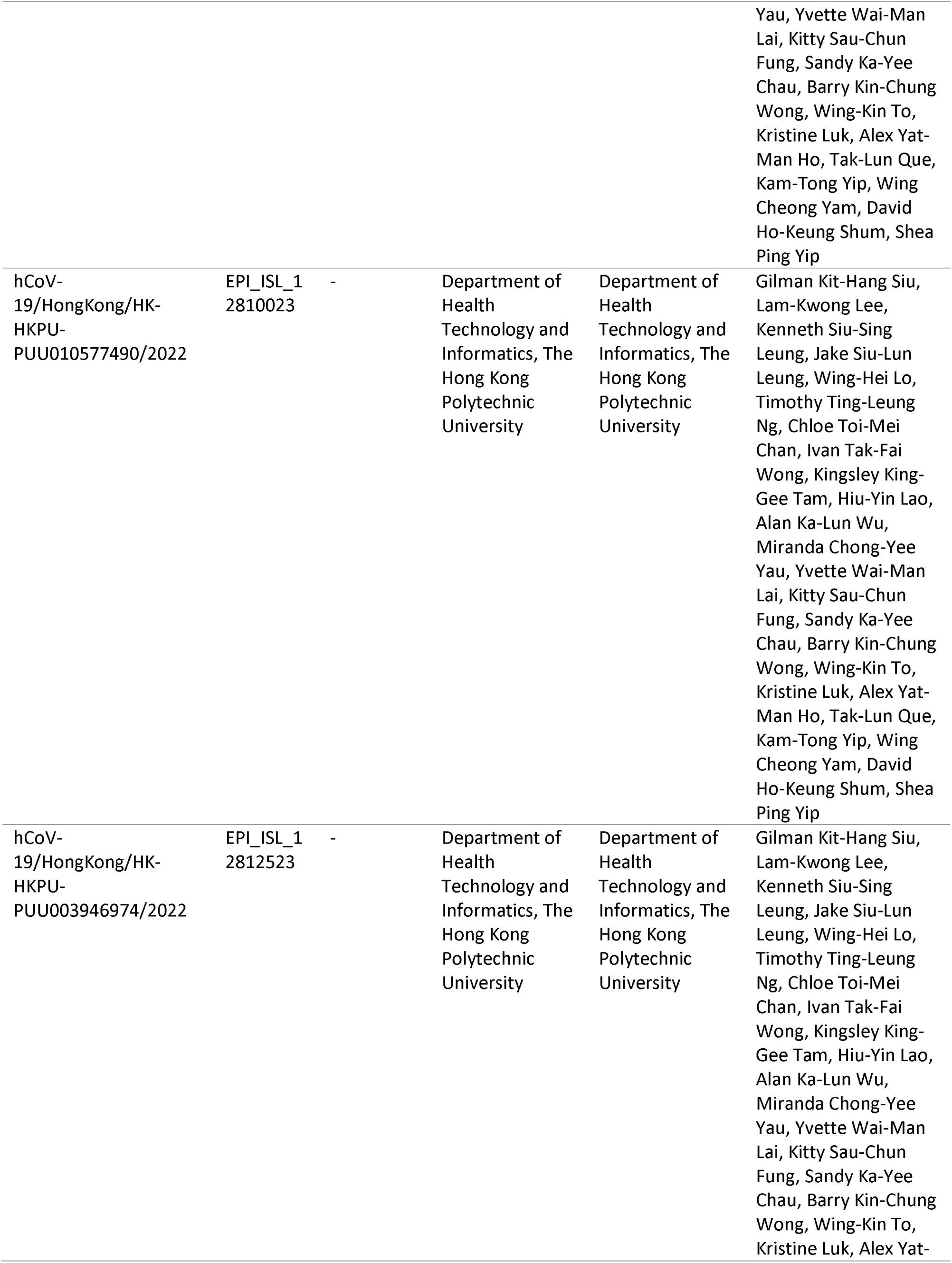
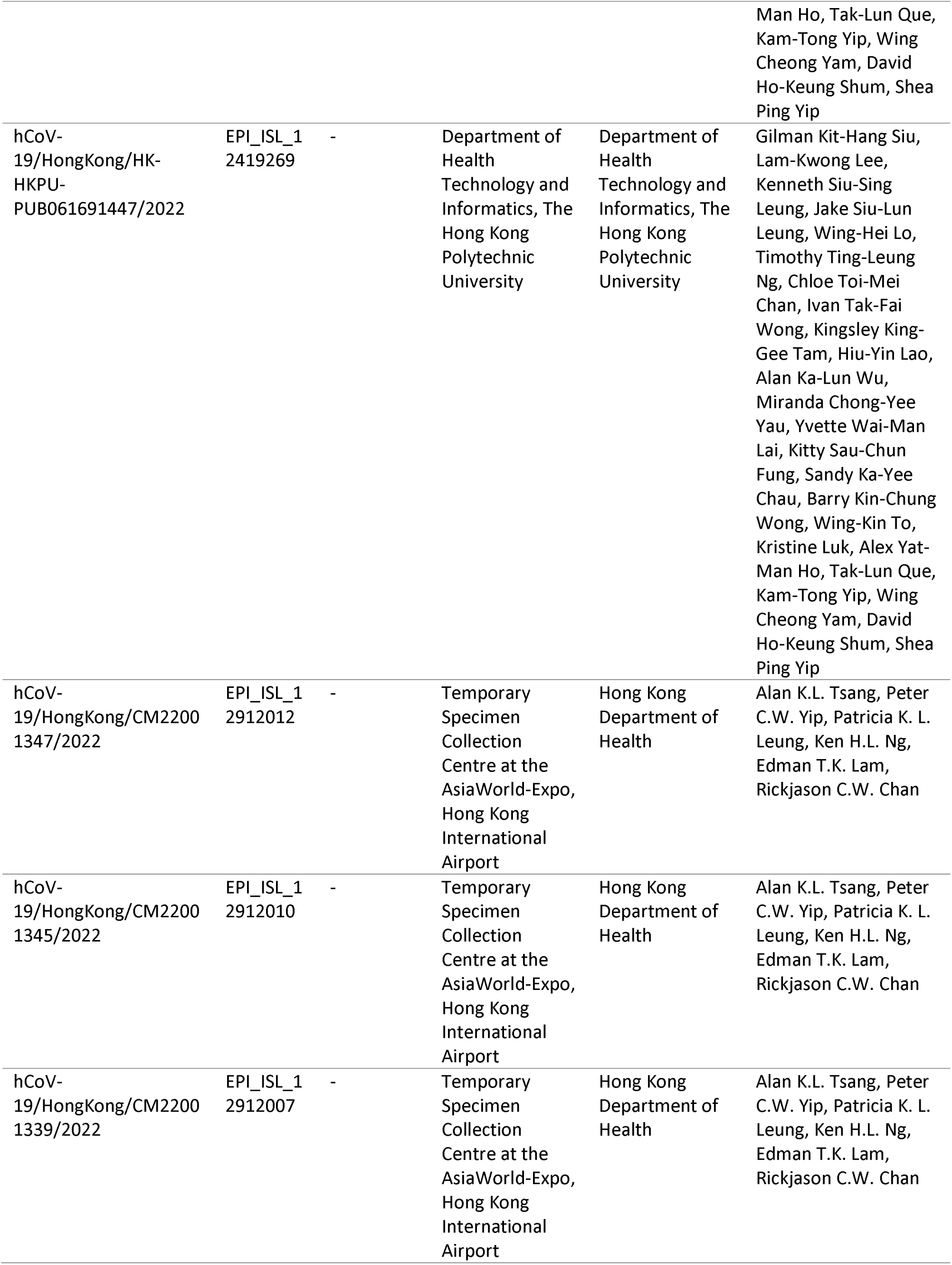
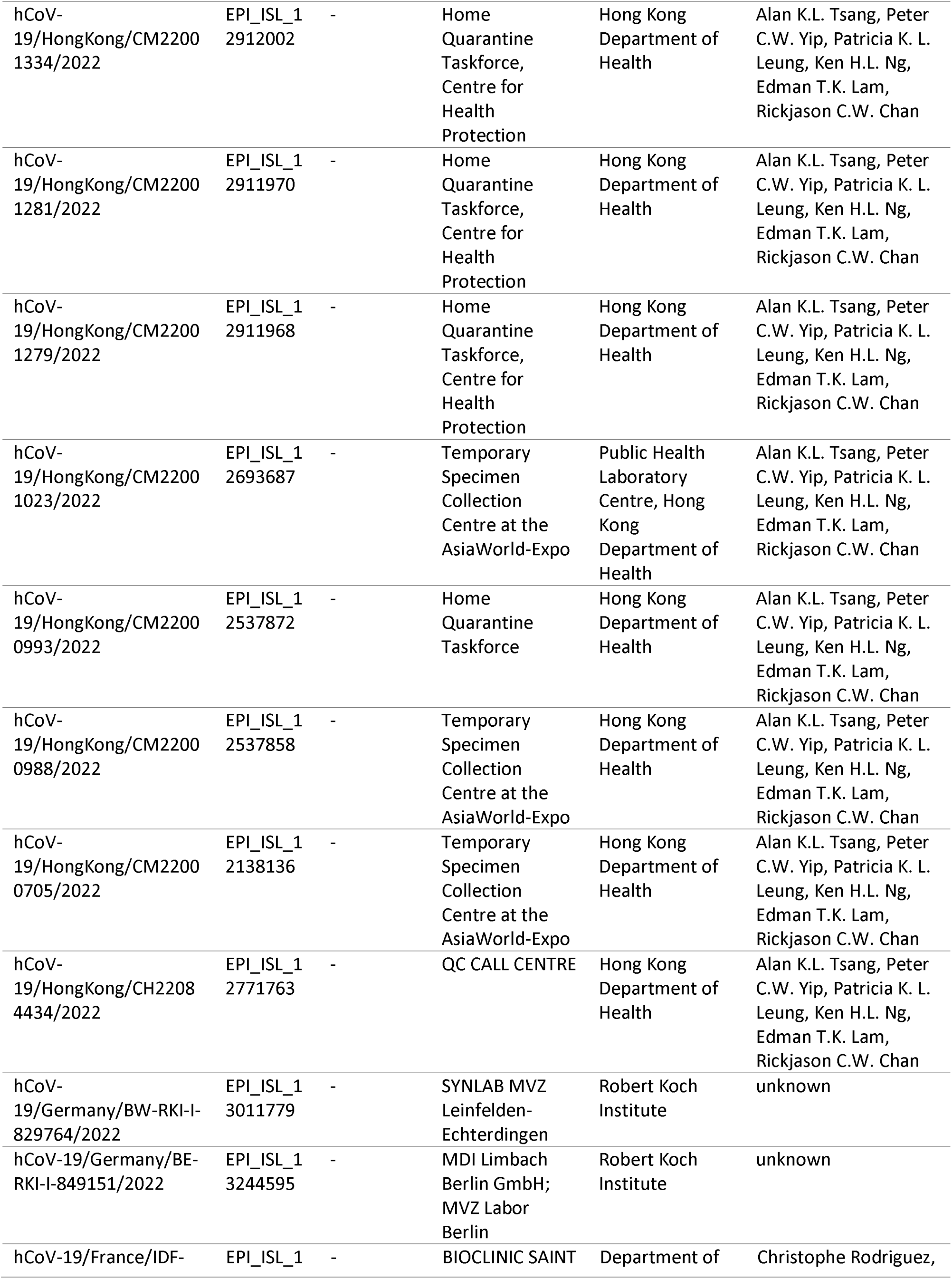
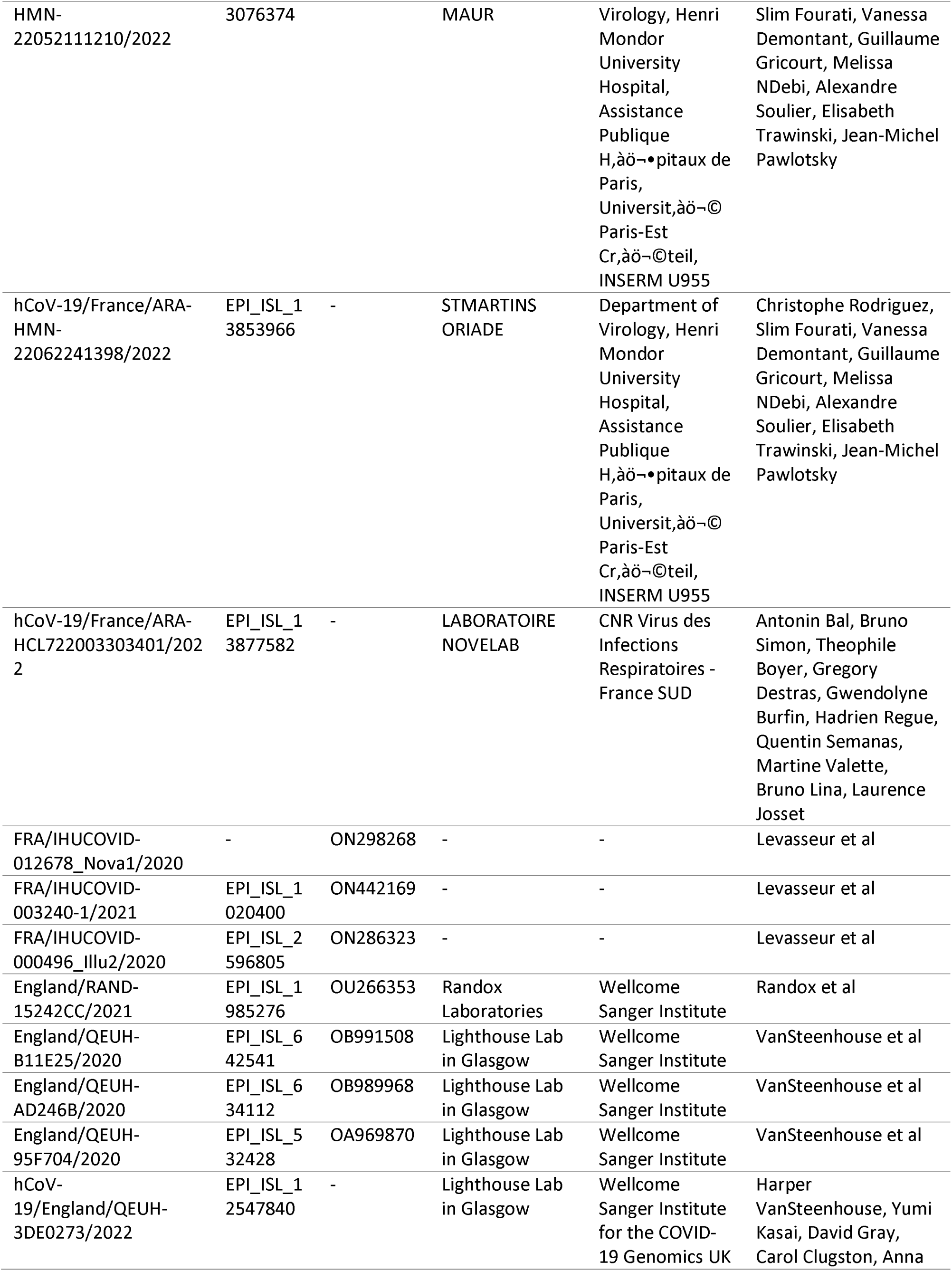
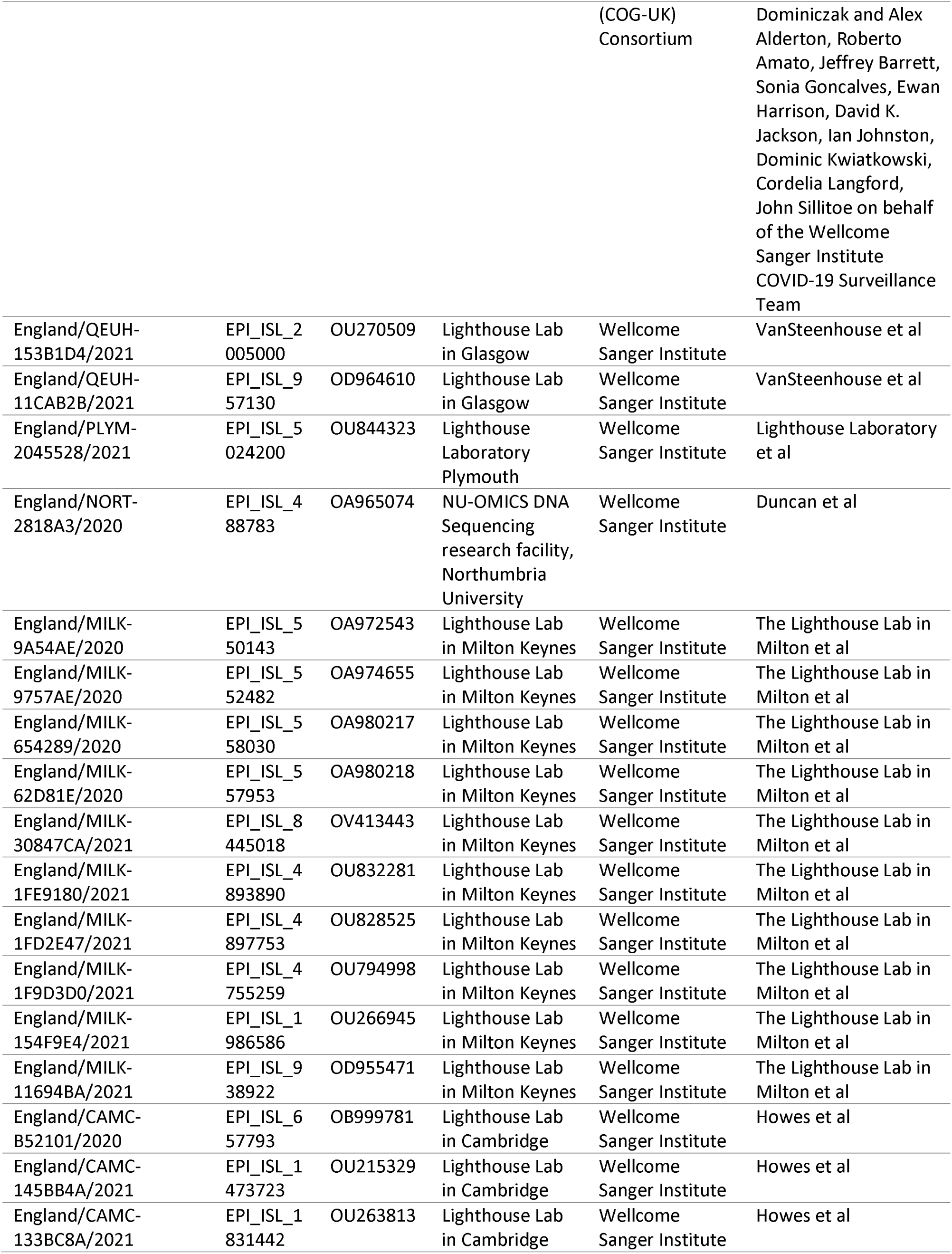
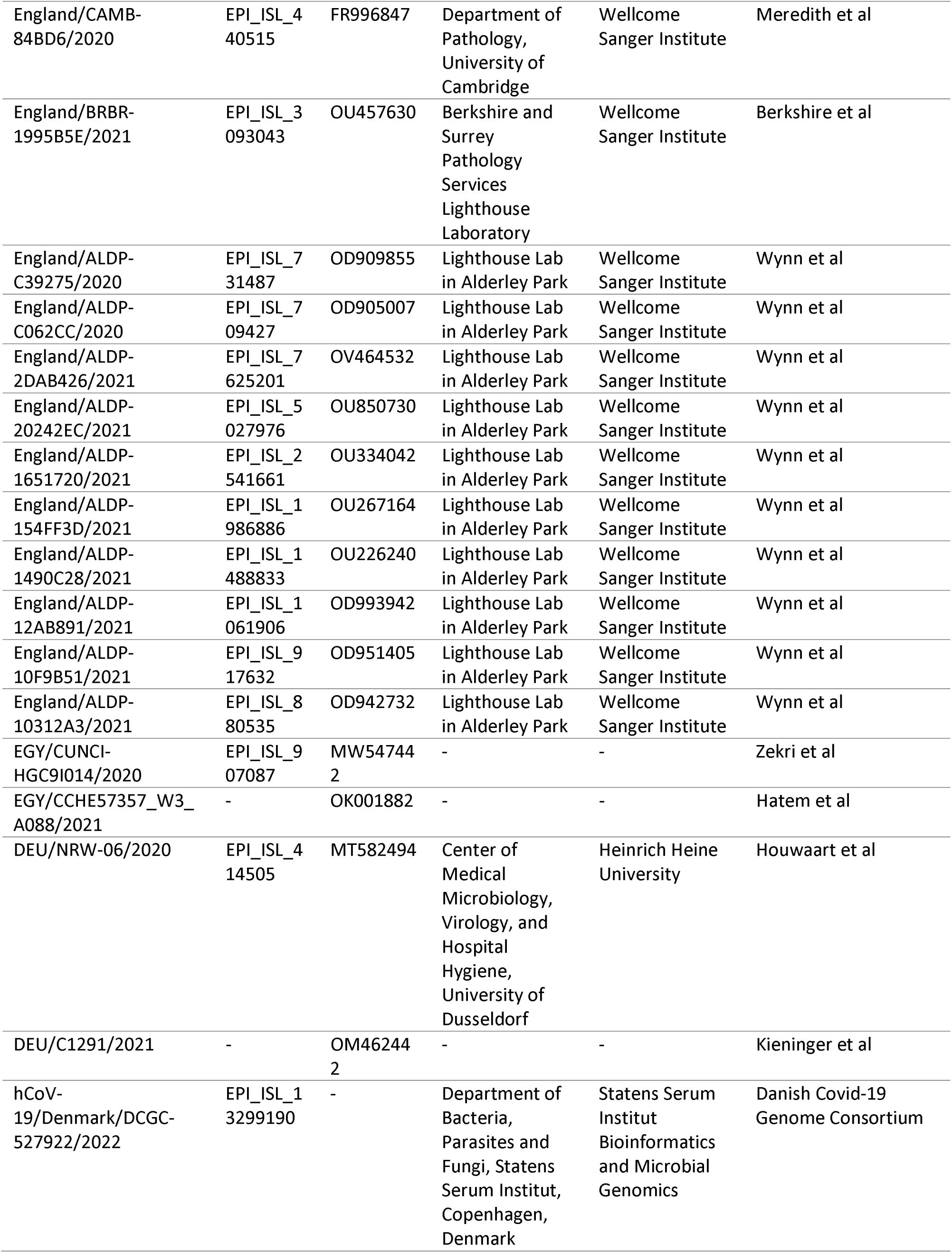
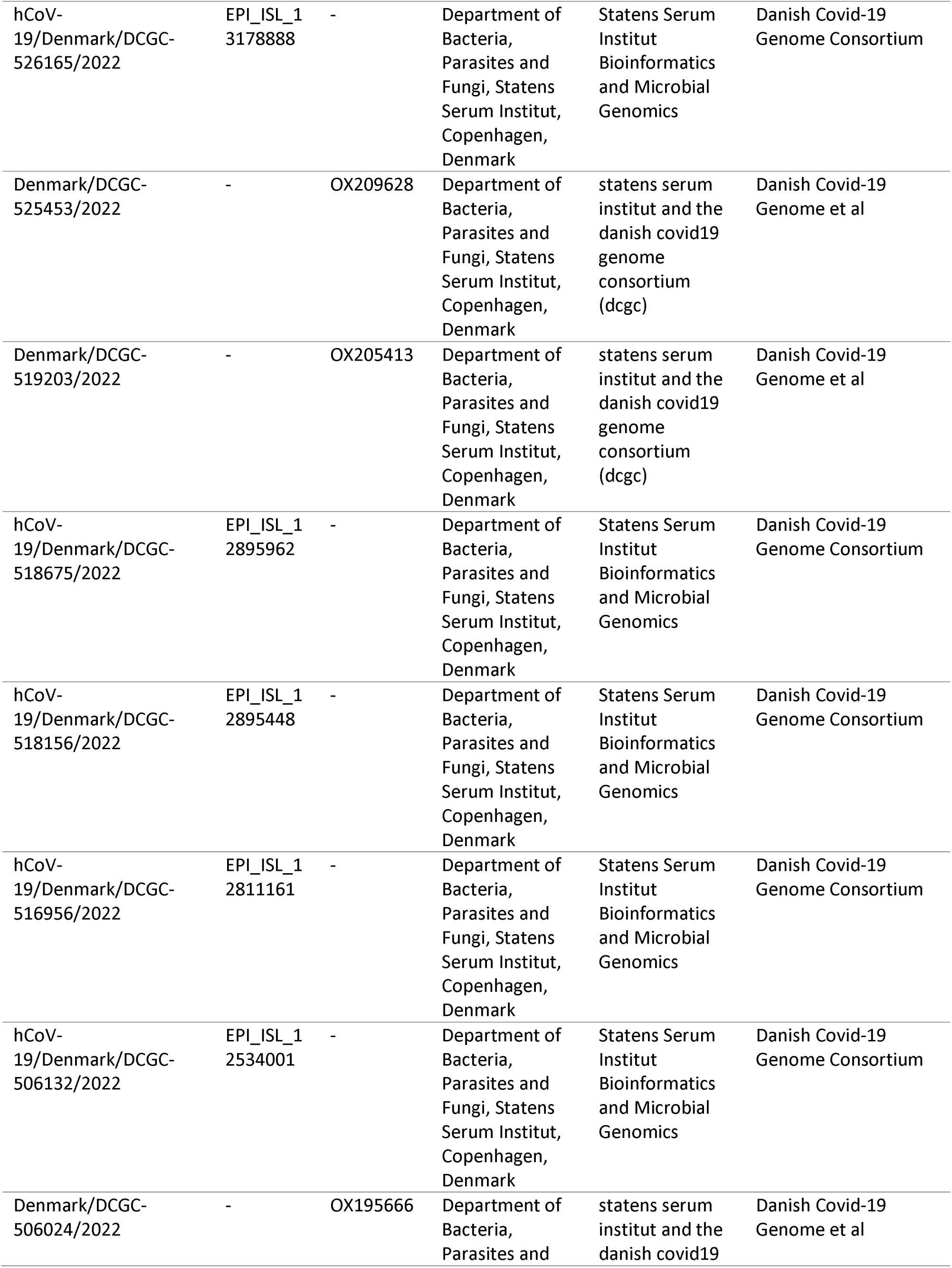
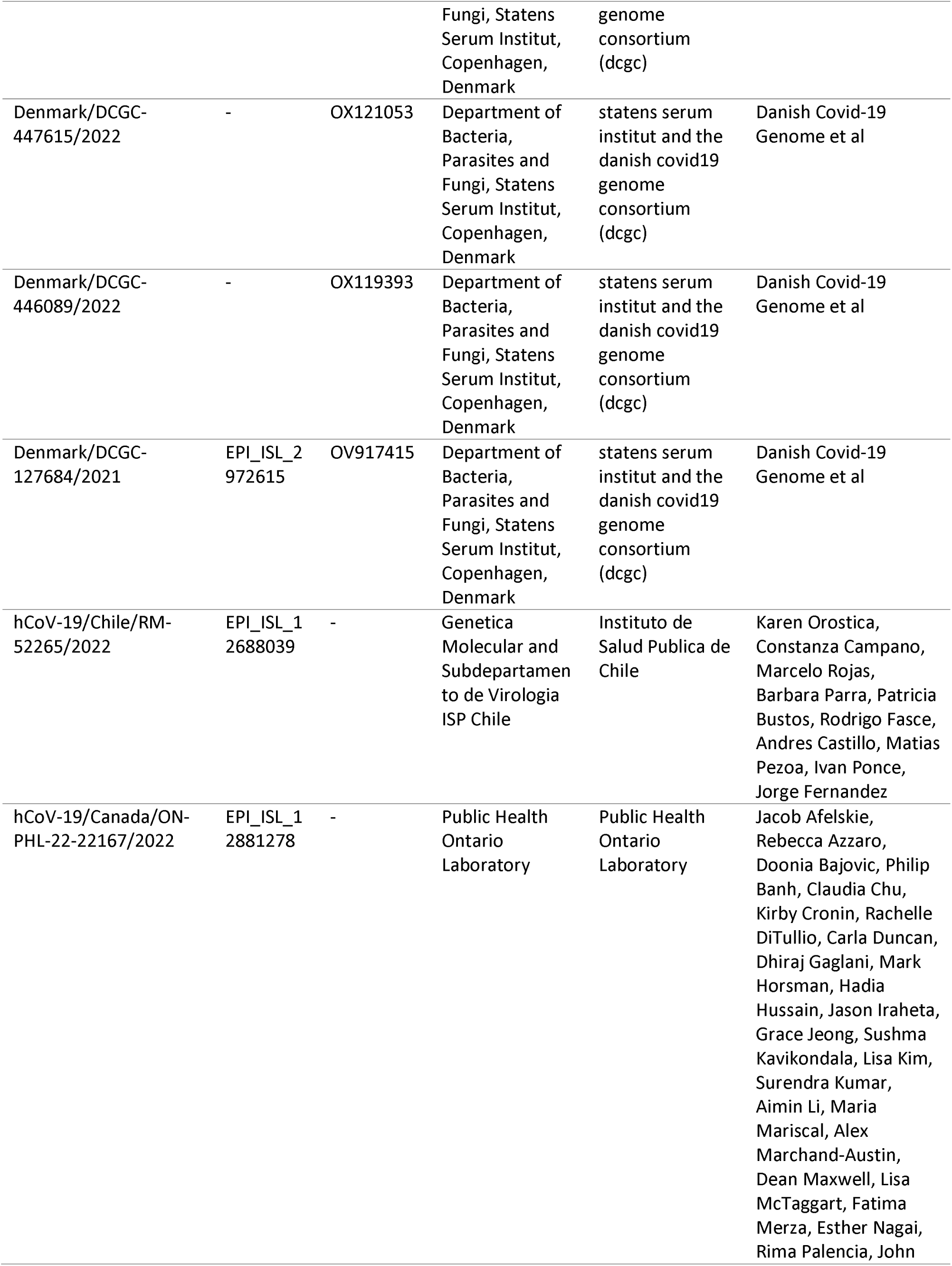
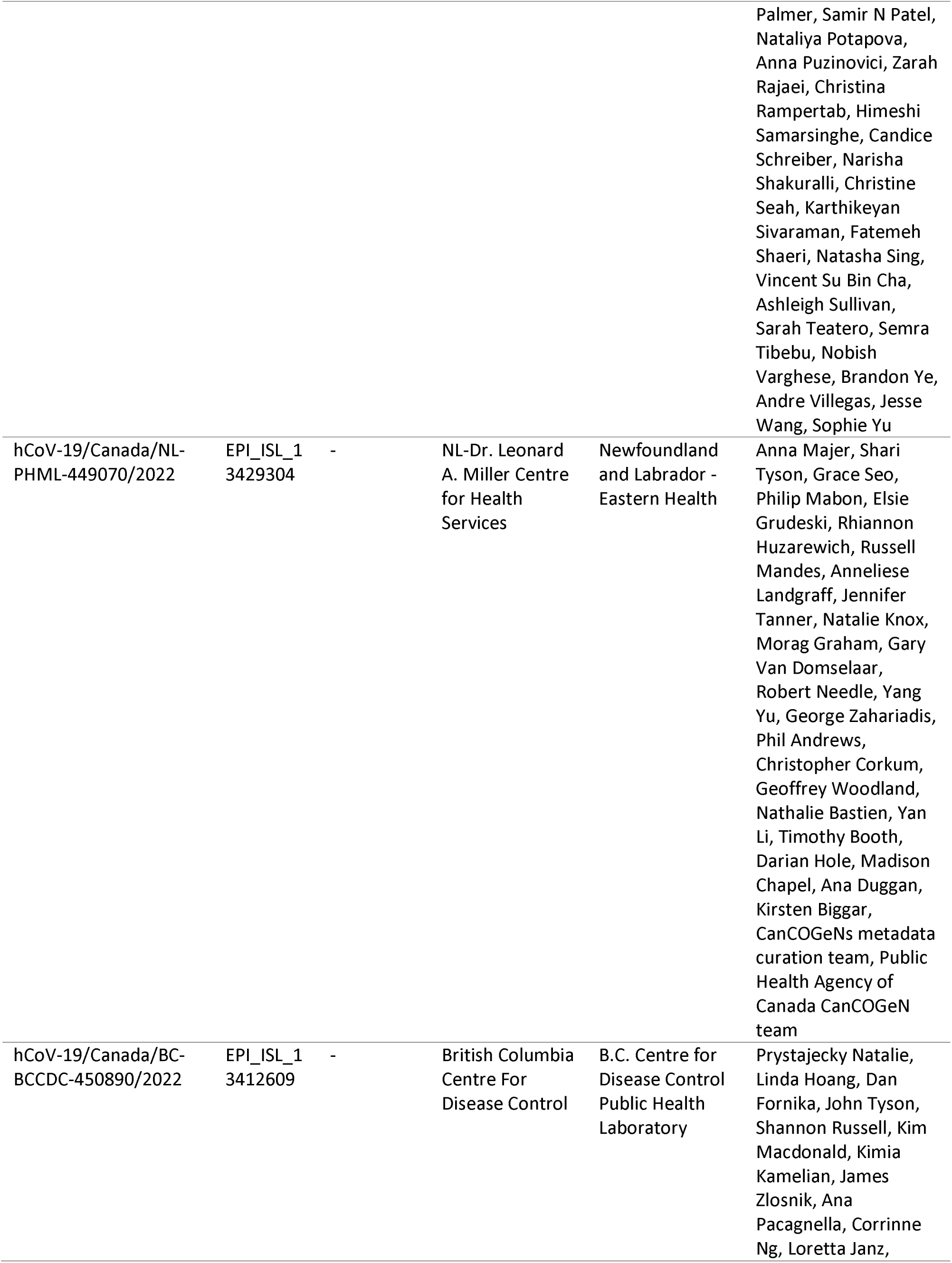
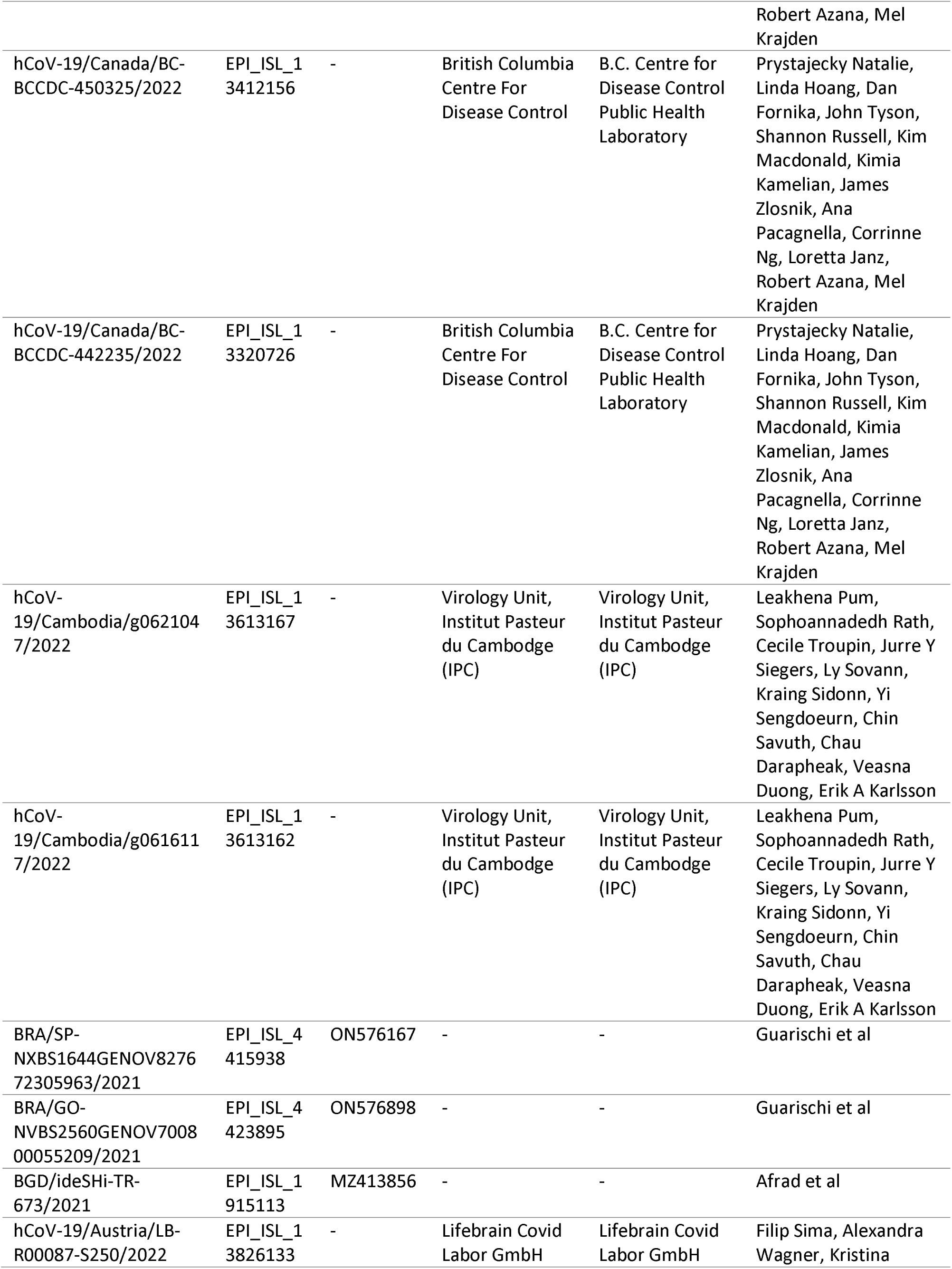
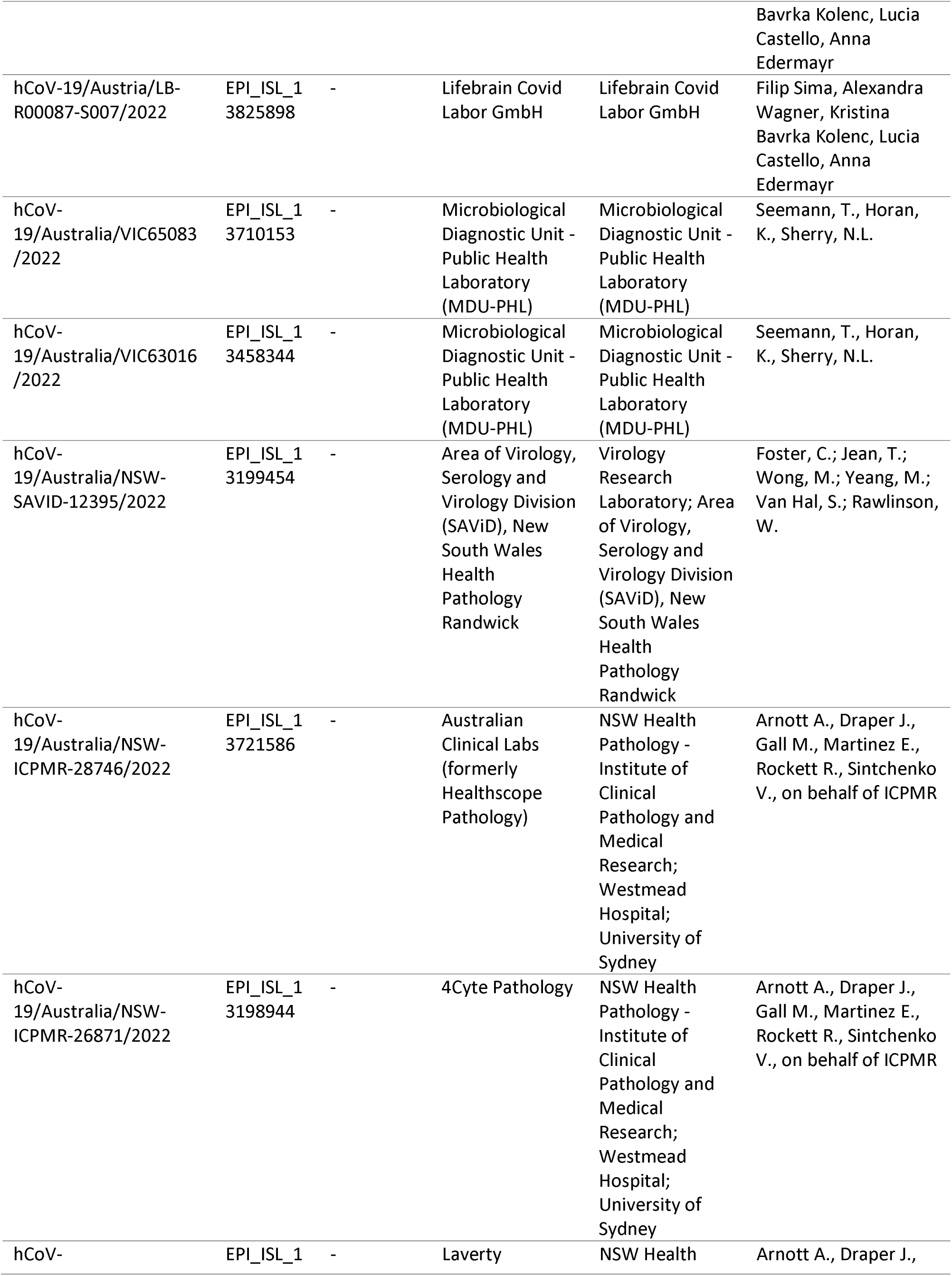
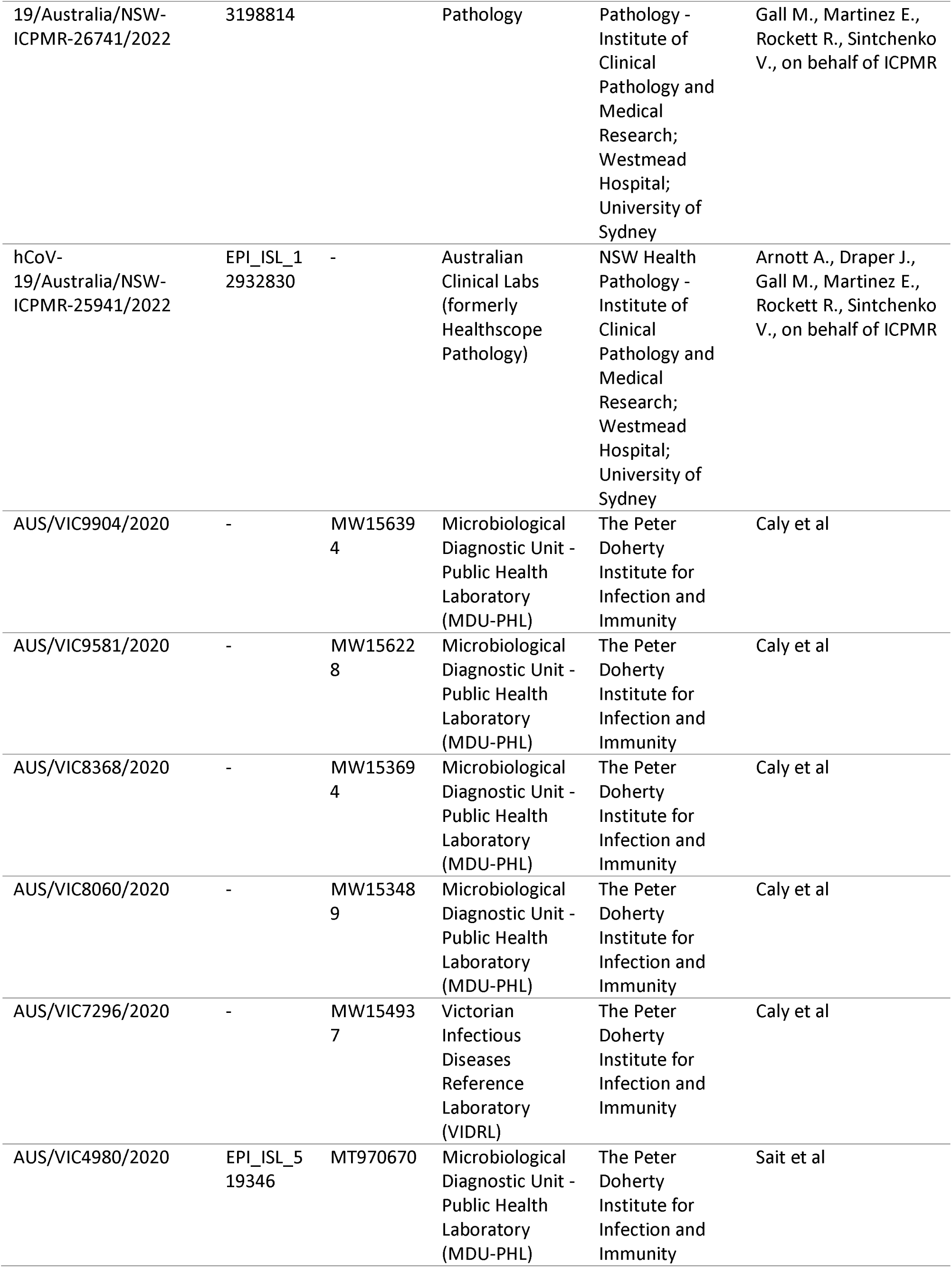
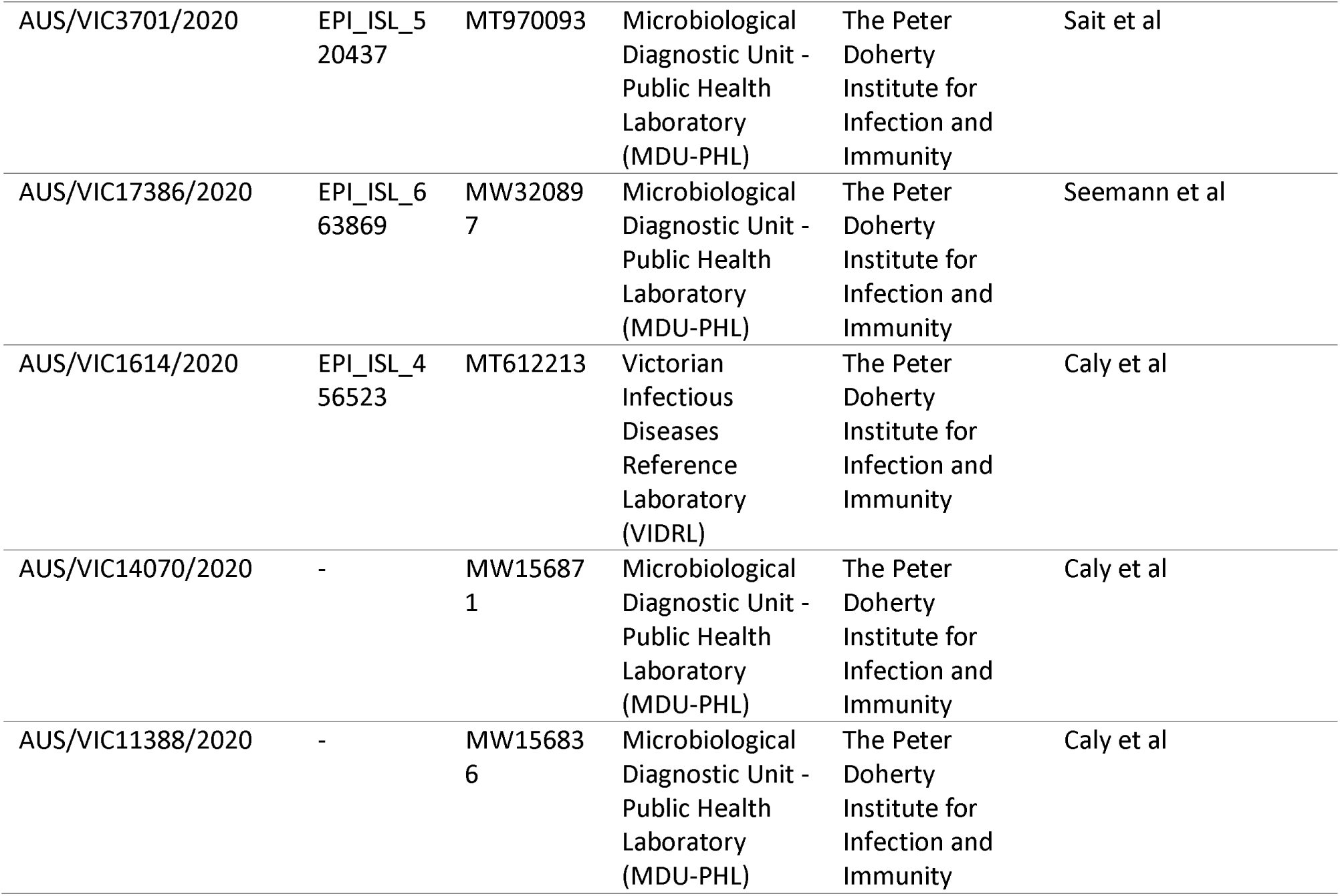
Acknowledgement of public sequences retrieved from GISAID and NCBI GenBank. We gratefully acknowledge the following Authors from the Originating laboratories responsible for obtaining the specimens, as well as the Submitting laboratories where the genome data were generated and shared via GISAID and GenBank, on which this research is based.

